# Increased Expression of *ZFPM2* Bypasses *SRY* to Drive 46,XX Testicular Development: A New Mechanism of 46,XX DSD

**DOI:** 10.1101/2024.05.27.24307993

**Authors:** Leah Ragno, Daphne Yang, Trisha R. Bhatti, Jonathan P. Bradfield, Diane K. Bowen, Katheryn L. Grand, Hakon Hakonarson, Thomas F. Kolon, Joseph Glessner, M. Celeste Simon, Nicolas Skuli, Maria G. Vogiatzi, Marie A. Guerraty, Matthew A. Deardorff, Louise C. Pyle

**Affiliations:** Children’s Hospital of Philadelphia, Philadelphia, Pennsylvania, 19104, United States

## Abstract

We present a patient with a novel cause of 46,XX ambiguous/androgenous genitalia Differences of Sex Development (DSD). Genome-wide array from blood showed 46,XX with ∼35% mosaic duplication of 76.5 Mb at chromosome 8q13.2-q24.3, containing 257 OMIM genes including *ZFPM2* and *CYP11B1*. Congenital adrenal hyperplasia testing was negative, testosterone was elevated, and the pro-testicular master regulator *SRY* was absent. The infant had a uterus, one streak ovary (<5% 8q duplication), and one testicle (75% 8q duplication). *We hypothesized that mosaic ZFPM2 duplication resulted in localized ZFPM2 overexpression and testicular development*. In typical testicular development, *ZFPM2* and its binding partner, *GATA4*, drive expression of the *SRY* master regulator. We completed RNA-seq of four Formalin-Fixed Paraffin-Embedded (FFPE) gonadal tissue samples from this mosaic individual to identify differentially expressed genes (DEGs). After quality control, two lines representing with and without the duplication were analyzed. In gonadal tissue containing the duplication, increased dosage of *ZFPM2* in a *Sry*-negative-46-XX individual appears to upregulate transcriptional activity of gonadal specific promoters such as *SOX9* and *AMH* via its protein interaction with known regulator of early testis development *GATA4. ZFPM2* is essential to the *GATA4*/*ZFPM2* transcription complex. Our results show that increased *ZFPM2* dosage enhances *ZFPM2*’s interaction with *GATA4* and results in upregulation of *SOX9* that is sufficient to initiate testis differentiation independent of *SRY. SOX9* (FC=39.2, *p*=6.1×10^−119^) and *SF-1* (FC=1.4, *p*=1.6×10^−3^) interact to produce the functional marker of fetal Sertoli cells *AMH* (FC=108.4, *p*=3.0×10^−54^) and inhibit female sexual differentiation. Several components of the testicular sex-development pathway were upregulated in addition to *SOX9* and *AMH*, including the pro-testicular transcription factor *MAP3K1* (FC=1.7, *p*=5.6×10^−17^), *DMRT1* (FC=13.9, *p*=2.1×10^−12^), *LHX9* (FC=2.5, *p*=5.0×10^−14^), *DHH* (FC=12.0, *p*=1.8×10^−30^), *PTGDS* (FC=2.5, *p*=3.4×10^−18^), and *SOX8* (FC=10.9, *p*=6.5×10^−10^). *ZFPM2* may function as a master temporal and spatial regulator of mammalian testicular organogenesis whose increased dosage elicits significant and cascading downstream effects. Further, components of the ovarian *WNT*-signaling pathway were repressed, including *LEF1* (FC=-3.7, *p*=1.4×10^−21^) and *FOXL2* (FC=-8.1, *p*=2.9×10^−39^). We have shown that increased *ZFPM2* dosage can induce 46,XX testicular development in a manner not dependent on *SRY*. This contravenes the previous understanding that *GATA4*/*ZFPM2* drives testicular development through *SRY. ZFPM2* may modulate numerous critical sex-development genes including transcription factors otherwise thought to be downstream of *SRY* (*MAP3K1, SOX9, AMH*). Findings from this single high-yield patient demonstrate that the primary role of *ZFPM2* in testicular development may be independent of *Sry*. This adds *ZFPM2* to the brief (<10) list of genes capable of directing testicular development in the 46,XX context, absent *SRY*. Overall, new understanding of these genes demonstrates that the role of *SRY* as a “master regulator” of testicular development may be less than previously thought.

## Introduction

Sex-specific gonadal development starts with activation of the ovary-specific pathway or the testis-specific pathway (Eggers, Ohnesorg, & Sinclair, 2014). In humans, the development of a typical male or female gonadal phenotype depends primarily on the presence of the Sex-determining Region Y (*SRY*) gene found on the Y chromosome (Eggers et al., 2014). When *SRY* is activated in males, it acts as a switch that diverts the fate of the undifferentiated gonadal primordia, the genital ridges, towards testis development. This sex-determining event sets in train a cascade of morphological changes, gene regulation, and molecular interactions that directs the differentiation of male characteristics (Wilhelm, Palmer, & Koopman, 2007). It is thought that in the absence of *SRY*, male sex organs cannot develop, and alternative molecular cascades and cellular events drive the genital ridges toward ovary development (Sinclair et al., 1990).

One of the general functions of *SRY* is to induce *SOX9* gene expression—*SOX9* expression is typically not induced absent *SRY*. The choice of somatic cell fate occurs following the upregulation of *SOX9* expression by *SRY*, and the subsequent maintenance of gonadal fate can be viewed as a battle for dominance between male (*DMRT1, SOX9*) and female (*FOXL2* and *WNT/β-catenin*) regulatory gene networks (Quinn & Koopman, 2012). Several models by which *SRY* activates *SOX9* are described, including a direct activation model by which *SRY* activates *SOX9* via cis-regulatory elements (Kanai, Hiramatsu, Matoba, & Kidokoro, 2005; Koopman, 1999).

Cases of aberrant sex determination involving atypical *SOX9* expression are well documented, including cases in which *SRY*-independent overexpression of *SOX9* triggers male-specific sex determination events—such as a case of *SRY*-negtive *SOX9* duplication leading to 46,XX male sex reversal (Huang, Wang, Ning, Lamb, & Bartley, 1999). Additionally, in mice, *SOX9* alone is sufficient to initiate testis differentiation absent *SRY*, making *SOX9* expression alone both necessary and sufficient for testicular development (Bishop et al., 2000; Qin & Bishop, 2005; Vidal, Chaboissier, de Rooij, & Schedl, 2001). Indeed, there is considerable evidence *SOX9* plays a pivotal role initiating or maintaining the upregulation of testis-specific genes while repressing ovarian-specific genes to direct the organogenetic fate of the bipotential gonads to that of testicular tissue.

Therefore, to accurately describe sex differentiation pathways, their overlap and interactions, and their spatial and temporal regulation, understanding direct or indirect activators and repressors of *SOX9* is critical. The *GATA4/ZFPM2* heterodimer complex is one transcription complex with potentially significant direct or indirect interaction with *SOX9. GATA4* is essential for testicular development and functions with its cofactor *ZFPM2*; *GATA4* and *ZFPM2* form a heterodimer complex that influences transcription of a number of genes in developmental systems, including genes that play a role in gonadal sexual differentiation in humans (Tevosian et al., 2002; van den Bergen et al., 2020).

In typical testicular development, *ZFPM2* and its binding partner *GATA4* drive expression of the *SRY* master regulator (Tevosian et al., 2002). In mice, there is evidence that *GATA4*/*ZFPM2* is requisite for *SOX9* expression (the next step downstream of *SRY*) and typical ovarian development. However, it is unclear whether *GATA4*/*ZFPM2* plays an essential role in testis differentiation subsequent to *SRY* activation. To drive *SRY*-negative XX female-to-male sex reversal, the ability of the *GATA4*/*ZFPM2* complex to regulate *SOX9* in the XX gonads (in the absence of the Y chromosome) must be independent from its ability to regulate *SRY*, the Y chromosome-linked gene.

Presently, there is substantial evidence of regulation of *SOX9* gene expression by *GATA4*/*ZFPM2* absent *SRY*. For example, *ZFPM2* was shown to be a limiting factor in the formation of a functional *GATA4*/*ZFPM2* transcription complex required for *SOX9* expression during gonadogenesis (Manuylov, Fujiwara, Adameyko, Poulat, & Tevosian, 2007). However, in mice, although *GATA4*/*ZFPM2* is necessary for testis determination, the mechanism of how this leads to testis formation is unclear (Bashamboo et al., 2014). Nonetheless, gonadal differentiation, sex determination, and normal *SRY* expression require direct interaction between transcription partners *GATA4* and *ZFPM2*, and abrogation of the *GATA4*/*ZFPM2* interaction or *ZFPM2* loss results in the equivalent defect in mouse gonadal differentiation (Tevosian et al., 2002; Viger, Mertineit, Trasler, & Nemer, 1998). Further, *GATA4*/*ZFPM2* function is required to maintain precise numbers of Sertoli cells in the developing gonads, and there is evidence that correct dosage of *GATA4* and *ZFPM2* transcription factors is critical for fetal testis development, with *SOX9* upregulation especially dependent on the proper dosage of *GATA4* and *ZFPM2* in mice (Bouma, Washburn, Albrecht, & Eicher, 2007; Manuylov et al., 2007).

Our data support an interaction between the *GATA4*/*ZFPM2* transcription complex and *SOX9* in a manner not dependent on *SRY*, suggesting that increased *ZFPM2* dosage can induce 46,XX testicular development absent *SRY*. This contravenes the previous understanding that *GATA4*/*ZFPM2* drives testicular development through transcriptional upregulation of *SRY*.

## Background

We present a patient with a novel cause of 46,XX ambiguous/androgenous genitalia Differences of Sex Development (DSD). Genome-wide array from blood showed 46,XX with ∼35% mosaic duplication of 76.5 Mb at chromosome 8q13.2-q24.3, containing 257 OMIM genes including *ZFPM2* and *CYP11B1*. Refined breakpoint analysis of the rearranged cell line showed a small 1.55 Mb deletion in the p arm of chromosome 8 and a large 76.46 Mb duplication in the q arm of chromosome 8 (**Figure 1**) (Pyle, Bhatti, Bowen, Kolon, & Deardorff, 2018). Congenital adrenal hyperplasia testing was negative, testosterone was elevated, and the pro-testicular master regulator *SRY* was absent. The infant had a uterus, one streak ovary (<5% 8q duplication), and one testicle (75% 8q duplication) (**Figure 2** and **Figure 3**). We hypothesized that mosaic *ZFPM2* duplication resulted in localized *ZFPM2* overexpression and testicular development. In typical testicular development, *ZFPM2* and its binding partner, *GATA4*, drive expression of the *SRY* master regulator.

**Figure 1:**
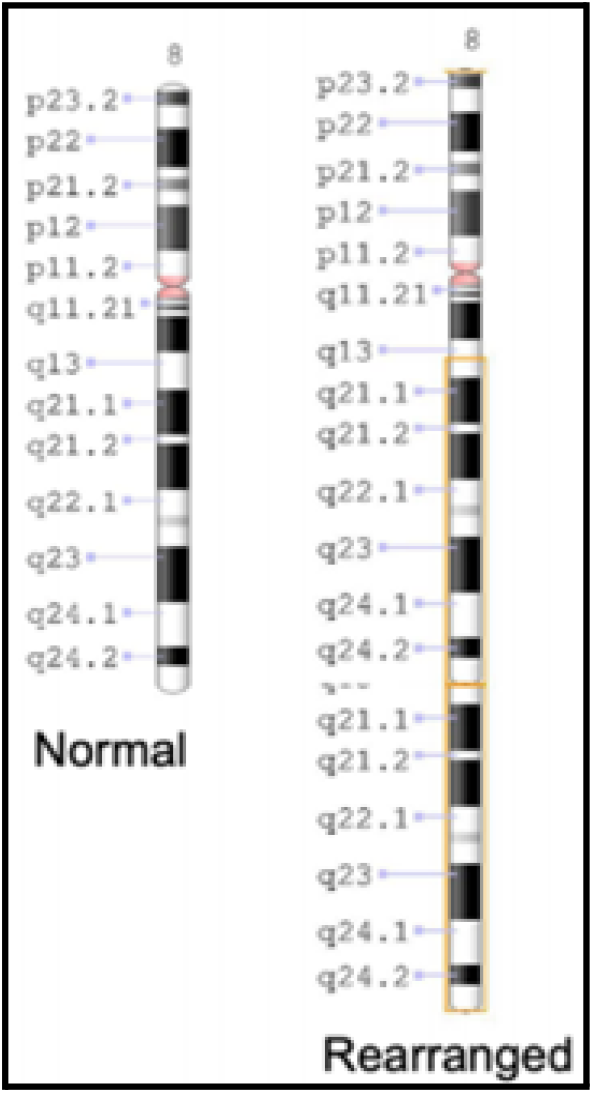
Rearranged chromosome 8. A small region near the 8p telomere is deleted, and the majority of 8q is duplicated (Adapted from Pyle et al., 2018).

**Figure 2:**
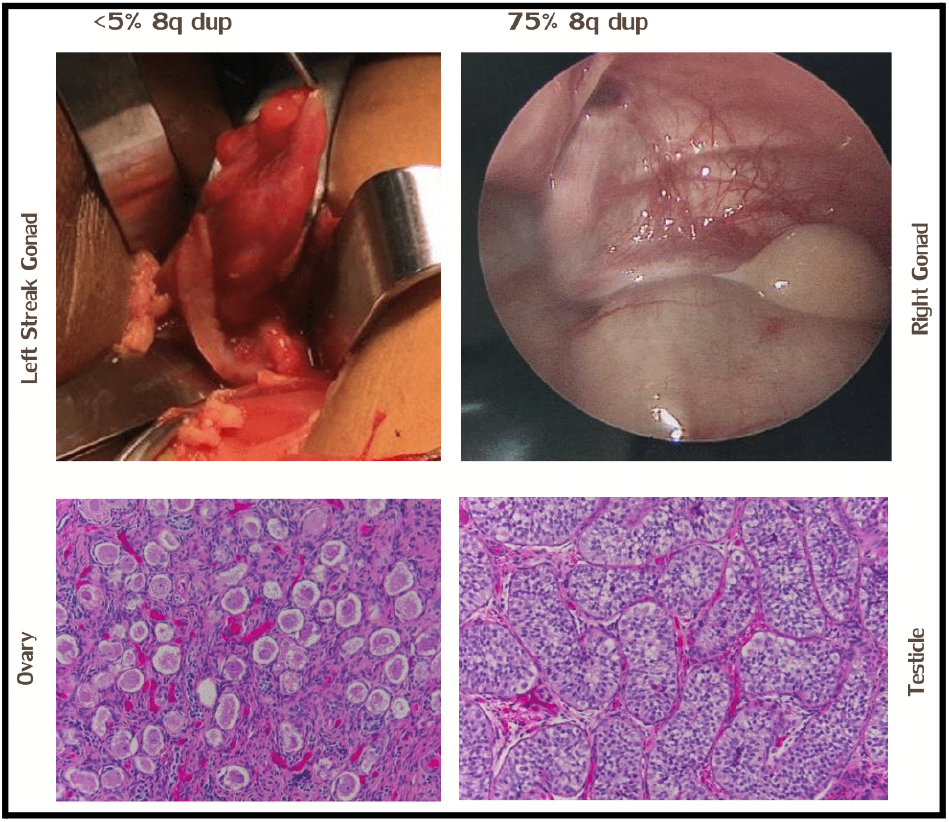
Surgical and histological imagery of left streak gonad and right gonad, with gonadal array revealing that fewer than 5% of the streak ovary cell population contained the chromosome 8 rearrangement, while 75% of the abnormal testicle cell population contained the rearrangement (Adapted from Pyle et al., 2018).

**Figure 3:**
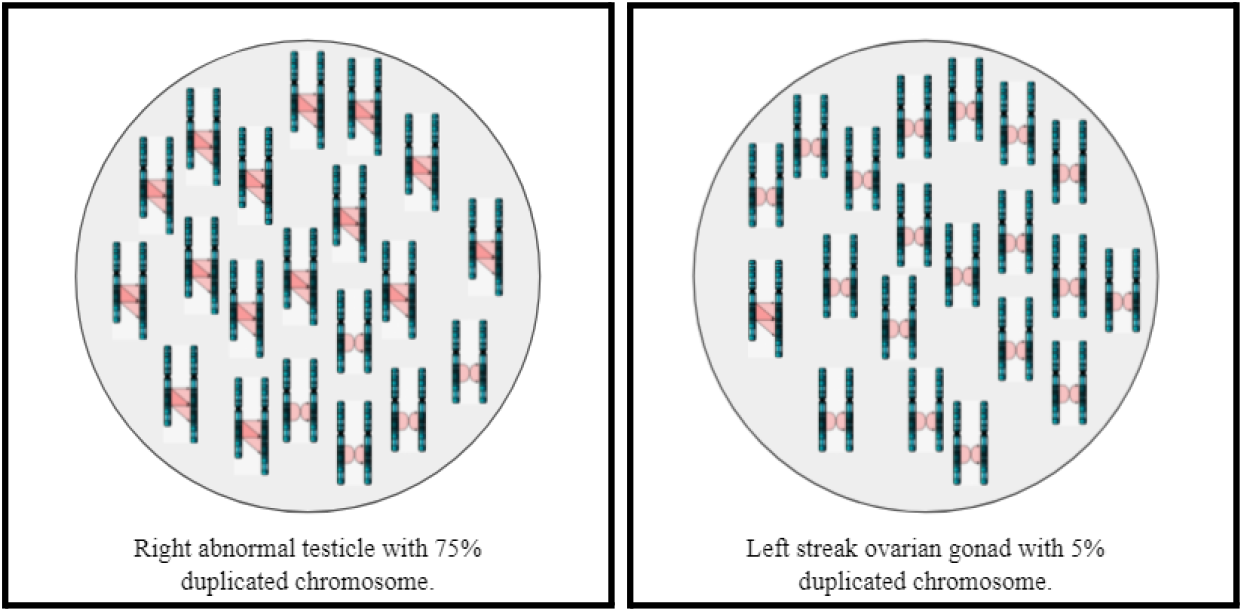
5% of the streak ovary cell population contained the chromosome 8 rearrangement, while 75% of the abnormal testicle cell population contained the rearrangement.

## Methods

We extracted RNA from formalin-fixed paraffin-embedded (FFPE) gonadal tissue samples from the patient of interest and completed RNA-seq of these four FFPE gonadal tissue samples from this mosaic individual to identify differentially expressed genes (DEGs). After quality control, two lines representing with and without the duplication were analyzed. Two samples corresponded to the left streak ovary, and two samples corresponded to the right testicular gonad. We used the Qiagen RNeasy FFPE Kit for purification of total RNA from our FFPE tissue sections. After RNA extraction, sequencing of these four samples and an initial data analysis to identify DEGs and associated statistics were completed.

### RNA-Seq

FastQC was used as a quality control check on raw sequence data from the high-throughput sequencing pipeline. After verifying the quality of the reads, adapter sequences were trimmed from the 3’ ends of reads. Mapping of large sets of high-throughput sequencing reads to a reference genome is one of the foundational steps in RNA-seq data analysis; the Spliced Transcripts Alignment to a Reference (STAR) was used for alignment of the high-throughput RNA-seq data to GRCh38. StringTie, a computational method that applies a network flow algorithm originally developed in optimization theory to assemble mapped reads into transcripts, was used for read assembly using the Gencode v40 GRCH38.p13 annotation file (Pertea et al., 2015). Compared with other leading transcript assembly programs including Cufflinks, IsoLasso, Scripture, and Traph, StringTie has been shown to produce more complete and accurate reconstructions of genes and better estimates of expression levels (Pertea et al., 2015). StringTie generated normalized, multi-mapping-corrected expression levels of known Ensembl genes.

### Differential Expression

Differential expression analysis (DEA) was performed using DESeq2. In comparative high-throughput sequencing assays, a fundamental task is the analysis of count data, such as read counts per gene in RNA-seq, for evidence of systematic changes across experimental conditions. In our case, we aimed to determine if there were systematic differences in gene read counts in FFPE samples representative of the duplication-containing right testicular gonad compared to FFPE samples representative of the patient’s left streak ovary. DESeq2 uses shrinkage estimation for dispersions and fold changes to improve stability and interpretability of estimates to enable a more quantitative analysis focused on the strength, rather than the mere presence, of differential expression (Love, Huber, & Anders, 2014). We used DESeq2’s standard median ratio method to estimate size factors and further normalize gene read counts.

### Sex-Development Gene List

To determine which of the DEGs were relevant to sex-development pathways, we developed a custom set of sex-development genes using available resources, such as KEGG, Reactome, and WikiPathways, as well as standardized literature searches and existing clinical resources, assembling a list of 135 genes expected to play a role in DSDs. The full custom list of sex-development genes is included in *Supplemental File 2*.

### Gene Set Enrichment Analysis (GSEA)

Lastly, we performed Gene Set Enrichment Analysis using a pre-ranked list of sex-development genes whose differential expression values were obtained from DESeq2 analysis of gene counts generated by StringTie analysis of aligned BAM files from each sample. List rankings were based on the metric of -log_10_(*p*-value)*sign(FC) (Zhang & Storey, 2018). The list contained sex-development genes from our custom sex-development gene list that successfully matched to DESeq2 output and had an associated human gene symbol. The final pre-ranked list contained 126 genes. Gene set size filters included a minimum gene set size of 10 and maximum gene set size of 500, resulting in filtering out 31,847 of 32,880 gene sets. The remaining 1,033 gene sets were used in the analysis. The pre-ranked sex-development gene list is included in *Supplemental File 2*.

## Results

The RNA-seq analysis pipeline indicated a total of 12,534 genes as differentially expressed with adjusted *p*-value ≤ 0.05 in the patient’s right testicular gonadal tissue versus the patient’s left streak ovary. Of these genes, 8,684 were protein-coding genes. The full list of DEGs is included in *Supplemental File 1*.

74 of the 135 sex-development genes were present in our list of DEGs with adjusted *p*-value ≤ 0.05. The full list of differentially expressed sex-development genes is included in *Supplemental File 2*. 51 of the 74 sex-development genes with statistically significant differential expression exhibited an absolute fold change (FC) value ≥ 1.5 and are included in the table below (**Table 1**).

**Table 1:** Differentially expressed sex-development genes with adjusted *p*-value ≤ 0.05 and abs(FC) ≥ 1.5.

| Sex-Development Gene Name | Gene description | Chromosome/scaffold name | Gene type | $\log_2\text{FC}$ | FC | $p\text{-adj.}$ |
| --- | --- | --- | --- | --- | --- | --- |
| *CHD4* | chromodomain helicase DNA binding protein 4 | 12 | Protein Coding | 2.48 | 5.57 | 5.69E-147 |
| *SOX9* | SRY-box transcription factor 9 | 17 | Protein Coding | 5.29 | 39.20 | 6.06E-119 |
| *STAR* | steroidogenic acute regulatory protein | 8 | Protein Coding | -1.29 | -2.44 | 1.93E-55 |
| *AMH* | anti-Mullerian hormone | 19 | Protein Coding | 6.76 | 108.42 | 3.00E-54 |
| *HHAT* | hedgehog acyltransferase | 1 | Protein Coding | -2.36 | -5.13 | 2.76E-50 |
| *DHCR24* | 24-dehydrocholesterol reductase | 1 | Protein Coding | 2.09 | 4.27 | 1.73E-46 |
| *BNC2* | basonuclin 2 | 9 | Protein Coding | -0.81 | -1.75 | 8.18E-41 |
| *FOXL2* | forkhead box L2 | 3 | Protein Coding | -3.02 | -8.12 | 2.86E-39 |
| *STAG3* | stromal antigen 3 | 7 | Protein Coding | -2.51 | -5.71 | 2.55E-38 |
| *ZEB2* | zinc finger E-box binding homeobox 2 | 2 | Protein Coding | 0.63 | 1.54 | 1.68E-35 |
| *ESCO2* | establishment of sister chromatid cohesion<br>N-acetyltransferase 2 | 8 | Protein Coding | -2.74 | -6.68 | 3.95E-31 |
| *EVC* | EvC ciliary complex subunit 1 | 4 | Protein Coding | -0.81 | -1.75 | 2.41E-29 |
| *HSD17B3* | hydroxysteroid 17-beta dehydrogenase 3 | 9 | Protein Coding | 5.18 | 36.25 | 7.78E-27 |
| *ANOS1* | anosmin 1 | X | Protein Coding | 2.02 | 4.05 | 1.96E-24 |
| *B3GLCT* | beta 3-glucosyltransferase | 13 | Protein Coding | -1.40 | -2.64 | 1.88E-21 |
| *FREM2* | FRAS1 related extracellular matrix 2 | 13 | Protein Coding | -1.05 | -2.07 | 3.54E-21 |
| *TP63* | tumor protein p63 | 3 | Protein Coding | -3.24 | -9.47 | 5.19E-20 |
| *VAMP7* | vesicle associated membrane protein 7 | X | Protein Coding | 4.53 | 23.11 | 7.65E-19 |
| *PDE4D* | phosphodiesterase 4D | 5 | Protein Coding | 1.72 | 3.30 | 5.78E-18 |
| *MAP3K1* | mitogen-activated protein kinase kinase kinase 1 | 5 | Protein Coding | 0.76 | 1.69 | 5.60E-17 |
| *OPHN1* | oligophrenin 1 | X | Protein Coding | 0.77 | 1.71 | 2.06E-15 |
| *WDR11* | WD repeat domain 11 | 10 | Protein Coding | 0.80 | 1.74 | 2.79E-15 |
| *WNT5A* | Wnt family member 5A | 3 | Protein Coding | -0.68 | -1.60 | 3.67E-15 |
| *HSD17B4* | hydroxysteroid 17-beta dehydrogenase 4 | 5 | Protein Coding | 0.90 | 1.86 | 1.66E-14 |
| *BBS5* | Bardet-Biedl syndrome 5 | 2 | Protein Coding | -1.67 | -3.18 | 3.71E-14 |
| *WWOX* | WW domain containing oxidoreductase | 16 | Protein Coding | -0.89 | -1.85 | 5.14E-14 |
| *SETBP1* | SET binding protein 1 | 18 | Protein Coding | 0.73 | 1.66 | 1.20E-13 |
| *DMRT1* | doublesex and mab-3 related transcription factor 1 | 9 | Protein Coding | 3.80 | 13.91 | 2.14E-12 |
| *LHCGR* | luteinizing hormone/choriogonadotropin receptor | 2 | Protein Coding | 1.96 | 3.88 | 1.50E-11 |
| *TWIST2* | twist family bHLH transcription factor 2 | 2 | Protein Coding | 3.66 | 12.68 | 2.14E-11 |
| *AKR1C2* | aldo-keto reductase family 1 member C2 | 10 | Protein Coding | 3.52 | 11.46 | 2.41E-10 |
| *GNRH1* | gonadotropin releasing hormone 1 | 8 | Protein Coding | 2.65 | 6.28 | 1.60E-09 |
| *HS6ST1* | heparan sulfate 6-O-sulfotransferase 1 | 2 | Protein Coding | 0.86 | 1.82 | 1.90E-09 |
| *BCOR* | BCL6 corepressor | X | Protein Coding | 0.83 | 1.78 | 6.58E-09 |
| *MID1* | midline 1 | X | Protein Coding | 1.11 | 2.16 | 1.48E-08 |
| *LEPR* | leptin receptor | 1 | Protein Coding | -0.69 | -1.61 | 2.92E-08 |
| *RBBP8* | RB binding protein 8, endonuclease | 18 | Protein Coding | -1.25 | -2.38 | 7.07E-08 |
| *MCM9* | minichromosome maintenance 9 homologous<br>recombination repair factor | 6 | Protein Coding | 0.93 | 1.90 | 1.42E-06 |
| *MKKS* | MKKS centrosomal shuttling protein | 20 | Protein Coding | 1.45 | 2.73 | 3.77E-06 |
| *TOE1* | target of EGR1, exonuclease | 1 | Protein Coding | 2.66 | 6.31 | 7.28E-06 |
| *FGFR3* | fibroblast growth factor receptor 3 | 4 | Protein Coding | 1.49 | 2.80 | 8.00E-06 |
| *TMEM70* | transmembrane protein 70 | 8 | Protein Coding | 1.14 | 2.20 | 0.00 |
| *HOXA10* | homeobox A10 | 7 | Protein Coding | 2.11 | 4.33 | 0.00 |
| *PITX2* | paired like homeodomain 2 | 4 | Protein Coding | -2.07 | -4.19 | 0.00 |
| *SALL1* | spalt like transcription factor 1 | 16 | Protein Coding | -0.94 | -1.92 | 0.00 |
| *HFE* | homeostatic iron regulator | 6 | Protein Coding | 0.84 | 1.79 | 0.00 |
| *TRIM32* | tripartite motif containing 32 | 9 | Protein Coding | 0.63 | 1.55 | 0.01 |
| *SCARF2* | scavenger receptor class F member 2 | 22 | Protein Coding | -1.10 | -2.15 | 0.01 |
| *DNMT3B* | DNA methyltransferase 3 beta | 20 | Protein Coding | 0.62 | 1.54 | 0.02 |
| *HOXB6* | homeobox B6 | 17 | Protein Coding | 1.32 | 2.50 | 0.02 |
| *BBS12* | Bardet-Biedl syndrome 12 | 4 | Protein Coding | 0.97 | 1.96 | 0.02 |

In gonadal tissue containing the duplication, increased dosage of *ZFPM2* in a *SRY*-negative 46,XX individual appears to upregulate transcriptional activity of gonadal specific promoters such as *SOX9* and *AMH* via its protein interaction with known regulator of early testis development *GATA4. ZFPM2* is essential to the *GATA4*/*ZFPM2* transcription complex; our results show that increased *ZFPM2* dosage enhances *ZFPM2*’s interaction with *GATA4* and results in upregulation of *SOX9* that is sufficient to initiate testis differentiation independent of *SRY. SOX9* (FC=39.2, *p*=6.1×10^−119^) and *SF-1* (FC=1.4, *p*=1.6×10^−3^) interact to produce the functional marker of fetal Sertoli cells *AMH* (FC=108.4, *p*=3.0×10^−54^) and inhibit female sexual differentiation. Several components of the testicular sex-development pathway were upregulated in addition to *SOX9* and *AMH*, including the pro-testicular transcription factor *MAP3K1* (FC=1.7, *p*=5.6×10^−17^), *DMRT1* (FC=13.9, *p*=2.1×10^−12^), *LHX9* (FC=2.5, *p*=5.0×10^−14^), *DHH* (FC=12.0, *p*=1.8×10^−30^), *PTGDS* (FC=2.5, *p*=3.4×10^−18^), and *SOX8* (FC=10.9, *p*=6.5×10^−10^).

*GATA4* has been shown to bind to a consensus site in the *AMH* promoter and activate expression of an *AMH* reporter construct *in vitro* (Viger et al., 1998). We observed significant upregulation of *AMH* in duplication-containing cells (FC=108.4, *p*=3.0×10^−54^). *AMH* is only expressed in immature Sertoli cells, whereas expression of *GATA4* typically remains activated in adult testes (Rajpert-De Meyts et al., 1999; Steger et al., 1999). However, expression of *AMH* in visibly undifferentiated Sertoli cells is consistent with testicular dysgenesis.

Our results are consistent with a model of the *GATA4*/*ZFPM2* transcription complex circumventing *SRY* to directly or indirectly activate *SOX9* gene expression and trigger a cascade of transcriptional events resulting in significant upregulation of male sex determination pathways along with repression of typical female sex determination pathways (**Figure 4**).

**Figure 4:**
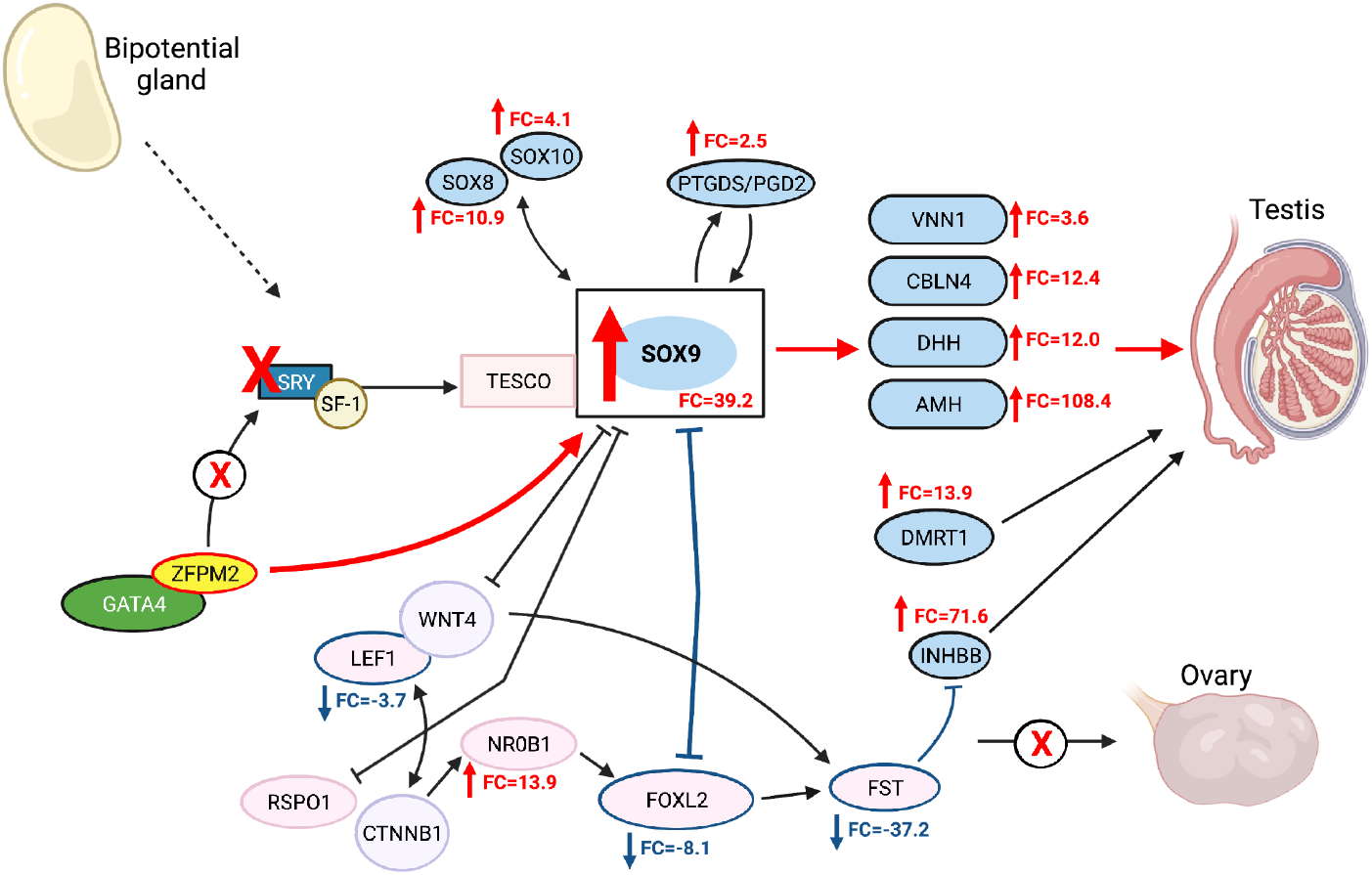
The *GATA4*/*ZFPM2* transcription complex circumvents *SRY* to activate *SOX9* gene expression. Indicated fold changes are statistically significant with adj. *p*-value ≤ 0.05.

### GSEA of sex-development genes

GSEA indicated 498 of 1,033 gene sets upregulated in samples representative of the right testicular gonad. One gene set unrelated to sex development was significantly enriched at false discovery rate (FDR) < 0.25. 26 gene sets were significantly enriched at nominal *p*-value ≤ 0.05, including the following gene sets possibly related to sex development: Human Phenotype (HP) Infertility, Gene Ontology Biological Process (GOBP) Male Sex Differentiation, HP Decreased Fertility in Males, HP Gonosomal Inheritance, and HP Hypogonadatropic Hypogonadism (**Table 2**). The full GSEA report using sex-development genes is included as *Supplemental File 3*.

**Table 2:**
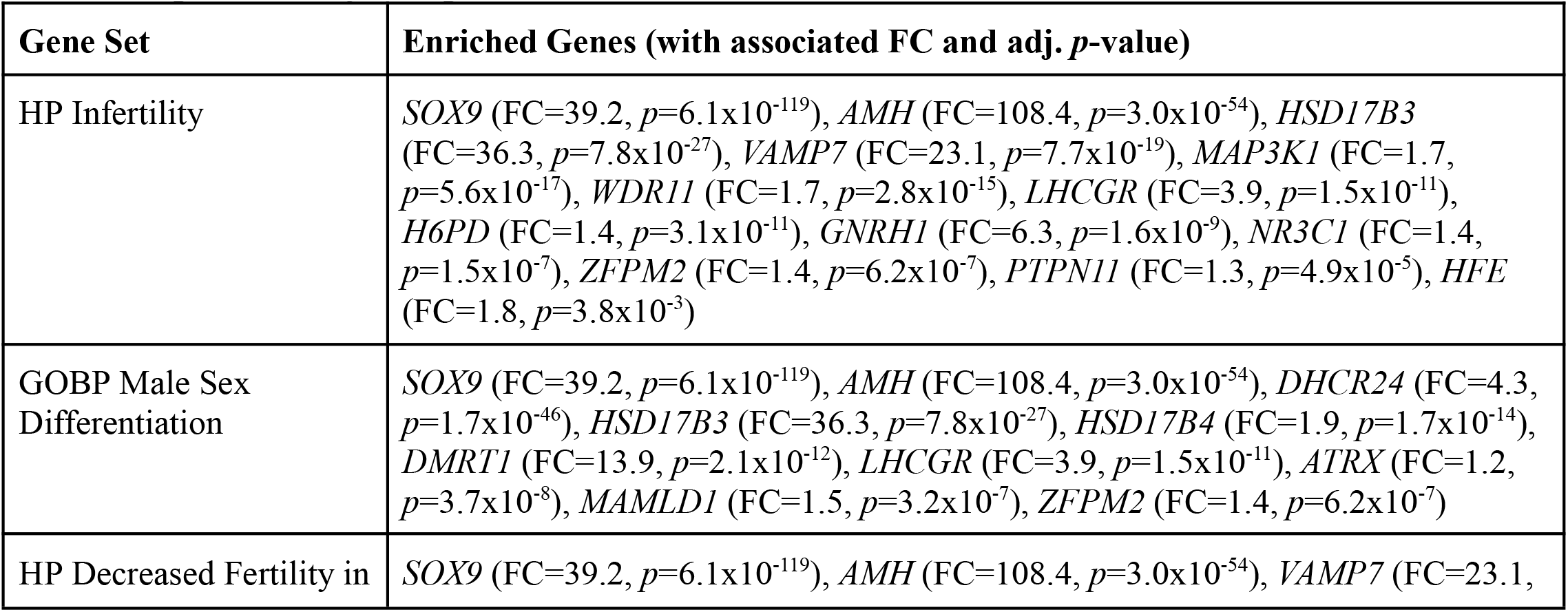
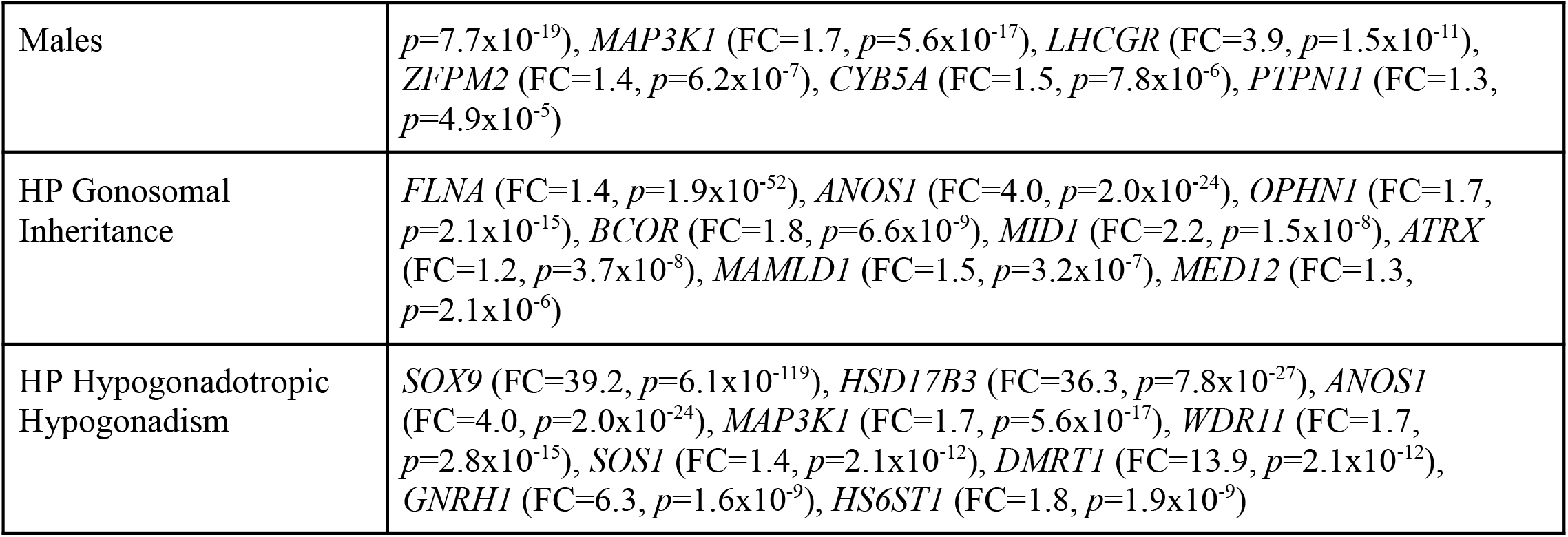
Gene sets related to sex development significantly enriched at nominal *p*-value ≤ 0.05 using sex-development genes in GSEA.

## Discussion

*ZFPM2* may function as a master temporal and spatial regulator of mammalian testicular organogenesis whose increased dosage elicits significant and cascading downstream effects. These effects are primarily exerted via activation of *SOX9*, whose duplication or increased dosage is implicated in cases of *SRY*-negative 46,XX sex reversal in humans, as well as non-human mammals (Huang et al., 1999; Rossi et al., 2014). Ovarian development is an active process involving antagonistic regulatory networks that suppress testicular development.

Increased dosage of *SOX9* appears sufficient to initiate testis differentiation absent *SRY* through its induction of male sex-development pathways and subsequent repression of female sex-development pathways (**Figure 5**). *PTGDS* (prostaglandin D2 synthase), for example, potentially acts as an amplification loop of *SOX9* expression, driving further *AMH* expression and repression of female reproductive development. Further, components of the ovarian *WNT*-signaling pathway were repressed, including *LEF1* (FC=-3.7, *p*=1.4×10^−21^) and *FOXL2* (FC=-8.1, *p*=2.9×10^−39^).

**Figure 5:**
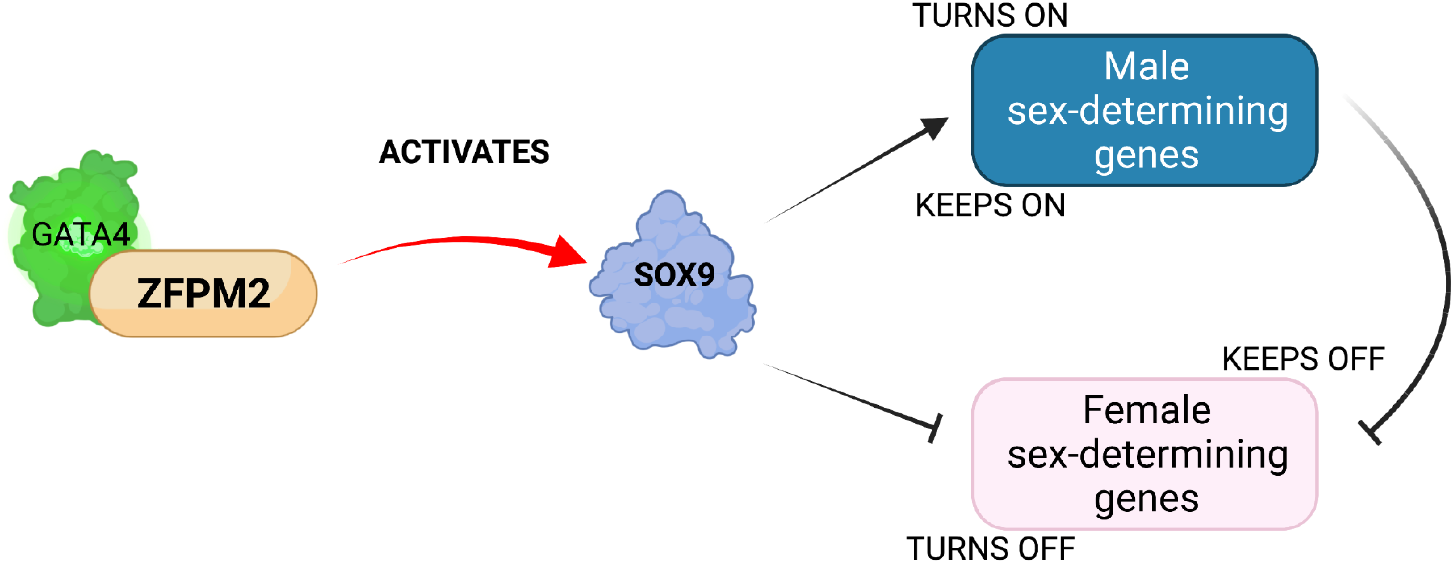
Increased dosage of *ZFPM2* upregulates pro-testicular *SOX9*, setting in motion a *SRY*-negative 46,XX sex reversal pathway. Activation of male sex-development pathways appears to result in concurrent repression of female sex-development pathways.

*SOX9* may be central to *ZFPM2*’s significant effects upon sex development, as *SOX9* has numerous regulatory sites throughout the human genome that may directly or indirectly interact with the *GATA4*/*ZFPM2* transcription complex. *SOX9* enhancer mutations are established as a significant potential cause of human DSD (Croft et al., 2018). In mice, *SRY* typically cooperates with *SF-1* to bind to and activate a *SOX9* enhancer known as Testis Specific Enhancer of *SOX9* (TES) and its core sequence TESCO (Gonen, Quinn, O’Neill, Koopman, & Lovell-Badge, 2017); (Sekido & Lovell-Badge, 2008). TES and its core TESCO bind relevant transcription factors and integrate positive and negative control over *SOX9* expression during gonadal development (Gonen et al., 2017). However, although normal levels of *SOX9* expression in the testis depend on TES/TESCO at both embryonic and adult stages, TES/TESCO deletion in mice does not lead to signs of sex reversal in either TES/TESCO-homozygous-deleted embryos or adults (Gonen et al., 2017). This implies TESCO is not the only enhancer active in *SOX9* expression. We posit the *GATA4*/*ZFPM2* transcription complex affects sex development via interaction with one or a combination of *SOX9* regulatory elements. Regarding TESCO, a combination of wild-type *ZFPM2* and *GATA4* proteins showed no difference in transactivation of TESCO when compared with *GATA4* alone, although the level of activity was significantly reduced in comparison with wild-type *ZFPM2* alone (Bashamboo et al., 2014).

There is thus an increasing need to elucidate the interaction of *GATA4*/*ZFPM2* with additional *SOX9* control elements. Presently, there is evidence for multiple enhancers upstream of *SOX9 (Yao et al., 2015)*. At least four 46,XX *SRY*-negative individuals have presented with testicular development and a duplication involving a genomic region located ∼600 kb upstream of *SOX9* (Benko et al., 2011; Cox, Willatt, Homfray, & Woods, 2011; Kim et al., 2015; Vetro et al., 2011; Xiao, Ji, Xing, Chen, & Tao, 2013). Croft et al. (2017) identified three possible enhancers of *SOX9* that in cell-based reporter assays responded to different combinations of testis-specific regulators—including *SRY, SF-1*, and *SOX9*—and that when duplicated or deleted resulted in sex reversal in *SRY*-negative 46,XX individuals. These enhancers include Sex Reversal Enhancer-A (eSR-A) in the XYSR genomic region (17q24.3), Sex Reversal Enhancer-B (eSR-B) in the RevSex genomic region, and eALDI. eSR-A and eSR-B are of particular interest as they have been found duplicated in *SRY*-negative 46,XX DSD patients and can be activated absent *SRY* (Gonen et al., 2017).

The *GATA4*/*ZFPM2* transcriptional complex may play a crucial role in human sex development and DSD through direct or indirect activation of *SOX9* via one or a combination of *SOX9*’s upstream regulatory elements absent *SRY*. eSR-A was first identified after two 46,XX DSD *SRY*-negative sex reversal patients each presented with a duplication overlapping a 32.5-kb genetic interval in the 2-Mb *SOX9* regulatory region known as XYSR (17q24.3)—the deletion of which has also been linked to 46,XY sex reversal (Gonen et al., 2017; Kim et al., 2015). The 5.2-kb region of overlap has a core gonadal enhancer for *SOX9* implicated in both 46,XX and 46,XY DSD (Gonen et al., 2017). eSR-B was identified after four 46,XX *SRY*-negative sex reversal DSD patients presented with duplications in the upstream regulatory region of *SOX9* known as RevSex (Benko et al., 2011; Hyon et al., 2015; Ohnesorg et al., 2017; Vetro et al., 2011; Xiao et al., 2013). eSR-B is the RevSex fragment showing overlap amongst these patients and with a highly conserved SOX9 binding motif (Gonen et al., 2017). The third putative *SOX9* enhancer element, eALDI, is located 1,259 bp immediately upstream of TESCO; in combination with *SF-1, SRY* strongly activates the eALDI enhancer, unlike eSR-A or eSR-B (Gonen et al., 2017). We posit that enhanced activation due to increased dosage of *ZFPM2* of one or a combination of these possible *SOX9* regulatory elements directly or indirectly by *GATA4*/*ZFPM2* might result in the synergistic gene activation of male sex determination pathways (**Figure 6**).

**Figure 6:**
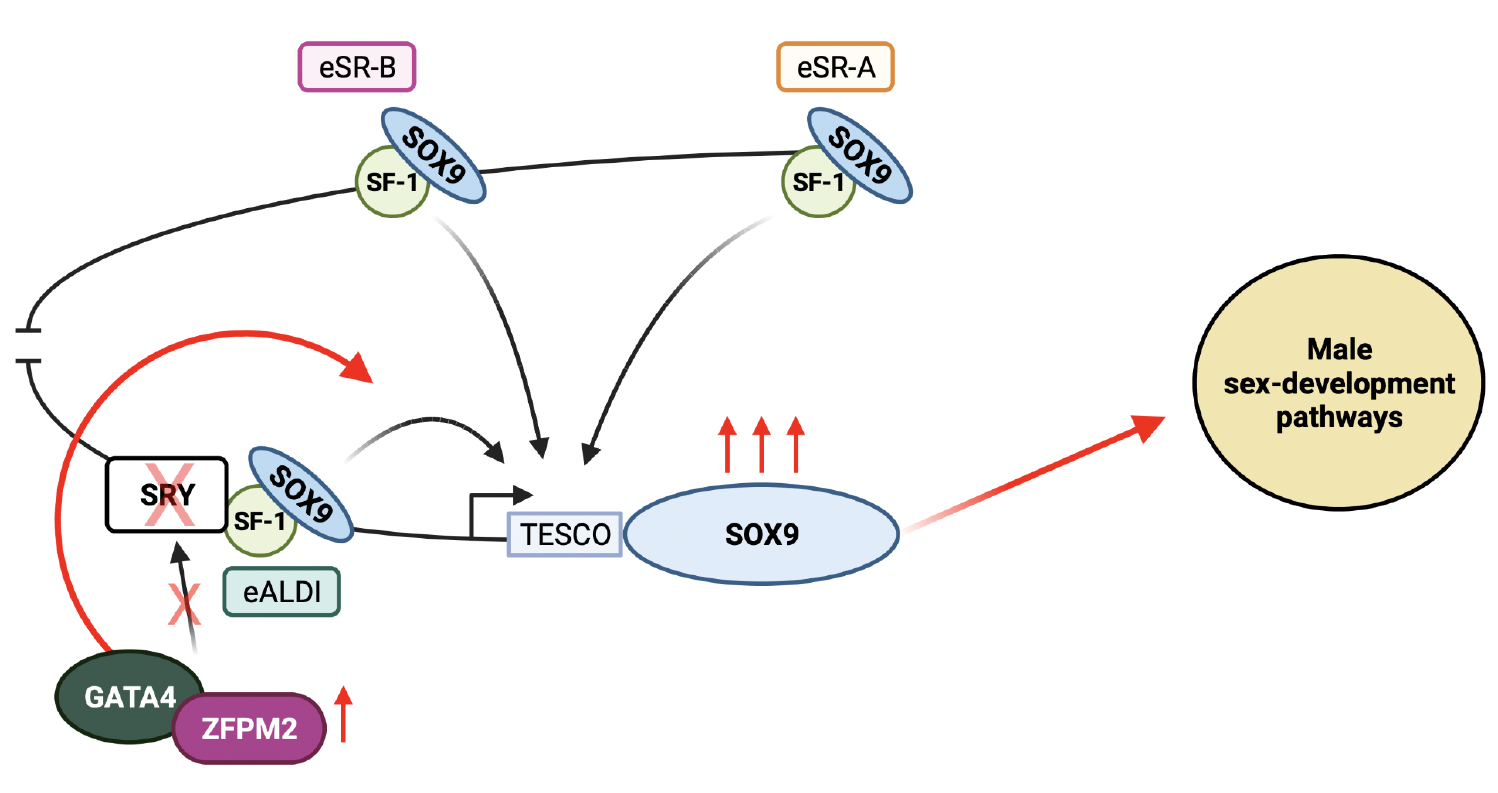
The *GATA4*/*ZFPM2* transcription complex circumvents *SRY* to either directly or indirectly upregulate *SOX9* gene expression.

## Conclusion

We have shown that increased *ZPFM2* dosage can induce 46,XX testicular development in a manner not dependent on *SRY*. This contravenes the previous understanding that *GATA4*/*ZFPM2* drives testicular development through *SRY*. In typical testicular development, *ZFPM2* and its binding partner, *GATA4*, drive expression of the *SRY* master regulator. However, *ZFPM2* may modulate numerous critical sex-development genes including transcription factors otherwise thought to be downstream of *SRY* (*MAP3K1, SOX9, AMH*). Findings from this single high-yield patient demonstrate that the primary role of *ZFPM2* in testicular development may be independent of *SRY*. The *GATA4*/*ZFPM2* transcription complex circumvents *SRY* to either directly or indirectly upregulate *SOX9* gene expression. Ultimately, absent *SRY*, increased *ZFPM2* dosage initiates a *SOX9-*mediated 46,XX sex reversal pathway. The downstream effects of *GATA4*/*ZFPM2*-mediated *SOX9* upregulation are significant, resulting in the upregulation of male sex-determination pathways with interrelated loss in ovarian transcription factor function and dosage. This adds *ZFPM2* to the brief (<10) list of genes capable of directing testicular development in the 46,XX context, absent *SRY*. Overall, new understanding of these genes demonstrates that the role of *SRY* as a “master regulator” of testicular development may be less than previously thought.

## Data Availability

All data produced in the present study are available upon reasonable request to the authors.

## Author Contributions

L.K.R., L.C.P., and D.Y. conceived the study. L.K.R. wrote the manuscript and developed all original graphics. L.K.R. performed the FFPE RNA-Seq preparation, DEA, and GSEA.

## Notes

### Competing Interest Statement

The authors have declared no competing interest.

### Funding Statement

This study did not receive any funding.

### Author Declarations

Ethics committee/IRB of Children's Hospital of Philadelphia gave ethical approval for this work.

