## Supplemental File 3 for "Increased Expression of *ZFPM2* Bypasses *SRY* to Drive 46,XX Testicular Development: A New Mechanism of 46,XX DSD": BUSSLINGER_GASTRIC_IMMUNE_CELLS.html

Details for gene set BUSSLINGER\_GASTRIC\_IMMUNE\_CELLS[GSEA]

|  || Dataset | SexDevelopmentGenesPRL\_remapped |
| Phenotype | NoPhenotypeAvailable |
| Upregulated in class | na\_pos |
| GeneSet | BUSSLINGER\_GASTRIC\_IMMUNE\_CELLS |
| Enrichment Score (ES) | 0.54861665 |
| Normalized Enrichment Score (NES) | 2.1389322 |
| Nominal p-value | 0.002057613 |
| FDR q-value | 0.5831791 |
| FWER p-Value | 0.711 |
Table: GSEA Results Summary

  

Fig 1: Enrichment plot: BUSSLINGER\_GASTRIC\_IMMUNE\_CELLS      
 Profile of the Running ES Score & Positions of GeneSet Members on the Rank Ordered List

  

| SYMBOL | TITLE | RANK IN GENE LIST | RANK METRIC SCORE | RUNNING ES | CORE ENRICHMENT || 1 | FLNA | filamin A [Source:HGNC Symbol;Acc:HGNC:3754] | 3 | 53.800 | 0.0648 | Yes |
| 2 | ZEB2 | zinc finger E-box binding homeobox 2 [Source:HGNC Symbol;Acc:HGNC:14881] | 5 | 36.500 | 0.1470 | Yes |
| 3 | SOS1 | SOS Ras/Rac guanine nucleotide exchange factor 1 [Source:HGNC Symbol;Acc:HGNC:11187] | 15 | 12.800 | 0.1597 | Yes |
| 4 | CREBBP | CREB binding protein [Source:HGNC Symbol;Acc:HGNC:2348] | 20 | 11.800 | 0.2158 | Yes |
| 5 | ATRX | ATRX chromatin remodeler [Source:HGNC Symbol;Acc:HGNC:886] | 27 | 8.350 | 0.2545 | Yes |
| 6 | KAT6B | lysine acetyltransferase 6B [Source:HGNC Symbol;Acc:HGNC:17582] | 28 | 8.070 | 0.3455 | Yes |
| 7 | NR3C1 | nuclear receptor subfamily 3 group C member 1 [Source:HGNC Symbol;Acc:HGNC:7978] | 29 | 7.690 | 0.4364 | Yes |
| 8 | UBR1 | ubiquitin protein ligase E3 component n-recognin 1 [Source:HGNC Symbol;Acc:HGNC:16808] | 38 | 5.250 | 0.4577 | Yes |
| 9 | PTPN11 | protein tyrosine phosphatase non-receptor type 11 [Source:HGNC Symbol;Acc:HGNC:9644] | 39 | 4.970 | 0.5486 | Yes |
| 10 | PTDSS1 | phosphatidylserine synthase 1 [Source:HGNC Symbol;Acc:HGNC:9587] | 71 | 0.100 | 0.3700 | No |
| 11 | EPG5 | ectopic P-granules autophagy protein 5 homolog [Source:HGNC Symbol;Acc:HGNC:29331] | 72 | 0.100 | 0.4609 | No |
Table: GSEA details [plain text format]

  

Fig 2: BUSSLINGER\_GASTRIC\_IMMUNE\_CELLS: Random ES distribution      
 Gene set null distribution of ES for **BUSSLINGER\_GASTRIC\_IMMUNE\_CELLS**

  
