## Supplemental File 3 for "Increased Expression of *ZFPM2* Bypasses *SRY* to Drive 46,XX Testicular Development: A New Mechanism of 46,XX DSD": DACOSTA_UV_RESPONSE_VIA_ERCC3_DN.html

Details for gene set DACOSTA\_UV\_RESPONSE\_VIA\_ERCC3\_DN[GSEA]

|  || Dataset | SexDevelopmentGenesPRL\_remapped |
| Phenotype | NoPhenotypeAvailable |
| Upregulated in class | na\_pos |
| GeneSet | DACOSTA\_UV\_RESPONSE\_VIA\_ERCC3\_DN |
| Enrichment Score (ES) | 0.5833901 |
| Normalized Enrichment Score (NES) | 2.4136128 |
| Nominal p-value | 0.0 |
| FDR q-value | 0.17965767 |
| FWER p-Value | 0.179 |
Table: GSEA Results Summary

  

Fig 1: Enrichment plot: DACOSTA\_UV\_RESPONSE\_VIA\_ERCC3\_DN      
 Profile of the Running ES Score & Positions of GeneSet Members on the Rank Ordered List

  

| SYMBOL | TITLE | RANK IN GENE LIST | RANK METRIC SCORE | RUNNING ES | CORE ENRICHMENT || 1 | SOX9 | SRY-box transcription factor 9 [Source:HGNC Symbol;Acc:HGNC:11204] | 1 | 121.000 | 0.0681 | Yes |
| 2 | ZEB2 | zinc finger E-box binding homeobox 2 [Source:HGNC Symbol;Acc:HGNC:14881] | 5 | 36.500 | 0.1184 | Yes |
| 3 | SOS1 | SOS Ras/Rac guanine nucleotide exchange factor 1 [Source:HGNC Symbol;Acc:HGNC:11187] | 15 | 12.800 | 0.1157 | Yes |
| 4 | PCNT | pericentrin [Source:HGNC Symbol;Acc:HGNC:16068] | 17 | 12.400 | 0.1838 | Yes |
| 5 | CREBBP | CREB binding protein [Source:HGNC Symbol;Acc:HGNC:2348] | 20 | 11.800 | 0.2430 | Yes |
| 6 | MID1 | midline 1 [Source:HGNC Symbol;Acc:HGNC:7095] | 26 | 8.770 | 0.2757 | Yes |
| 7 | ATRX | ATRX chromatin remodeler [Source:HGNC Symbol;Acc:HGNC:886] | 27 | 8.350 | 0.3526 | Yes |
| 8 | KAT6B | lysine acetyltransferase 6B [Source:HGNC Symbol;Acc:HGNC:17582] | 28 | 8.070 | 0.4295 | Yes |
| 9 | NR3C1 | nuclear receptor subfamily 3 group C member 1 [Source:HGNC Symbol;Acc:HGNC:7978] | 29 | 7.690 | 0.5065 | Yes |
| 10 | MAMLD1 | mastermind like domain containing 1 [Source:HGNC Symbol;Acc:HGNC:2568] | 30 | 7.350 | 0.5834 | Yes |
| 11 | NEK1 | NIMA related kinase 1 [Source:HGNC Symbol;Acc:HGNC:7744] | 75 | -0.090 | 0.2709 | No |
| 12 | SPECC1L | sperm antigen with calponin homology and coiled-coil domains 1 like [Source:HGNC Symbol;Acc:HGNC:29022] | 109 | -4.400 | 0.0558 | No |
| 13 | WNT5A | Wnt family member 5A [Source:HGNC Symbol;Acc:HGNC:12784] | 115 | -15.700 | 0.0885 | No |
Table: GSEA details [plain text format]

  

Fig 2: DACOSTA\_UV\_RESPONSE\_VIA\_ERCC3\_DN: Random ES distribution      
 Gene set null distribution of ES for **DACOSTA\_UV\_RESPONSE\_VIA\_ERCC3\_DN**

  
