## Supplemental File 3 for "Increased Expression of *ZFPM2* Bypasses *SRY* to Drive 46,XX Testicular Development: A New Mechanism of 46,XX DSD": GOBP_EMBRYONIC_APPENDAGE_MORPHOGENESIS.html

Details for gene set GOBP\_EMBRYONIC\_APPENDAGE\_MORPHOGENESIS[GSEA]

|  || Dataset | SexDevelopmentGenesPRL\_remapped |
| Phenotype | NoPhenotypeAvailable |
| Upregulated in class | na\_neg |
| GeneSet | GOBP\_EMBRYONIC\_APPENDAGE\_MORPHOGENESIS |
| Enrichment Score (ES) | -0.4107143 |
| Normalized Enrichment Score (NES) | -1.7315018 |
| Nominal p-value | 0.022177419 |
| FDR q-value | 0.5236232 |
| FWER p-Value | 1.0 |
Table: GSEA Results Summary

  

Fig 1: Enrichment plot: GOBP\_EMBRYONIC\_APPENDAGE\_MORPHOGENESIS      
 Profile of the Running ES Score & Positions of GeneSet Members on the Rank Ordered List

  

| SYMBOL | TITLE | RANK IN GENE LIST | RANK METRIC SCORE | RUNNING ES | CORE ENRICHMENT || 1 | CREBBP | CREB binding protein [Source:HGNC Symbol;Acc:HGNC:2348] | 20 | 11.800 | -0.1071 | No |
| 2 | HOXA10 | homeobox A10 [Source:HGNC Symbol;Acc:HGNC:5100] | 41 | 3.830 | -0.2143 | No |
| 3 | BMP4 | bone morphogenetic protein 4 [Source:HGNC Symbol;Acc:HGNC:1071] | 61 | 0.750 | -0.3125 | No |
| 4 | FRAS1 | Fraser extracellular matrix complex subunit 1 [Source:HGNC Symbol;Acc:HGNC:19185] | 73 | 0.010 | -0.3393 | Yes |
| 5 | DYNC2H1 | dynein cytoplasmic 2 heavy chain 1 [Source:HGNC Symbol;Acc:HGNC:2962] | 79 | -0.230 | -0.3125 | Yes |
| 6 | MKS1 | MKS transition zone complex subunit 1 [Source:HGNC Symbol;Acc:HGNC:7121] | 88 | -0.480 | -0.3125 | Yes |
| 7 | ROR2 | receptor tyrosine kinase like orphan receptor 2 [Source:HGNC Symbol;Acc:HGNC:10257] | 91 | -0.580 | -0.2589 | Yes |
| 8 | GLI3 | GLI family zinc finger 3 [Source:HGNC Symbol;Acc:HGNC:4319] | 104 | -2.920 | -0.2946 | Yes |
| 9 | CHD7 | chromodomain helicase DNA binding protein 7 [Source:HGNC Symbol;Acc:HGNC:20626] | 105 | -3.230 | -0.2232 | Yes |
| 10 | SALL1 | spalt like transcription factor 1 [Source:HGNC Symbol;Acc:HGNC:10524] | 106 | -3.630 | -0.1518 | Yes |
| 11 | PBX1 | PBX homeobox 1 [Source:HGNC Symbol;Acc:HGNC:8632] | 112 | -12.600 | -0.1250 | Yes |
| 12 | WNT5A | Wnt family member 5A [Source:HGNC Symbol;Acc:HGNC:12784] | 115 | -15.700 | -0.0714 | Yes |
| 13 | TP63 | tumor protein p63 [Source:HGNC Symbol;Acc:HGNC:15979] | 116 | -20.700 | 0.0000 | Yes |
| 14 | FREM2 | FRAS1 related extracellular matrix 2 [Source:HGNC Symbol;Acc:HGNC:25396] | 117 | -21.900 | 0.0714 | Yes |
Table: GSEA details [plain text format]

  

Fig 2: GOBP\_EMBRYONIC\_APPENDAGE\_MORPHOGENESIS: Random ES distribution      
 Gene set null distribution of ES for **GOBP\_EMBRYONIC\_APPENDAGE\_MORPHOGENESIS**

  
