## Supplemental File 3 for "Increased Expression of *ZFPM2* Bypasses *SRY* to Drive 46,XX Testicular Development: A New Mechanism of 46,XX DSD": GOBP_IMMUNE_RESPONSE.html

Details for gene set GOBP\_IMMUNE\_RESPONSE[GSEA]

|  || Dataset | SexDevelopmentGenesPRL\_remapped |
| Phenotype | NoPhenotypeAvailable |
| Upregulated in class | na\_pos |
| GeneSet | GOBP\_IMMUNE\_RESPONSE |
| Enrichment Score (ES) | 0.4810345 |
| Normalized Enrichment Score (NES) | 1.7779622 |
| Nominal p-value | 0.028282829 |
| FDR q-value | 0.88046753 |
| FWER p-Value | 1.0 |
Table: GSEA Results Summary

  

Fig 1: Enrichment plot: GOBP\_IMMUNE\_RESPONSE      
 Profile of the Running ES Score & Positions of GeneSet Members on the Rank Ordered List

  

| SYMBOL | TITLE | RANK IN GENE LIST | RANK METRIC SCORE | RUNNING ES | CORE ENRICHMENT || 1 | VAMP7 | vesicle associated membrane protein 7 [Source:HGNC Symbol;Acc:HGNC:11486] | 8 | 19.500 | 0.0310 | Yes |
| 2 | PDE4D | phosphodiesterase 4D [Source:HGNC Symbol;Acc:HGNC:8783] | 9 | 18.600 | 0.1310 | Yes |
| 3 | MAP3K1 | mitogen-activated protein kinase kinase kinase 1 [Source:HGNC Symbol;Acc:HGNC:6848] | 10 | 17.600 | 0.2310 | Yes |
| 4 | SOS1 | SOS Ras/Rac guanine nucleotide exchange factor 1 [Source:HGNC Symbol;Acc:HGNC:11187] | 15 | 12.800 | 0.2966 | Yes |
| 5 | CREBBP | CREB binding protein [Source:HGNC Symbol;Acc:HGNC:2348] | 20 | 11.800 | 0.3621 | Yes |
| 6 | PTPN11 | protein tyrosine phosphatase non-receptor type 11 [Source:HGNC Symbol;Acc:HGNC:9644] | 39 | 4.970 | 0.3069 | Yes |
| 7 | HFE | homeostatic iron regulator [Source:HGNC Symbol;Acc:HGNC:4886] | 43 | 2.890 | 0.3810 | Yes |
| 8 | TRIM32 | tripartite motif containing 32 [Source:HGNC Symbol;Acc:HGNC:16380] | 44 | 2.410 | 0.4810 | Yes |
| 9 | EPG5 | ectopic P-granules autophagy protein 5 homolog [Source:HGNC Symbol;Acc:HGNC:29331] | 72 | 0.100 | 0.3483 | No |
| 10 | WNT5A | Wnt family member 5A [Source:HGNC Symbol;Acc:HGNC:12784] | 115 | -15.700 | 0.0862 | No |
Table: GSEA details [plain text format]

  

Fig 2: GOBP\_IMMUNE\_RESPONSE: Random ES distribution      
 Gene set null distribution of ES for **GOBP\_IMMUNE\_RESPONSE**

  
