## Supplemental File 3 for "Increased Expression of *ZFPM2* Bypasses *SRY* to Drive 46,XX Testicular Development: A New Mechanism of 46,XX DSD": GOBP_MALE_SEX_DIFFERENTIATION.html

Details for gene set GOBP\_MALE\_SEX\_DIFFERENTIATION[GSEA]

|  || Dataset | SexDevelopmentGenesPRL\_remapped |
| Phenotype | NoPhenotypeAvailable |
| Upregulated in class | na\_pos |
| GeneSet | GOBP\_MALE\_SEX\_DIFFERENTIATION |
| Enrichment Score (ES) | 0.38640043 |
| Normalized Enrichment Score (NES) | 1.8340257 |
| Nominal p-value | 0.01010101 |
| FDR q-value | 1.0 |
| FWER p-Value | 0.999 |
Table: GSEA Results Summary

  

Fig 1: Enrichment plot: GOBP\_MALE\_SEX\_DIFFERENTIATION      
 Profile of the Running ES Score & Positions of GeneSet Members on the Rank Ordered List

  

| SYMBOL | TITLE | RANK IN GENE LIST | RANK METRIC SCORE | RUNNING ES | CORE ENRICHMENT || 1 | SOX9 | SRY-box transcription factor 9 [Source:HGNC Symbol;Acc:HGNC:11204] | 1 | 121.000 | 0.0496 | Yes |
| 2 | AMH | anti-Mullerian hormone [Source:HGNC Symbol;Acc:HGNC:464] | 2 | 55.600 | 0.1085 | Yes |
| 3 | DHCR24 | 24-dehydrocholesterol reductase [Source:HGNC Symbol;Acc:HGNC:2859] | 4 | 47.700 | 0.1581 | Yes |
| 4 | HSD17B3 | hydroxysteroid 17-beta dehydrogenase 3 [Source:HGNC Symbol;Acc:HGNC:5212] | 6 | 27.700 | 0.2078 | Yes |
| 5 | HSD17B4 | hydroxysteroid 17-beta dehydrogenase 4 [Source:HGNC Symbol;Acc:HGNC:5213] | 13 | 15.000 | 0.2115 | Yes |
| 6 | DMRT1 | doublesex and mab-3 related transcription factor 1 [Source:HGNC Symbol;Acc:HGNC:2934] | 16 | 12.800 | 0.2520 | Yes |
| 7 | LHCGR | luteinizing hormone/choriogonadotropin receptor [Source:HGNC Symbol;Acc:HGNC:6585] | 18 | 11.900 | 0.3017 | Yes |
| 8 | ATRX | ATRX chromatin remodeler [Source:HGNC Symbol;Acc:HGNC:886] | 27 | 8.350 | 0.2871 | Yes |
| 9 | MAMLD1 | mastermind like domain containing 1 [Source:HGNC Symbol;Acc:HGNC:2568] | 30 | 7.350 | 0.3276 | Yes |
| 10 | ZFPM2 | "zinc finger protein, FOG family member 2 [Source:HGNC Symbol;Acc:HGNC:16700]" | 31 | 7.040 | 0.3864 | Yes |
| 11 | HOXA10 | homeobox A10 [Source:HGNC Symbol;Acc:HGNC:5100] | 41 | 3.830 | 0.3627 | No |
| 12 | FGF10 | fibroblast growth factor 10 [Source:HGNC Symbol;Acc:HGNC:3666] | 59 | 0.890 | 0.2655 | No |
| 13 | AR | androgen receptor [Source:HGNC Symbol;Acc:HGNC:644] | 63 | 0.490 | 0.2968 | No |
| 14 | SEMA3A | semaphorin 3A [Source:HGNC Symbol;Acc:HGNC:10723] | 67 | 0.310 | 0.3281 | No |
| 15 | WNT4 | Wnt family member 4 [Source:HGNC Symbol;Acc:HGNC:12783] | 82 | -0.310 | 0.2585 | No |
| 16 | ROR2 | receptor tyrosine kinase like orphan receptor 2 [Source:HGNC Symbol;Acc:HGNC:10257] | 91 | -0.580 | 0.2439 | No |
| 17 | WNT5A | Wnt family member 5A [Source:HGNC Symbol;Acc:HGNC:12784] | 115 | -15.700 | 0.0917 | No |
Table: GSEA details [plain text format]

  

Fig 2: GOBP\_MALE\_SEX\_DIFFERENTIATION: Random ES distribution      
 Gene set null distribution of ES for **GOBP\_MALE\_SEX\_DIFFERENTIATION**

  
