## Supplemental File 3 for "Increased Expression of *ZFPM2* Bypasses *SRY* to Drive 46,XX Testicular Development: A New Mechanism of 46,XX DSD": GOBP_MORPHOGENESIS_OF_A_BRANCHING_STRUCTURE.html

Details for gene set GOBP\_MORPHOGENESIS\_OF\_A\_BRANCHING\_STRUCTURE[GSEA]

|  || Dataset | SexDevelopmentGenesPRL\_remapped |
| Phenotype | NoPhenotypeAvailable |
| Upregulated in class | na\_neg |
| GeneSet | GOBP\_MORPHOGENESIS\_OF\_A\_BRANCHING\_STRUCTURE |
| Enrichment Score (ES) | -0.43635124 |
| Normalized Enrichment Score (NES) | -1.8257024 |
| Nominal p-value | 0.012219959 |
| FDR q-value | 0.49980047 |
| FWER p-Value | 1.0 |
Table: GSEA Results Summary

  

Fig 1: Enrichment plot: GOBP\_MORPHOGENESIS\_OF\_A\_BRANCHING\_STRUCTURE      
 Profile of the Running ES Score & Positions of GeneSet Members on the Rank Ordered List

  

| SYMBOL | TITLE | RANK IN GENE LIST | RANK METRIC SCORE | RUNNING ES | CORE ENRICHMENT || 1 | SOX9 | SRY-box transcription factor 9 [Source:HGNC Symbol;Acc:HGNC:11204] | 1 | 121.000 | 0.0681 | No |
| 2 | FGF10 | fibroblast growth factor 10 [Source:HGNC Symbol;Acc:HGNC:3666] | 59 | 0.890 | -0.3594 | Yes |
| 3 | FGFR2 | fibroblast growth factor receptor 2 [Source:HGNC Symbol;Acc:HGNC:3689] | 60 | 0.760 | -0.2825 | Yes |
| 4 | BMP4 | bone morphogenetic protein 4 [Source:HGNC Symbol;Acc:HGNC:1071] | 61 | 0.750 | -0.2056 | Yes |
| 5 | AR | androgen receptor [Source:HGNC Symbol;Acc:HGNC:644] | 63 | 0.490 | -0.1375 | Yes |
| 6 | SEMA3A | semaphorin 3A [Source:HGNC Symbol;Acc:HGNC:10723] | 67 | 0.310 | -0.0871 | Yes |
| 7 | WNT4 | Wnt family member 4 [Source:HGNC Symbol;Acc:HGNC:12783] | 82 | -0.310 | -0.1341 | Yes |
| 8 | MKS1 | MKS transition zone complex subunit 1 [Source:HGNC Symbol;Acc:HGNC:7121] | 88 | -0.480 | -0.1014 | Yes |
| 9 | GLI3 | GLI family zinc finger 3 [Source:HGNC Symbol;Acc:HGNC:4319] | 104 | -2.920 | -0.1572 | Yes |
| 10 | SALL1 | spalt like transcription factor 1 [Source:HGNC Symbol;Acc:HGNC:10524] | 106 | -3.630 | -0.0892 | Yes |
| 11 | PBX1 | PBX homeobox 1 [Source:HGNC Symbol;Acc:HGNC:8632] | 112 | -12.600 | -0.0565 | Yes |
| 12 | WNT5A | Wnt family member 5A [Source:HGNC Symbol;Acc:HGNC:12784] | 115 | -15.700 | 0.0027 | Yes |
| 13 | TP63 | tumor protein p63 [Source:HGNC Symbol;Acc:HGNC:15979] | 116 | -20.700 | 0.0796 | Yes |
Table: GSEA details [plain text format]

  

Fig 2: GOBP\_MORPHOGENESIS\_OF\_A\_BRANCHING\_STRUCTURE: Random ES distribution      
 Gene set null distribution of ES for **GOBP\_MORPHOGENESIS\_OF\_A\_BRANCHING\_STRUCTURE**

  
