## Supplemental File 3 for "Increased Expression of *ZFPM2* Bypasses *SRY* to Drive 46,XX Testicular Development: A New Mechanism of 46,XX DSD": GOBP_POSITIVE_REGULATION_OF_GENE_EXPRESSION.html

Details for gene set GOBP\_POSITIVE\_REGULATION\_OF\_GENE\_EXPRESSION[GSEA]

|  || Dataset | SexDevelopmentGenesPRL\_remapped |
| Phenotype | NoPhenotypeAvailable |
| Upregulated in class | na\_pos |
| GeneSet | GOBP\_POSITIVE\_REGULATION\_OF\_GENE\_EXPRESSION |
| Enrichment Score (ES) | 0.40095305 |
| Normalized Enrichment Score (NES) | 1.6683867 |
| Nominal p-value | 0.033663366 |
| FDR q-value | 0.8374539 |
| FWER p-Value | 1.0 |
Table: GSEA Results Summary

  

Fig 1: Enrichment plot: GOBP\_POSITIVE\_REGULATION\_OF\_GENE\_EXPRESSION      
 Profile of the Running ES Score & Positions of GeneSet Members on the Rank Ordered List

  

| SYMBOL | TITLE | RANK IN GENE LIST | RANK METRIC SCORE | RUNNING ES | CORE ENRICHMENT || 1 | SOX9 | SRY-box transcription factor 9 [Source:HGNC Symbol;Acc:HGNC:11204] | 1 | 121.000 | 0.0681 | Yes |
| 2 | AMH | anti-Mullerian hormone [Source:HGNC Symbol;Acc:HGNC:464] | 2 | 55.600 | 0.1450 | Yes |
| 3 | PDE4D | phosphodiesterase 4D [Source:HGNC Symbol;Acc:HGNC:8783] | 9 | 18.600 | 0.1688 | Yes |
| 4 | PTPN11 | protein tyrosine phosphatase non-receptor type 11 [Source:HGNC Symbol;Acc:HGNC:9644] | 39 | 4.970 | -0.0109 | Yes |
| 5 | HFE | homeostatic iron regulator [Source:HGNC Symbol;Acc:HGNC:4886] | 43 | 2.890 | 0.0395 | Yes |
| 6 | TRIM32 | tripartite motif containing 32 [Source:HGNC Symbol;Acc:HGNC:16380] | 44 | 2.410 | 0.1164 | Yes |
| 7 | DNMT3B | DNA methyltransferase 3 beta [Source:HGNC Symbol;Acc:HGNC:2979] | 45 | 2.110 | 0.1933 | Yes |
| 8 | LMNA | lamin A/C [Source:HGNC Symbol;Acc:HGNC:6636] | 52 | 1.560 | 0.2172 | Yes |
| 9 | FGF10 | fibroblast growth factor 10 [Source:HGNC Symbol;Acc:HGNC:3666] | 59 | 0.890 | 0.2410 | Yes |
| 10 | BMP4 | bone morphogenetic protein 4 [Source:HGNC Symbol;Acc:HGNC:1071] | 61 | 0.750 | 0.3091 | Yes |
| 11 | AR | androgen receptor [Source:HGNC Symbol;Acc:HGNC:644] | 63 | 0.490 | 0.3771 | Yes |
| 12 | ATF3 | activating transcription factor 3 [Source:HGNC Symbol;Acc:HGNC:785] | 70 | 0.100 | 0.4010 | Yes |
| 13 | WNT5A | Wnt family member 5A [Source:HGNC Symbol;Acc:HGNC:12784] | 115 | -15.700 | 0.0885 | No |
Table: GSEA details [plain text format]

  

Fig 2: GOBP\_POSITIVE\_REGULATION\_OF\_GENE\_EXPRESSION: Random ES distribution      
 Gene set null distribution of ES for **GOBP\_POSITIVE\_REGULATION\_OF\_GENE\_EXPRESSION**

  
