## Supplemental File 3 for "Increased Expression of *ZFPM2* Bypasses *SRY* to Drive 46,XX Testicular Development: A New Mechanism of 46,XX DSD": GOBP_RESPONSE_TO_OXYGEN_CONTAINING_COMPOUND.html

Details for gene set GOBP\_RESPONSE\_TO\_OXYGEN\_CONTAINING\_COMPOUND[GSEA]

|  || Dataset | SexDevelopmentGenesPRL\_remapped |
| Phenotype | NoPhenotypeAvailable |
| Upregulated in class | na\_pos |
| GeneSet | GOBP\_RESPONSE\_TO\_OXYGEN\_CONTAINING\_COMPOUND |
| Enrichment Score (ES) | 0.33152977 |
| Normalized Enrichment Score (NES) | 1.6221907 |
| Nominal p-value | 0.05009634 |
| FDR q-value | 0.83546066 |
| FWER p-Value | 1.0 |
Table: GSEA Results Summary

  

Fig 1: Enrichment plot: GOBP\_RESPONSE\_TO\_OXYGEN\_CONTAINING\_COMPOUND      
 Profile of the Running ES Score & Positions of GeneSet Members on the Rank Ordered List

  

| SYMBOL | TITLE | RANK IN GENE LIST | RANK METRIC SCORE | RUNNING ES | CORE ENRICHMENT || 1 | SOX9 | SRY-box transcription factor 9 [Source:HGNC Symbol;Acc:HGNC:11204] | 1 | 121.000 | 0.0433 | Yes |
| 2 | FLNA | filamin A [Source:HGNC Symbol;Acc:HGNC:3754] | 3 | 53.800 | 0.0866 | Yes |
| 3 | LHCGR | luteinizing hormone/choriogonadotropin receptor [Source:HGNC Symbol;Acc:HGNC:6585] | 18 | 11.900 | 0.0084 | Yes |
| 4 | H6PD | hexose-6-phosphate dehydrogenase/glucose 1-dehydrogenase [Source:HGNC Symbol;Acc:HGNC:4795] | 21 | 11.600 | 0.0423 | Yes |
| 5 | AKR1C2 | aldo-keto reductase family 1 member C2 [Source:HGNC Symbol;Acc:HGNC:385] | 22 | 10.700 | 0.0949 | Yes |
| 6 | GNRH1 | gonadotropin releasing hormone 1 [Source:HGNC Symbol;Acc:HGNC:4419] | 23 | 9.790 | 0.1476 | Yes |
| 7 | NR3C1 | nuclear receptor subfamily 3 group C member 1 [Source:HGNC Symbol;Acc:HGNC:7978] | 29 | 7.690 | 0.1535 | Yes |
| 8 | UBR1 | ubiquitin protein ligase E3 component n-recognin 1 [Source:HGNC Symbol;Acc:HGNC:16808] | 38 | 5.250 | 0.1313 | Yes |
| 9 | PTPN11 | protein tyrosine phosphatase non-receptor type 11 [Source:HGNC Symbol;Acc:HGNC:9644] | 39 | 4.970 | 0.1840 | Yes |
| 10 | HOXA10 | homeobox A10 [Source:HGNC Symbol;Acc:HGNC:5100] | 41 | 3.830 | 0.2273 | Yes |
| 11 | NSMF | NMDA receptor synaptonuclear signaling and neuronal migration factor [Source:HGNC Symbol;Acc:HGNC:29843] | 55 | 1.360 | 0.1584 | Yes |
| 12 | CYP11A1 | cytochrome P450 family 11 subfamily A member 1 [Source:HGNC Symbol;Acc:HGNC:2590] | 56 | 1.050 | 0.2110 | Yes |
| 13 | FGF10 | fibroblast growth factor 10 [Source:HGNC Symbol;Acc:HGNC:3666] | 59 | 0.890 | 0.2450 | Yes |
| 14 | FGFR2 | fibroblast growth factor receptor 2 [Source:HGNC Symbol;Acc:HGNC:3689] | 60 | 0.760 | 0.2976 | Yes |
| 15 | AR | androgen receptor [Source:HGNC Symbol;Acc:HGNC:644] | 63 | 0.490 | 0.3315 | Yes |
| 16 | CYP19A1 | cytochrome P450 family 19 subfamily A member 1 [Source:HGNC Symbol;Acc:HGNC:2594] | 81 | -0.300 | 0.2253 | No |
| 17 | AKR1C4 | aldo-keto reductase family 1 member C4 [Source:HGNC Symbol;Acc:HGNC:387] | 93 | -0.670 | 0.1751 | No |
| 18 | POR | cytochrome p450 oxidoreductase [Source:HGNC Symbol;Acc:HGNC:9208] | 108 | -3.990 | 0.0969 | No |
| 19 | WNT5A | Wnt family member 5A [Source:HGNC Symbol;Acc:HGNC:12784] | 115 | -15.700 | 0.0935 | No |
Table: GSEA details [plain text format]

  

Fig 2: GOBP\_RESPONSE\_TO\_OXYGEN\_CONTAINING\_COMPOUND: Random ES distribution      
 Gene set null distribution of ES for **GOBP\_RESPONSE\_TO\_OXYGEN\_CONTAINING\_COMPOUND**

  
