## Supplemental File 3 for "Increased Expression of *ZFPM2* Bypasses *SRY* to Drive 46,XX Testicular Development: A New Mechanism of 46,XX DSD": GOBP_SENSORY_ORGAN_DEVELOPMENT.html

Details for gene set GOBP\_SENSORY\_ORGAN\_DEVELOPMENT[GSEA]

|  || Dataset | SexDevelopmentGenesPRL\_remapped |
| Phenotype | NoPhenotypeAvailable |
| Upregulated in class | na\_neg |
| GeneSet | GOBP\_SENSORY\_ORGAN\_DEVELOPMENT |
| Enrichment Score (ES) | -0.38095242 |
| Normalized Enrichment Score (NES) | -1.9316115 |
| Nominal p-value | 0.0037664783 |
| FDR q-value | 0.44048163 |
| FWER p-Value | 0.985 |
Table: GSEA Results Summary

  

Fig 1: Enrichment plot: GOBP\_SENSORY\_ORGAN\_DEVELOPMENT      
 Profile of the Running ES Score & Positions of GeneSet Members on the Rank Ordered List

  

| SYMBOL | TITLE | RANK IN GENE LIST | RANK METRIC SCORE | RUNNING ES | CORE ENRICHMENT || 1 | SOX9 | SRY-box transcription factor 9 [Source:HGNC Symbol;Acc:HGNC:11204] | 1 | 121.000 | 0.0381 | No |
| 2 | SOS1 | SOS Ras/Rac guanine nucleotide exchange factor 1 [Source:HGNC Symbol;Acc:HGNC:11187] | 15 | 12.800 | -0.0381 | No |
| 3 | PTPN11 | protein tyrosine phosphatase non-receptor type 11 [Source:HGNC Symbol;Acc:HGNC:9644] | 39 | 4.970 | -0.2095 | No |
| 4 | HESX1 | HESX homeobox 1 [Source:HGNC Symbol;Acc:HGNC:4877] | 51 | 1.570 | -0.2667 | No |
| 5 | FGF10 | fibroblast growth factor 10 [Source:HGNC Symbol;Acc:HGNC:3666] | 59 | 0.890 | -0.2857 | No |
| 6 | FGFR2 | fibroblast growth factor receptor 2 [Source:HGNC Symbol;Acc:HGNC:3689] | 60 | 0.760 | -0.2381 | No |
| 7 | BMP4 | bone morphogenetic protein 4 [Source:HGNC Symbol;Acc:HGNC:1071] | 61 | 0.750 | -0.1905 | No |
| 8 | BBS7 | Bardet-Biedl syndrome 7 [Source:HGNC Symbol;Acc:HGNC:18758] | 66 | 0.400 | -0.1810 | No |
| 9 | MKS1 | MKS transition zone complex subunit 1 [Source:HGNC Symbol;Acc:HGNC:7121] | 88 | -0.480 | -0.3333 | Yes |
| 10 | CDKN1C | cyclin dependent kinase inhibitor 1C [Source:HGNC Symbol;Acc:HGNC:1786] | 89 | -0.530 | -0.2857 | Yes |
| 11 | ROR2 | receptor tyrosine kinase like orphan receptor 2 [Source:HGNC Symbol;Acc:HGNC:10257] | 91 | -0.580 | -0.2476 | Yes |
| 12 | TTC8 | tetratricopeptide repeat domain 8 [Source:HGNC Symbol;Acc:HGNC:20087] | 92 | -0.640 | -0.2000 | Yes |
| 13 | BBS4 | Bardet-Biedl syndrome 4 [Source:HGNC Symbol;Acc:HGNC:969] | 100 | -1.360 | -0.2190 | Yes |
| 14 | GLI3 | GLI family zinc finger 3 [Source:HGNC Symbol;Acc:HGNC:4319] | 104 | -2.920 | -0.2000 | Yes |
| 15 | CHD7 | chromodomain helicase DNA binding protein 7 [Source:HGNC Symbol;Acc:HGNC:20626] | 105 | -3.230 | -0.1524 | Yes |
| 16 | PITX2 | paired like homeodomain 2 [Source:HGNC Symbol;Acc:HGNC:9005] | 107 | -3.730 | -0.1143 | Yes |
| 17 | PBX1 | PBX homeobox 1 [Source:HGNC Symbol;Acc:HGNC:8632] | 112 | -12.600 | -0.1048 | Yes |
| 18 | WNT5A | Wnt family member 5A [Source:HGNC Symbol;Acc:HGNC:12784] | 115 | -15.700 | -0.0762 | Yes |
| 19 | FREM2 | FRAS1 related extracellular matrix 2 [Source:HGNC Symbol;Acc:HGNC:25396] | 117 | -21.900 | -0.0381 | Yes |
| 20 | FOXL2 | forkhead box L2 [Source:HGNC Symbol;Acc:HGNC:1092] | 122 | -40.400 | -0.0286 | Yes |
| 21 | BNC2 | basonuclin 2 [Source:HGNC Symbol;Acc:HGNC:30988] | 123 | -42.000 | 0.0190 | Yes |
Table: GSEA details [plain text format]

  

Fig 2: GOBP\_SENSORY\_ORGAN\_DEVELOPMENT: Random ES distribution      
 Gene set null distribution of ES for **GOBP\_SENSORY\_ORGAN\_DEVELOPMENT**

  
