## Supplemental File 3 for "Increased Expression of *ZFPM2* Bypasses *SRY* to Drive 46,XX Testicular Development: A New Mechanism of 46,XX DSD": GOBP_SKELETAL_SYSTEM_DEVELOPMENT.html

Details for gene set GOBP\_SKELETAL\_SYSTEM\_DEVELOPMENT[GSEA]

|  || Dataset | SexDevelopmentGenesPRL\_remapped |
| Phenotype | NoPhenotypeAvailable |
| Upregulated in class | na\_neg |
| GeneSet | GOBP\_SKELETAL\_SYSTEM\_DEVELOPMENT |
| Enrichment Score (ES) | -0.34655973 |
| Normalized Enrichment Score (NES) | -1.8156984 |
| Nominal p-value | 0.015414258 |
| FDR q-value | 0.4691986 |
| FWER p-Value | 1.0 |
Table: GSEA Results Summary

  

Fig 1: Enrichment plot: GOBP\_SKELETAL\_SYSTEM\_DEVELOPMENT      
 Profile of the Running ES Score & Positions of GeneSet Members on the Rank Ordered List

  

| SYMBOL | TITLE | RANK IN GENE LIST | RANK METRIC SCORE | RUNNING ES | CORE ENRICHMENT || 1 | SOX9 | SRY-box transcription factor 9 [Source:HGNC Symbol;Acc:HGNC:11204] | 1 | 121.000 | 0.0338 | No |
| 2 | MED12 | mediator complex subunit 12 [Source:HGNC Symbol;Acc:HGNC:11957] | 33 | 6.470 | -0.2237 | No |
| 3 | MKKS | MKKS centrosomal shuttling protein [Source:HGNC Symbol;Acc:HGNC:7108] | 34 | 6.190 | -0.1802 | No |
| 4 | FGFR3 | fibroblast growth factor receptor 3 [Source:HGNC Symbol;Acc:HGNC:3690] | 37 | 5.830 | -0.1562 | No |
| 5 | PTPN11 | protein tyrosine phosphatase non-receptor type 11 [Source:HGNC Symbol;Acc:HGNC:9644] | 39 | 4.970 | -0.1224 | No |
| 6 | HOXA10 | homeobox A10 [Source:HGNC Symbol;Acc:HGNC:5100] | 41 | 3.830 | -0.0886 | No |
| 7 | HOXB6 | homeobox B6 [Source:HGNC Symbol;Acc:HGNC:5117] | 46 | 2.100 | -0.0840 | No |
| 8 | FGFR2 | fibroblast growth factor receptor 2 [Source:HGNC Symbol;Acc:HGNC:3689] | 60 | 0.760 | -0.1667 | No |
| 9 | BMP4 | bone morphogenetic protein 4 [Source:HGNC Symbol;Acc:HGNC:1071] | 61 | 0.750 | -0.1233 | No |
| 10 | HOXA4 | homeobox A4 [Source:HGNC Symbol;Acc:HGNC:5105] | 85 | -0.350 | -0.3031 | Yes |
| 11 | MKS1 | MKS transition zone complex subunit 1 [Source:HGNC Symbol;Acc:HGNC:7121] | 88 | -0.480 | -0.2790 | Yes |
| 12 | CDKN1C | cyclin dependent kinase inhibitor 1C [Source:HGNC Symbol;Acc:HGNC:1786] | 89 | -0.530 | -0.2355 | Yes |
| 13 | FGFR1 | fibroblast growth factor receptor 1 [Source:HGNC Symbol;Acc:HGNC:3688] | 90 | -0.570 | -0.1921 | Yes |
| 14 | ROR2 | receptor tyrosine kinase like orphan receptor 2 [Source:HGNC Symbol;Acc:HGNC:10257] | 91 | -0.580 | -0.1486 | Yes |
| 15 | GLI3 | GLI family zinc finger 3 [Source:HGNC Symbol;Acc:HGNC:4319] | 104 | -2.920 | -0.2216 | Yes |
| 16 | CHD7 | chromodomain helicase DNA binding protein 7 [Source:HGNC Symbol;Acc:HGNC:20626] | 105 | -3.230 | -0.1781 | Yes |
| 17 | PITX2 | paired like homeodomain 2 [Source:HGNC Symbol;Acc:HGNC:9005] | 107 | -3.730 | -0.1444 | Yes |
| 18 | POR | cytochrome p450 oxidoreductase [Source:HGNC Symbol;Acc:HGNC:9208] | 108 | -3.990 | -0.1009 | Yes |
| 19 | PBX1 | PBX homeobox 1 [Source:HGNC Symbol;Acc:HGNC:8632] | 112 | -12.600 | -0.0865 | Yes |
| 20 | WWOX | WW domain containing oxidoreductase [Source:HGNC Symbol;Acc:HGNC:12799] | 113 | -14.500 | -0.0431 | Yes |
| 21 | WNT5A | Wnt family member 5A [Source:HGNC Symbol;Acc:HGNC:12784] | 115 | -15.700 | -0.0093 | Yes |
| 22 | TP63 | tumor protein p63 [Source:HGNC Symbol;Acc:HGNC:15979] | 116 | -20.700 | 0.0342 | Yes |
| 23 | EVC | EvC ciliary complex subunit 1 [Source:HGNC Symbol;Acc:HGNC:3497] | 119 | -30.300 | 0.0583 | Yes |
Table: GSEA details [plain text format]

  

Fig 2: GOBP\_SKELETAL\_SYSTEM\_DEVELOPMENT: Random ES distribution      
 Gene set null distribution of ES for **GOBP\_SKELETAL\_SYSTEM\_DEVELOPMENT**

  
