## Supplemental File 3 for "Increased Expression of *ZFPM2* Bypasses *SRY* to Drive 46,XX Testicular Development: A New Mechanism of 46,XX DSD": GOCC_CATALYTIC_COMPLEX.html

Details for gene set GOCC\_CATALYTIC\_COMPLEX[GSEA]

|  || Dataset | SexDevelopmentGenesPRL\_remapped |
| Phenotype | NoPhenotypeAvailable |
| Upregulated in class | na\_pos |
| GeneSet | GOCC\_CATALYTIC\_COMPLEX |
| Enrichment Score (ES) | 0.4254386 |
| Normalized Enrichment Score (NES) | 1.7119213 |
| Nominal p-value | 0.038910504 |
| FDR q-value | 0.8488784 |
| FWER p-Value | 1.0 |
Table: GSEA Results Summary

  

Fig 1: Enrichment plot: GOCC\_CATALYTIC\_COMPLEX      
 Profile of the Running ES Score & Positions of GeneSet Members on the Rank Ordered List

  

| SYMBOL | TITLE | RANK IN GENE LIST | RANK METRIC SCORE | RUNNING ES | CORE ENRICHMENT || 1 | CHD4 | chromodomain helicase DNA binding protein 4 [Source:HGNC Symbol;Acc:HGNC:1919] | 0 | 149.000 | 0.0833 | Yes |
| 2 | SOS1 | SOS Ras/Rac guanine nucleotide exchange factor 1 [Source:HGNC Symbol;Acc:HGNC:11187] | 15 | 12.800 | 0.0439 | Yes |
| 3 | CREBBP | CREB binding protein [Source:HGNC Symbol;Acc:HGNC:2348] | 20 | 11.800 | 0.0921 | Yes |
| 4 | BCOR | BCL6 corepressor [Source:HGNC Symbol;Acc:HGNC:20893] | 25 | 9.140 | 0.1404 | Yes |
| 5 | KAT6B | lysine acetyltransferase 6B [Source:HGNC Symbol;Acc:HGNC:17582] | 28 | 8.070 | 0.2061 | Yes |
| 6 | MED12 | mediator complex subunit 12 [Source:HGNC Symbol;Acc:HGNC:11957] | 33 | 6.470 | 0.2544 | Yes |
| 7 | UBR1 | ubiquitin protein ligase E3 component n-recognin 1 [Source:HGNC Symbol;Acc:HGNC:16808] | 38 | 5.250 | 0.3026 | Yes |
| 8 | FBXL4 | F-box and leucine rich repeat protein 4 [Source:HGNC Symbol;Acc:HGNC:13601] | 42 | 3.740 | 0.3596 | Yes |
| 9 | DNMT3B | DNA methyltransferase 3 beta [Source:HGNC Symbol;Acc:HGNC:2979] | 45 | 2.110 | 0.4254 | Yes |
| 10 | DYNC2H1 | dynein cytoplasmic 2 heavy chain 1 [Source:HGNC Symbol;Acc:HGNC:2962] | 79 | -0.230 | 0.2193 | No |
| 11 | CBX2 | chromobox 2 [Source:HGNC Symbol;Acc:HGNC:1552] | 87 | -0.410 | 0.2412 | No |
| 12 | CUL7 | cullin 7 [Source:HGNC Symbol;Acc:HGNC:21024] | 96 | -0.950 | 0.2544 | No |
Table: GSEA details [plain text format]

  

Fig 2: GOCC\_CATALYTIC\_COMPLEX: Random ES distribution      
 Gene set null distribution of ES for **GOCC\_CATALYTIC\_COMPLEX**

  
