## Supplemental File 3 for "Increased Expression of *ZFPM2* Bypasses *SRY* to Drive 46,XX Testicular Development: A New Mechanism of 46,XX DSD": GOCC_PLASMA_MEMBRANE_REGION.html

Details for gene set GOCC\_PLASMA\_MEMBRANE\_REGION[GSEA]

|  || Dataset | SexDevelopmentGenesPRL\_remapped |
| Phenotype | NoPhenotypeAvailable |
| Upregulated in class | na\_neg |
| GeneSet | GOCC\_PLASMA\_MEMBRANE\_REGION |
| Enrichment Score (ES) | -0.4385965 |
| Normalized Enrichment Score (NES) | -1.7613256 |
| Nominal p-value | 0.025423728 |
| FDR q-value | 0.5077113 |
| FWER p-Value | 1.0 |
Table: GSEA Results Summary

  

Fig 1: Enrichment plot: GOCC\_PLASMA\_MEMBRANE\_REGION      
 Profile of the Running ES Score & Positions of GeneSet Members on the Rank Ordered List

  

| SYMBOL | TITLE | RANK IN GENE LIST | RANK METRIC SCORE | RUNNING ES | CORE ENRICHMENT || 1 | PDE4D | phosphodiesterase 4D [Source:HGNC Symbol;Acc:HGNC:8783] | 9 | 18.600 | 0.0044 | No |
| 2 | BBS9 | Bardet-Biedl syndrome 9 [Source:HGNC Symbol;Acc:HGNC:30000] | 47 | 2.000 | -0.2368 | No |
| 3 | BBS2 | Bardet-Biedl syndrome 2 [Source:HGNC Symbol;Acc:HGNC:967] | 48 | 1.960 | -0.1535 | No |
| 4 | BBS7 | Bardet-Biedl syndrome 7 [Source:HGNC Symbol;Acc:HGNC:18758] | 66 | 0.400 | -0.2193 | No |
| 5 | TTC8 | tetratricopeptide repeat domain 8 [Source:HGNC Symbol;Acc:HGNC:20087] | 92 | -0.640 | -0.3553 | Yes |
| 6 | GRIP1 | glutamate receptor interacting protein 1 [Source:HGNC Symbol;Acc:HGNC:18708] | 94 | -0.750 | -0.2807 | Yes |
| 7 | EVC2 | EvC ciliary complex subunit 2 [Source:HGNC Symbol;Acc:HGNC:19747] | 95 | -0.890 | -0.1974 | Yes |
| 8 | TCTN3 | tectonic family member 3 [Source:HGNC Symbol;Acc:HGNC:24519] | 97 | -1.040 | -0.1228 | Yes |
| 9 | BBS4 | Bardet-Biedl syndrome 4 [Source:HGNC Symbol;Acc:HGNC:969] | 100 | -1.360 | -0.0570 | Yes |
| 10 | LEPR | leptin receptor [Source:HGNC Symbol;Acc:HGNC:6554] | 111 | -8.460 | -0.0614 | Yes |
| 11 | BBS5 | Bardet-Biedl syndrome 5 [Source:HGNC Symbol;Acc:HGNC:970] | 114 | -14.600 | 0.0044 | Yes |
| 12 | EVC | EvC ciliary complex subunit 1 [Source:HGNC Symbol;Acc:HGNC:3497] | 119 | -30.300 | 0.0526 | Yes |
Table: GSEA details [plain text format]

  

Fig 2: GOCC\_PLASMA\_MEMBRANE\_REGION: Random ES distribution      
 Gene set null distribution of ES for **GOCC\_PLASMA\_MEMBRANE\_REGION**

  
