## Supplemental File 3 for "Increased Expression of *ZFPM2* Bypasses *SRY* to Drive 46,XX Testicular Development: A New Mechanism of 46,XX DSD": gsea_report_for_na_neg_1653344355978.html

Report for na\_neg 1653344355978 [GSEA]

| GS  follow link to MSigDB | GS DETAILS | SIZE | ES | NES | NOM p-val | FDR q-val | FWER p-val | RANK AT MAX | LEADING EDGE || 1 | HP\_FINGER\_SYNDACTYLY | Details ... | 39 | -0.36 | -2.24 | 0.000 | 0.612 | 0.438 | 83 | tags=90%, list=66%, signal=182% |
| 2 | HP\_CLEFT\_LIP | Details ... | 29 | -0.39 | -2.21 | 0.002 | 0.392 | 0.525 | 76 | tags=90%, list=60%, signal=174% |
| 3 | HP\_CLEFT\_UPPER\_LIP | Details ... | 26 | -0.38 | -2.12 | 0.002 | 0.450 | 0.725 | 55 | tags=73%, list=44%, signal=103% |
| 4 | HP\_ABNORMALITY\_OF\_GLOBE\_SIZE | Details ... | 19 | -0.42 | -2.07 | 0.004 | 0.481 | 0.837 | 69 | tags=89%, list=55%, signal=168% |
| 5 | HP\_SYNDACTYLY | Details ... | 49 | -0.31 | -2.04 | 0.002 | 0.465 | 0.892 | 80 | tags=82%, list=63%, signal=137% |
| 6 | HP\_SMALL\_NAIL | Details ... | 12 | -0.51 | -2.04 | 0.002 | 0.392 | 0.893 | 69 | tags=100%, list=55%, signal=200% |
| 7 | HP\_ABNORMAL\_GROWTH\_HORMONE\_LEVEL | Details ... | 10 | -0.55 | -2.03 | 0.002 | 0.344 | 0.897 | 38 | tags=80%, list=30%, signal=105% |
| 8 | HP\_APLASIA\_HYPOPLASIA\_OF\_THE\_NAILS | Details ... | 14 | -0.46 | -1.97 | 0.006 | 0.455 | 0.965 | 75 | tags=100%, list=60%, signal=220% |
| 9 | HP\_APLASIA\_HYPOPLASIA\_AFFECTING\_THE\_EYE | Details ... | 25 | -0.35 | -1.96 | 0.006 | 0.420 | 0.971 | 41 | tags=60%, list=33%, signal=71% |
| 10 | GOBP\_SENSORY\_ORGAN\_DEVELOPMENT | Details ... | 21 | -0.38 | -1.93 | 0.004 | 0.440 | 0.985 | 39 | tags=62%, list=31%, signal=75% |
| 11 | HP\_HAND\_POLYDACTYLY | Details ... | 30 | -0.33 | -1.87 | 0.019 | 0.581 | 0.996 | 83 | tags=90%, list=66%, signal=201% |
| 12 | HP\_VERTEBRAL\_SEGMENTATION\_DEFECT | Details ... | 13 | -0.45 | -1.86 | 0.017 | 0.539 | 0.996 | 37 | tags=69%, list=29%, signal=88% |
| 13 | GOBP\_UROGENITAL\_SYSTEM\_DEVELOPMENT | Details ... | 20 | -0.38 | -1.86 | 0.014 | 0.516 | 0.999 | 68 | tags=85%, list=54%, signal=155% |
| 14 | HP\_ABNORMAL\_RETINAL\_MORPHOLOGY | Details ... | 33 | -0.31 | -1.84 | 0.016 | 0.515 | 1.000 | 83 | tags=88%, list=66%, signal=190% |
| 15 | HP\_OLIGOHYDRAMNIOS | Details ... | 10 | -0.49 | -1.84 | 0.017 | 0.501 | 1.000 | 45 | tags=80%, list=36%, signal=115% |
| 16 | GOBP\_MORPHOGENESIS\_OF\_A\_BRANCHING\_STRUCTURE | Details ... | 13 | -0.44 | -1.83 | 0.012 | 0.500 | 1.000 | 68 | tags=92%, list=54%, signal=180% |
| 17 | HP\_ABNORMAL\_FINGERNAIL\_MORPHOLOGY | Details ... | 12 | -0.46 | -1.82 | 0.016 | 0.492 | 1.000 | 75 | tags=100%, list=60%, signal=224% |
| 18 | GOBP\_SKELETAL\_SYSTEM\_DEVELOPMENT | Details ... | 23 | -0.35 | -1.82 | 0.015 | 0.469 | 1.000 | 42 | tags=61%, list=33%, signal=75% |
| 19 | HP\_DUPLICATION\_OF\_HAND\_BONES | Details ... | 36 | -0.29 | -1.80 | 0.020 | 0.472 | 1.000 | 83 | tags=86%, list=66%, signal=180% |
| 20 | HP\_ABNORMAL\_LOCALIZATION\_OF\_KIDNEY | Details ... | 16 | -0.39 | -1.79 | 0.015 | 0.476 | 1.000 | 37 | tags=63%, list=29%, signal=77% |
| 21 | HP\_APLASIA\_HYPOPLASIA\_OF\_THE\_CEREBELLAR\_VERMIS | Details ... | 21 | -0.34 | -1.76 | 0.023 | 0.526 | 1.000 | 55 | tags=71%, list=44%, signal=106% |
| 22 | GOCC\_PLASMA\_MEMBRANE\_REGION | Details ... | 12 | -0.44 | -1.76 | 0.025 | 0.508 | 1.000 | 35 | tags=67%, list=28%, signal=84% |
| 23 | HP\_ABNORMAL\_CEREBELLAR\_VERMIS\_MORPHOLOGY | Details ... | 21 | -0.34 | -1.74 | 0.028 | 0.536 | 1.000 | 55 | tags=71%, list=44%, signal=106% |
| 24 | WP\_CILIOPATHIES | Details ... | 20 | -0.36 | -1.74 | 0.027 | 0.523 | 1.000 | 83 | tags=95%, list=66%, signal=234% |
| 25 | GOBP\_EMBRYONIC\_APPENDAGE\_MORPHOGENESIS | Details ... | 14 | -0.41 | -1.73 | 0.022 | 0.524 | 1.000 | 54 | tags=79%, list=43%, signal=122% |
| 26 | HP\_APLASIA\_HYPOPLASIA\_INVOLVING\_BONES\_OF\_THE\_UPPER\_LIMBS | Details ... | 41 | -0.27 | -1.72 | 0.033 | 0.541 | 1.000 | 76 | tags=78%, list=60%, signal=133% |
| 27 | HP\_BIFID\_TONGUE |  | 10 | -0.47 | -1.71 | 0.022 | 0.532 | 1.000 | 73 | tags=100%, list=58%, signal=219% |
| 28 | HP\_DUPLICATION\_INVOLVING\_BONES\_OF\_THE\_FEET |  | 17 | -0.36 | -1.71 | 0.023 | 0.515 | 1.000 | 80 | tags=94%, list=63%, signal=223% |
| 29 | GOBP\_APPENDAGE\_DEVELOPMENT |  | 21 | -0.33 | -1.71 | 0.017 | 0.509 | 1.000 | 68 | tags=81%, list=54%, signal=147% |
| 30 | HP\_POLYDACTYLY |  | 34 | -0.28 | -1.70 | 0.032 | 0.504 | 1.000 | 83 | tags=85%, list=66%, signal=182% |
| 31 | HP\_DILATED\_FOURTH\_VENTRICLE |  | 12 | -0.42 | -1.70 | 0.031 | 0.500 | 1.000 | 48 | tags=75%, list=38%, signal=110% |
| 32 | GOBP\_IMMUNE\_SYSTEM\_DEVELOPMENT |  | 15 | -0.38 | -1.70 | 0.034 | 0.486 | 1.000 | 68 | tags=87%, list=54%, signal=166% |
| 33 | GOBP\_GLAND\_DEVELOPMENT |  | 18 | -0.35 | -1.69 | 0.028 | 0.500 | 1.000 | 68 | tags=83%, list=54%, signal=155% |
| 34 | HP\_LIMB\_UNDERGROWTH |  | 27 | -0.30 | -1.67 | 0.032 | 0.524 | 1.000 | 74 | tags=81%, list=59%, signal=155% |
| 35 | CTTTAAR\_UNKNOWN |  | 14 | -0.38 | -1.67 | 0.037 | 0.509 | 1.000 | 21 | tags=50%, list=17%, signal=53% |
| 36 | TRAVAGLINI\_LUNG\_CILIATED\_CELL |  | 12 | -0.41 | -1.67 | 0.051 | 0.496 | 1.000 | 80 | tags=100%, list=63%, signal=248% |
| 37 | HP\_HYPOPLASTIC\_FEMALE\_EXTERNAL\_GENITALIA |  | 12 | -0.42 | -1.66 | 0.031 | 0.497 | 1.000 | 69 | tags=92%, list=55%, signal=183% |
| 38 | NUP153\_TARGET\_GENES |  | 13 | -0.39 | -1.65 | 0.037 | 0.508 | 1.000 | 44 | tags=69%, list=35%, signal=95% |
| 39 | HP\_ABNORMAL\_TOENAIL\_MORPHOLOGY |  | 10 | -0.45 | -1.65 | 0.037 | 0.514 | 1.000 | 75 | tags=100%, list=60%, signal=227% |
| 40 | HP\_CONICAL\_INCISOR |  | 11 | -0.41 | -1.64 | 0.034 | 0.513 | 1.000 | 68 | tags=91%, list=54%, signal=180% |
| 41 | HP\_HEPATIC\_FIBROSIS |  | 18 | -0.34 | -1.64 | 0.033 | 0.510 | 1.000 | 83 | tags=94%, list=66%, signal=237% |
| 42 | BARX2\_TARGET\_GENES |  | 13 | -0.39 | -1.63 | 0.043 | 0.514 | 1.000 | 44 | tags=69%, list=35%, signal=95% |
| 43 | HP\_SHORT\_LONG\_BONE |  | 23 | -0.31 | -1.63 | 0.035 | 0.512 | 1.000 | 73 | tags=83%, list=58%, signal=161% |
| 44 | MEISSNER\_BRAIN\_HCP\_WITH\_H3K4ME3\_AND\_H3K27ME3 |  | 18 | -0.34 | -1.62 | 0.035 | 0.517 | 1.000 | 69 | tags=83%, list=55%, signal=158% |
| 45 | WP\_GENES\_RELATED\_TO\_PRIMARY\_CILIUM\_DEVELOPMENT\_BASED\_ON\_CRISPR |  | 16 | -0.36 | -1.61 | 0.050 | 0.516 | 1.000 | 80 | tags=94%, list=63%, signal=224% |
| 46 | GOBP\_SENSORY\_SYSTEM\_DEVELOPMENT |  | 17 | -0.34 | -1.61 | 0.042 | 0.526 | 1.000 | 38 | tags=59%, list=30%, signal=73% |
| 47 | HP\_CONICAL\_TOOTH |  | 11 | -0.41 | -1.61 | 0.048 | 0.515 | 1.000 | 68 | tags=91%, list=54%, signal=180% |
| 48 | HP\_ABNORMALITY\_OF\_RETINAL\_PIGMENTATION |  | 18 | -0.34 | -1.60 | 0.039 | 0.508 | 1.000 | 83 | tags=94%, list=66%, signal=237% |
| 49 | GOBP\_SPECIFICATION\_OF\_SYMMETRY |  | 11 | -0.41 | -1.60 | 0.046 | 0.508 | 1.000 | 68 | tags=91%, list=54%, signal=180% |
| 50 | GOBP\_BRANCHING\_MORPHOGENESIS\_OF\_AN\_EPITHELIAL\_TUBE |  | 11 | -0.41 | -1.60 | 0.044 | 0.505 | 1.000 | 68 | tags=91%, list=54%, signal=180% |
| 51 | HP\_CEREBELLAR\_HYPOPLASIA |  | 26 | -0.30 | -1.59 | 0.046 | 0.500 | 1.000 | 73 | tags=81%, list=58%, signal=152% |
| 52 | HP\_CORNEAL\_OPACITY |  | 17 | -0.34 | -1.59 | 0.043 | 0.493 | 1.000 | 75 | tags=88%, list=60%, signal=189% |
| 53 | HP\_EXTERNAL\_EAR\_MALFORMATION |  | 13 | -0.39 | -1.59 | 0.058 | 0.487 | 1.000 | 54 | tags=77%, list=43%, signal=121% |
| 54 | HP\_ABNORMAL\_LABIA\_MAJORA\_MORPHOLOGY |  | 11 | -0.40 | -1.58 | 0.046 | 0.495 | 1.000 | 69 | tags=91%, list=55%, signal=183% |
| 55 | HP\_HYPOPITUITARISM |  | 17 | -0.34 | -1.57 | 0.044 | 0.522 | 1.000 | 38 | tags=59%, list=30%, signal=73% |
| 56 | HP\_ABNORMALITY\_OF\_PRENATAL\_DEVELOPMENT\_OR\_BIRTH |  | 42 | -0.25 | -1.56 | 0.038 | 0.524 | 1.000 | 76 | tags=76%, list=60%, signal=128% |
| 57 | GOBP\_REGIONALIZATION |  | 17 | -0.34 | -1.56 | 0.061 | 0.531 | 1.000 | 68 | tags=82%, list=54%, signal=155% |
| 58 | GOBP\_SMOOTHENED\_SIGNALING\_PATHWAY |  | 14 | -0.37 | -1.55 | 0.051 | 0.526 | 1.000 | 68 | tags=86%, list=54%, signal=166% |
| 59 | HP\_ANAL\_STENOSIS |  | 10 | -0.41 | -1.55 | 0.052 | 0.519 | 1.000 | 54 | tags=80%, list=43%, signal=129% |
| 60 | GOBP\_SENSORY\_ORGAN\_MORPHOGENESIS |  | 13 | -0.38 | -1.55 | 0.063 | 0.522 | 1.000 | 36 | tags=62%, list=29%, signal=77% |
| 61 | HP\_ENCEPHALOCELE |  | 10 | -0.41 | -1.55 | 0.049 | 0.513 | 1.000 | 54 | tags=80%, list=43%, signal=129% |
| 62 | HP\_ABNORMALITY\_OF\_THE\_5TH\_TOE |  | 15 | -0.35 | -1.55 | 0.072 | 0.513 | 1.000 | 80 | tags=93%, list=63%, signal=225% |
| 63 | GOBP\_ANIMAL\_ORGAN\_MORPHOGENESIS |  | 37 | -0.24 | -1.54 | 0.057 | 0.527 | 1.000 | 68 | tags=70%, list=54%, signal=108% |
| 64 | HP\_RENAL\_HYPOPLASIA |  | 19 | -0.32 | -1.53 | 0.070 | 0.528 | 1.000 | 73 | tags=84%, list=58%, signal=170% |
| 65 | GOBP\_EMBRYONIC\_MORPHOGENESIS |  | 30 | -0.26 | -1.53 | 0.072 | 0.524 | 1.000 | 68 | tags=73%, list=54%, signal=121% |
| 66 | HP\_ABNORMAL\_VERTEBRAL\_MORPHOLOGY |  | 27 | -0.28 | -1.53 | 0.063 | 0.522 | 1.000 | 76 | tags=81%, list=60%, signal=161% |
| 67 | HP\_ABNORMAL\_FINGER\_PHALANX\_MORPHOLOGY |  | 56 | -0.22 | -1.53 | 0.071 | 0.519 | 1.000 | 80 | tags=75%, list=63%, signal=114% |
| 68 | HP\_ENLARGED\_POSTERIOR\_FOSSA |  | 13 | -0.36 | -1.52 | 0.059 | 0.516 | 1.000 | 48 | tags=69%, list=38%, signal=100% |
| 69 | HP\_ANOPHTHALMIA |  | 10 | -0.42 | -1.52 | 0.067 | 0.516 | 1.000 | 66 | tags=90%, list=52%, signal=174% |
| 70 | GOBP\_EPITHELIAL\_CELL\_PROLIFERATION |  | 10 | -0.40 | -1.51 | 0.067 | 0.525 | 1.000 | 68 | tags=90%, list=54%, signal=180% |
| 71 | GOBP\_MAMMARY\_GLAND\_DEVELOPMENT |  | 10 | -0.40 | -1.51 | 0.070 | 0.532 | 1.000 | 68 | tags=90%, list=54%, signal=180% |
| 72 | WP\_DEVELOPMENT\_OF\_URETERIC\_COLLECTION\_SYSTEM |  | 10 | -0.41 | -1.50 | 0.066 | 0.533 | 1.000 | 67 | tags=90%, list=53%, signal=177% |
| 73 | HP\_HIGHLY\_ARCHED\_EYEBROW |  | 12 | -0.37 | -1.50 | 0.063 | 0.526 | 1.000 | 43 | tags=67%, list=34%, signal=92% |
| 74 | GOBP\_REGULATION\_OF\_APOPTOTIC\_SIGNALING\_PATHWAY |  | 11 | -0.38 | -1.50 | 0.077 | 0.521 | 1.000 | 83 | tags=100%, list=66%, signal=267% |
| 75 | HP\_ABNORMAL\_FORM\_OF\_THE\_VERTEBRAL\_BODIES |  | 18 | -0.32 | -1.50 | 0.065 | 0.515 | 1.000 | 43 | tags=61%, list=34%, signal=80% |
| 76 | HP\_APLASIA\_HYPOPLASIA\_OF\_THE\_EXTREMITIES |  | 49 | -0.23 | -1.50 | 0.057 | 0.524 | 1.000 | 76 | tags=73%, list=60%, signal=113% |
| 77 | HP\_RETINAL\_DYSTROPHY |  | 18 | -0.31 | -1.49 | 0.067 | 0.524 | 1.000 | 80 | tags=89%, list=63%, signal=209% |
| 78 | GOBP\_CAMERA\_TYPE\_EYE\_DEVELOPMENT |  | 13 | -0.36 | -1.49 | 0.085 | 0.518 | 1.000 | 38 | tags=62%, list=30%, signal=79% |
| 79 | GOCC\_CILIUM |  | 19 | -0.32 | -1.49 | 0.079 | 0.515 | 1.000 | 80 | tags=89%, list=63%, signal=208% |
| 80 | HP\_FUNCTIONAL\_ABNORMALITY\_OF\_THE\_MIDDLE\_EAR |  | 22 | -0.30 | -1.49 | 0.086 | 0.512 | 1.000 | 79 | tags=86%, list=63%, signal=191% |
| 81 | HP\_CEREBELLAR\_CYST |  | 13 | -0.36 | -1.49 | 0.069 | 0.506 | 1.000 | 48 | tags=69%, list=38%, signal=100% |
| 82 | HP\_ABNORMALITY\_OF\_THE\_OUTER\_EAR |  | 78 | -0.23 | -1.49 | 0.065 | 0.505 | 1.000 | 45 | tags=44%, list=36%, signal=26% |
| 83 | HP\_HYPODONTIA |  | 16 | -0.33 | -1.48 | 0.066 | 0.519 | 1.000 | 75 | tags=88%, list=60%, signal=189% |
| 84 | HP\_SYNOSTOSIS\_OF\_CARPALS\_TARSALS |  | 11 | -0.38 | -1.47 | 0.070 | 0.518 | 1.000 | 37 | tags=64%, list=29%, signal=82% |
| 85 | GOBP\_GLAND\_MORPHOGENESIS |  | 10 | -0.40 | -1.47 | 0.075 | 0.514 | 1.000 | 68 | tags=90%, list=54%, signal=180% |
| 86 | HP\_HYPOPLASTIC\_LABIA\_MAJORA |  | 10 | -0.39 | -1.47 | 0.087 | 0.514 | 1.000 | 69 | tags=90%, list=55%, signal=183% |
| 87 | HP\_POSTAXIAL\_POLYDACTYLY |  | 24 | -0.28 | -1.47 | 0.077 | 0.514 | 1.000 | 83 | tags=88%, list=66%, signal=208% |
| 88 | HP\_CENTRAL\_NERVOUS\_SYSTEM\_CYST |  | 15 | -0.34 | -1.46 | 0.086 | 0.515 | 1.000 | 73 | tags=87%, list=58%, signal=182% |
| 89 | HP\_CLINODACTYLY |  | 42 | -0.23 | -1.46 | 0.063 | 0.514 | 1.000 | 39 | tags=45%, list=31%, signal=44% |
| 90 | HP\_ABNORMAL\_CHORIORETINAL\_MORPHOLOGY |  | 12 | -0.36 | -1.46 | 0.074 | 0.514 | 1.000 | 44 | tags=67%, list=35%, signal=93% |
| 91 | HP\_ABNORMAL\_TONGUE\_MORPHOLOGY |  | 26 | -0.27 | -1.46 | 0.093 | 0.511 | 1.000 | 76 | tags=81%, list=60%, signal=162% |
| 92 | HP\_LOW\_SET\_POSTERIORLY\_ROTATED\_EARS |  | 30 | -0.25 | -1.46 | 0.069 | 0.512 | 1.000 | 44 | tags=53%, list=35%, signal=62% |
| 93 | HP\_ABNORMALITY\_OF\_THE\_LIVER |  | 37 | -0.24 | -1.45 | 0.082 | 0.516 | 1.000 | 89 | tags=86%, list=71%, signal=208% |
| 94 | HP\_RENAL\_CYST |  | 31 | -0.25 | -1.45 | 0.102 | 0.511 | 1.000 | 83 | tags=84%, list=66%, signal=185% |
| 95 | HP\_TOE\_SYNDACTYLY |  | 28 | -0.27 | -1.45 | 0.080 | 0.506 | 1.000 | 74 | tags=79%, list=59%, signal=148% |
| 96 | HP\_ABNORMALITY\_OF\_THE\_NAIL |  | 30 | -0.24 | -1.44 | 0.098 | 0.529 | 1.000 | 91 | tags=90%, list=72%, signal=247% |
| 97 | HP\_ABNORMAL\_NUMBER\_OF\_INCISORS |  | 10 | -0.39 | -1.43 | 0.100 | 0.529 | 1.000 | 69 | tags=90%, list=55%, signal=183% |
| 98 | HP\_ABNORMAL\_ORAL\_FRENULUM\_MORPHOLOGY |  | 12 | -0.36 | -1.43 | 0.085 | 0.540 | 1.000 | 75 | tags=92%, list=60%, signal=205% |
| 99 | HP\_ABNORMALITY\_OF\_THE\_HYPOTHALAMUS\_PITUITARY\_AXIS |  | 36 | -0.23 | -1.42 | 0.097 | 0.545 | 1.000 | 43 | tags=50%, list=34%, signal=54% |
| 100 | GOBP\_EMBRYONIC\_ORGAN\_DEVELOPMENT |  | 24 | -0.27 | -1.42 | 0.082 | 0.542 | 1.000 | 42 | tags=54%, list=33%, signal=66% |
| 101 | HP\_PIGMENTARY\_RETINOPATHY |  | 15 | -0.32 | -1.42 | 0.102 | 0.546 | 1.000 | 83 | tags=93%, list=66%, signal=241% |
| 102 | HP\_ABNORMAL\_CHOROID\_MORPHOLOGY |  | 12 | -0.36 | -1.42 | 0.107 | 0.543 | 1.000 | 44 | tags=67%, list=35%, signal=93% |
| 103 | GOBP\_MORPHOGENESIS\_OF\_AN\_EPITHELIUM |  | 27 | -0.26 | -1.41 | 0.107 | 0.546 | 1.000 | 69 | tags=74%, list=55%, signal=129% |
| 104 | HP\_ABNORMALITY\_IRIS\_MORPHOLOGY |  | 21 | -0.28 | -1.41 | 0.082 | 0.544 | 1.000 | 44 | tags=57%, list=35%, signal=73% |
| 105 | HP\_MULTICYSTIC\_KIDNEY\_DYSPLASIA |  | 23 | -0.27 | -1.41 | 0.114 | 0.553 | 1.000 | 83 | tags=87%, list=66%, signal=208% |
| 106 | HP\_ABNORMALITY\_OF\_THE\_URETER |  | 31 | -0.24 | -1.40 | 0.105 | 0.549 | 1.000 | 39 | tags=48%, list=31%, signal=53% |
| 107 | HP\_ABNORMAL\_RENAL\_MORPHOLOGY |  | 68 | -0.21 | -1.40 | 0.108 | 0.544 | 1.000 | 39 | tags=40%, list=31%, signal=26% |
| 108 | HP\_ABNORMAL\_5TH\_FINGER\_MORPHOLOGY |  | 49 | -0.21 | -1.40 | 0.120 | 0.552 | 1.000 | 39 | tags=43%, list=31%, signal=38% |
| 109 | HP\_RENAL\_HYPOPLASIA\_APLASIA |  | 37 | -0.23 | -1.40 | 0.110 | 0.555 | 1.000 | 76 | tags=76%, list=60%, signal=135% |
| 110 | HP\_ABNORMALITY\_OF\_LONG\_BONE\_MORPHOLOGY |  | 48 | -0.22 | -1.40 | 0.130 | 0.552 | 1.000 | 76 | tags=73%, list=60%, signal=114% |
| 111 | HP\_ABNORMALITY\_OF\_THE\_CHOANAE |  | 18 | -0.29 | -1.39 | 0.094 | 0.552 | 1.000 | 75 | tags=83%, list=60%, signal=176% |
| 112 | HP\_APLASIA\_HYPOPLASIA\_INVOLVING\_FOREARM\_BONES |  | 13 | -0.33 | -1.39 | 0.110 | 0.549 | 1.000 | 70 | tags=85%, list=56%, signal=171% |
| 113 | HP\_ABNORMAL\_METATARSAL\_MORPHOLOGY |  | 16 | -0.30 | -1.39 | 0.120 | 0.549 | 1.000 | 70 | tags=81%, list=56%, signal=160% |
| 114 | HP\_POSTAXIAL\_HAND\_POLYDACTYLY |  | 23 | -0.27 | -1.39 | 0.115 | 0.548 | 1.000 | 83 | tags=87%, list=66%, signal=208% |
| 115 | HP\_COLOBOMA |  | 17 | -0.29 | -1.38 | 0.128 | 0.550 | 1.000 | 43 | tags=59%, list=34%, signal=77% |
| 116 | HP\_APLASIA\_HYPOPLASIA\_OF\_FINGERS |  | 31 | -0.24 | -1.38 | 0.116 | 0.550 | 1.000 | 76 | tags=77%, list=60%, signal=147% |
| 117 | HP\_APLASIA\_HYPOPLASIA\_OF\_THE\_CEREBELLUM |  | 28 | -0.24 | -1.38 | 0.117 | 0.547 | 1.000 | 76 | tags=79%, list=60%, signal=154% |
| 118 | HP\_ABNORMALITY\_OF\_THE\_TONGUE |  | 28 | -0.24 | -1.38 | 0.111 | 0.545 | 1.000 | 76 | tags=79%, list=60%, signal=154% |
| 119 | HP\_INCREASED\_BLOOD\_PRESSURE |  | 21 | -0.28 | -1.38 | 0.112 | 0.543 | 1.000 | 98 | tags=100%, list=78%, signal=375% |
| 120 | GOBP\_TISSUE\_MORPHOGENESIS |  | 29 | -0.24 | -1.37 | 0.122 | 0.553 | 1.000 | 69 | tags=72%, list=55%, signal=123% |
| 121 | HP\_CUTANEOUS\_SYNDACTYLY |  | 12 | -0.34 | -1.37 | 0.128 | 0.552 | 1.000 | 67 | tags=83%, list=53%, signal=161% |
| 122 | GOBP\_APPENDAGE\_MORPHOGENESIS |  | 18 | -0.29 | -1.37 | 0.123 | 0.548 | 1.000 | 68 | tags=78%, list=54%, signal=145% |
| 123 | HP\_SPECIFIC\_LEARNING\_DISABILITY |  | 11 | -0.36 | -1.37 | 0.114 | 0.548 | 1.000 | 63 | tags=82%, list=50%, signal=149% |
| 124 | HP\_WIDE\_NOSE |  | 10 | -0.36 | -1.35 | 0.133 | 0.582 | 1.000 | 73 | tags=90%, list=58%, signal=197% |
| 125 | GOBP\_PATTERN\_SPECIFICATION\_PROCESS |  | 26 | -0.25 | -1.35 | 0.147 | 0.579 | 1.000 | 68 | tags=73%, list=54%, signal=126% |
| 126 | HP\_ABNORMALITY\_OF\_THE\_PITUITARY\_GLAND |  | 34 | -0.22 | -1.34 | 0.150 | 0.593 | 1.000 | 40 | tags=47%, list=32%, signal=50% |
| 127 | HP\_SKELETAL\_MUSCLE\_ATROPHY |  | 20 | -0.26 | -1.34 | 0.136 | 0.590 | 1.000 | 93 | tags=95%, list=74%, signal=305% |
| 128 | GOBP\_EMBRYO\_DEVELOPMENT\_ENDING\_IN\_BIRTH\_OR\_EGG\_HATCHING |  | 20 | -0.27 | -1.34 | 0.133 | 0.588 | 1.000 | 42 | tags=55%, list=33%, signal=69% |
| 129 | HP\_APLASIA\_HYPOPLASIA\_INVOLVING\_BONES\_OF\_THE\_FEET |  | 24 | -0.25 | -1.34 | 0.124 | 0.587 | 1.000 | 75 | tags=79%, list=60%, signal=158% |
| 130 | GOBP\_EMBRYO\_DEVELOPMENT |  | 38 | -0.22 | -1.34 | 0.152 | 0.583 | 1.000 | 68 | tags=68%, list=54%, signal=104% |
| 131 | HP\_ABNORMALITY\_OF\_THE\_DENTITION |  | 62 | -0.19 | -1.33 | 0.139 | 0.607 | 1.000 | 76 | tags=69%, list=60%, signal=89% |
| 132 | HP\_ABNORMAL\_INCISOR\_MORPHOLOGY |  | 12 | -0.33 | -1.33 | 0.153 | 0.606 | 1.000 | 68 | tags=83%, list=54%, signal=164% |
| 133 | GOCC\_TRANSCRIPTION\_REGULATOR\_COMPLEX |  | 10 | -0.35 | -1.32 | 0.126 | 0.617 | 1.000 | 23 | tags=50%, list=18%, signal=56% |
| 134 | GOBP\_APOPTOTIC\_SIGNALING\_PATHWAY |  | 12 | -0.32 | -1.32 | 0.137 | 0.617 | 1.000 | 90 | tags=100%, list=71%, signal=317% |
| 135 | HP\_ABNORMALITY\_OF\_TOE |  | 51 | -0.20 | -1.31 | 0.153 | 0.620 | 1.000 | 70 | tags=67%, list=56%, signal=89% |
| 136 | GOBP\_POSITIVE\_REGULATION\_OF\_PROTEIN\_METABOLIC\_PROCESS |  | 18 | -0.28 | -1.31 | 0.138 | 0.619 | 1.000 | 90 | tags=94%, list=71%, signal=283% |
| 137 | HP\_ABNORMALITY\_OF\_THE\_BLADDER |  | 24 | -0.25 | -1.31 | 0.180 | 0.619 | 1.000 | 39 | tags=50%, list=31%, signal=59% |
| 138 | HP\_ABNORMAL\_LIVER\_MORPHOLOGY |  | 30 | -0.23 | -1.31 | 0.147 | 0.615 | 1.000 | 88 | tags=87%, list=70%, signal=219% |
| 139 | HP\_ABNORMALITY\_OF\_THE\_PINNA |  | 53 | -0.19 | -1.31 | 0.138 | 0.612 | 1.000 | 32 | tags=36%, list=25%, signal=28% |
| 140 | REACTOME\_CILIUM\_ASSEMBLY |  | 16 | -0.28 | -1.31 | 0.168 | 0.614 | 1.000 | 80 | tags=88%, list=63%, signal=209% |
| 141 | HP\_ABNORMAL\_EYELID\_MORPHOLOGY |  | 72 | -0.20 | -1.30 | 0.149 | 0.613 | 1.000 | 27 | tags=29%, list=21%, signal=16% |
| 142 | HP\_POSTERIORLY\_ROTATED\_EARS |  | 39 | -0.21 | -1.30 | 0.176 | 0.610 | 1.000 | 44 | tags=49%, list=35%, signal=52% |
| 143 | HP\_ABNORMAL\_NUMBER\_OF\_TEETH |  | 36 | -0.22 | -1.30 | 0.162 | 0.606 | 1.000 | 76 | tags=75%, list=60%, signal=135% |
| 144 | HP\_ABNORMALITY\_OF\_THE\_AUDITORY\_CANAL |  | 11 | -0.34 | -1.30 | 0.159 | 0.611 | 1.000 | 54 | tags=73%, list=43%, signal=116% |
| 145 | HP\_ABNORMALITY\_OF\_THE\_ABDOMINAL\_ORGANS |  | 42 | -0.20 | -1.30 | 0.133 | 0.613 | 1.000 | 89 | tags=83%, list=71%, signal=189% |
| 146 | HP\_ROD\_CONE\_DYSTROPHY |  | 12 | -0.32 | -1.29 | 0.166 | 0.620 | 1.000 | 80 | tags=92%, list=63%, signal=227% |
| 147 | HP\_POSTAXIAL\_FOOT\_POLYDACTYLY |  | 12 | -0.32 | -1.29 | 0.179 | 0.621 | 1.000 | 80 | tags=92%, list=63%, signal=227% |
| 148 | HP\_ABSENT\_TOE |  | 10 | -0.34 | -1.29 | 0.145 | 0.625 | 1.000 | 75 | tags=90%, list=60%, signal=205% |
| 149 | GOBP\_EPITHELIAL\_TUBE\_MORPHOGENESIS |  | 17 | -0.27 | -1.28 | 0.171 | 0.638 | 1.000 | 68 | tags=76%, list=54%, signal=144% |
| 150 | HP\_ABNORMAL\_SYSTEMIC\_BLOOD\_PRESSURE |  | 27 | -0.23 | -1.28 | 0.172 | 0.635 | 1.000 | 100 | tags=96%, list=79%, signal=367% |
| 151 | GOBP\_TELENCEPHALON\_DEVELOPMENT |  | 11 | -0.33 | -1.28 | 0.178 | 0.632 | 1.000 | 66 | tags=82%, list=52%, signal=157% |
| 152 | HP\_ECTOPIC\_ANUS |  | 20 | -0.25 | -1.28 | 0.184 | 0.628 | 1.000 | 56 | tags=65%, list=44%, signal=98% |
| 153 | GOBP\_FOREBRAIN\_DEVELOPMENT |  | 19 | -0.26 | -1.28 | 0.188 | 0.625 | 1.000 | 79 | tags=84%, list=63%, signal=192% |
| 154 | HP\_OCULAR\_ANTERIOR\_SEGMENT\_DYSGENESIS |  | 14 | -0.30 | -1.28 | 0.191 | 0.625 | 1.000 | 39 | tags=57%, list=31%, signal=74% |
| 155 | HP\_CRANIOSYNOSTOSIS |  | 19 | -0.26 | -1.27 | 0.182 | 0.629 | 1.000 | 26 | tags=42%, list=21%, signal=45% |
| 156 | HP\_ABNORMALITY\_OF\_THE\_UTERUS |  | 36 | -0.21 | -1.27 | 0.150 | 0.626 | 1.000 | 73 | tags=72%, list=58%, signal=123% |
| 157 | HP\_ABNORMAL\_UVEA\_MORPHOLOGY |  | 22 | -0.25 | -1.27 | 0.183 | 0.631 | 1.000 | 44 | tags=55%, list=35%, signal=69% |
| 158 | GOBP\_REGULATION\_OF\_HYDROLASE\_ACTIVITY |  | 15 | -0.28 | -1.26 | 0.183 | 0.641 | 1.000 | 45 | tags=60%, list=36%, signal=82% |
| 159 | KDM5D\_TARGET\_GENES |  | 10 | -0.34 | -1.25 | 0.198 | 0.655 | 1.000 | 75 | tags=90%, list=60%, signal=205% |
| 160 | GOBP\_POSITIVE\_REGULATION\_OF\_TRANSCRIPTION\_BY\_RNA\_POLYMERASE\_II |  | 25 | -0.23 | -1.25 | 0.197 | 0.668 | 1.000 | 23 | tags=36%, list=18%, signal=35% |
| 161 | HP\_ABNORMALITY\_OF\_THE\_VASCULATURE\_OF\_THE\_EYE |  | 11 | -0.32 | -1.24 | 0.195 | 0.670 | 1.000 | 90 | tags=100%, list=71%, signal=319% |
| 162 | GOBP\_REGULATION\_OF\_WNT\_SIGNALING\_PATHWAY |  | 11 | -0.31 | -1.24 | 0.209 | 0.667 | 1.000 | 68 | tags=82%, list=54%, signal=162% |
| 163 | GOBP\_MESENCHYMAL\_CELL\_DIFFERENTIATION |  | 13 | -0.30 | -1.24 | 0.184 | 0.663 | 1.000 | 74 | tags=85%, list=59%, signal=184% |
| 164 | HP\_ABNORMAL\_URETER\_MORPHOLOGY |  | 17 | -0.26 | -1.24 | 0.209 | 0.662 | 1.000 | 39 | tags=53%, list=31%, signal=66% |
| 165 | HP\_ABNORMAL\_ESOPHAGUS\_MORPHOLOGY |  | 10 | -0.33 | -1.24 | 0.197 | 0.658 | 1.000 | 76 | tags=90%, list=60%, signal=209% |
| 166 | HP\_OMPHALOCELE |  | 16 | -0.28 | -1.24 | 0.217 | 0.660 | 1.000 | 73 | tags=81%, list=58%, signal=169% |
| 167 | HP\_INTRAUTERINE\_GROWTH\_RETARDATION |  | 30 | -0.22 | -1.24 | 0.210 | 0.660 | 1.000 | 39 | tags=47%, list=31%, signal=51% |
| 168 | GOBP\_NEGATIVE\_REGULATION\_OF\_RESPONSE\_TO\_STIMULUS |  | 24 | -0.23 | -1.24 | 0.208 | 0.657 | 1.000 | 93 | tags=92%, list=74%, signal=283% |
| 169 | HP\_ANORECTAL\_ANOMALY |  | 32 | -0.21 | -1.23 | 0.215 | 0.664 | 1.000 | 56 | tags=59%, list=44%, signal=80% |
| 170 | HP\_ABNORMALITY\_OF\_UPPER\_LIP |  | 51 | -0.19 | -1.23 | 0.206 | 0.661 | 1.000 | 76 | tags=71%, list=60%, signal=106% |
| 171 | HP\_FINGER\_CLINODACTYLY |  | 34 | -0.20 | -1.23 | 0.206 | 0.659 | 1.000 | 38 | tags=44%, list=30%, signal=46% |
| 172 | GOBP\_POSITIVE\_REGULATION\_OF\_CELLULAR\_BIOSYNTHETIC\_PROCESS |  | 34 | -0.20 | -1.23 | 0.204 | 0.658 | 1.000 | 23 | tags=32%, list=18%, signal=29% |
| 173 | HP\_ABNORMALITY\_OF\_THE\_ANUS |  | 32 | -0.21 | -1.23 | 0.207 | 0.656 | 1.000 | 56 | tags=59%, list=44%, signal=80% |
| 174 | HP\_ABNORMALITY\_OF\_THE\_PALPEBRAL\_FISSURES |  | 60 | -0.18 | -1.23 | 0.212 | 0.654 | 1.000 | 27 | tags=30%, list=21%, signal=20% |
| 175 | IRF9\_TARGET\_GENES |  | 11 | -0.32 | -1.23 | 0.228 | 0.654 | 1.000 | 67 | tags=82%, list=53%, signal=159% |
| 176 | HP\_GASTROINTESTINAL\_ATRESIA |  | 10 | -0.33 | -1.22 | 0.232 | 0.673 | 1.000 | 76 | tags=90%, list=60%, signal=209% |
| 177 | HP\_NAIL\_DYSPLASIA |  | 16 | -0.27 | -1.21 | 0.227 | 0.676 | 1.000 | 74 | tags=81%, list=59%, signal=172% |
| 178 | GOBP\_MORPHOGENESIS\_OF\_EMBRYONIC\_EPITHELIUM |  | 11 | -0.31 | -1.21 | 0.234 | 0.674 | 1.000 | 68 | tags=82%, list=54%, signal=162% |
| 179 | GOBP\_SENSORY\_PERCEPTION |  | 11 | -0.31 | -1.21 | 0.220 | 0.670 | 1.000 | 80 | tags=91%, list=63%, signal=227% |
| 180 | HP\_ORAL\_CLEFT |  | 56 | -0.18 | -1.21 | 0.227 | 0.684 | 1.000 | 76 | tags=70%, list=60%, signal=98% |
| 181 | GOBP\_REGULATION\_OF\_ANATOMICAL\_STRUCTURE\_MORPHOGENESIS |  | 15 | -0.27 | -1.20 | 0.226 | 0.689 | 1.000 | 72 | tags=80%, list=57%, signal=164% |
| 182 | HP\_MEDIAL\_FLARING\_OF\_THE\_EYEBROW |  | 12 | -0.30 | -1.20 | 0.216 | 0.691 | 1.000 | 93 | tags=100%, list=74%, signal=345% |
| 183 | GOBP\_ENDOCRINE\_SYSTEM\_DEVELOPMENT |  | 10 | -0.33 | -1.20 | 0.242 | 0.692 | 1.000 | 76 | tags=90%, list=60%, signal=209% |
| 184 | OSMAN\_BLOOD\_CHAD63\_KH\_AGE\_18\_50YO\_HIGH\_DOSE\_SUBJECTS\_24HR\_DN |  | 17 | -0.25 | -1.20 | 0.230 | 0.689 | 1.000 | 85 | tags=88%, list=67%, signal=235% |
| 185 | HP\_ABNORMALITY\_OF\_FINGER |  | 71 | -0.18 | -1.19 | 0.242 | 0.691 | 1.000 | 80 | tags=70%, list=63%, signal=84% |
| 186 | HP\_POSITIONAL\_FOOT\_DEFORMITY |  | 30 | -0.20 | -1.19 | 0.223 | 0.690 | 1.000 | 53 | tags=57%, list=42%, signal=75% |
| 187 | HP\_ABNORMAL\_INTERNAL\_GENITALIA |  | 76 | -0.18 | -1.18 | 0.235 | 0.708 | 1.000 | 80 | tags=70%, list=63%, signal=76% |
| 188 | GOBP\_EPITHELIUM\_DEVELOPMENT |  | 37 | -0.19 | -1.18 | 0.235 | 0.710 | 1.000 | 69 | tags=68%, list=55%, signal=106% |
| 189 | GOBP\_TUBE\_DEVELOPMENT |  | 28 | -0.21 | -1.18 | 0.231 | 0.717 | 1.000 | 39 | tags=46%, list=31%, signal=52% |
| 190 | HP\_ABNORMAL\_THUMB\_MORPHOLOGY |  | 29 | -0.20 | -1.17 | 0.255 | 0.725 | 1.000 | 73 | tags=72%, list=58%, signal=133% |
| 191 | GOBP\_REGULATION\_OF\_GROWTH |  | 13 | -0.27 | -1.17 | 0.245 | 0.722 | 1.000 | 96 | tags=100%, list=76%, signal=377% |
| 192 | HP\_APLASIA\_HYPOPLASIA\_INVOLVING\_BONES\_OF\_THE\_LOWER\_LIMBS |  | 28 | -0.21 | -1.17 | 0.261 | 0.735 | 1.000 | 75 | tags=75%, list=60%, signal=144% |
| 193 | CUI\_TCF21\_TARGETS\_2\_UP |  | 10 | -0.31 | -1.17 | 0.251 | 0.734 | 1.000 | 66 | tags=80%, list=52%, signal=155% |
| 194 | HP\_SMALL\_FOR\_GESTATIONAL\_AGE |  | 15 | -0.27 | -1.16 | 0.260 | 0.732 | 1.000 | 97 | tags=100%, list=77%, signal=383% |
| 195 | HP\_PREAXIAL\_POLYDACTYLY |  | 12 | -0.29 | -1.16 | 0.274 | 0.747 | 1.000 | 73 | tags=83%, list=58%, signal=179% |
| 196 | HP\_ABNORMAL\_UTERUS\_MORPHOLOGY |  | 15 | -0.26 | -1.16 | 0.257 | 0.744 | 1.000 | 73 | tags=80%, list=58%, signal=168% |
| 197 | GOBP\_CELL\_SURFACE\_RECEPTOR\_SIGNALING\_PATHWAY\_INVOLVED\_IN\_CELL\_CELL\_SIGNALING |  | 16 | -0.25 | -1.15 | 0.269 | 0.757 | 1.000 | 68 | tags=75%, list=54%, signal=142% |
| 198 | HP\_ABNORMALITY\_OF\_ADRENAL\_MORPHOLOGY |  | 13 | -0.28 | -1.15 | 0.294 | 0.761 | 1.000 | 76 | tags=85%, list=60%, signal=191% |
| 199 | GOBP\_CONNECTIVE\_TISSUE\_DEVELOPMENT |  | 11 | -0.29 | -1.15 | 0.279 | 0.757 | 1.000 | 36 | tags=55%, list=29%, signal=70% |
| 200 | GOBP\_EAR\_DEVELOPMENT |  | 13 | -0.28 | -1.15 | 0.273 | 0.754 | 1.000 | 76 | tags=85%, list=60%, signal=191% |
| 201 | HP\_ABNORMAL\_ANATOMIC\_LOCATION\_OF\_THE\_HEART |  | 12 | -0.28 | -1.15 | 0.270 | 0.752 | 1.000 | 53 | tags=67%, list=42%, signal=104% |
| 202 | HP\_GENERALIZED\_HIRSUTISM |  | 16 | -0.26 | -1.15 | 0.286 | 0.749 | 1.000 | 83 | tags=88%, list=66%, signal=224% |
| 203 | GOBP\_NEGATIVE\_REGULATION\_OF\_MOLECULAR\_FUNCTION |  | 13 | -0.27 | -1.14 | 0.274 | 0.750 | 1.000 | 38 | tags=54%, list=30%, signal=69% |
| 204 | HP\_ABNORMALITY\_OF\_PELVIC\_GIRDLE\_BONE\_MORPHOLOGY |  | 33 | -0.19 | -1.14 | 0.284 | 0.748 | 1.000 | 37 | tags=42%, list=29%, signal=44% |
| 205 | MANNO\_MIDBRAIN\_NEUROTYPES\_HRGL3 |  | 10 | -0.31 | -1.14 | 0.278 | 0.752 | 1.000 | 28 | tags=50%, list=22%, signal=59% |
| 206 | GOBP\_POSITIVE\_REGULATION\_OF\_PROTEIN\_MODIFICATION\_PROCESS |  | 15 | -0.26 | -1.14 | 0.290 | 0.753 | 1.000 | 90 | tags=93%, list=71%, signal=288% |
| 207 | GOBP\_NEGATIVE\_REGULATION\_OF\_SIGNALING |  | 19 | -0.23 | -1.14 | 0.304 | 0.753 | 1.000 | 89 | tags=89%, list=71%, signal=259% |
| 208 | HP\_BICORNUATE\_UTERUS |  | 11 | -0.30 | -1.14 | 0.305 | 0.749 | 1.000 | 70 | tags=82%, list=56%, signal=168% |
| 209 | DODD\_NASOPHARYNGEAL\_CARCINOMA\_DN |  | 11 | -0.30 | -1.13 | 0.285 | 0.748 | 1.000 | 93 | tags=100%, list=74%, signal=348% |
| 210 | WTGAAAT\_UNKNOWN |  | 11 | -0.29 | -1.13 | 0.323 | 0.750 | 1.000 | 82 | tags=91%, list=65%, signal=238% |
| 211 | HP\_ABNORMALITY\_OF\_THE\_LENS |  | 26 | -0.21 | -1.13 | 0.283 | 0.756 | 1.000 | 87 | tags=85%, list=69%, signal=217% |
| 212 | REACTOME\_BBSOME\_MEDIATED\_CARGO\_TARGETING\_TO\_CILIUM |  | 10 | -0.30 | -1.13 | 0.281 | 0.754 | 1.000 | 80 | tags=90%, list=63%, signal=227% |
| 213 | REACTOME\_CARGO\_TRAFFICKING\_TO\_THE\_PERICILIARY\_MEMBRANE |  | 10 | -0.30 | -1.13 | 0.279 | 0.754 | 1.000 | 80 | tags=90%, list=63%, signal=227% |
| 214 | GOBP\_TISSUE\_DEVELOPMENT |  | 52 | -0.17 | -1.13 | 0.293 | 0.751 | 1.000 | 39 | tags=40%, list=31%, signal=34% |
| 215 | HP\_ABNORMALITY\_OF\_THE\_ELBOW |  | 23 | -0.21 | -1.12 | 0.302 | 0.754 | 1.000 | 45 | tags=52%, list=36%, signal=66% |
| 216 | HP\_ABNORMALITY\_OF\_THE\_CLITORIS |  | 25 | -0.21 | -1.12 | 0.282 | 0.755 | 1.000 | 71 | tags=72%, list=56%, signal=132% |
| 217 | HP\_MICROMELIA |  | 16 | -0.24 | -1.12 | 0.312 | 0.758 | 1.000 | 38 | tags=50%, list=30%, signal=63% |
| 218 | HP\_ABNORMAL\_CORNEA\_MORPHOLOGY |  | 31 | -0.20 | -1.12 | 0.310 | 0.757 | 1.000 | 80 | tags=77%, list=63%, signal=160% |
| 219 | MIR6867\_5P |  | 12 | -0.28 | -1.12 | 0.289 | 0.756 | 1.000 | 22 | tags=42%, list=17%, signal=46% |
| 220 | GOBP\_CYTOKINE\_MEDIATED\_SIGNALING\_PATHWAY |  | 10 | -0.30 | -1.11 | 0.319 | 0.761 | 1.000 | 29 | tags=50%, list=23%, signal=60% |
| 221 | GOMF\_CIS\_REGULATORY\_REGION\_SEQUENCE\_SPECIFIC\_DNA\_BINDING |  | 19 | -0.23 | -1.11 | 0.300 | 0.762 | 1.000 | 23 | tags=37%, list=18%, signal=38% |
| 222 | GOBP\_NERVOUS\_SYSTEM\_PROCESS |  | 14 | -0.26 | -1.11 | 0.300 | 0.759 | 1.000 | 80 | tags=86%, list=63%, signal=209% |
| 223 | HP\_ABNORMALITY\_OF\_THE\_MIDDLE\_EAR |  | 27 | -0.20 | -1.11 | 0.318 | 0.760 | 1.000 | 79 | tags=78%, list=63%, signal=164% |
| 224 | GOBP\_PROSTATE\_GLAND\_DEVELOPMENT |  | 10 | -0.29 | -1.11 | 0.330 | 0.757 | 1.000 | 68 | tags=80%, list=54%, signal=160% |
| 225 | HP\_SPINA\_BIFIDA |  | 10 | -0.30 | -1.11 | 0.298 | 0.757 | 1.000 | 54 | tags=70%, list=43%, signal=113% |
| 226 | GOBP\_TUBE\_MORPHOGENESIS |  | 25 | -0.20 | -1.11 | 0.324 | 0.757 | 1.000 | 31 | tags=40%, list=25%, signal=43% |
| 227 | HP\_ABNORMALITY\_OF\_THE\_MIDFACE |  | 39 | -0.17 | -1.11 | 0.330 | 0.755 | 1.000 | 44 | tags=46%, list=35%, signal=49% |
| 228 | GOBP\_NEURON\_DIFFERENTIATION |  | 29 | -0.19 | -1.11 | 0.288 | 0.752 | 1.000 | 39 | tags=45%, list=31%, signal=50% |
| 229 | HP\_ABNORMAL\_NEURAL\_TUBE\_MORPHOLOGY |  | 12 | -0.27 | -1.11 | 0.308 | 0.749 | 1.000 | 54 | tags=67%, list=43%, signal=106% |
| 230 | GOMF\_SEQUENCE\_SPECIFIC\_DNA\_BINDING |  | 23 | -0.21 | -1.10 | 0.338 | 0.756 | 1.000 | 23 | tags=35%, list=18%, signal=35% |
| 231 | GOBP\_POSITIVE\_REGULATION\_OF\_NUCLEOBASE\_CONTAINING\_COMPOUND\_METABOLIC\_PROCESS |  | 32 | -0.18 | -1.10 | 0.327 | 0.755 | 1.000 | 23 | tags=31%, list=18%, signal=29% |
| 232 | GRYDER\_PAX3FOXO1\_ENHANCERS\_IN\_TADS |  | 16 | -0.25 | -1.10 | 0.323 | 0.754 | 1.000 | 29 | tags=44%, list=23%, signal=50% |
| 233 | HP\_ABNORMALITY\_OF\_THE\_SPINAL\_CORD |  | 12 | -0.27 | -1.10 | 0.334 | 0.751 | 1.000 | 54 | tags=67%, list=43%, signal=106% |
| 234 | HP\_ABNORMALITY\_OF\_THE\_PALM |  | 30 | -0.19 | -1.10 | 0.331 | 0.750 | 1.000 | 75 | tags=73%, list=60%, signal=138% |
| 235 | HP\_APLASIA\_HYPOPLASIA\_OF\_THE\_EAR |  | 16 | -0.24 | -1.10 | 0.321 | 0.747 | 1.000 | 30 | tags=44%, list=24%, signal=50% |
| 236 | HP\_OPTIC\_ATROPHY |  | 19 | -0.22 | -1.09 | 0.331 | 0.756 | 1.000 | 70 | tags=74%, list=56%, signal=141% |
| 237 | HP\_ABNORMAL\_LACRIMAL\_DUCT\_MORPHOLOGY |  | 10 | -0.29 | -1.09 | 0.331 | 0.753 | 1.000 | 68 | tags=80%, list=54%, signal=160% |
| 238 | HP\_ABNORMAL\_VISUAL\_ELECTROPHYSIOLOGY |  | 15 | -0.25 | -1.09 | 0.312 | 0.750 | 1.000 | 83 | tags=87%, list=66%, signal=224% |
| 239 | HP\_ABNORMALITY\_OF\_THE\_DIENCEPHALON |  | 11 | -0.28 | -1.09 | 0.326 | 0.749 | 1.000 | 37 | tags=55%, list=29%, signal=70% |
| 240 | HP\_ABNORMAL\_ELECTRORETINOGRAM |  | 15 | -0.25 | -1.09 | 0.327 | 0.749 | 1.000 | 83 | tags=87%, list=66%, signal=224% |
| 241 | GOBP\_EMBRYONIC\_ORGAN\_MORPHOGENESIS |  | 17 | -0.23 | -1.09 | 0.357 | 0.747 | 1.000 | 94 | tags=94%, list=75%, signal=321% |
| 242 | HP\_ABNORMAL\_METACARPAL\_MORPHOLOGY |  | 19 | -0.22 | -1.09 | 0.362 | 0.744 | 1.000 | 37 | tags=47%, list=29%, signal=57% |
| 243 | HP\_ABNORMAL\_HAND\_MORPHOLOGY |  | 28 | -0.20 | -1.09 | 0.336 | 0.741 | 1.000 | 76 | tags=75%, list=60%, signal=147% |
| 244 | HP\_APLASIA\_HYPOPLASIA\_OF\_TOE |  | 18 | -0.22 | -1.09 | 0.313 | 0.743 | 1.000 | 75 | tags=78%, list=60%, signal=165% |
| 245 | REACTOME\_ORGANELLE\_BIOGENESIS\_AND\_MAINTENANCE |  | 17 | -0.23 | -1.09 | 0.326 | 0.745 | 1.000 | 80 | tags=82%, list=63%, signal=195% |
| 246 | HP\_ABNORMALITY\_OF\_THE\_FEMALE\_GENITALIA |  | 85 | -0.17 | -1.09 | 0.331 | 0.743 | 1.000 | 80 | tags=68%, list=63%, signal=61% |
| 247 | GOBP\_FAT\_CELL\_DIFFERENTIATION |  | 11 | -0.27 | -1.08 | 0.337 | 0.741 | 1.000 | 96 | tags=100%, list=76%, signal=383% |
| 248 | HP\_ABNORMALITY\_OF\_DENTAL\_STRUCTURE |  | 21 | -0.21 | -1.08 | 0.323 | 0.743 | 1.000 | 75 | tags=76%, list=60%, signal=157% |
| 249 | HP\_ABNORMALITY\_OF\_FACIAL\_SOFT\_TISSUE |  | 15 | -0.25 | -1.08 | 0.335 | 0.740 | 1.000 | 83 | tags=87%, list=66%, signal=224% |
| 250 | HP\_ABNORMAL\_LABIA\_MORPHOLOGY |  | 20 | -0.22 | -1.08 | 0.331 | 0.738 | 1.000 | 22 | tags=35%, list=17%, signal=36% |
| 251 | HP\_ABNORMALITY\_OF\_THE\_LARYNX |  | 27 | -0.20 | -1.08 | 0.352 | 0.739 | 1.000 | 75 | tags=74%, list=60%, signal=144% |
| 252 | HP\_ABNORMAL\_THORAX\_MORPHOLOGY |  | 48 | -0.16 | -1.07 | 0.339 | 0.750 | 1.000 | 75 | tags=69%, list=60%, signal=105% |
| 253 | HP\_ABNORMALITY\_OF\_THE\_METACARPAL\_BONES |  | 25 | -0.19 | -1.07 | 0.339 | 0.749 | 1.000 | 37 | tags=44%, list=29%, signal=50% |
| 254 | LEE\_BMP2\_TARGETS\_UP |  | 10 | -0.28 | -1.07 | 0.341 | 0.748 | 1.000 | 31 | tags=50%, list=25%, signal=61% |
| 255 | NABA\_MATRISOME |  | 10 | -0.29 | -1.07 | 0.358 | 0.747 | 1.000 | 68 | tags=80%, list=54%, signal=160% |
| 256 | HP\_ABNORMALITY\_OF\_THE\_RIBS |  | 24 | -0.20 | -1.07 | 0.346 | 0.747 | 1.000 | 75 | tags=75%, list=60%, signal=150% |
| 257 | HP\_FEEDING\_DIFFICULTIES\_IN\_INFANCY |  | 29 | -0.18 | -1.07 | 0.343 | 0.748 | 1.000 | 70 | tags=69%, list=56%, signal=119% |
| 258 | ZSCAN30\_TARGET\_GENES |  | 16 | -0.23 | -1.06 | 0.362 | 0.758 | 1.000 | 23 | tags=38%, list=18%, signal=40% |
| 259 | HP\_ABNORMALITY\_OF\_THE\_LOWER\_URINARY\_TRACT |  | 70 | -0.16 | -1.06 | 0.349 | 0.761 | 1.000 | 48 | tags=44%, list=38%, signal=32% |
| 260 | HP\_ABNORMAL\_EYEBROW\_MORPHOLOGY |  | 38 | -0.18 | -1.06 | 0.375 | 0.760 | 1.000 | 85 | tags=79%, list=67%, signal=169% |
| 261 | HP\_ABNORMALITY\_OF\_THE\_KIDNEY |  | 77 | -0.16 | -1.06 | 0.350 | 0.760 | 1.000 | 39 | tags=36%, list=31%, signal=20% |
| 262 | HP\_ABNORMAL\_ANTERIOR\_EYE\_SEGMENT\_MORPHOLOGY |  | 42 | -0.17 | -1.06 | 0.338 | 0.760 | 1.000 | 80 | tags=74%, list=63%, signal=135% |
| 263 | GOMF\_DNA\_BINDING\_TRANSCRIPTION\_FACTOR\_BINDING |  | 23 | -0.20 | -1.05 | 0.359 | 0.766 | 1.000 | 35 | tags=43%, list=28%, signal=49% |
| 264 | GOMF\_TRANSCRIPTION\_FACTOR\_BINDING |  | 23 | -0.20 | -1.05 | 0.346 | 0.767 | 1.000 | 35 | tags=43%, list=28%, signal=49% |
| 265 | HP\_RESPIRATORY\_INSUFFICIENCY |  | 21 | -0.21 | -1.05 | 0.345 | 0.768 | 1.000 | 75 | tags=76%, list=60%, signal=157% |
| 266 | HP\_ANAL\_ATRESIA |  | 24 | -0.19 | -1.05 | 0.391 | 0.774 | 1.000 | 102 | tags=96%, list=81%, signal=407% |
| 267 | HP\_ABNORMAL\_RIB\_CAGE\_MORPHOLOGY |  | 32 | -0.18 | -1.04 | 0.391 | 0.783 | 1.000 | 75 | tags=72%, list=60%, signal=132% |
| 268 | GOBP\_DEVELOPMENTAL\_GROWTH |  | 29 | -0.18 | -1.04 | 0.384 | 0.784 | 1.000 | 27 | tags=34%, list=21%, signal=34% |
| 269 | HP\_ABNORMALITY\_OF\_EPIPHYSIS\_MORPHOLOGY |  | 21 | -0.20 | -1.04 | 0.344 | 0.783 | 1.000 | 52 | tags=57%, list=41%, signal=81% |
| 270 | TAATTA\_CHX10\_01 |  | 17 | -0.22 | -1.04 | 0.376 | 0.780 | 1.000 | 21 | tags=35%, list=17%, signal=37% |
| 271 | BRUINS\_UVC\_RESPONSE\_LATE |  | 12 | -0.25 | -1.03 | 0.385 | 0.785 | 1.000 | 88 | tags=92%, list=70%, signal=275% |
| 272 | HP\_ABNORMALITY\_OF\_THYROID\_PHYSIOLOGY |  | 19 | -0.21 | -1.03 | 0.382 | 0.783 | 1.000 | 38 | tags=47%, list=30%, signal=58% |
| 273 | MZF1\_TARGET\_GENES |  | 11 | -0.27 | -1.03 | 0.395 | 0.780 | 1.000 | 27 | tags=45%, list=21%, signal=53% |
| 274 | HP\_ABNORMALITY\_OF\_THE\_BILIARY\_SYSTEM |  | 13 | -0.25 | -1.03 | 0.411 | 0.783 | 1.000 | 89 | tags=92%, list=71%, signal=282% |
| 275 | HP\_ABNORMAL\_EYELASH\_MORPHOLOGY |  | 14 | -0.24 | -1.03 | 0.388 | 0.785 | 1.000 | 82 | tags=86%, list=65%, signal=218% |
| 276 | HP\_GLAUCOMA |  | 21 | -0.20 | -1.03 | 0.390 | 0.785 | 1.000 | 70 | tags=71%, list=56%, signal=134% |
| 277 | GOBP\_REGULATION\_OF\_DEVELOPMENTAL\_GROWTH |  | 11 | -0.27 | -1.03 | 0.397 | 0.785 | 1.000 | 96 | tags=100%, list=76%, signal=383% |
| 278 | HP\_ABNORMAL\_CORTICAL\_GYRATION |  | 11 | -0.27 | -1.03 | 0.388 | 0.783 | 1.000 | 62 | tags=73%, list=49%, signal=131% |
| 279 | HP\_ABNORMAL\_ODONTOID\_TISSUE\_MORPHOLOGY |  | 17 | -0.22 | -1.03 | 0.384 | 0.780 | 1.000 | 73 | tags=76%, list=58%, signal=157% |
| 280 | HP\_SHORT\_NOSE |  | 19 | -0.21 | -1.03 | 0.398 | 0.778 | 1.000 | 38 | tags=47%, list=30%, signal=58% |
| 281 | HP\_PROPORTIONATE\_SHORT\_STATURE |  | 13 | -0.25 | -1.03 | 0.395 | 0.775 | 1.000 | 41 | tags=54%, list=33%, signal=72% |
| 282 | HP\_ABNORMAL\_URETER\_PHYSIOLOGY |  | 17 | -0.22 | -1.02 | 0.405 | 0.775 | 1.000 | 22 | tags=35%, list=17%, signal=37% |
| 283 | GGGTGGRR\_PAX4\_03 |  | 16 | -0.22 | -1.02 | 0.426 | 0.787 | 1.000 | 24 | tags=38%, list=19%, signal=40% |
| 284 | GOBP\_CELL\_CELL\_SIGNALING\_BY\_WNT |  | 15 | -0.23 | -1.02 | 0.419 | 0.788 | 1.000 | 76 | tags=80%, list=60%, signal=178% |
| 285 | HP\_IRIS\_COLOBOMA |  | 13 | -0.25 | -1.02 | 0.413 | 0.786 | 1.000 | 70 | tags=77%, list=56%, signal=155% |
| 286 | MIR6809\_3P |  | 10 | -0.27 | -1.01 | 0.431 | 0.788 | 1.000 | 33 | tags=50%, list=26%, signal=62% |
| 287 | HP\_HYDROURETER |  | 10 | -0.28 | -1.01 | 0.449 | 0.786 | 1.000 | 32 | tags=50%, list=25%, signal=62% |
| 288 | HP\_COGNITIVE\_IMPAIRMENT |  | 17 | -0.22 | -1.01 | 0.421 | 0.783 | 1.000 | 44 | tags=53%, list=35%, signal=70% |
| 289 | TRAVAGLINI\_LUNG\_PROXIMAL\_CILIATED\_CELL |  | 13 | -0.24 | -1.01 | 0.416 | 0.782 | 1.000 | 80 | tags=85%, list=63%, signal=208% |
| 290 | ZBTB7B\_TARGET\_GENES |  | 23 | -0.19 | -1.01 | 0.403 | 0.784 | 1.000 | 80 | tags=78%, list=63%, signal=175% |
| 291 | HP\_ABNORMALITY\_OF\_EARLOBE |  | 15 | -0.23 | -1.01 | 0.417 | 0.784 | 1.000 | 26 | tags=40%, list=21%, signal=44% |
| 292 | GOBP\_GROWTH |  | 32 | -0.17 | -1.01 | 0.456 | 0.785 | 1.000 | 28 | tags=34%, list=22%, signal=33% |
| 293 | HP\_ABNORMAL\_NASOLACRIMAL\_SYSTEM\_MORPHOLOGY |  | 15 | -0.23 | -1.00 | 0.435 | 0.793 | 1.000 | 68 | tags=73%, list=54%, signal=140% |
| 294 | HP\_ABNORMALITY\_OF\_THE\_CERVICAL\_SPINE |  | 35 | -0.16 | -1.00 | 0.431 | 0.793 | 1.000 | 94 | tags=86%, list=75%, signal=244% |
| 295 | HP\_APLASIA\_HYPOPLASIA\_OF\_THE\_RADIUS |  | 10 | -0.27 | -1.00 | 0.442 | 0.791 | 1.000 | 70 | tags=80%, list=56%, signal=166% |
| 296 | HP\_DEVIATION\_OF\_THE\_HAND\_OR\_OF\_FINGERS\_OF\_THE\_HAND |  | 42 | -0.15 | -1.00 | 0.378 | 0.788 | 1.000 | 39 | tags=40%, list=31%, signal=39% |
| 297 | HP\_ADVANCED\_ERUPTION\_OF\_TEETH |  | 11 | -0.25 | -1.00 | 0.453 | 0.799 | 1.000 | 75 | tags=82%, list=60%, signal=184% |
| 298 | LHX9\_TARGET\_GENES |  | 12 | -0.25 | -1.00 | 0.436 | 0.796 | 1.000 | 99 | tags=100%, list=79%, signal=422% |
| 299 | HP\_ABNORMALITY\_OF\_THE\_AMNIOTIC\_FLUID |  | 27 | -0.18 | -0.99 | 0.434 | 0.797 | 1.000 | 39 | tags=44%, list=31%, signal=51% |
| 300 | GOBP\_NEUROGENESIS |  | 36 | -0.17 | -0.99 | 0.438 | 0.797 | 1.000 | 77 | tags=72%, list=61%, signal=133% |
| 301 | HP\_BRACHYDACTYLY |  | 38 | -0.16 | -0.99 | 0.456 | 0.794 | 1.000 | 37 | tags=39%, list=29%, signal=39% |
| 302 | HP\_PROMINENT\_NASAL\_BRIDGE |  | 23 | -0.19 | -0.99 | 0.433 | 0.799 | 1.000 | 102 | tags=96%, list=81%, signal=411% |
| 303 | HP\_ABNORMAL\_VENTRICULAR\_SEPTUM\_MORPHOLOGY |  | 34 | -0.17 | -0.99 | 0.434 | 0.801 | 1.000 | 45 | tags=47%, list=36%, signal=53% |
| 304 | GOMF\_RNA\_POLYMERASE\_II\_SPECIFIC\_DNA\_BINDING\_TRANSCRIPTION\_FACTOR\_BINDING |  | 18 | -0.20 | -0.99 | 0.439 | 0.799 | 1.000 | 35 | tags=44%, list=28%, signal=53% |
| 305 | HP\_ANTERIORLY\_PLACED\_ANUS |  | 13 | -0.23 | -0.99 | 0.469 | 0.802 | 1.000 | 23 | tags=38%, list=18%, signal=42% |
| 306 | GOCC\_CHROMOSOME |  | 32 | -0.16 | -0.98 | 0.485 | 0.800 | 1.000 | 21 | tags=28%, list=17%, signal=25% |
| 307 | HP\_ABNORMALITY\_OF\_THE\_INCISOR |  | 15 | -0.22 | -0.98 | 0.448 | 0.809 | 1.000 | 69 | tags=73%, list=55%, signal=143% |
| 308 | GOBP\_LIPID\_BIOSYNTHETIC\_PROCESS |  | 13 | -0.24 | -0.98 | 0.475 | 0.808 | 1.000 | 71 | tags=77%, list=56%, signal=158% |
| 309 | GOBP\_PROGRAMMED\_CELL\_DEATH |  | 26 | -0.18 | -0.98 | 0.448 | 0.807 | 1.000 | 75 | tags=73%, list=60%, signal=143% |
| 310 | GOBP\_REGULATION\_OF\_CELL\_DEATH |  | 23 | -0.19 | -0.97 | 0.453 | 0.819 | 1.000 | 75 | tags=74%, list=60%, signal=149% |
| 311 | HP\_DISPROPORTIONATE\_SHORT\_LIMB\_SHORT\_STATURE |  | 12 | -0.24 | -0.97 | 0.490 | 0.817 | 1.000 | 37 | tags=50%, list=29%, signal=64% |
| 312 | HP\_ABNORMALITY\_OF\_THE\_URINARY\_SYSTEM\_PHYSIOLOGY |  | 42 | -0.15 | -0.97 | 0.441 | 0.819 | 1.000 | 87 | tags=79%, list=69%, signal=169% |
| 313 | GOBP\_ANATOMICAL\_STRUCTURE\_FORMATION\_INVOLVED\_IN\_MORPHOGENESIS |  | 22 | -0.18 | -0.97 | 0.483 | 0.817 | 1.000 | 68 | tags=68%, list=54%, signal=122% |
| 314 | HP\_SHORT\_PALM |  | 14 | -0.22 | -0.97 | 0.448 | 0.817 | 1.000 | 75 | tags=79%, list=60%, signal=173% |
| 315 | HP\_APLASIA\_HYPOPLASIA\_INVOLVING\_THE\_SKELETON |  | 63 | -0.14 | -0.97 | 0.425 | 0.817 | 1.000 | 76 | tags=67%, list=60%, signal=84% |
| 316 | HP\_EXTERNAL\_GENITAL\_HYPOPLASIA |  | 76 | -0.14 | -0.96 | 0.490 | 0.817 | 1.000 | 80 | tags=68%, list=63%, signal=74% |
| 317 | HP\_SHORT\_DIGIT |  | 48 | -0.15 | -0.96 | 0.464 | 0.821 | 1.000 | 76 | tags=69%, list=60%, signal=107% |
| 318 | GOCC\_CHROMATIN |  | 27 | -0.18 | -0.96 | 0.487 | 0.828 | 1.000 | 21 | tags=30%, list=17%, signal=28% |
| 319 | GOBP\_CELL\_PROJECTION\_ORGANIZATION |  | 34 | -0.16 | -0.96 | 0.497 | 0.829 | 1.000 | 83 | tags=76%, list=66%, signal=164% |
| 320 | HP\_AGENESIS\_OF\_CORPUS\_CALLOSUM |  | 26 | -0.17 | -0.95 | 0.510 | 0.829 | 1.000 | 76 | tags=73%, list=60%, signal=146% |
| 321 | TTANTCA\_UNKNOWN |  | 15 | -0.22 | -0.95 | 0.476 | 0.827 | 1.000 | 69 | tags=73%, list=55%, signal=143% |
| 322 | GOBP\_NEURON\_DEVELOPMENT |  | 22 | -0.19 | -0.95 | 0.525 | 0.833 | 1.000 | 39 | tags=45%, list=31%, signal=54% |
| 323 | GAO\_LARGE\_INTESTINE\_ADULT\_CJ\_IMMUNE\_CELLS |  | 12 | -0.24 | -0.95 | 0.511 | 0.838 | 1.000 | 37 | tags=50%, list=29%, signal=64% |
| 324 | HP\_ABNORMALITY\_OF\_THE\_PHALANGES\_OF\_THE\_TOES |  | 14 | -0.22 | -0.94 | 0.487 | 0.844 | 1.000 | 75 | tags=79%, list=60%, signal=173% |
| 325 | HP\_DECREASED\_HEAD\_CIRCUMFERENCE |  | 40 | -0.15 | -0.94 | 0.495 | 0.843 | 1.000 | 92 | tags=83%, list=73%, signal=209% |
| 326 | HP\_CEREBELLAR\_MALFORMATION |  | 22 | -0.19 | -0.94 | 0.494 | 0.841 | 1.000 | 39 | tags=45%, list=31%, signal=54% |
| 327 | GOBP\_CILIUM\_ORGANIZATION |  | 19 | -0.19 | -0.94 | 0.522 | 0.841 | 1.000 | 80 | tags=79%, list=63%, signal=184% |
| 328 | HP\_HERNIA |  | 40 | -0.15 | -0.94 | 0.511 | 0.846 | 1.000 | 76 | tags=70%, list=60%, signal=120% |
| 329 | HP\_JOINT\_DISLOCATION |  | 15 | -0.21 | -0.93 | 0.513 | 0.856 | 1.000 | 37 | tags=47%, list=29%, signal=58% |
| 330 | HP\_ABNORMAL\_LOWER\_LIMB\_BONE\_MORPHOLOGY |  | 49 | -0.14 | -0.93 | 0.535 | 0.856 | 1.000 | 80 | tags=71%, list=63%, signal=120% |
| 331 | HP\_ABNORMAL\_MYOCARDIUM\_MORPHOLOGY |  | 16 | -0.20 | -0.93 | 0.556 | 0.857 | 1.000 | 89 | tags=88%, list=71%, signal=260% |
| 332 | HP\_ABNORMAL\_LUNG\_DEVELOPMENT |  | 25 | -0.17 | -0.93 | 0.542 | 0.856 | 1.000 | 54 | tags=56%, list=43%, signal=79% |
| 333 | HP\_DEVIATION\_OF\_TOES |  | 13 | -0.23 | -0.93 | 0.512 | 0.857 | 1.000 | 24 | tags=38%, list=19%, signal=43% |
| 334 | GOBP\_NEGATIVE\_REGULATION\_OF\_CELL\_POPULATION\_PROLIFERATION |  | 13 | -0.22 | -0.92 | 0.521 | 0.868 | 1.000 | 83 | tags=85%, list=66%, signal=222% |
| 335 | HP\_ABNORMAL\_UMBILICUS\_MORPHOLOGY |  | 26 | -0.17 | -0.92 | 0.517 | 0.867 | 1.000 | 23 | tags=31%, list=18%, signal=30% |
| 336 | GOBP\_MUSCLE\_STRUCTURE\_DEVELOPMENT |  | 13 | -0.22 | -0.92 | 0.565 | 0.868 | 1.000 | 83 | tags=85%, list=66%, signal=222% |
| 337 | GOBP\_HEART\_DEVELOPMENT |  | 23 | -0.17 | -0.91 | 0.546 | 0.872 | 1.000 | 27 | tags=35%, list=21%, signal=36% |
| 338 | HP\_ABNORMALITY\_OF\_THE\_THYROID\_GLAND |  | 20 | -0.19 | -0.91 | 0.532 | 0.875 | 1.000 | 38 | tags=45%, list=30%, signal=54% |
| 339 | DODD\_NASOPHARYNGEAL\_CARCINOMA\_UP |  | 10 | -0.25 | -0.91 | 0.541 | 0.873 | 1.000 | 48 | tags=60%, list=38%, signal=89% |
| 340 | HP\_ABNORMAL\_FOREARM\_BONE\_MORPHOLOGY |  | 14 | -0.21 | -0.91 | 0.502 | 0.872 | 1.000 | 31 | tags=43%, list=25%, signal=51% |
| 341 | HP\_ECTRODACTYLY |  | 12 | -0.23 | -0.91 | 0.558 | 0.870 | 1.000 | 38 | tags=50%, list=30%, signal=65% |
| 342 | HP\_HYDROCEPHALUS |  | 20 | -0.18 | -0.91 | 0.549 | 0.869 | 1.000 | 70 | tags=70%, list=56%, signal=133% |
| 343 | GOBP\_POSITIVE\_REGULATION\_OF\_MAPK\_CASCADE |  | 11 | -0.23 | -0.91 | 0.554 | 0.869 | 1.000 | 101 | tags=100%, list=80%, signal=460% |
| 344 | GOBP\_CARTILAGE\_DEVELOPMENT |  | 10 | -0.24 | -0.91 | 0.590 | 0.868 | 1.000 | 23 | tags=40%, list=18%, signal=45% |
| 345 | HP\_ABNORMAL\_SIZE\_OF\_THE\_PALPEBRAL\_FISSURES |  | 21 | -0.18 | -0.91 | 0.524 | 0.868 | 1.000 | 12 | tags=24%, list=10%, signal=22% |
| 346 | MIR200B\_3P |  | 10 | -0.24 | -0.91 | 0.527 | 0.866 | 1.000 | 23 | tags=40%, list=18%, signal=45% |
| 347 | HP\_ABNORMALITY\_OF\_MUSCLE\_SIZE |  | 28 | -0.16 | -0.90 | 0.546 | 0.869 | 1.000 | 93 | tags=86%, list=74%, signal=255% |
| 348 | HP\_ABNORMALITY\_OF\_THE\_CEREBROSPINAL\_FLUID |  | 20 | -0.18 | -0.90 | 0.571 | 0.867 | 1.000 | 70 | tags=70%, list=56%, signal=133% |
| 349 | WP\_JOUBERT\_SYNDROME |  | 11 | -0.23 | -0.90 | 0.559 | 0.866 | 1.000 | 43 | tags=55%, list=34%, signal=76% |
| 350 | HP\_ABNORMALITY\_OF\_LIMB\_EPIPHYSIS\_MORPHOLOGY |  | 11 | -0.23 | -0.90 | 0.556 | 0.864 | 1.000 | 32 | tags=45%, list=25%, signal=56% |
| 351 | GOBP\_POSITIVE\_REGULATION\_OF\_DEVELOPMENTAL\_PROCESS |  | 24 | -0.17 | -0.90 | 0.568 | 0.863 | 1.000 | 68 | tags=67%, list=54%, signal=117% |
| 352 | MIR200C\_3P |  | 10 | -0.24 | -0.90 | 0.567 | 0.865 | 1.000 | 23 | tags=40%, list=18%, signal=45% |
| 353 | HP\_ABNORMALITY\_OF\_THE\_NASAL\_ALAE |  | 41 | -0.14 | -0.90 | 0.566 | 0.869 | 1.000 | 44 | tags=44%, list=35%, signal=46% |
| 354 | GOBP\_POSITIVE\_REGULATION\_OF\_RNA\_METABOLIC\_PROCESS |  | 31 | -0.15 | -0.90 | 0.570 | 0.868 | 1.000 | 23 | tags=29%, list=18%, signal=27% |
| 355 | GOBP\_REGULATION\_OF\_CELL\_DIFFERENTIATION |  | 26 | -0.16 | -0.89 | 0.582 | 0.881 | 1.000 | 38 | tags=42%, list=30%, signal=48% |
| 356 | HP\_BROAD\_FINGER |  | 10 | -0.23 | -0.89 | 0.592 | 0.882 | 1.000 | 37 | tags=50%, list=29%, signal=65% |
| 357 | HP\_APLASIA\_HYPOPLASIA\_INVOLVING\_THE\_METACARPAL\_BONES |  | 15 | -0.21 | -0.89 | 0.592 | 0.881 | 1.000 | 37 | tags=47%, list=29%, signal=58% |
| 358 | GOBP\_VASCULATURE\_DEVELOPMENT |  | 14 | -0.21 | -0.89 | 0.567 | 0.878 | 1.000 | 68 | tags=71%, list=54%, signal=138% |
| 359 | GOBP\_POSITIVE\_REGULATION\_OF\_CELL\_DIFFERENTIATION |  | 13 | -0.21 | -0.89 | 0.580 | 0.877 | 1.000 | 45 | tags=54%, list=36%, signal=75% |
| 360 | GOBP\_POSITIVE\_REGULATION\_OF\_MOLECULAR\_FUNCTION |  | 16 | -0.19 | -0.89 | 0.587 | 0.875 | 1.000 | 90 | tags=88%, list=71%, signal=267% |
| 361 | HP\_ABNORMAL\_TRACHEOBRONCHIAL\_MORPHOLOGY |  | 16 | -0.19 | -0.88 | 0.600 | 0.875 | 1.000 | 82 | tags=81%, list=65%, signal=203% |
| 362 | GOCC\_CILIARY\_BASAL\_BODY |  | 11 | -0.23 | -0.88 | 0.616 | 0.874 | 1.000 | 43 | tags=55%, list=34%, signal=76% |
| 363 | GOBP\_MESENCHYME\_DEVELOPMENT |  | 16 | -0.20 | -0.88 | 0.581 | 0.873 | 1.000 | 74 | tags=75%, list=59%, signal=159% |
| 364 | GOBP\_MULTICELLULAR\_ORGANISMAL\_HOMEOSTASIS |  | 13 | -0.22 | -0.88 | 0.591 | 0.876 | 1.000 | 93 | tags=92%, list=74%, signal=316% |
| 365 | HP\_NEOPLASM |  | 36 | -0.14 | -0.88 | 0.595 | 0.875 | 1.000 | 23 | tags=28%, list=18%, signal=24% |
| 366 | RTAAACA\_FREAC2\_01 |  | 11 | -0.23 | -0.88 | 0.592 | 0.874 | 1.000 | 101 | tags=100%, list=80%, signal=460% |
| 367 | GOBP\_CELL\_ACTIVATION |  | 14 | -0.21 | -0.88 | 0.578 | 0.874 | 1.000 | 23 | tags=36%, list=18%, signal=39% |
| 368 | HP\_AMBIGUOUS\_GENITALIA |  | 41 | -0.14 | -0.87 | 0.604 | 0.880 | 1.000 | 54 | tags=51%, list=43%, signal=60% |
| 369 | HP\_ABNORMALITY\_OF\_URINE\_HOMEOSTASIS |  | 10 | -0.24 | -0.87 | 0.600 | 0.881 | 1.000 | 87 | tags=90%, list=69%, signal=268% |
| 370 | HP\_ABNORMALITY\_OF\_FEMALE\_EXTERNAL\_GENITALIA |  | 40 | -0.14 | -0.87 | 0.616 | 0.881 | 1.000 | 71 | tags=65%, list=56%, signal=102% |
| 371 | HP\_ABNORMALITY\_OF\_THE\_ANTIHELIX |  | 10 | -0.23 | -0.87 | 0.602 | 0.884 | 1.000 | 37 | tags=50%, list=29%, signal=65% |
| 372 | HP\_NATAL\_TOOTH |  | 10 | -0.23 | -0.87 | 0.611 | 0.882 | 1.000 | 75 | tags=80%, list=60%, signal=182% |
| 373 | GOBP\_CENTRAL\_NERVOUS\_SYSTEM\_DEVELOPMENT |  | 27 | -0.15 | -0.87 | 0.595 | 0.879 | 1.000 | 79 | tags=74%, list=63%, signal=156% |
| 374 | GOBP\_RESPONSE\_TO\_CYTOKINE |  | 12 | -0.21 | -0.87 | 0.626 | 0.881 | 1.000 | 29 | tags=42%, list=23%, signal=49% |
| 375 | SKIL\_TARGET\_GENES |  | 13 | -0.21 | -0.87 | 0.597 | 0.881 | 1.000 | 103 | tags=100%, list=82%, signal=491% |
| 376 | UBN1\_TARGET\_GENES |  | 12 | -0.21 | -0.86 | 0.621 | 0.882 | 1.000 | 61 | tags=67%, list=48%, signal=117% |
| 377 | HP\_TREMOR |  | 16 | -0.19 | -0.86 | 0.620 | 0.881 | 1.000 | 43 | tags=50%, list=34%, signal=66% |
| 378 | GOBP\_HEAD\_DEVELOPMENT |  | 26 | -0.15 | -0.86 | 0.636 | 0.883 | 1.000 | 68 | tags=65%, list=54%, signal=113% |
| 379 | HP\_BROAD\_THUMB |  | 10 | -0.23 | -0.86 | 0.627 | 0.883 | 1.000 | 37 | tags=50%, list=29%, signal=65% |
| 380 | HP\_INGUINAL\_HERNIA |  | 18 | -0.18 | -0.86 | 0.583 | 0.884 | 1.000 | 38 | tags=44%, list=30%, signal=55% |
| 381 | HP\_RENAL\_AGENESIS |  | 23 | -0.16 | -0.85 | 0.635 | 0.892 | 1.000 | 39 | tags=43%, list=31%, signal=51% |
| 382 | HP\_NEVUS |  | 10 | -0.22 | -0.85 | 0.658 | 0.891 | 1.000 | 38 | tags=50%, list=30%, signal=66% |
| 383 | HP\_ABNORMAL\_LIP\_MORPHOLOGY |  | 59 | -0.12 | -0.85 | 0.637 | 0.890 | 1.000 | 76 | tags=66%, list=60%, signal=89% |
| 384 | GOBP\_POSITIVE\_REGULATION\_OF\_PROTEIN\_PHOSPHORYLATION |  | 11 | -0.22 | -0.85 | 0.640 | 0.891 | 1.000 | 90 | tags=91%, list=71%, signal=290% |
| 385 | KEGG\_PATHWAYS\_IN\_CANCER |  | 11 | -0.21 | -0.85 | 0.647 | 0.895 | 1.000 | 68 | tags=73%, list=54%, signal=144% |
| 386 | MEBARKI\_HCC\_PROGENITOR\_FZD8CRD\_UP |  | 13 | -0.21 | -0.84 | 0.656 | 0.897 | 1.000 | 65 | tags=69%, list=52%, signal=128% |
| 387 | GOBP\_CIRCULATORY\_SYSTEM\_DEVELOPMENT |  | 28 | -0.15 | -0.84 | 0.653 | 0.898 | 1.000 | 31 | tags=36%, list=25%, signal=37% |
| 388 | GOBP\_ORGAN\_GROWTH |  | 12 | -0.21 | -0.84 | 0.664 | 0.896 | 1.000 | 19 | tags=33%, list=15%, signal=36% |
| 389 | HP\_SEVERE\_SHORT\_STATURE |  | 10 | -0.23 | -0.84 | 0.640 | 0.894 | 1.000 | 63 | tags=70%, list=50%, signal=129% |
| 390 | GOBP\_ORGANELLE\_ASSEMBLY |  | 18 | -0.18 | -0.84 | 0.620 | 0.893 | 1.000 | 80 | tags=78%, list=63%, signal=183% |
| 391 | HP\_ECLABION |  | 10 | -0.23 | -0.84 | 0.646 | 0.892 | 1.000 | 37 | tags=50%, list=29%, signal=65% |
| 392 | GOBP\_BIOLOGICAL\_ADHESION |  | 14 | -0.20 | -0.84 | 0.642 | 0.893 | 1.000 | 24 | tags=36%, list=19%, signal=39% |
| 393 | HP\_ABNORMAL\_MIDDLE\_PHALANX\_MORPHOLOGY\_OF\_THE\_HAND |  | 13 | -0.20 | -0.84 | 0.647 | 0.891 | 1.000 | 75 | tags=77%, list=60%, signal=170% |
| 394 | GOBP\_POSITIVE\_REGULATION\_OF\_MULTICELLULAR\_ORGANISMAL\_PROCESS |  | 25 | -0.15 | -0.84 | 0.677 | 0.891 | 1.000 | 36 | tags=40%, list=29%, signal=45% |
| 395 | HP\_PAIN |  | 11 | -0.22 | -0.84 | 0.687 | 0.889 | 1.000 | 102 | tags=100%, list=81%, signal=479% |
| 396 | GOCC\_NUCLEOLUS |  | 10 | -0.22 | -0.84 | 0.660 | 0.890 | 1.000 | 26 | tags=40%, list=21%, signal=46% |
| 397 | HP\_DISPROPORTIONATE\_SHORT\_STATURE |  | 13 | -0.20 | -0.84 | 0.675 | 0.889 | 1.000 | 37 | tags=46%, list=29%, signal=59% |
| 398 | GOBP\_TUBE\_FORMATION |  | 10 | -0.22 | -0.83 | 0.631 | 0.888 | 1.000 | 76 | tags=80%, list=60%, signal=186% |
| 399 | HP\_CONSTITUTIONAL\_SYMPTOM |  | 19 | -0.17 | -0.83 | 0.658 | 0.894 | 1.000 | 102 | tags=95%, list=81%, signal=422% |
| 400 | GOBP\_FEMALE\_SEX\_DIFFERENTIATION |  | 11 | -0.22 | -0.83 | 0.651 | 0.892 | 1.000 | 22 | tags=36%, list=17%, signal=40% |
| 401 | UBP1\_TARGET\_GENES |  | 13 | -0.20 | -0.83 | 0.660 | 0.896 | 1.000 | 66 | tags=69%, list=52%, signal=130% |
| 402 | HP\_BROAD\_LONG\_BONES |  | 13 | -0.20 | -0.83 | 0.682 | 0.895 | 1.000 | 37 | tags=46%, list=29%, signal=59% |
| 403 | HP\_ABNORMAL\_CIRCULATING\_HORMONE\_CONCENTRATION |  | 42 | -0.13 | -0.83 | 0.637 | 0.894 | 1.000 | 23 | tags=26%, list=18%, signal=21% |
| 404 | HP\_ABNORMAL\_TRACHEAL\_MORPHOLOGY |  | 13 | -0.19 | -0.82 | 0.693 | 0.894 | 1.000 | 76 | tags=77%, list=60%, signal=174% |
| 405 | HP\_ABNORMALITY\_OF\_THE\_NECK |  | 39 | -0.13 | -0.82 | 0.661 | 0.893 | 1.000 | 83 | tags=74%, list=66%, signal=150% |
| 406 | MEF2C\_TARGET\_GENES |  | 11 | -0.21 | -0.81 | 0.675 | 0.908 | 1.000 | 57 | tags=64%, list=45%, signal=106% |
| 407 | GOMF\_DNA\_BINDING\_TRANSCRIPTION\_FACTOR\_ACTIVITY |  | 19 | -0.17 | -0.81 | 0.691 | 0.909 | 1.000 | 23 | tags=32%, list=18%, signal=33% |
| 408 | HP\_HIGH\_NARROW\_PALATE |  | 12 | -0.20 | -0.81 | 0.684 | 0.909 | 1.000 | 62 | tags=67%, list=49%, signal=119% |
| 409 | HP\_ABNORMAL\_CARDIAC\_VENTRICLE\_MORPHOLOGY |  | 39 | -0.13 | -0.81 | 0.716 | 0.910 | 1.000 | 77 | tags=69%, list=61%, signal=123% |
| 410 | GOBP\_REGULATION\_OF\_CELL\_POPULATION\_PROLIFERATION |  | 24 | -0.15 | -0.81 | 0.667 | 0.909 | 1.000 | 75 | tags=71%, list=60%, signal=142% |
| 411 | HP\_ABNORMALITY\_OF\_THE\_URETHRA |  | 65 | -0.12 | -0.81 | 0.685 | 0.910 | 1.000 | 75 | tags=65%, list=60%, signal=77% |
| 412 | GOBP\_POSITIVE\_REGULATION\_OF\_PHOSPHORUS\_METABOLIC\_PROCESS |  | 15 | -0.18 | -0.80 | 0.700 | 0.911 | 1.000 | 90 | tags=87%, list=71%, signal=267% |
| 413 | HP\_RESPIRATORY\_TRACT\_INFECTION |  | 16 | -0.17 | -0.80 | 0.716 | 0.911 | 1.000 | 37 | tags=44%, list=29%, signal=54% |
| 414 | GOBP\_NITROGEN\_COMPOUND\_TRANSPORT |  | 22 | -0.15 | -0.80 | 0.707 | 0.910 | 1.000 | 88 | tags=82%, list=70%, signal=224% |
| 415 | GOBP\_STEROID\_BIOSYNTHETIC\_PROCESS |  | 11 | -0.21 | -0.80 | 0.727 | 0.913 | 1.000 | 46 | tags=55%, list=37%, signal=78% |
| 416 | HP\_PROTRUDING\_EAR |  | 14 | -0.19 | -0.80 | 0.663 | 0.912 | 1.000 | 52 | tags=57%, list=41%, signal=86% |
| 417 | HP\_CONE\_SHAPED\_EPIPHYSIS |  | 10 | -0.21 | -0.80 | 0.739 | 0.911 | 1.000 | 52 | tags=60%, list=41%, signal=94% |
| 418 | GOBP\_EPITHELIAL\_TUBE\_FORMATION |  | 10 | -0.22 | -0.80 | 0.722 | 0.912 | 1.000 | 76 | tags=80%, list=60%, signal=186% |
| 419 | HP\_ABNORMALITY\_OF\_DIGESTIVE\_SYSTEM\_MORPHOLOGY |  | 54 | -0.12 | -0.79 | 0.669 | 0.916 | 1.000 | 102 | tags=87%, list=81%, signal=261% |
| 420 | GOBP\_CELL\_CYCLE\_PROCESS |  | 19 | -0.16 | -0.79 | 0.716 | 0.917 | 1.000 | 17 | tags=26%, list=13%, signal=26% |
| 421 | HP\_ABNORMALITY\_OF\_UPPER\_LIMB\_JOINT |  | 34 | -0.13 | -0.79 | 0.711 | 0.915 | 1.000 | 37 | tags=38%, list=29%, signal=40% |
| 422 | HP\_APLASIA\_HYPOPLASIA\_OF\_THE\_LUNGS |  | 24 | -0.15 | -0.79 | 0.732 | 0.913 | 1.000 | 54 | tags=54%, list=43%, signal=77% |
| 423 | HP\_APLASIA\_HYPOPLASIA\_OF\_THE\_UTERUS |  | 19 | -0.16 | -0.79 | 0.736 | 0.918 | 1.000 | 77 | tags=74%, list=61%, signal=161% |
| 424 | GOBP\_SKELETAL\_SYSTEM\_MORPHOGENESIS |  | 12 | -0.20 | -0.79 | 0.720 | 0.917 | 1.000 | 94 | tags=92%, list=75%, signal=327% |
| 425 | HP\_ABNORMAL\_CEREBRAL\_CORTEX\_MORPHOLOGY |  | 24 | -0.15 | -0.79 | 0.706 | 0.917 | 1.000 | 70 | tags=67%, list=56%, signal=121% |
| 426 | GOBP\_OSSIFICATION |  | 15 | -0.18 | -0.79 | 0.722 | 0.915 | 1.000 | 23 | tags=33%, list=18%, signal=36% |
| 427 | HP\_APLASIA\_HYPOPLASIA\_AFFECTING\_BONES\_OF\_THE\_AXIAL\_SKELETON |  | 57 | -0.11 | -0.78 | 0.742 | 0.918 | 1.000 | 75 | tags=65%, list=60%, signal=88% |
| 428 | ZFHX3\_TARGET\_GENES |  | 11 | -0.20 | -0.78 | 0.755 | 0.918 | 1.000 | 58 | tags=64%, list=46%, signal=108% |
| 429 | HP\_HYPOPLASIA\_OF\_THE\_UTERUS |  | 12 | -0.19 | -0.78 | 0.737 | 0.917 | 1.000 | 53 | tags=58%, list=42%, signal=91% |
| 430 | BENPORATH\_SOX2\_TARGETS |  | 12 | -0.20 | -0.78 | 0.722 | 0.917 | 1.000 | 94 | tags=92%, list=75%, signal=327% |
| 431 | HP\_ABNORMALITY\_OF\_SKIN\_PHYSIOLOGY |  | 11 | -0.20 | -0.78 | 0.748 | 0.918 | 1.000 | 70 | tags=73%, list=56%, signal=149% |
| 432 | HP\_ABNORMALITY\_OF\_CRANIAL\_SUTURES |  | 30 | -0.14 | -0.78 | 0.735 | 0.918 | 1.000 | 76 | tags=70%, list=60%, signal=134% |
| 433 | HP\_ABNORMALITY\_OF\_MALAR\_BONES |  | 22 | -0.15 | -0.77 | 0.764 | 0.925 | 1.000 | 37 | tags=41%, list=29%, signal=48% |
| 434 | HP\_BIFID\_SCROTUM |  | 16 | -0.17 | -0.77 | 0.743 | 0.925 | 1.000 | 77 | tags=75%, list=61%, signal=168% |
| 435 | BENPORATH\_NANOG\_TARGETS |  | 12 | -0.20 | -0.77 | 0.729 | 0.925 | 1.000 | 94 | tags=92%, list=75%, signal=327% |
| 436 | HP\_DISPLACEMENT\_OF\_THE\_URETHRAL\_MEATUS |  | 60 | -0.11 | -0.77 | 0.780 | 0.923 | 1.000 | 23 | tags=23%, list=18%, signal=15% |
| 437 | HP\_UNDERDEVELOPED\_NASAL\_ALAE |  | 12 | -0.19 | -0.77 | 0.748 | 0.921 | 1.000 | 11 | tags=25%, list=9%, signal=25% |
| 438 | HP\_ABNORMALITY\_OF\_THE\_OPTIC\_NERVE |  | 24 | -0.14 | -0.77 | 0.738 | 0.919 | 1.000 | 102 | tags=92%, list=81%, signal=390% |
| 439 | HP\_BLEPHAROPHIMOSIS |  | 15 | -0.17 | -0.76 | 0.760 | 0.923 | 1.000 | 24 | tags=33%, list=19%, signal=36% |
| 440 | HP\_DYSPHAGIA |  | 18 | -0.16 | -0.76 | 0.756 | 0.921 | 1.000 | 68 | tags=67%, list=54%, signal=124% |
| 441 | HP\_CONGENITAL\_DIAPHRAGMATIC\_HERNIA |  | 12 | -0.19 | -0.76 | 0.737 | 0.921 | 1.000 | 74 | tags=75%, list=59%, signal=164% |
| 442 | GOBP\_TAXIS |  | 12 | -0.19 | -0.76 | 0.761 | 0.922 | 1.000 | 74 | tags=75%, list=59%, signal=164% |
| 443 | GOMF\_DNA\_BINDING\_TRANSCRIPTION\_ACTIVATOR\_ACTIVITY |  | 10 | -0.21 | -0.76 | 0.777 | 0.923 | 1.000 | 15 | tags=30%, list=12%, signal=31% |
| 444 | GOBP\_ORGANELLE\_FISSION |  | 10 | -0.20 | -0.76 | 0.768 | 0.923 | 1.000 | 66 | tags=70%, list=52%, signal=135% |
| 445 | GOBP\_POSITIVE\_REGULATION\_OF\_CELL\_POPULATION\_PROLIFERATION |  | 15 | -0.17 | -0.76 | 0.766 | 0.921 | 1.000 | 74 | tags=73%, list=59%, signal=157% |
| 446 | GOBP\_POSITIVE\_REGULATION\_OF\_TRANSPORT |  | 10 | -0.20 | -0.76 | 0.779 | 0.919 | 1.000 | 28 | tags=40%, list=22%, signal=47% |
| 447 | HP\_ABNORMALITY\_OF\_THE\_DIAPHRAGM |  | 12 | -0.19 | -0.75 | 0.773 | 0.923 | 1.000 | 74 | tags=75%, list=59%, signal=164% |
| 448 | HP\_DOWNSLANTED\_PALPEBRAL\_FISSURES |  | 42 | -0.12 | -0.75 | 0.724 | 0.924 | 1.000 | 27 | tags=29%, list=21%, signal=24% |
| 449 | HP\_ABNORMAL\_SOFT\_PALATE\_MORPHOLOGY |  | 20 | -0.15 | -0.75 | 0.785 | 0.925 | 1.000 | 73 | tags=70%, list=58%, signal=140% |
| 450 | HP\_APLASIA\_HYPOPLASIA\_OF\_THE\_PHALANGES\_OF\_THE\_HAND |  | 26 | -0.13 | -0.75 | 0.786 | 0.928 | 1.000 | 75 | tags=69%, list=60%, signal=136% |
| 451 | HP\_ABNORMAL\_CONJUGATE\_EYE\_MOVEMENT |  | 41 | -0.11 | -0.74 | 0.776 | 0.937 | 1.000 | 22 | tags=24%, list=17%, signal=20% |
| 452 | HP\_GLUCOSE\_INTOLERANCE |  | 16 | -0.17 | -0.73 | 0.815 | 0.944 | 1.000 | 93 | tags=88%, list=74%, signal=292% |
| 453 | HP\_IMPAIRMENT\_IN\_PERSONALITY\_FUNCTIONING |  | 21 | -0.14 | -0.73 | 0.744 | 0.943 | 1.000 | 28 | tags=33%, list=22%, signal=36% |
| 454 | AEBP2\_TARGET\_GENES |  | 12 | -0.18 | -0.73 | 0.807 | 0.945 | 1.000 | 65 | tags=67%, list=52%, signal=125% |
| 455 | HP\_ABNORMAL\_UPPER\_LIMB\_BONE\_MORPHOLOGY |  | 38 | -0.12 | -0.73 | 0.811 | 0.943 | 1.000 | 37 | tags=37%, list=29%, signal=36% |
| 456 | GOCC\_NEURON\_PROJECTION |  | 14 | -0.17 | -0.73 | 0.794 | 0.944 | 1.000 | 36 | tags=43%, list=29%, signal=53% |
| 457 | HP\_ABNORMAL\_RENAL\_PHYSIOLOGY |  | 29 | -0.12 | -0.73 | 0.817 | 0.946 | 1.000 | 28 | tags=31%, list=22%, signal=31% |
| 458 | GOBP\_REGULATION\_OF\_MULTICELLULAR\_ORGANISMAL\_DEVELOPMENT |  | 21 | -0.14 | -0.72 | 0.767 | 0.947 | 1.000 | 88 | tags=81%, list=70%, signal=224% |
| 459 | AHRR\_TARGET\_GENES |  | 13 | -0.17 | -0.72 | 0.831 | 0.947 | 1.000 | 69 | tags=69%, list=55%, signal=137% |
| 460 | HP\_ABNORMAL\_VAGINA\_MORPHOLOGY |  | 30 | -0.13 | -0.72 | 0.799 | 0.947 | 1.000 | 73 | tags=67%, list=58%, signal=121% |
| 461 | GOBP\_HEART\_MORPHOGENESIS |  | 14 | -0.17 | -0.72 | 0.779 | 0.947 | 1.000 | 27 | tags=36%, list=21%, signal=40% |
| 462 | GRYDER\_PAX3FOXO1\_ENHANCERS\_KO\_DOWN |  | 10 | -0.19 | -0.72 | 0.832 | 0.950 | 1.000 | 29 | tags=40%, list=23%, signal=48% |
| 463 | HP\_SHORT\_MIDDLE\_PHALANX\_OF\_FINGER |  | 10 | -0.19 | -0.71 | 0.837 | 0.950 | 1.000 | 67 | tags=70%, list=53%, signal=138% |
| 464 | HP\_ABNORMALITY\_OF\_FACIAL\_SKELETON |  | 58 | -0.10 | -0.71 | 0.860 | 0.951 | 1.000 | 70 | tags=60%, list=56%, signal=73% |
| 465 | GOBP\_PROTEOLYSIS |  | 16 | -0.16 | -0.71 | 0.837 | 0.950 | 1.000 | 31 | tags=38%, list=25%, signal=43% |
| 466 | HP\_MORTALITY\_AGING |  | 18 | -0.15 | -0.71 | 0.794 | 0.948 | 1.000 | 90 | tags=83%, list=71%, signal=250% |
| 467 | HP\_BRACHYCEPHALY |  | 16 | -0.16 | -0.71 | 0.843 | 0.947 | 1.000 | 94 | tags=88%, list=75%, signal=301% |
| 468 | HP\_ABNORMALITY\_OF\_THE\_CHEEK |  | 16 | -0.16 | -0.71 | 0.836 | 0.948 | 1.000 | 39 | tags=44%, list=31%, signal=55% |
| 469 | GOCC\_MITOCHONDRION |  | 11 | -0.18 | -0.71 | 0.806 | 0.947 | 1.000 | 106 | tags=100%, list=84%, signal=575% |
| 470 | HP\_TETRALOGY\_OF\_FALLOT |  | 11 | -0.18 | -0.70 | 0.845 | 0.951 | 1.000 | 37 | tags=45%, list=29%, signal=59% |
| 471 | GOBP\_CELL\_CYCLE |  | 25 | -0.13 | -0.70 | 0.837 | 0.952 | 1.000 | 18 | tags=24%, list=14%, signal=22% |
| 472 | GOBP\_LYMPHOCYTE\_ACTIVATION |  | 12 | -0.18 | -0.69 | 0.853 | 0.958 | 1.000 | 23 | tags=33%, list=18%, signal=37% |
| 473 | HP\_MICRODONTIA |  | 16 | -0.15 | -0.69 | 0.845 | 0.962 | 1.000 | 110 | tags=100%, list=87%, signal=688% |
| 474 | GOBP\_CANONICAL\_WNT\_SIGNALING\_PATHWAY |  | 12 | -0.17 | -0.69 | 0.850 | 0.961 | 1.000 | 76 | tags=75%, list=60%, signal=171% |
| 475 | HP\_ABNORMALITY\_OF\_THE\_SKULL\_BASE |  | 19 | -0.14 | -0.69 | 0.848 | 0.962 | 1.000 | 39 | tags=42%, list=31%, signal=52% |
| 476 | HP\_BLINDNESS |  | 11 | -0.17 | -0.69 | 0.871 | 0.961 | 1.000 | 107 | tags=100%, list=85%, signal=605% |
| 477 | HP\_ABNORMAL\_LARYNX\_MORPHOLOGY |  | 22 | -0.13 | -0.69 | 0.843 | 0.959 | 1.000 | 33 | tags=36%, list=26%, signal=41% |
| 478 | GOBP\_REGULATION\_OF\_CELL\_CYCLE\_PROCESS |  | 12 | -0.17 | -0.68 | 0.866 | 0.959 | 1.000 | 45 | tags=50%, list=36%, signal=70% |
| 479 | GOBP\_MITOTIC\_CELL\_CYCLE\_PROCESS |  | 11 | -0.18 | -0.68 | 0.849 | 0.957 | 1.000 | 38 | tags=45%, list=30%, signal=59% |
| 480 | HP\_ABNORMALITY\_OF\_THE\_ZYGOMATIC\_BONE |  | 23 | -0.13 | -0.68 | 0.840 | 0.957 | 1.000 | 37 | tags=39%, list=29%, signal=45% |
| 481 | REACTOME\_POST\_TRANSLATIONAL\_PROTEIN\_MODIFICATION |  | 11 | -0.17 | -0.68 | 0.887 | 0.960 | 1.000 | 107 | tags=100%, list=85%, signal=605% |
| 482 | HP\_TRIANGULAR\_SHAPED\_PHALANGES\_OF\_THE\_HAND |  | 17 | -0.15 | -0.68 | 0.864 | 0.959 | 1.000 | 37 | tags=41%, list=29%, signal=50% |
| 483 | HP\_LIMITED\_ELBOW\_MOVEMENT |  | 12 | -0.17 | -0.68 | 0.854 | 0.958 | 1.000 | 24 | tags=33%, list=19%, signal=37% |
| 484 | ZNF618\_TARGET\_GENES |  | 14 | -0.15 | -0.68 | 0.847 | 0.956 | 1.000 | 29 | tags=36%, list=23%, signal=41% |
| 485 | HP\_ABNORMALITY\_OF\_DENTAL\_MORPHOLOGY |  | 26 | -0.12 | -0.67 | 0.842 | 0.960 | 1.000 | 110 | tags=96%, list=87%, signal=601% |
| 486 | HP\_DENTAL\_CROWDING |  | 17 | -0.15 | -0.67 | 0.843 | 0.960 | 1.000 | 37 | tags=41%, list=29%, signal=50% |
| 487 | GOBP\_NEGATIVE\_REGULATION\_OF\_CELL\_DIFFERENTIATION |  | 14 | -0.15 | -0.67 | 0.840 | 0.960 | 1.000 | 29 | tags=36%, list=23%, signal=41% |
| 488 | SFMBT1\_TARGET\_GENES |  | 17 | -0.14 | -0.67 | 0.876 | 0.962 | 1.000 | 30 | tags=35%, list=24%, signal=40% |
| 489 | HP\_TALL\_STATURE |  | 10 | -0.18 | -0.66 | 0.888 | 0.961 | 1.000 | 94 | tags=90%, list=75%, signal=326% |
| 490 | HP\_ABNORMALITY\_OF\_LOWER\_LIMB\_JOINT |  | 30 | -0.11 | -0.66 | 0.862 | 0.960 | 1.000 | 32 | tags=33%, list=25%, signal=34% |
| 491 | HP\_REDUCED\_VISUAL\_ACUITY |  | 17 | -0.14 | -0.66 | 0.862 | 0.958 | 1.000 | 67 | tags=65%, list=53%, signal=120% |
| 492 | GOBP\_CELLULAR\_RESPONSE\_TO\_STRESS |  | 20 | -0.13 | -0.66 | 0.898 | 0.961 | 1.000 | 107 | tags=95%, list=85%, signal=530% |
| 493 | GOCC\_ENDOPLASMIC\_RETICULUM |  | 20 | -0.13 | -0.65 | 0.865 | 0.963 | 1.000 | 12 | tags=20%, list=10%, signal=19% |
| 494 | HP\_ABNORMALITY\_OF\_HINDBRAIN\_MORPHOLOGY |  | 39 | -0.10 | -0.65 | 0.902 | 0.967 | 1.000 | 76 | tags=67%, list=60%, signal=116% |
| 495 | HP\_NARROW\_MOUTH |  | 13 | -0.16 | -0.65 | 0.875 | 0.966 | 1.000 | 22 | tags=31%, list=17%, signal=33% |
| 496 | GOBP\_CELLULAR\_COMPONENT\_MORPHOGENESIS |  | 14 | -0.15 | -0.65 | 0.880 | 0.964 | 1.000 | 74 | tags=71%, list=59%, signal=154% |
| 497 | HP\_ABNORMALITY\_OF\_THE\_MAXILLA |  | 15 | -0.14 | -0.65 | 0.891 | 0.963 | 1.000 | 69 | tags=67%, list=55%, signal=130% |
| 498 | HP\_ABNORMAL\_FACIAL\_SHAPE |  | 50 | -0.10 | -0.65 | 0.871 | 0.962 | 1.000 | 44 | tags=40%, list=35%, signal=37% |
| 499 | HP\_ABNORMALITY\_OF\_THE\_SUPRAORBITAL\_RIDGES |  | 10 | -0.17 | -0.64 | 0.930 | 0.969 | 1.000 | 44 | tags=50%, list=35%, signal=71% |
| 500 | HP\_ABNORMAL\_GLUCOSE\_HOMEOSTASIS |  | 23 | -0.12 | -0.64 | 0.917 | 0.970 | 1.000 | 98 | tags=87%, list=78%, signal=320% |
| 501 | HP\_ABNORMALITY\_OF\_THE\_ABDOMINAL\_WALL |  | 39 | -0.10 | -0.63 | 0.893 | 0.969 | 1.000 | 76 | tags=67%, list=60%, signal=116% |
| 502 | BENPORATH\_ES\_WITH\_H3K27ME3 |  | 11 | -0.16 | -0.63 | 0.887 | 0.969 | 1.000 | 5 | tags=18%, list=4%, signal=17% |
| 503 | HAY\_BONE\_MARROW\_STROMAL |  | 17 | -0.14 | -0.62 | 0.919 | 0.977 | 1.000 | 75 | tags=71%, list=60%, signal=151% |
| 504 | HP\_WIDE\_NASAL\_BRIDGE |  | 39 | -0.10 | -0.62 | 0.911 | 0.976 | 1.000 | 44 | tags=41%, list=35%, signal=44% |
| 505 | MIR374B\_5P |  | 10 | -0.16 | -0.61 | 0.929 | 0.983 | 1.000 | 20 | tags=30%, list=16%, signal=33% |
| 506 | GOBP\_POSITIVE\_REGULATION\_OF\_SIGNALING |  | 31 | -0.11 | -0.61 | 0.933 | 0.984 | 1.000 | 68 | tags=61%, list=54%, signal=100% |
| 507 | WANG\_MLL\_TARGETS |  | 10 | -0.16 | -0.61 | 0.913 | 0.983 | 1.000 | 108 | tags=100%, list=86%, signal=644% |
| 508 | HP\_RADIAL\_DEVIATION\_OF\_THE\_HAND\_OR\_OF\_FINGERS\_OF\_THE\_HAND |  | 14 | -0.14 | -0.61 | 0.900 | 0.982 | 1.000 | 102 | tags=93%, list=81%, signal=433% |
| 509 | GOBP\_BLOOD\_VESSEL\_MORPHOGENESIS |  | 12 | -0.15 | -0.61 | 0.930 | 0.980 | 1.000 | 68 | tags=67%, list=54%, signal=131% |
| 510 | HP\_ABNORMAL\_OVARIAN\_PHYSIOLOGY |  | 11 | -0.16 | -0.60 | 0.931 | 0.983 | 1.000 | 6 | tags=18%, list=5%, signal=17% |
| 511 | HP\_APLASIA\_HYPOPLASIA\_INVOLVING\_THE\_NOSE |  | 15 | -0.14 | -0.60 | 0.950 | 0.982 | 1.000 | 11 | tags=20%, list=9%, signal=19% |
| 512 | HP\_WIDE\_INTERMAMILLARY\_DISTANCE |  | 15 | -0.14 | -0.60 | 0.946 | 0.981 | 1.000 | 11 | tags=20%, list=9%, signal=19% |
| 513 | HP\_ABNORMAL\_CARDIAC\_SEPTUM\_MORPHOLOGY |  | 44 | -0.09 | -0.60 | 0.938 | 0.980 | 1.000 | 45 | tags=41%, list=36%, signal=41% |
| 514 | HP\_ABNORMALITY\_OF\_THE\_UPPER\_RESPIRATORY\_TRACT |  | 35 | -0.10 | -0.59 | 0.947 | 0.984 | 1.000 | 75 | tags=66%, list=60%, signal=117% |
| 515 | HP\_ABNORMALITY\_OF\_THE\_HELIX |  | 17 | -0.13 | -0.59 | 0.940 | 0.982 | 1.000 | 113 | tags=100%, list=90%, signal=838% |
| 516 | HP\_LONG\_PHILTRUM |  | 18 | -0.12 | -0.58 | 0.939 | 0.985 | 1.000 | 37 | tags=39%, list=29%, signal=47% |
| 517 | BENPORATH\_EED\_TARGETS |  | 13 | -0.14 | -0.57 | 0.949 | 0.988 | 1.000 | 5 | tags=15%, list=4%, signal=14% |
| 518 | HP\_ABNORMAL\_INVOLUNTARY\_EYE\_MOVEMENTS |  | 43 | -0.09 | -0.57 | 0.941 | 0.986 | 1.000 | 93 | tags=79%, list=74%, signal=199% |
| 519 | ZSCAN29\_TARGET\_GENES |  | 11 | -0.15 | -0.57 | 0.949 | 0.986 | 1.000 | 30 | tags=36%, list=24%, signal=44% |
| 520 | MURARO\_PANCREAS\_BETA\_CELL |  | 14 | -0.13 | -0.57 | 0.937 | 0.985 | 1.000 | 103 | tags=93%, list=82%, signal=452% |
| 521 | HP\_THICKENED\_SKIN |  | 16 | -0.12 | -0.56 | 0.958 | 0.990 | 1.000 | 90 | tags=81%, list=71%, signal=248% |
| 522 | CTTTGA\_LEF1\_Q2 |  | 16 | -0.12 | -0.55 | 0.972 | 0.992 | 1.000 | 90 | tags=81%, list=71%, signal=248% |
| 523 | GOBP\_MITOTIC\_CELL\_CYCLE |  | 12 | -0.14 | -0.54 | 0.961 | 0.992 | 1.000 | 38 | tags=42%, list=30%, signal=54% |
| 524 | HP\_ABNORMALITY\_OF\_THE\_OVARY |  | 41 | -0.08 | -0.54 | 0.975 | 0.992 | 1.000 | 83 | tags=71%, list=66%, signal=140% |
| 525 | HP\_UPSLANTED\_PALPEBRAL\_FISSURE |  | 18 | -0.11 | -0.54 | 0.963 | 0.993 | 1.000 | 73 | tags=67%, list=58%, signal=136% |
| 526 | HP\_ABNORMAL\_BLOOD\_ION\_CONCENTRATION |  | 10 | -0.14 | -0.53 | 0.986 | 0.995 | 1.000 | 98 | tags=90%, list=78%, signal=373% |
| 527 | GOBP\_CELL\_PART\_MORPHOGENESIS |  | 13 | -0.13 | -0.52 | 0.978 | 0.994 | 1.000 | 35 | tags=38%, list=28%, signal=48% |
| 528 | GOBP\_INTRACELLULAR\_TRANSPORT |  | 20 | -0.11 | -0.52 | 0.980 | 0.993 | 1.000 | 78 | tags=70%, list=62%, signal=155% |
| 529 | HP\_LIMB\_JOINT\_CONTRACTURE |  | 24 | -0.10 | -0.52 | 0.978 | 0.992 | 1.000 | 75 | tags=67%, list=60%, signal=133% |
| 530 | HP\_CEREBRAL\_CORTICAL\_ATROPHY |  | 13 | -0.12 | -0.51 | 0.978 | 0.992 | 1.000 | 55 | tags=54%, list=44%, signal=86% |
| 531 | HP\_CONTRACTURES\_OF\_THE\_JOINTS\_OF\_THE\_UPPER\_LIMBS |  | 23 | -0.10 | -0.50 | 0.992 | 0.994 | 1.000 | 24 | tags=26%, list=19%, signal=26% |
| 532 | HP\_ABNORMALITY\_OF\_MOUTH\_SIZE |  | 25 | -0.09 | -0.49 | 0.982 | 0.994 | 1.000 | 22 | tags=24%, list=17%, signal=23% |
| 533 | HP\_ABNORMALITY\_OF\_DENTAL\_ERUPTION |  | 30 | -0.09 | -0.49 | 0.988 | 0.993 | 1.000 | 39 | tags=37%, list=31%, signal=40% |
| 534 | HP\_ABNORMALITY\_OF\_THE\_HALLUX |  | 19 | -0.10 | -0.47 | 0.994 | 0.995 | 1.000 | 37 | tags=37%, list=29%, signal=44% |
| 535 | HP\_ABNORMAL\_SACRUM\_MORPHOLOGY |  | 10 | -0.12 | -0.44 | 0.996 | 0.997 | 1.000 | 12 | tags=20%, list=10%, signal=20% |
Table: Gene sets enriched in phenotype **na **[plain text format]****

  
