## Supplemental File 3 for "Increased Expression of *ZFPM2* Bypasses *SRY* to Drive 46,XX Testicular Development: A New Mechanism of 46,XX DSD": gsea_report_for_na_pos_1653344355978.html

Report for na\_pos 1653344355978 [GSEA]

| GS  follow link to MSigDB | GS DETAILS | SIZE | ES | NES | NOM p-val | FDR q-val | FWER p-val | RANK AT MAX | LEADING EDGE || 1 | DACOSTA\_UV\_RESPONSE\_VIA\_ERCC3\_DN | Details ... | 13 | 0.58 | 2.41 | 0.000 | 0.180 | 0.179 | 30 | tags=77%, list=24%, signal=91% |
| 2 | BUSSLINGER\_GASTRIC\_IMMUNE\_CELLS | Details ... | 11 | 0.55 | 2.14 | 0.002 | 0.583 | 0.711 | 39 | tags=82%, list=31%, signal=108% |
| 3 | HP\_INFERTILITY | Details ... | 18 | 0.44 | 2.10 | 0.002 | 0.490 | 0.788 | 43 | tags=72%, list=34%, signal=94% |
| 4 | HP\_THICK\_VERMILION\_BORDER | Details ... | 10 | 0.52 | 1.97 | 0.012 | 0.823 | 0.973 | 39 | tags=80%, list=31%, signal=107% |
| 5 | NAB2\_TARGET\_GENES | Details ... | 10 | 0.52 | 1.94 | 0.006 | 0.781 | 0.987 | 52 | tags=90%, list=41%, signal=141% |
| 6 | HP\_ABNORMALITY\_OF\_HAIR\_TEXTURE | Details ... | 11 | 0.48 | 1.86 | 0.018 | 1.000 | 0.999 | 58 | tags=91%, list=46%, signal=154% |
| 7 | GOBP\_MALE\_SEX\_DIFFERENTIATION | Details ... | 17 | 0.39 | 1.83 | 0.010 | 1.000 | 0.999 | 31 | tags=59%, list=25%, signal=67% |
| 8 | HP\_UPPER\_MOTOR\_NEURON\_DYSFUNCTION | Details ... | 25 | 0.33 | 1.79 | 0.010 | 1.000 | 1.000 | 36 | tags=56%, list=29%, signal=63% |
| 9 | HP\_GONOSOMAL\_INHERITANCE | Details ... | 12 | 0.44 | 1.79 | 0.010 | 1.000 | 1.000 | 33 | tags=67%, list=26%, signal=82% |
| 10 | HP\_INTELLECTUAL\_DISABILITY\_SEVERE | Details ... | 11 | 0.47 | 1.78 | 0.014 | 0.961 | 1.000 | 37 | tags=73%, list=29%, signal=94% |
| 11 | GOBP\_IMMUNE\_RESPONSE | Details ... | 10 | 0.48 | 1.78 | 0.028 | 0.880 | 1.000 | 44 | tags=80%, list=35%, signal=113% |
| 12 | HP\_DECREASED\_FERTILITY\_IN\_MALES | Details ... | 11 | 0.45 | 1.76 | 0.021 | 0.874 | 1.000 | 39 | tags=73%, list=31%, signal=96% |
| 13 | HP\_SHORT\_THUMB | Details ... | 10 | 0.45 | 1.74 | 0.018 | 0.903 | 1.000 | 60 | tags=90%, list=48%, signal=158% |
| 14 | HP\_SPASTICITY | Details ... | 16 | 0.38 | 1.74 | 0.020 | 0.862 | 1.000 | 36 | tags=63%, list=29%, signal=76% |
| 15 | GOMF\_TRANSITION\_METAL\_ION\_BINDING | Details ... | 11 | 0.44 | 1.73 | 0.024 | 0.833 | 1.000 | 63 | tags=91%, list=50%, signal=166% |
| 16 | GOCC\_CATALYTIC\_COMPLEX | Details ... | 12 | 0.43 | 1.71 | 0.039 | 0.849 | 1.000 | 45 | tags=75%, list=36%, signal=106% |
| 17 | HP\_ABNORMAL\_FEMORAL\_NECK\_HEAD\_MORPHOLOGY | Details ... | 11 | 0.44 | 1.68 | 0.028 | 0.917 | 1.000 | 52 | tags=82%, list=41%, signal=127% |
| 18 | HP\_HETEROTROPIA | Details ... | 10 | 0.45 | 1.68 | 0.036 | 0.875 | 1.000 | 60 | tags=90%, list=48%, signal=158% |
| 19 | RYTTCCTG\_ETS2\_B | Details ... | 12 | 0.42 | 1.67 | 0.033 | 0.877 | 1.000 | 46 | tags=75%, list=37%, signal=107% |
| 20 | GOBP\_POSITIVE\_REGULATION\_OF\_GENE\_EXPRESSION | Details ... | 13 | 0.40 | 1.67 | 0.034 | 0.837 | 1.000 | 70 | tags=92%, list=56%, signal=186% |
| 21 | HP\_ABNORMALITY\_OF\_THE\_BREAST | Details ... | 42 | 0.26 | 1.65 | 0.029 | 0.877 | 1.000 | 67 | tags=71%, list=53%, signal=102% |
| 22 | HP\_HYPERACTIVITY | Details ... | 14 | 0.38 | 1.64 | 0.041 | 0.874 | 1.000 | 37 | tags=64%, list=29%, signal=81% |
| 23 | HP\_ABNORMAL\_ILIUM\_MORPHOLOGY | Details ... | 10 | 0.43 | 1.63 | 0.042 | 0.856 | 1.000 | 37 | tags=70%, list=29%, signal=91% |
| 24 | HP\_HYPERTONIA | Details ... | 17 | 0.34 | 1.62 | 0.049 | 0.868 | 1.000 | 36 | tags=59%, list=29%, signal=71% |
| 25 | GOBP\_RESPONSE\_TO\_OXYGEN\_CONTAINING\_COMPOUND | Details ... | 19 | 0.33 | 1.62 | 0.050 | 0.835 | 1.000 | 63 | tags=79%, list=50%, signal=134% |
| 26 | HP\_HYPOGONADOTROPIC\_HYPOGONADISM | Details ... | 19 | 0.32 | 1.58 | 0.049 | 0.996 | 1.000 | 24 | tags=47%, list=19%, signal=50% |
| 27 | HP\_MALE\_INFERTILITY |  | 10 | 0.42 | 1.57 | 0.058 | 1.000 | 1.000 | 39 | tags=70%, list=31%, signal=93% |
| 28 | HP\_FUNCTIONAL\_ABNORMALITY\_OF\_THE\_INNER\_EAR |  | 42 | 0.24 | 1.56 | 0.043 | 0.993 | 1.000 | 78 | tags=79%, list=62%, signal=138% |
| 29 | ZNF740\_TARGET\_GENES |  | 10 | 0.41 | 1.55 | 0.061 | 0.992 | 1.000 | 52 | tags=80%, list=41%, signal=125% |
| 30 | HP\_ABNORMALITY\_OF\_BONE\_MINERAL\_DENSITY |  | 30 | 0.26 | 1.54 | 0.063 | 1.000 | 1.000 | 62 | tags=70%, list=49%, signal=105% |
| 31 | GOBP\_NEGATIVE\_REGULATION\_OF\_PROTEIN\_METABOLIC\_PROCESS |  | 11 | 0.40 | 1.53 | 0.060 | 1.000 | 1.000 | 45 | tags=73%, list=36%, signal=103% |
| 32 | HP\_NEOPLASM\_BY\_HISTOLOGY |  | 17 | 0.33 | 1.52 | 0.069 | 1.000 | 1.000 | 15 | tags=41%, list=12%, signal=40% |
| 33 | HP\_ABNORMALITY\_OF\_THE\_PUBIC\_HAIR |  | 13 | 0.36 | 1.51 | 0.062 | 1.000 | 1.000 | 36 | tags=62%, list=29%, signal=77% |
| 34 | DIAZ\_CHRONIC\_MYELOGENOUS\_LEUKEMIA\_UP |  | 14 | 0.36 | 1.50 | 0.055 | 1.000 | 1.000 | 13 | tags=43%, list=10%, signal=42% |
| 35 | GOBP\_REGULATION\_OF\_INTRACELLULAR\_SIGNAL\_TRANSDUCTION |  | 21 | 0.30 | 1.49 | 0.080 | 1.000 | 1.000 | 70 | tags=81%, list=56%, signal=152% |
| 36 | HP\_SPARSE\_HAIR |  | 34 | 0.25 | 1.49 | 0.080 | 1.000 | 1.000 | 65 | tags=71%, list=52%, signal=106% |
| 37 | HP\_ABNORMAL\_SPERMATOGENESIS |  | 20 | 0.30 | 1.48 | 0.089 | 1.000 | 1.000 | 43 | tags=60%, list=34%, signal=77% |
| 38 | HP\_HYPOPLASIA\_OF\_THE\_CORPUS\_CALLOSUM |  | 13 | 0.34 | 1.47 | 0.091 | 1.000 | 1.000 | 57 | tags=77%, list=45%, signal=126% |
| 39 | SUPT16H\_TARGET\_GENES |  | 12 | 0.36 | 1.47 | 0.070 | 1.000 | 1.000 | 52 | tags=75%, list=41%, signal=116% |
| 40 | HP\_DECREASED\_FERTILITY |  | 30 | 0.25 | 1.46 | 0.081 | 1.000 | 1.000 | 67 | tags=73%, list=53%, signal=119% |
| 41 | HP\_ABNORMAL\_ENCHONDRAL\_OSSIFICATION |  | 11 | 0.37 | 1.45 | 0.086 | 1.000 | 1.000 | 83 | tags=100%, list=66%, signal=267% |
| 42 | HP\_JOINT\_HYPERMOBILITY |  | 17 | 0.31 | 1.45 | 0.076 | 1.000 | 1.000 | 54 | tags=71%, list=43%, signal=107% |
| 43 | HP\_OSTEOPOROSIS |  | 22 | 0.28 | 1.44 | 0.087 | 1.000 | 1.000 | 56 | tags=68%, list=44%, signal=101% |
| 44 | HP\_ABNORMALITY\_OF\_THE\_AXILLARY\_HAIR |  | 11 | 0.38 | 1.43 | 0.092 | 1.000 | 1.000 | 36 | tags=64%, list=29%, signal=81% |
| 45 | BLALOCK\_ALZHEIMERS\_DISEASE\_DN |  | 13 | 0.35 | 1.43 | 0.106 | 1.000 | 1.000 | 86 | tags=100%, list=68%, signal=283% |
| 46 | HP\_ABNORMALITY\_OF\_FLUID\_REGULATION |  | 18 | 0.30 | 1.42 | 0.085 | 1.000 | 1.000 | 79 | tags=89%, list=63%, signal=204% |
| 47 | HP\_VASCULAR\_SKIN\_ABNORMALITY |  | 11 | 0.37 | 1.42 | 0.113 | 1.000 | 1.000 | 71 | tags=91%, list=56%, signal=190% |
| 48 | HP\_ABSENCE\_OF\_SECONDARY\_SEX\_CHARACTERISTICS |  | 15 | 0.32 | 1.41 | 0.102 | 1.000 | 1.000 | 56 | tags=73%, list=44%, signal=116% |
| 49 | MIR548AA\_MIR548AP\_3P\_MIR548T\_3P |  | 16 | 0.31 | 1.41 | 0.110 | 1.000 | 1.000 | 52 | tags=69%, list=41%, signal=102% |
| 50 | GOMF\_MOLECULAR\_FUNCTION\_REGULATOR |  | 15 | 0.31 | 1.41 | 0.106 | 1.000 | 1.000 | 23 | tags=47%, list=18%, signal=50% |
| 51 | HP\_EPICANTHUS |  | 31 | 0.24 | 1.41 | 0.117 | 1.000 | 1.000 | 45 | tags=55%, list=36%, signal=64% |
| 52 | HP\_MACULE |  | 13 | 0.33 | 1.40 | 0.107 | 1.000 | 1.000 | 39 | tags=62%, list=31%, signal=80% |
| 53 | HP\_PES\_PLANUS |  | 17 | 0.30 | 1.40 | 0.104 | 1.000 | 1.000 | 33 | tags=53%, list=26%, signal=62% |
| 54 | GOBP\_POSITIVE\_REGULATION\_OF\_INTRACELLULAR\_SIGNAL\_TRANSDUCTION |  | 16 | 0.31 | 1.39 | 0.123 | 1.000 | 1.000 | 67 | tags=81%, list=53%, signal=151% |
| 55 | GOBP\_MULTICELLULAR\_ORGANISM\_REPRODUCTION |  | 17 | 0.30 | 1.39 | 0.116 | 1.000 | 1.000 | 48 | tags=65%, list=38%, signal=90% |
| 56 | GOBP\_CELLULAR\_RESPONSE\_TO\_OXYGEN\_CONTAINING\_COMPOUND |  | 14 | 0.31 | 1.39 | 0.109 | 1.000 | 1.000 | 63 | tags=79%, list=50%, signal=140% |
| 57 | HP\_ABNORMAL\_BONE\_STRUCTURE |  | 36 | 0.23 | 1.39 | 0.112 | 1.000 | 1.000 | 83 | tags=83%, list=66%, signal=174% |
| 58 | GOBP\_CELLULAR\_RESPONSE\_TO\_NITROGEN\_COMPOUND |  | 10 | 0.38 | 1.39 | 0.124 | 1.000 | 1.000 | 56 | tags=80%, list=44%, signal=133% |
| 59 | MURARO\_PANCREAS\_DUCTAL\_CELL |  | 13 | 0.33 | 1.38 | 0.111 | 1.000 | 1.000 | 10 | tags=38%, list=8%, signal=37% |
| 60 | HP\_ABNORMALITY\_OF\_THE\_NASAL\_TIP |  | 17 | 0.30 | 1.38 | 0.126 | 0.992 | 1.000 | 33 | tags=53%, list=26%, signal=62% |
| 61 | GOBP\_ORGANONITROGEN\_COMPOUND\_CATABOLIC\_PROCESS |  | 12 | 0.34 | 1.38 | 0.120 | 0.983 | 1.000 | 44 | tags=67%, list=35%, signal=93% |
| 62 | GOBP\_SMALL\_MOLECULE\_METABOLIC\_PROCESS |  | 19 | 0.28 | 1.37 | 0.124 | 0.988 | 1.000 | 22 | tags=42%, list=17%, signal=43% |
| 63 | ZNF350\_TARGET\_GENES |  | 10 | 0.37 | 1.37 | 0.118 | 0.990 | 1.000 | 32 | tags=60%, list=25%, signal=74% |
| 64 | HP\_CAFE\_AU\_LAIT\_SPOT |  | 11 | 0.35 | 1.37 | 0.131 | 0.989 | 1.000 | 39 | tags=64%, list=31%, signal=84% |
| 65 | HP\_FUNCTIONAL\_ABNORMALITY\_OF\_MALE\_INTERNAL\_GENITALIA |  | 21 | 0.27 | 1.36 | 0.108 | 0.989 | 1.000 | 31 | tags=48%, list=25%, signal=53% |
| 66 | GOBP\_PROTEIN\_CONTAINING\_COMPLEX\_ORGANIZATION |  | 19 | 0.28 | 1.36 | 0.150 | 0.985 | 1.000 | 49 | tags=63%, list=39%, signal=88% |
| 67 | HP\_BOWING\_OF\_THE\_LEGS |  | 17 | 0.29 | 1.36 | 0.131 | 0.976 | 1.000 | 27 | tags=47%, list=21%, signal=52% |
| 68 | GOBERT\_OLIGODENDROCYTE\_DIFFERENTIATION\_DN |  | 10 | 0.37 | 1.36 | 0.120 | 0.967 | 1.000 | 70 | tags=90%, list=56%, signal=186% |
| 69 | HP\_ABNORMALITY\_OF\_SKELETAL\_MATURATION |  | 44 | 0.21 | 1.35 | 0.129 | 0.981 | 1.000 | 39 | tags=45%, list=31%, signal=43% |
| 70 | MIR520D\_5P |  | 11 | 0.34 | 1.35 | 0.145 | 0.967 | 1.000 | 63 | tags=82%, list=50%, signal=149% |
| 71 | GOBP\_REGULATION\_OF\_CELLULAR\_LOCALIZATION |  | 11 | 0.34 | 1.35 | 0.129 | 0.954 | 1.000 | 63 | tags=82%, list=50%, signal=149% |
| 72 | MIR524\_5P |  | 11 | 0.34 | 1.35 | 0.136 | 0.945 | 1.000 | 63 | tags=82%, list=50%, signal=149% |
| 73 | GOBP\_CHROMATIN\_ORGANIZATION |  | 11 | 0.34 | 1.34 | 0.139 | 0.955 | 1.000 | 29 | tags=55%, list=23%, signal=65% |
| 74 | MIR95\_5P |  | 11 | 0.35 | 1.34 | 0.129 | 0.953 | 1.000 | 39 | tags=64%, list=31%, signal=84% |
| 75 | GOBP\_DEVELOPMENT\_OF\_PRIMARY\_SEXUAL\_CHARACTERISTICS |  | 20 | 0.26 | 1.32 | 0.147 | 1.000 | 1.000 | 34 | tags=50%, list=27%, signal=58% |
| 76 | HP\_MOTOR\_DELAY |  | 19 | 0.26 | 1.32 | 0.153 | 1.000 | 1.000 | 37 | tags=53%, list=29%, signal=63% |
| 77 | HP\_ABNORMALITY\_OF\_THE\_MUSCULATURE\_OF\_THE\_LIMBS |  | 11 | 0.34 | 1.31 | 0.167 | 1.000 | 1.000 | 63 | tags=82%, list=50%, signal=149% |
| 78 | HP\_AGANGLIONIC\_MEGACOLON |  | 10 | 0.35 | 1.31 | 0.163 | 1.000 | 1.000 | 34 | tags=60%, list=27%, signal=76% |
| 79 | HP\_ABNORMALITY\_OF\_THE\_SENSE\_OF\_SMELL |  | 15 | 0.30 | 1.31 | 0.128 | 0.997 | 1.000 | 67 | tags=80%, list=53%, signal=151% |
| 80 | GOCC\_NUCLEAR\_BODY |  | 11 | 0.34 | 1.31 | 0.152 | 0.988 | 1.000 | 63 | tags=82%, list=50%, signal=149% |
| 81 | HP\_ABNORMAL\_CIRCULATING\_METABOLITE\_CONCENTRATION |  | 28 | 0.22 | 1.30 | 0.149 | 1.000 | 1.000 | 90 | tags=89%, list=71%, signal=243% |
| 82 | GOBP\_NEGATIVE\_REGULATION\_OF\_CELLULAR\_COMPONENT\_ORGANIZATION |  | 12 | 0.32 | 1.30 | 0.165 | 1.000 | 1.000 | 67 | tags=83%, list=53%, signal=161% |
| 83 | MAFG\_TARGET\_GENES |  | 12 | 0.32 | 1.29 | 0.189 | 1.000 | 1.000 | 15 | tags=42%, list=12%, signal=43% |
| 84 | REACTOME\_DISEASES\_OF\_SIGNAL\_TRANSDUCTION\_BY\_GROWTH\_FACTOR\_RECEPTORS\_AND\_SECOND\_MESSENGERS |  | 10 | 0.34 | 1.28 | 0.178 | 1.000 | 1.000 | 60 | tags=80%, list=48%, signal=141% |
| 85 | HP\_DYSPNEA |  | 13 | 0.31 | 1.28 | 0.171 | 1.000 | 1.000 | 90 | tags=100%, list=71%, signal=314% |
| 86 | REACTOME\_METABOLISM\_OF\_STEROIDS |  | 10 | 0.34 | 1.28 | 0.197 | 1.000 | 1.000 | 22 | tags=50%, list=17%, signal=56% |
| 87 | HP\_AZOOSPERMIA |  | 17 | 0.28 | 1.27 | 0.190 | 1.000 | 1.000 | 43 | tags=59%, list=34%, signal=77% |
| 88 | HP\_SHORT\_ATTENTION\_SPAN |  | 12 | 0.31 | 1.26 | 0.193 | 1.000 | 1.000 | 37 | tags=58%, list=29%, signal=75% |
| 89 | GRAESSMANN\_APOPTOSIS\_BY\_DOXORUBICIN\_DN |  | 24 | 0.24 | 1.26 | 0.175 | 1.000 | 1.000 | 48 | tags=58%, list=38%, signal=76% |
| 90 | HP\_ABNORMALITY\_OF\_FEMUR\_MORPHOLOGY |  | 18 | 0.26 | 1.26 | 0.170 | 1.000 | 1.000 | 27 | tags=44%, list=21%, signal=48% |
| 91 | GOBP\_ENZYME\_LINKED\_RECEPTOR\_PROTEIN\_SIGNALING\_PATHWAY |  | 21 | 0.25 | 1.26 | 0.182 | 1.000 | 1.000 | 63 | tags=71%, list=50%, signal=119% |
| 92 | HP\_ARNOLD\_CHIARI\_MALFORMATION |  | 11 | 0.32 | 1.25 | 0.187 | 1.000 | 1.000 | 20 | tags=45%, list=16%, signal=49% |
| 93 | HP\_IRREGULAR\_HYPERPIGMENTATION |  | 14 | 0.29 | 1.25 | 0.165 | 1.000 | 1.000 | 39 | tags=57%, list=31%, signal=74% |
| 94 | GRAESSMANN\_APOPTOSIS\_BY\_DOXORUBICIN\_UP |  | 10 | 0.33 | 1.25 | 0.209 | 1.000 | 1.000 | 74 | tags=90%, list=59%, signal=201% |
| 95 | HP\_ONSET |  | 18 | 0.26 | 1.25 | 0.192 | 1.000 | 1.000 | 76 | tags=83%, list=60%, signal=180% |
| 96 | HP\_LONG\_FINGERS |  | 11 | 0.32 | 1.24 | 0.218 | 1.000 | 1.000 | 65 | tags=82%, list=52%, signal=154% |
| 97 | HP\_JOINT\_LAXITY |  | 18 | 0.26 | 1.22 | 0.203 | 1.000 | 1.000 | 55 | tags=67%, list=44%, signal=101% |
| 98 | HP\_ABNORMALITY\_OF\_SECONDARY\_SEXUAL\_HAIR |  | 15 | 0.27 | 1.22 | 0.205 | 1.000 | 1.000 | 36 | tags=53%, list=29%, signal=66% |
| 99 | HP\_ABNORMAL\_REFLEX |  | 19 | 0.25 | 1.22 | 0.205 | 1.000 | 1.000 | 92 | tags=95%, list=73%, signal=298% |
| 100 | MIR1277\_5P |  | 10 | 0.32 | 1.22 | 0.214 | 1.000 | 1.000 | 75 | tags=90%, list=60%, signal=205% |
| 101 | HP\_HYPOPLASIA\_OF\_TEETH |  | 11 | 0.30 | 1.22 | 0.201 | 1.000 | 1.000 | 90 | tags=100%, list=71%, signal=319% |
| 102 | HP\_ABNORMALITY\_OF\_PRIMARY\_TEETH |  | 10 | 0.33 | 1.22 | 0.223 | 1.000 | 1.000 | 74 | tags=90%, list=59%, signal=201% |
| 103 | GOBP\_MAPK\_CASCADE |  | 15 | 0.27 | 1.21 | 0.211 | 1.000 | 1.000 | 70 | tags=80%, list=56%, signal=159% |
| 104 | HP\_DECREASED\_CIRCULATING\_ANDROGEN\_CONCENTRATION |  | 16 | 0.27 | 1.21 | 0.237 | 1.000 | 1.000 | 56 | tags=69%, list=44%, signal=108% |
| 105 | HP\_ABNORMAL\_CIRCULATING\_GONADOTROPIN\_CONCENTRATION |  | 18 | 0.25 | 1.21 | 0.219 | 1.000 | 1.000 | 63 | tags=72%, list=50%, signal=124% |
| 106 | HP\_GERM\_CELL\_NEOPLASIA |  | 10 | 0.32 | 1.21 | 0.230 | 1.000 | 1.000 | 37 | tags=60%, list=29%, signal=78% |
| 107 | HP\_ABNORMALITY\_OF\_REFRACTION |  | 19 | 0.25 | 1.21 | 0.236 | 1.000 | 1.000 | 39 | tags=53%, list=31%, signal=65% |
| 108 | HP\_TALIPES\_EQUINOVARUS |  | 21 | 0.24 | 1.20 | 0.207 | 1.000 | 1.000 | 28 | tags=43%, list=22%, signal=46% |
| 109 | GOBP\_SEX\_DIFFERENTIATION |  | 28 | 0.21 | 1.20 | 0.230 | 1.000 | 1.000 | 41 | tags=50%, list=33%, signal=58% |
| 110 | HP\_FAILURE\_TO\_THRIVE |  | 36 | 0.20 | 1.20 | 0.209 | 1.000 | 1.000 | 72 | tags=72%, list=57%, signal=120% |
| 111 | HP\_ABNORMALITY\_OF\_THE\_SCROTUM |  | 34 | 0.20 | 1.20 | 0.238 | 1.000 | 1.000 | 36 | tags=44%, list=29%, signal=45% |
| 112 | HP\_ABNORMALITY\_OF\_UPPER\_LIP\_VERMILLION |  | 18 | 0.25 | 1.19 | 0.229 | 1.000 | 1.000 | 35 | tags=50%, list=28%, signal=59% |
| 113 | HP\_ADRENAL\_INSUFFICIENCY |  | 14 | 0.28 | 1.19 | 0.218 | 1.000 | 1.000 | 31 | tags=50%, list=25%, signal=59% |
| 114 | RYBP\_TARGET\_GENES |  | 15 | 0.27 | 1.19 | 0.243 | 1.000 | 1.000 | 45 | tags=60%, list=36%, signal=82% |
| 115 | GOBP\_PROTEIN\_CATABOLIC\_PROCESS |  | 11 | 0.31 | 1.18 | 0.246 | 1.000 | 1.000 | 44 | tags=64%, list=35%, signal=89% |
| 116 | HP\_CUPPED\_EAR |  | 12 | 0.29 | 1.18 | 0.233 | 1.000 | 1.000 | 60 | tags=75%, list=48%, signal=130% |
| 117 | MIR5688 |  | 10 | 0.31 | 1.18 | 0.246 | 1.000 | 1.000 | 51 | tags=70%, list=40%, signal=108% |
| 118 | HP\_ABNORMALITY\_OF\_ADRENAL\_PHYSIOLOGY |  | 14 | 0.28 | 1.18 | 0.244 | 1.000 | 1.000 | 31 | tags=50%, list=25%, signal=59% |
| 119 | HP\_GAIT\_DISTURBANCE |  | 24 | 0.22 | 1.18 | 0.240 | 1.000 | 1.000 | 13 | tags=29%, list=10%, signal=26% |
| 120 | GOCC\_ORGANELLE\_SUBCOMPARTMENT |  | 15 | 0.26 | 1.17 | 0.250 | 1.000 | 1.000 | 12 | tags=33%, list=10%, signal=32% |
| 121 | HP\_ABNORMALITY\_OF\_THE\_CEREBRAL\_SUBCORTEX |  | 40 | 0.19 | 1.17 | 0.253 | 1.000 | 1.000 | 90 | tags=85%, list=71%, signal=203% |
| 122 | HP\_ABNORMAL\_ELASTICITY\_OF\_SKIN |  | 12 | 0.29 | 1.17 | 0.248 | 1.000 | 1.000 | 71 | tags=83%, list=56%, signal=173% |
| 123 | GOBP\_RESPONSE\_TO\_NITROGEN\_COMPOUND |  | 11 | 0.30 | 1.17 | 0.258 | 1.000 | 1.000 | 56 | tags=73%, list=44%, signal=119% |
| 124 | GOBP\_MICROTUBULE\_BASED\_PROCESS |  | 14 | 0.27 | 1.17 | 0.260 | 1.000 | 1.000 | 59 | tags=71%, list=47%, signal=119% |
| 125 | HP\_FLAT\_FACE |  | 13 | 0.28 | 1.16 | 0.275 | 1.000 | 1.000 | 45 | tags=62%, list=36%, signal=86% |
| 126 | HP\_MALE\_PSEUDOHERMAPHRODITISM |  | 16 | 0.25 | 1.16 | 0.275 | 1.000 | 1.000 | 27 | tags=44%, list=21%, signal=49% |
| 127 | GOMF\_TRANSCRIPTION\_COREGULATOR\_ACTIVITY |  | 11 | 0.30 | 1.16 | 0.276 | 1.000 | 1.000 | 45 | tags=64%, list=36%, signal=90% |
| 128 | HP\_ABNORMALITY\_OF\_THE\_NASAL\_DORSUM |  | 23 | 0.22 | 1.16 | 0.270 | 1.000 | 1.000 | 53 | tags=61%, list=42%, signal=86% |
| 129 | HP\_ABNORMAL\_MALE\_REPRODUCTIVE\_SYSTEM\_PHYSIOLOGY |  | 23 | 0.22 | 1.15 | 0.268 | 1.000 | 1.000 | 31 | tags=43%, list=25%, signal=47% |
| 130 | BRUINS\_UVC\_RESPONSE\_VIA\_TP53\_GROUP\_A |  | 11 | 0.29 | 1.14 | 0.285 | 1.000 | 1.000 | 57 | tags=73%, list=45%, signal=121% |
| 131 | HP\_THIN\_UPPER\_LIP\_VERMILION |  | 14 | 0.26 | 1.14 | 0.262 | 1.000 | 1.000 | 33 | tags=50%, list=26%, signal=60% |
| 132 | HP\_ABNORMALITY\_OF\_THE\_CHIN |  | 16 | 0.25 | 1.13 | 0.290 | 1.000 | 1.000 | 19 | tags=38%, list=15%, signal=39% |
| 133 | HP\_JOINT\_HYPERFLEXIBILITY |  | 15 | 0.26 | 1.13 | 0.309 | 1.000 | 1.000 | 54 | tags=67%, list=43%, signal=103% |
| 134 | HP\_ABNORMALITY\_OF\_THE\_HAIRLINE |  | 12 | 0.28 | 1.12 | 0.286 | 1.000 | 1.000 | 83 | tags=92%, list=66%, signal=243% |
| 135 | GOBP\_GAMETE\_GENERATION |  | 15 | 0.26 | 1.12 | 0.300 | 1.000 | 1.000 | 63 | tags=73%, list=50%, signal=129% |
| 136 | HP\_AUTISTIC\_BEHAVIOR |  | 14 | 0.26 | 1.12 | 0.293 | 1.000 | 1.000 | 33 | tags=50%, list=26%, signal=60% |
| 137 | HP\_PECTUS\_EXCAVATUM |  | 19 | 0.23 | 1.11 | 0.309 | 1.000 | 1.000 | 54 | tags=63%, list=43%, signal=94% |
| 138 | GOMF\_SIGNALING\_RECEPTOR\_BINDING |  | 17 | 0.24 | 1.11 | 0.300 | 1.000 | 1.000 | 91 | tags=94%, list=72%, signal=293% |
| 139 | GOBP\_ORGANIC\_HYDROXY\_COMPOUND\_METABOLIC\_PROCESS |  | 15 | 0.25 | 1.11 | 0.327 | 1.000 | 1.000 | 22 | tags=40%, list=17%, signal=43% |
| 140 | GOBP\_CYTOSKELETON\_ORGANIZATION |  | 16 | 0.24 | 1.11 | 0.310 | 1.000 | 1.000 | 59 | tags=69%, list=47%, signal=113% |
| 141 | BLALOCK\_ALZHEIMERS\_DISEASE\_UP |  | 17 | 0.24 | 1.11 | 0.317 | 1.000 | 1.000 | 62 | tags=71%, list=49%, signal=120% |
| 142 | LAKE\_ADULT\_KIDNEY\_C5\_PROXIMAL\_TUBULE\_EPITHELIAL\_CELLS\_STRESS\_INFLAM |  | 10 | 0.29 | 1.11 | 0.330 | 1.000 | 1.000 | 28 | tags=50%, list=22%, signal=59% |
| 143 | HP\_ABNORMAL\_DERMATOGLYPHICS |  | 23 | 0.21 | 1.11 | 0.291 | 1.000 | 1.000 | 65 | tags=70%, list=52%, signal=117% |
| 144 | HP\_ABNORMALITY\_OF\_NEURONAL\_MIGRATION |  | 19 | 0.22 | 1.11 | 0.305 | 1.000 | 1.000 | 101 | tags=100%, list=80%, signal=428% |
| 145 | HP\_ABNORMALITY\_OF\_SKIN\_PIGMENTATION |  | 23 | 0.21 | 1.10 | 0.310 | 1.000 | 1.000 | 43 | tags=52%, list=34%, signal=65% |
| 146 | GOBP\_LOCOMOTION |  | 28 | 0.19 | 1.10 | 0.331 | 1.000 | 1.000 | 61 | tags=64%, list=48%, signal=97% |
| 147 | HP\_HYPERGONADOTROPIC\_HYPOGONADISM |  | 11 | 0.28 | 1.10 | 0.315 | 1.000 | 1.000 | 36 | tags=55%, list=29%, signal=70% |
| 148 | GOBP\_NEGATIVE\_REGULATION\_OF\_NUCLEOBASE\_CONTAINING\_COMPOUND\_METABOLIC\_PROCESS |  | 27 | 0.20 | 1.10 | 0.324 | 1.000 | 1.000 | 31 | tags=41%, list=25%, signal=42% |
| 149 | HP\_INTESTINAL\_MALROTATION |  | 14 | 0.25 | 1.10 | 0.306 | 1.000 | 1.000 | 88 | tags=93%, list=70%, signal=274% |
| 150 | HP\_ABNORMALITY\_OF\_THE\_KNEE |  | 16 | 0.24 | 1.09 | 0.340 | 1.000 | 1.000 | 28 | tags=44%, list=22%, signal=49% |
| 151 | HP\_ABNORMAL\_INFLAMMATORY\_RESPONSE |  | 26 | 0.20 | 1.09 | 0.330 | 1.000 | 1.000 | 76 | tags=77%, list=60%, signal=154% |
| 152 | GOBP\_REGULATION\_OF\_IMMUNE\_SYSTEM\_PROCESS |  | 16 | 0.24 | 1.09 | 0.334 | 1.000 | 1.000 | 20 | tags=38%, list=16%, signal=39% |
| 153 | MIR607 |  | 10 | 0.30 | 1.09 | 0.328 | 1.000 | 1.000 | 40 | tags=60%, list=32%, signal=81% |
| 154 | YCATTAA\_UNKNOWN |  | 11 | 0.28 | 1.08 | 0.337 | 1.000 | 1.000 | 59 | tags=73%, list=47%, signal=125% |
| 155 | GOBP\_ORGANIC\_HYDROXY\_COMPOUND\_BIOSYNTHETIC\_PROCESS |  | 12 | 0.27 | 1.08 | 0.352 | 1.000 | 1.000 | 21 | tags=42%, list=17%, signal=45% |
| 156 | HP\_KYPHOSIS |  | 13 | 0.26 | 1.08 | 0.329 | 1.000 | 1.000 | 57 | tags=69%, list=45%, signal=113% |
| 157 | HP\_GENERALIZED\_HYPOTONIA |  | 26 | 0.20 | 1.08 | 0.359 | 1.000 | 1.000 | 42 | tags=50%, list=33%, signal=60% |
| 158 | REACTOME\_RNA\_POLYMERASE\_II\_TRANSCRIPTION |  | 15 | 0.25 | 1.07 | 0.371 | 1.000 | 1.000 | 39 | tags=53%, list=31%, signal=68% |
| 159 | GOMF\_OXIDOREDUCTASE\_ACTIVITY |  | 12 | 0.26 | 1.06 | 0.347 | 1.000 | 1.000 | 22 | tags=42%, list=17%, signal=46% |
| 160 | ZNF423\_TARGET\_GENES |  | 13 | 0.25 | 1.06 | 0.384 | 1.000 | 1.000 | 48 | tags=62%, list=38%, signal=89% |
| 161 | HP\_SINGLE\_TRANSVERSE\_PALMAR\_CREASE |  | 15 | 0.24 | 1.06 | 0.350 | 1.000 | 1.000 | 65 | tags=73%, list=52%, signal=133% |
| 162 | HP\_ABNORMAL\_CORPUS\_CALLOSUM\_MORPHOLOGY |  | 38 | 0.17 | 1.06 | 0.363 | 1.000 | 1.000 | 90 | tags=84%, list=71%, signal=206% |
| 163 | GOBP\_REGULATION\_OF\_REPRODUCTIVE\_PROCESS |  | 10 | 0.28 | 1.06 | 0.356 | 1.000 | 1.000 | 67 | tags=80%, list=53%, signal=157% |
| 164 | HP\_ABNORMAL\_HEART\_VALVE\_MORPHOLOGY |  | 20 | 0.21 | 1.05 | 0.369 | 1.000 | 1.000 | 52 | tags=60%, list=41%, signal=86% |
| 165 | HP\_ANOSMIA |  | 13 | 0.26 | 1.05 | 0.374 | 1.000 | 1.000 | 67 | tags=77%, list=53%, signal=147% |
| 166 | HP\_ABNORMALLY\_LAX\_OR\_HYPEREXTENSIBLE\_SKIN |  | 11 | 0.27 | 1.05 | 0.390 | 1.000 | 1.000 | 71 | tags=82%, list=56%, signal=171% |
| 167 | HP\_MUSCLE\_WEAKNESS |  | 22 | 0.21 | 1.05 | 0.368 | 1.000 | 1.000 | 75 | tags=77%, list=60%, signal=158% |
| 168 | HP\_PREMATURELY\_AGED\_APPEARANCE |  | 11 | 0.27 | 1.05 | 0.392 | 1.000 | 1.000 | 71 | tags=82%, list=56%, signal=171% |
| 169 | HP\_ABNORMAL\_HAIR\_PATTERN |  | 20 | 0.22 | 1.04 | 0.365 | 1.000 | 1.000 | 39 | tags=50%, list=31%, signal=61% |
| 170 | HP\_ABNORMALITY\_OF\_THE\_CALF |  | 20 | 0.21 | 1.04 | 0.402 | 1.000 | 1.000 | 27 | tags=40%, list=21%, signal=43% |
| 171 | GOBP\_REPRODUCTIVE\_SYSTEM\_DEVELOPMENT |  | 32 | 0.18 | 1.04 | 0.400 | 1.000 | 1.000 | 41 | tags=47%, list=33%, signal=52% |
| 172 | HP\_ABNORMALITY\_OF\_CARDIOVASCULAR\_SYSTEM\_ELECTROPHYSIOLOGY |  | 15 | 0.24 | 1.04 | 0.386 | 1.000 | 1.000 | 90 | tags=93%, list=71%, signal=288% |
| 173 | HP\_DEPRESSED\_NASAL\_RIDGE |  | 12 | 0.26 | 1.04 | 0.365 | 1.000 | 1.000 | 53 | tags=67%, list=42%, signal=104% |
| 174 | HP\_ATTENTION\_DEFICIT\_HYPERACTIVITY\_DISORDER |  | 11 | 0.27 | 1.04 | 0.400 | 1.000 | 1.000 | 37 | tags=55%, list=29%, signal=70% |
| 175 | HP\_ABNORMALITY\_OF\_THE\_FRONTAL\_HAIRLINE |  | 11 | 0.27 | 1.04 | 0.395 | 1.000 | 1.000 | 83 | tags=91%, list=66%, signal=243% |
| 176 | HP\_ABNORMAL\_CIRCULATING\_FOLLICLE\_STIMULATING\_HORMONE\_CONCENTRATION |  | 13 | 0.25 | 1.03 | 0.410 | 1.000 | 1.000 | 10 | tags=31%, list=8%, signal=30% |
| 177 | HP\_ABNORMAL\_PATTERN\_OF\_RESPIRATION |  | 23 | 0.20 | 1.03 | 0.401 | 1.000 | 1.000 | 99 | tags=96%, list=79%, signal=365% |
| 178 | HP\_HEPATOMEGALY |  | 12 | 0.25 | 1.03 | 0.369 | 1.000 | 1.000 | 54 | tags=67%, list=43%, signal=106% |
| 179 | HP\_LARGE\_FACE |  | 12 | 0.25 | 1.03 | 0.377 | 1.000 | 1.000 | 33 | tags=50%, list=26%, signal=61% |
| 180 | HP\_MYOPIA |  | 10 | 0.27 | 1.03 | 0.409 | 1.000 | 1.000 | 68 | tags=80%, list=54%, signal=160% |
| 181 | GOCC\_NUCLEAR\_PROTEIN\_CONTAINING\_COMPLEX |  | 12 | 0.25 | 1.03 | 0.390 | 1.000 | 1.000 | 33 | tags=50%, list=26%, signal=61% |
| 182 | KDM7A\_TARGET\_GENES |  | 13 | 0.25 | 1.03 | 0.400 | 1.000 | 1.000 | 39 | tags=54%, list=31%, signal=70% |
| 183 | HP\_ABNORMAL\_ORAL\_PHYSIOLOGY |  | 12 | 0.25 | 1.03 | 0.393 | 1.000 | 1.000 | 75 | tags=83%, list=60%, signal=186% |
| 184 | MODULE\_220 |  | 15 | 0.23 | 1.02 | 0.399 | 1.000 | 1.000 | 7 | tags=27%, list=6%, signal=25% |
| 185 | HP\_ABNORMAL\_CIRCULATING\_LIPID\_CONCENTRATION |  | 11 | 0.27 | 1.02 | 0.421 | 1.000 | 1.000 | 83 | tags=91%, list=66%, signal=243% |
| 186 | HP\_NEOPLASM\_OF\_THE\_GENITOURINARY\_TRACT |  | 14 | 0.23 | 1.02 | 0.394 | 1.000 | 1.000 | 63 | tags=71%, list=50%, signal=127% |
| 187 | GOBP\_CHROMOSOME\_ORGANIZATION |  | 15 | 0.23 | 1.02 | 0.405 | 1.000 | 1.000 | 32 | tags=47%, list=25%, signal=55% |
| 188 | WTTGKCTG\_UNKNOWN |  | 11 | 0.27 | 1.02 | 0.424 | 1.000 | 1.000 | 83 | tags=91%, list=66%, signal=243% |
| 189 | GOBP\_NEGATIVE\_REGULATION\_OF\_BIOSYNTHETIC\_PROCESS |  | 28 | 0.18 | 1.02 | 0.445 | 1.000 | 1.000 | 31 | tags=39%, list=25%, signal=41% |
| 190 | MIR548AJ\_3P\_MIR548X\_3P |  | 14 | 0.23 | 1.02 | 0.404 | 1.000 | 1.000 | 45 | tags=57%, list=36%, signal=79% |
| 191 | HP\_SLANTING\_OF\_THE\_PALPEBRAL\_FISSURE |  | 52 | 0.15 | 1.01 | 0.402 | 1.000 | 1.000 | 68 | tags=63%, list=54%, signal=81% |
| 192 | GOBP\_MACROMOLECULE\_CATABOLIC\_PROCESS |  | 12 | 0.25 | 1.01 | 0.427 | 1.000 | 1.000 | 44 | tags=58%, list=35%, signal=81% |
| 193 | HP\_INCREASED\_BODY\_WEIGHT |  | 33 | 0.17 | 1.01 | 0.406 | 1.000 | 1.000 | 52 | tags=55%, list=41%, signal=69% |
| 194 | HP\_ABNORMALITY\_OF\_THE\_PHILTRUM |  | 34 | 0.17 | 1.01 | 0.413 | 1.000 | 1.000 | 28 | tags=35%, list=22%, signal=33% |
| 195 | HP\_REGIONAL\_ABNORMALITY\_OF\_SKIN |  | 27 | 0.18 | 1.01 | 0.410 | 1.000 | 1.000 | 65 | tags=67%, list=52%, signal=108% |
| 196 | REACTOME\_DEVELOPMENTAL\_BIOLOGY |  | 18 | 0.21 | 1.01 | 0.417 | 1.000 | 1.000 | 39 | tags=50%, list=31%, signal=62% |
| 197 | HP\_ABNORMALITY\_OF\_THE\_ULNA |  | 18 | 0.21 | 1.01 | 0.432 | 1.000 | 1.000 | 39 | tags=50%, list=31%, signal=62% |
| 198 | HP\_GONADAL\_NEOPLASM |  | 14 | 0.23 | 1.01 | 0.385 | 1.000 | 1.000 | 63 | tags=71%, list=50%, signal=127% |
| 199 | GOBP\_PHOSPHORYLATION |  | 20 | 0.20 | 1.00 | 0.427 | 1.000 | 1.000 | 91 | tags=90%, list=72%, signal=273% |
| 200 | HP\_PROMINENT\_FOREHEAD |  | 15 | 0.22 | 1.00 | 0.438 | 1.000 | 1.000 | 33 | tags=47%, list=26%, signal=56% |
| 201 | HP\_HYPERTELORISM |  | 50 | 0.15 | 1.00 | 0.404 | 1.000 | 1.000 | 28 | tags=32%, list=22%, signal=25% |
| 202 | HP\_CRYPTORCHIDISM |  | 87 | 0.16 | 1.00 | 0.426 | 1.000 | 1.000 | 68 | tags=60%, list=54%, signal=40% |
| 203 | HP\_ABNORMAL\_NIPPLE\_MORPHOLOGY |  | 22 | 0.19 | 1.00 | 0.433 | 1.000 | 1.000 | 65 | tags=68%, list=52%, signal=116% |
| 204 | HP\_APLASIA\_HYPOPLASIA\_INVOLVING\_BONES\_OF\_THE\_THORAX |  | 25 | 0.19 | 1.00 | 0.429 | 1.000 | 1.000 | 96 | tags=92%, list=76%, signal=310% |
| 205 | GOBP\_RESPONSE\_TO\_HORMONE |  | 19 | 0.21 | 0.99 | 0.479 | 1.000 | 1.000 | 63 | tags=68%, list=50%, signal=116% |
| 206 | HP\_GENITAL\_NEOPLASM |  | 14 | 0.23 | 0.99 | 0.416 | 1.000 | 1.000 | 63 | tags=71%, list=50%, signal=127% |
| 207 | ZNF274\_TARGET\_GENES |  | 10 | 0.27 | 0.99 | 0.452 | 1.000 | 1.000 | 56 | tags=70%, list=44%, signal=116% |
| 208 | HP\_OTITIS\_MEDIA |  | 11 | 0.26 | 0.99 | 0.480 | 1.000 | 1.000 | 61 | tags=73%, list=48%, signal=129% |
| 209 | GOBP\_RESPONSE\_TO\_ENDOGENOUS\_STIMULUS |  | 35 | 0.17 | 0.99 | 0.434 | 1.000 | 1.000 | 63 | tags=63%, list=50%, signal=91% |
| 210 | HP\_APLASIA\_HYPOPLASIA\_OF\_THE\_BREASTS |  | 15 | 0.22 | 0.99 | 0.443 | 1.000 | 1.000 | 67 | tags=73%, list=53%, signal=138% |
| 211 | HP\_PUBERTY\_AND\_GONADAL\_DISORDERS |  | 61 | 0.15 | 0.99 | 0.458 | 1.000 | 1.000 | 68 | tags=62%, list=54%, signal=70% |
| 212 | HP\_GASTROESOPHAGEAL\_REFLUX |  | 15 | 0.22 | 0.99 | 0.452 | 1.000 | 1.000 | 42 | tags=53%, list=33%, signal=70% |
| 213 | HP\_GYNECOMASTIA |  | 23 | 0.19 | 0.99 | 0.453 | 1.000 | 1.000 | 12 | tags=26%, list=10%, signal=24% |
| 214 | PAX3\_TARGET\_GENES |  | 16 | 0.21 | 0.98 | 0.464 | 1.000 | 1.000 | 31 | tags=44%, list=25%, signal=51% |
| 215 | HP\_HIGH\_PALATE |  | 39 | 0.15 | 0.98 | 0.443 | 1.000 | 1.000 | 76 | tags=72%, list=60%, signal=125% |
| 216 | GOMF\_CHROMATIN\_BINDING |  | 15 | 0.22 | 0.98 | 0.463 | 1.000 | 1.000 | 33 | tags=47%, list=26%, signal=56% |
| 217 | HP\_BROAD\_FOREHEAD |  | 11 | 0.25 | 0.98 | 0.445 | 1.000 | 1.000 | 96 | tags=100%, list=76%, signal=383% |
| 218 | HP\_ABNORMAL\_SHAPE\_OF\_THE\_FRONTAL\_REGION |  | 27 | 0.18 | 0.98 | 0.454 | 1.000 | 1.000 | 14 | tags=26%, list=11%, signal=23% |
| 219 | HP\_PARAPLEGIA\_PARAPARESIS |  | 12 | 0.24 | 0.97 | 0.467 | 1.000 | 1.000 | 35 | tags=50%, list=28%, signal=63% |
| 220 | HP\_PES\_CAVUS |  | 12 | 0.25 | 0.97 | 0.447 | 1.000 | 1.000 | 13 | tags=33%, list=10%, signal=34% |
| 221 | HP\_ABNORMALITY\_OF\_THE\_TESTIS\_SIZE |  | 28 | 0.17 | 0.97 | 0.506 | 1.000 | 1.000 | 36 | tags=43%, list=29%, signal=47% |
| 222 | HP\_APNEA |  | 13 | 0.23 | 0.97 | 0.466 | 1.000 | 1.000 | 99 | tags=100%, list=79%, signal=419% |
| 223 | HP\_ABNORMAL\_FEMALE\_REPRODUCTIVE\_SYSTEM\_PHYSIOLOGY |  | 39 | 0.16 | 0.97 | 0.479 | 1.000 | 1.000 | 63 | tags=62%, list=50%, signal=85% |
| 224 | HP\_ABNORMALITY\_OF\_REPRODUCTIVE\_SYSTEM\_PHYSIOLOGY |  | 63 | 0.14 | 0.96 | 0.410 | 1.000 | 1.000 | 68 | tags=62%, list=54%, signal=67% |
| 225 | HP\_ABNORMALITY\_OF\_THE\_MENSTRUAL\_CYCLE |  | 38 | 0.15 | 0.96 | 0.500 | 1.000 | 1.000 | 55 | tags=55%, list=44%, signal=68% |
| 226 | HP\_ERECTILE\_DYSFUNCTION |  | 12 | 0.23 | 0.96 | 0.487 | 1.000 | 1.000 | 67 | tags=75%, list=53%, signal=145% |
| 227 | GOBP\_VESICLE\_MEDIATED\_TRANSPORT |  | 13 | 0.23 | 0.96 | 0.514 | 1.000 | 1.000 | 12 | tags=31%, list=10%, signal=30% |
| 228 | HP\_JOINT\_STIFFNESS |  | 11 | 0.24 | 0.96 | 0.495 | 1.000 | 1.000 | 74 | tags=82%, list=59%, signal=181% |
| 229 | REACTOME\_METABOLISM\_OF\_LIPIDS |  | 13 | 0.23 | 0.96 | 0.508 | 1.000 | 1.000 | 22 | tags=38%, list=17%, signal=42% |
| 230 | HP\_ABNORMALITY\_OF\_TIBIA\_MORPHOLOGY |  | 11 | 0.24 | 0.96 | 0.482 | 1.000 | 1.000 | 17 | tags=36%, list=13%, signal=38% |
| 231 | HP\_ABNORMAL\_CIRCULATING\_ESTROGEN\_LEVEL |  | 12 | 0.23 | 0.95 | 0.516 | 1.000 | 1.000 | 36 | tags=50%, list=29%, signal=63% |
| 232 | HP\_DELAYED\_PUBERTY |  | 25 | 0.18 | 0.95 | 0.481 | 1.000 | 1.000 | 67 | tags=68%, list=53%, signal=116% |
| 233 | HP\_LARYNGOMALACIA |  | 11 | 0.25 | 0.95 | 0.501 | 1.000 | 1.000 | 28 | tags=45%, list=22%, signal=53% |
| 234 | GOBP\_REGULATION\_OF\_CELLULAR\_COMPONENT\_MOVEMENT |  | 19 | 0.19 | 0.94 | 0.506 | 1.000 | 1.000 | 91 | tags=89%, list=72%, signal=274% |
| 235 | HP\_ABNORMAL\_SCALP\_MORPHOLOGY |  | 22 | 0.18 | 0.94 | 0.509 | 1.000 | 1.000 | 83 | tags=82%, list=66%, signal=198% |
| 236 | GOBP\_NEGATIVE\_REGULATION\_OF\_CELL\_DEATH |  | 16 | 0.21 | 0.94 | 0.501 | 1.000 | 1.000 | 63 | tags=69%, list=50%, signal=120% |
| 237 | GOBP\_REGULATION\_OF\_PROTEIN\_MODIFICATION\_PROCESS |  | 20 | 0.19 | 0.94 | 0.515 | 1.000 | 1.000 | 61 | tags=65%, list=48%, signal=106% |
| 238 | SMID\_BREAST\_CANCER\_BASAL\_DN |  | 11 | 0.24 | 0.94 | 0.509 | 1.000 | 1.000 | 63 | tags=73%, list=50%, signal=133% |
| 239 | HP\_MALE\_SEXUAL\_DYSFUNCTION |  | 12 | 0.23 | 0.94 | 0.499 | 1.000 | 1.000 | 67 | tags=75%, list=53%, signal=145% |
| 240 | GOBP\_AXON\_DEVELOPMENT |  | 12 | 0.23 | 0.94 | 0.511 | 1.000 | 1.000 | 67 | tags=75%, list=53%, signal=145% |
| 241 | GOBP\_MULTI\_ORGANISM\_REPRODUCTIVE\_PROCESS |  | 16 | 0.21 | 0.94 | 0.537 | 1.000 | 1.000 | 63 | tags=69%, list=50%, signal=120% |
| 242 | HP\_ABNORMAL\_CIRCULATING\_ANDROGEN\_LEVEL |  | 20 | 0.19 | 0.94 | 0.486 | 1.000 | 1.000 | 36 | tags=45%, list=29%, signal=53% |
| 243 | HP\_ABNORMAL\_TENDON\_MORPHOLOGY |  | 35 | 0.16 | 0.93 | 0.519 | 1.000 | 1.000 | 28 | tags=34%, list=22%, signal=32% |
| 244 | GOBP\_CELL\_MORPHOGENESIS |  | 16 | 0.20 | 0.93 | 0.523 | 1.000 | 1.000 | 16 | tags=31%, list=13%, signal=31% |
| 245 | HP\_ABNORMAL\_SEX\_DETERMINATION |  | 12 | 0.23 | 0.93 | 0.531 | 1.000 | 1.000 | 36 | tags=50%, list=29%, signal=63% |
| 246 | HP\_VENTRICULOMEGALY |  | 30 | 0.16 | 0.93 | 0.519 | 1.000 | 1.000 | 17 | tags=27%, list=13%, signal=23% |
| 247 | GOBP\_HORMONE\_METABOLIC\_PROCESS |  | 12 | 0.23 | 0.93 | 0.511 | 1.000 | 1.000 | 57 | tags=67%, list=45%, signal=110% |
| 248 | MORC2\_TARGET\_GENES |  | 11 | 0.24 | 0.93 | 0.507 | 1.000 | 1.000 | 40 | tags=55%, list=32%, signal=73% |
| 249 | HP\_ABNORMALITY\_OF\_THE\_PHARYNX |  | 10 | 0.25 | 0.93 | 0.530 | 1.000 | 1.000 | 33 | tags=50%, list=26%, signal=62% |
| 250 | HP\_VISUAL\_IMPAIRMENT |  | 36 | 0.16 | 0.93 | 0.508 | 1.000 | 1.000 | 27 | tags=33%, list=21%, signal=30% |
| 251 | HP\_ABNORMAL\_AGGRESSIVE\_IMPULSIVE\_OR\_VIOLENT\_BEHAVIOR |  | 10 | 0.25 | 0.93 | 0.517 | 1.000 | 1.000 | 33 | tags=50%, list=26%, signal=62% |
| 252 | HP\_ABNORMALITY\_OF\_THE\_GINGIVA |  | 12 | 0.23 | 0.93 | 0.534 | 1.000 | 1.000 | 25 | tags=42%, list=20%, signal=47% |
| 253 | HP\_INCREASED\_CIRCULATING\_GONADOTROPIN\_LEVEL |  | 15 | 0.21 | 0.92 | 0.519 | 1.000 | 1.000 | 18 | tags=33%, list=14%, signal=34% |
| 254 | HP\_BROAD\_TOE |  | 15 | 0.21 | 0.92 | 0.516 | 1.000 | 1.000 | 60 | tags=67%, list=48%, signal=112% |
| 255 | GOBP\_CELL\_MIGRATION |  | 24 | 0.18 | 0.92 | 0.514 | 1.000 | 1.000 | 23 | tags=33%, list=18%, signal=33% |
| 256 | HOXA2\_TARGET\_GENES |  | 14 | 0.21 | 0.92 | 0.521 | 1.000 | 1.000 | 92 | tags=93%, list=73%, signal=306% |
| 257 | GOBP\_CELLULAR\_MACROMOLECULE\_LOCALIZATION |  | 25 | 0.17 | 0.92 | 0.528 | 1.000 | 1.000 | 68 | tags=68%, list=54%, signal=118% |
| 258 | HP\_HYPERPITUITARISM |  | 15 | 0.21 | 0.92 | 0.515 | 1.000 | 1.000 | 18 | tags=33%, list=14%, signal=34% |
| 259 | HP\_ABNORMAL\_SCAPULA\_MORPHOLOGY |  | 14 | 0.21 | 0.92 | 0.545 | 1.000 | 1.000 | 65 | tags=71%, list=52%, signal=131% |
| 260 | HP\_ABNORMALITY\_OF\_THE\_PULMONARY\_VASCULATURE |  | 11 | 0.24 | 0.91 | 0.529 | 1.000 | 1.000 | 52 | tags=64%, list=41%, signal=99% |
| 261 | HP\_ABNORMAL\_PALMAR\_DERMATOGLYPHICS |  | 19 | 0.19 | 0.91 | 0.567 | 1.000 | 1.000 | 65 | tags=68%, list=52%, signal=120% |
| 262 | GOMF\_TRANSCRIPTION\_REGULATOR\_ACTIVITY |  | 30 | 0.16 | 0.91 | 0.534 | 1.000 | 1.000 | 51 | tags=53%, list=40%, signal=68% |
| 263 | HP\_PATENT\_DUCTUS\_ARTERIOSUS |  | 26 | 0.17 | 0.91 | 0.538 | 1.000 | 1.000 | 60 | tags=62%, list=48%, signal=93% |
| 264 | HP\_ABNORMAL\_CELLULAR\_PHENOTYPE |  | 11 | 0.24 | 0.90 | 0.577 | 1.000 | 1.000 | 52 | tags=64%, list=41%, signal=99% |
| 265 | HP\_ABNORMAL\_ABDOMEN\_MORPHOLOGY |  | 17 | 0.19 | 0.90 | 0.587 | 1.000 | 1.000 | 89 | tags=88%, list=71%, signal=260% |
| 266 | HP\_MICROPENIS |  | 47 | 0.14 | 0.90 | 0.556 | 1.000 | 1.000 | 39 | tags=40%, list=31%, signal=37% |
| 267 | GOMF\_ADENYL\_NUCLEOTIDE\_BINDING |  | 18 | 0.19 | 0.90 | 0.548 | 1.000 | 1.000 | 91 | tags=89%, list=72%, signal=274% |
| 268 | HP\_DECREASED\_TESTICULAR\_SIZE |  | 26 | 0.16 | 0.90 | 0.578 | 1.000 | 1.000 | 31 | tags=38%, list=25%, signal=40% |
| 269 | HP\_ABNORMAL\_INTESTINE\_MORPHOLOGY |  | 35 | 0.15 | 0.90 | 0.567 | 1.000 | 1.000 | 90 | tags=83%, list=71%, signal=209% |
| 270 | GTGCCTT\_MIR506 |  | 13 | 0.22 | 0.90 | 0.553 | 1.000 | 1.000 | 91 | tags=92%, list=72%, signal=298% |
| 271 | GOBP\_HOMEOSTATIC\_PROCESS |  | 20 | 0.18 | 0.89 | 0.569 | 1.000 | 1.000 | 49 | tags=55%, list=39%, signal=76% |
| 272 | GOBP\_REGULATION\_OF\_PHOSPHORUS\_METABOLIC\_PROCESS |  | 18 | 0.19 | 0.89 | 0.504 | 1.000 | 1.000 | 63 | tags=67%, list=50%, signal=114% |
| 273 | HP\_INCREASED\_HEAD\_CIRCUMFERENCE |  | 23 | 0.17 | 0.89 | 0.579 | 1.000 | 1.000 | 91 | tags=87%, list=72%, signal=256% |
| 274 | GOBP\_REGULATION\_OF\_ORGANELLE\_ORGANIZATION |  | 13 | 0.20 | 0.88 | 0.568 | 1.000 | 1.000 | 34 | tags=46%, list=27%, signal=57% |
| 275 | LET\_7A\_3P |  | 10 | 0.23 | 0.88 | 0.580 | 1.000 | 1.000 | 60 | tags=70%, list=48%, signal=123% |
| 276 | LET\_7B\_3P |  | 10 | 0.23 | 0.88 | 0.598 | 1.000 | 1.000 | 60 | tags=70%, list=48%, signal=123% |
| 277 | HP\_ABNORMAL\_FEAR\_ANXIETY\_RELATED\_BEHAVIOR |  | 14 | 0.21 | 0.87 | 0.571 | 1.000 | 1.000 | 30 | tags=43%, list=24%, signal=50% |
| 278 | MIR98\_3P |  | 10 | 0.23 | 0.87 | 0.596 | 1.000 | 1.000 | 60 | tags=70%, list=48%, signal=123% |
| 279 | HP\_HYPERPIGMENTATION\_OF\_THE\_SKIN |  | 18 | 0.19 | 0.87 | 0.588 | 1.000 | 1.000 | 56 | tags=61%, list=44%, signal=94% |
| 280 | HP\_ABNORMALITY\_OF\_COORDINATION |  | 25 | 0.16 | 0.87 | 0.601 | 1.000 | 1.000 | 99 | tags=92%, list=79%, signal=344% |
| 281 | HP\_HYPOGONADISM |  | 48 | 0.13 | 0.87 | 0.596 | 1.000 | 1.000 | 52 | tags=50%, list=41%, signal=53% |
| 282 | HP\_THIN\_VERMILION\_BORDER |  | 22 | 0.17 | 0.87 | 0.617 | 1.000 | 1.000 | 33 | tags=41%, list=26%, signal=46% |
| 283 | GOBP\_STEROID\_METABOLIC\_PROCESS |  | 14 | 0.20 | 0.87 | 0.590 | 1.000 | 1.000 | 22 | tags=36%, list=17%, signal=38% |
| 284 | ZBTB24\_TARGET\_GENES |  | 11 | 0.22 | 0.87 | 0.609 | 1.000 | 1.000 | 88 | tags=91%, list=70%, signal=275% |
| 285 | HP\_APLASIA\_HYPOPLASIA\_OF\_THE\_VAGINA |  | 10 | 0.23 | 0.87 | 0.624 | 1.000 | 1.000 | 10 | tags=30%, list=8%, signal=30% |
| 286 | HP\_BREAST\_HYPOPLASIA |  | 13 | 0.21 | 0.86 | 0.648 | 1.000 | 1.000 | 24 | tags=38%, list=19%, signal=43% |
| 287 | HP\_ABNORMALITY\_OF\_THE\_FOREHEAD |  | 49 | 0.13 | 0.86 | 0.614 | 1.000 | 1.000 | 20 | tags=24%, list=16%, signal=18% |
| 288 | LET\_7F\_1\_3P |  | 10 | 0.23 | 0.86 | 0.629 | 1.000 | 1.000 | 60 | tags=70%, list=48%, signal=123% |
| 289 | HP\_FLEXION\_CONTRACTURE\_OF\_DIGIT |  | 28 | 0.15 | 0.85 | 0.627 | 1.000 | 1.000 | 61 | tags=61%, list=48%, signal=92% |
| 290 | GOBP\_CELL\_MORPHOGENESIS\_INVOLVED\_IN\_DIFFERENTIATION |  | 14 | 0.20 | 0.85 | 0.597 | 1.000 | 1.000 | 67 | tags=71%, list=53%, signal=136% |
| 291 | HP\_TELECANTHUS |  | 19 | 0.17 | 0.85 | 0.629 | 1.000 | 1.000 | 60 | tags=63%, list=48%, signal=102% |
| 292 | HP\_ABNORMALITY\_OF\_THE\_PERIPHERAL\_NERVOUS\_SYSTEM |  | 17 | 0.19 | 0.85 | 0.613 | 1.000 | 1.000 | 90 | tags=88%, list=71%, signal=267% |
| 293 | GOBP\_TRANSMEMBRANE\_RECEPTOR\_PROTEIN\_SERINE\_THREONINE\_KINASE\_SIGNALING\_PATHWAY |  | 11 | 0.22 | 0.85 | 0.644 | 1.000 | 1.000 | 20 | tags=36%, list=16%, signal=39% |
| 294 | HP\_HIGH\_FOREHEAD |  | 16 | 0.19 | 0.85 | 0.646 | 1.000 | 1.000 | 65 | tags=69%, list=52%, signal=124% |
| 295 | MIR3662 |  | 13 | 0.21 | 0.85 | 0.621 | 1.000 | 1.000 | 43 | tags=54%, list=34%, signal=73% |
| 296 | HP\_ABNORMALITY\_OF\_GLOBE\_LOCATION |  | 57 | 0.12 | 0.85 | 0.647 | 1.000 | 1.000 | 28 | tags=30%, list=22%, signal=21% |
| 297 | HP\_ABNORMALITY\_OF\_THE\_SPLEEN |  | 19 | 0.17 | 0.84 | 0.629 | 1.000 | 1.000 | 60 | tags=63%, list=48%, signal=102% |
| 298 | GOBP\_RESPIRATORY\_SYSTEM\_DEVELOPMENT |  | 10 | 0.23 | 0.84 | 0.650 | 1.000 | 1.000 | 61 | tags=70%, list=48%, signal=125% |
| 299 | HP\_ABNORMAL\_EMOTION\_AFFECT\_BEHAVIOR |  | 14 | 0.20 | 0.84 | 0.614 | 1.000 | 1.000 | 76 | tags=79%, list=60%, signal=176% |
| 300 | HP\_LOCALIZED\_SKIN\_LESION |  | 36 | 0.14 | 0.84 | 0.645 | 1.000 | 1.000 | 39 | tags=42%, list=31%, signal=43% |
| 301 | ZBED5\_TARGET\_GENES |  | 10 | 0.23 | 0.84 | 0.674 | 1.000 | 1.000 | 48 | tags=60%, list=38%, signal=89% |
| 302 | HP\_HYPOPLASIA\_OF\_PENIS |  | 63 | 0.13 | 0.84 | 0.555 | 1.000 | 1.000 | 117 | tags=100%, list=93%, signal=700% |
| 303 | HP\_SHORT\_PHILTRUM |  | 19 | 0.18 | 0.84 | 0.669 | 1.000 | 1.000 | 20 | tags=32%, list=16%, signal=32% |
| 304 | HP\_SKELETAL\_DYSPLASIA |  | 16 | 0.18 | 0.84 | 0.648 | 1.000 | 1.000 | 105 | tags=100%, list=83%, signal=524% |
| 305 | GOBP\_TRANSMEMBRANE\_RECEPTOR\_PROTEIN\_TYROSINE\_KINASE\_SIGNALING\_PATHWAY |  | 12 | 0.21 | 0.83 | 0.638 | 1.000 | 1.000 | 91 | tags=92%, list=72%, signal=299% |
| 306 | MIR30D\_5P |  | 11 | 0.21 | 0.83 | 0.657 | 1.000 | 1.000 | 9 | tags=27%, list=7%, signal=27% |
| 307 | HP\_SLEEP\_DISTURBANCE |  | 10 | 0.22 | 0.83 | 0.659 | 1.000 | 1.000 | 99 | tags=100%, list=79%, signal=430% |
| 308 | MIR30B\_5P\_MIR30C\_5P |  | 11 | 0.21 | 0.83 | 0.664 | 1.000 | 1.000 | 9 | tags=27%, list=7%, signal=27% |
| 309 | MIR30A\_5P |  | 11 | 0.21 | 0.83 | 0.654 | 1.000 | 1.000 | 9 | tags=27%, list=7%, signal=27% |
| 310 | MIR30E\_5P |  | 11 | 0.21 | 0.83 | 0.654 | 1.000 | 1.000 | 9 | tags=27%, list=7%, signal=27% |
| 311 | GOBP\_REGULATION\_OF\_TRANSPORT |  | 17 | 0.17 | 0.82 | 0.663 | 1.000 | 1.000 | 17 | tags=29%, list=13%, signal=29% |
| 312 | HP\_PTOSIS |  | 38 | 0.13 | 0.82 | 0.695 | 1.000 | 1.000 | 27 | tags=32%, list=21%, signal=28% |
| 313 | HP\_FEMALE\_SEXUAL\_DYSFUNCTION |  | 10 | 0.22 | 0.82 | 0.687 | 1.000 | 1.000 | 24 | tags=40%, list=19%, signal=45% |
| 314 | HP\_DELAYED\_CRANIAL\_SUTURE\_CLOSURE |  | 12 | 0.20 | 0.82 | 0.683 | 1.000 | 1.000 | 60 | tags=67%, list=48%, signal=115% |
| 315 | GOBP\_ACTIN\_FILAMENT\_BASED\_PROCESS |  | 10 | 0.22 | 0.82 | 0.651 | 1.000 | 1.000 | 11 | tags=30%, list=9%, signal=30% |
| 316 | HP\_UNUSUAL\_INFECTION |  | 27 | 0.15 | 0.82 | 0.677 | 1.000 | 1.000 | 45 | tags=48%, list=36%, signal=59% |
| 317 | HP\_DELAYED\_ERUPTION\_OF\_TEETH |  | 23 | 0.15 | 0.81 | 0.675 | 1.000 | 1.000 | 38 | tags=43%, list=30%, signal=51% |
| 318 | GOBP\_RESPONSE\_TO\_ORGANIC\_CYCLIC\_COMPOUND |  | 12 | 0.20 | 0.81 | 0.653 | 1.000 | 1.000 | 81 | tags=83%, list=64%, signal=211% |
| 319 | GOBP\_CELLULAR\_RESPONSE\_TO\_ENDOGENOUS\_STIMULUS |  | 31 | 0.14 | 0.81 | 0.698 | 1.000 | 1.000 | 63 | tags=61%, list=50%, signal=92% |
| 320 | GOBP\_CELLULAR\_RESPONSE\_TO\_HORMONE\_STIMULUS |  | 15 | 0.18 | 0.81 | 0.683 | 1.000 | 1.000 | 63 | tags=67%, list=50%, signal=117% |
| 321 | HP\_ABNORMAL\_CLAVICLE\_MORPHOLOGY |  | 18 | 0.17 | 0.80 | 0.666 | 1.000 | 1.000 | 79 | tags=78%, list=63%, signal=179% |
| 322 | HP\_ABNORMAL\_CIRCULATING\_SEX\_HORMONE\_CONCENTRATION |  | 22 | 0.16 | 0.80 | 0.714 | 1.000 | 1.000 | 63 | tags=64%, list=50%, signal=105% |
| 323 | CEBPZ\_TARGET\_GENES |  | 12 | 0.20 | 0.80 | 0.718 | 1.000 | 1.000 | 102 | tags=100%, list=81%, signal=475% |
| 324 | HP\_ABNORMAL\_STERNUM\_MORPHOLOGY |  | 28 | 0.14 | 0.80 | 0.708 | 1.000 | 1.000 | 39 | tags=43%, list=31%, signal=48% |
| 325 | HP\_DEATH\_IN\_INFANCY |  | 13 | 0.19 | 0.80 | 0.738 | 1.000 | 1.000 | 94 | tags=92%, list=75%, signal=326% |
| 326 | HP\_ABNORMALITY\_OF\_THE\_FOREARM |  | 23 | 0.15 | 0.80 | 0.711 | 1.000 | 1.000 | 60 | tags=61%, list=48%, signal=95% |
| 327 | HP\_ABNORMAL\_HEART\_VALVE\_PHYSIOLOGY |  | 16 | 0.18 | 0.80 | 0.686 | 1.000 | 1.000 | 90 | tags=88%, list=71%, signal=267% |
| 328 | HP\_PRIMARY\_AMENORRHEA |  | 27 | 0.14 | 0.79 | 0.708 | 1.000 | 1.000 | 36 | tags=41%, list=29%, signal=45% |
| 329 | HP\_SPLENOMEGALY |  | 10 | 0.21 | 0.79 | 0.717 | 1.000 | 1.000 | 88 | tags=90%, list=70%, signal=275% |
| 330 | HP\_ELEVATED\_CIRCULATING\_FOLLICLE\_STIMULATING\_HORMONE\_LEVEL |  | 11 | 0.20 | 0.79 | 0.701 | 1.000 | 1.000 | 10 | tags=27%, list=8%, signal=27% |
| 331 | HP\_LANGUAGE\_IMPAIRMENT |  | 25 | 0.15 | 0.79 | 0.715 | 1.000 | 1.000 | 90 | tags=84%, list=71%, signal=236% |
| 332 | GEORGES\_TARGETS\_OF\_MIR192\_AND\_MIR215 |  | 13 | 0.19 | 0.79 | 0.718 | 1.000 | 1.000 | 75 | tags=77%, list=60%, signal=170% |
| 333 | HP\_GENERALIZED\_ABNORMALITY\_OF\_SKIN |  | 27 | 0.14 | 0.78 | 0.741 | 1.000 | 1.000 | 60 | tags=59%, list=48%, signal=89% |
| 334 | HP\_PARAPLEGIA |  | 11 | 0.21 | 0.78 | 0.761 | 1.000 | 1.000 | 67 | tags=73%, list=53%, signal=142% |
| 335 | GATTGGY\_NFY\_Q6\_01 |  | 15 | 0.17 | 0.78 | 0.742 | 1.000 | 1.000 | 5 | tags=20%, list=4%, signal=18% |
| 336 | GOBP\_MULTICELLULAR\_ORGANISM\_GROWTH |  | 12 | 0.19 | 0.78 | 0.731 | 1.000 | 1.000 | 51 | tags=58%, list=40%, signal=89% |
| 337 | HP\_ABNORMAL\_BLOOD\_GLUCOSE\_CONCENTRATION |  | 11 | 0.20 | 0.77 | 0.766 | 1.000 | 1.000 | 56 | tags=64%, list=44%, signal=105% |
| 338 | GOBP\_REGULATION\_OF\_HORMONE\_LEVELS |  | 15 | 0.17 | 0.77 | 0.751 | 1.000 | 1.000 | 22 | tags=33%, list=17%, signal=36% |
| 339 | GOBP\_REGULATION\_OF\_PROTEIN\_LOCALIZATION |  | 10 | 0.21 | 0.77 | 0.749 | 1.000 | 1.000 | 63 | tags=70%, list=50%, signal=129% |
| 340 | ZNF407\_TARGET\_GENES |  | 11 | 0.20 | 0.77 | 0.745 | 1.000 | 1.000 | 79 | tags=82%, list=63%, signal=200% |
| 341 | HP\_DEPRESSED\_NASAL\_BRIDGE |  | 46 | 0.12 | 0.77 | 0.763 | 1.000 | 1.000 | 28 | tags=30%, list=22%, signal=25% |
| 342 | HP\_SMALL\_FACE |  | 14 | 0.18 | 0.77 | 0.708 | 1.000 | 1.000 | 42 | tags=50%, list=33%, signal=67% |
| 343 | HP\_ABNORMAL\_SYSTEMIC\_ARTERIAL\_MORPHOLOGY |  | 21 | 0.15 | 0.77 | 0.730 | 1.000 | 1.000 | 91 | tags=86%, list=72%, signal=257% |
| 344 | HP\_ABNORMAL\_RENAL\_PELVIS\_MORPHOLOGY |  | 29 | 0.14 | 0.77 | 0.744 | 1.000 | 1.000 | 38 | tags=41%, list=30%, signal=46% |
| 345 | HP\_FEEDING\_DIFFICULTIES |  | 37 | 0.13 | 0.77 | 0.744 | 1.000 | 1.000 | 90 | tags=81%, list=71%, signal=200% |
| 346 | HP\_SYNOSTOSIS\_OF\_JOINTS |  | 20 | 0.16 | 0.77 | 0.741 | 1.000 | 1.000 | 39 | tags=45%, list=31%, signal=55% |
| 347 | GOBP\_ORGANELLE\_LOCALIZATION |  | 11 | 0.20 | 0.77 | 0.743 | 1.000 | 1.000 | 68 | tags=73%, list=54%, signal=144% |
| 348 | HP\_DEPRESSION |  | 12 | 0.19 | 0.77 | 0.736 | 1.000 | 1.000 | 30 | tags=42%, list=24%, signal=49% |
| 349 | GOBP\_ALCOHOL\_METABOLIC\_PROCESS |  | 11 | 0.20 | 0.76 | 0.739 | 1.000 | 1.000 | 22 | tags=36%, list=17%, signal=40% |
| 350 | GOBP\_REPRODUCTION |  | 42 | 0.12 | 0.76 | 0.709 | 1.000 | 1.000 | 34 | tags=36%, list=27%, signal=33% |
| 351 | HP\_ATROPHY\_DEGENERATION\_AFFECTING\_THE\_CENTRAL\_NERVOUS\_SYSTEM |  | 19 | 0.16 | 0.76 | 0.746 | 1.000 | 1.000 | 42 | tags=47%, list=33%, signal=60% |
| 352 | HP\_OSTEOPENIA |  | 14 | 0.18 | 0.76 | 0.742 | 1.000 | 1.000 | 105 | tags=100%, list=83%, signal=533% |
| 353 | GOBP\_CELL\_CELL\_SIGNALING |  | 31 | 0.13 | 0.76 | 0.779 | 1.000 | 1.000 | 11 | tags=19%, list=9%, signal=16% |
| 354 | GOBP\_CELL\_PROJECTION\_ASSEMBLY |  | 19 | 0.15 | 0.76 | 0.782 | 1.000 | 1.000 | 102 | tags=95%, list=81%, signal=422% |
| 355 | NAKAYA\_PLASMACYTOID\_DENDRITIC\_CELL\_FLUMIST\_AGE\_18\_50YO\_7DY\_DN |  | 12 | 0.19 | 0.76 | 0.750 | 1.000 | 1.000 | 9 | tags=25%, list=7%, signal=24% |
| 356 | HP\_SYNOSTOSIS\_INVOLVING\_BONES\_OF\_THE\_UPPER\_LIMBS |  | 15 | 0.17 | 0.76 | 0.767 | 1.000 | 1.000 | 39 | tags=47%, list=31%, signal=60% |
| 357 | GOBP\_EPITHELIAL\_CELL\_DIFFERENTIATION |  | 20 | 0.15 | 0.76 | 0.729 | 1.000 | 1.000 | 27 | tags=35%, list=21%, signal=37% |
| 358 | HP\_ABNORMALITY\_OF\_THE\_PERIORBITAL\_REGION |  | 14 | 0.18 | 0.76 | 0.719 | 1.000 | 1.000 | 60 | tags=64%, list=48%, signal=109% |
| 359 | AACTTT\_UNKNOWN |  | 29 | 0.13 | 0.76 | 0.765 | 1.000 | 1.000 | 73 | tags=69%, list=58%, signal=126% |
| 360 | HP\_ABNORMAL\_PALATE\_MORPHOLOGY |  | 65 | 0.11 | 0.76 | 0.760 | 1.000 | 1.000 | 91 | tags=78%, list=72%, signal=137% |
| 361 | HP\_SCROTAL\_HYPOPLASIA |  | 14 | 0.18 | 0.76 | 0.737 | 1.000 | 1.000 | 33 | tags=43%, list=26%, signal=52% |
| 362 | HP\_CONGENITAL\_MALFORMATION\_OF\_THE\_GREAT\_ARTERIES |  | 31 | 0.13 | 0.76 | 0.769 | 1.000 | 1.000 | 60 | tags=58%, list=48%, signal=84% |
| 363 | HES2\_TARGET\_GENES |  | 11 | 0.20 | 0.76 | 0.762 | 1.000 | 1.000 | 45 | tags=55%, list=36%, signal=77% |
| 364 | GOBP\_NEGATIVE\_REGULATION\_OF\_TRANSCRIPTION\_BY\_RNA\_POLYMERASE\_II |  | 21 | 0.15 | 0.75 | 0.743 | 1.000 | 1.000 | 31 | tags=38%, list=25%, signal=42% |
| 365 | HP\_SKELETAL\_MUSCLE\_HYPERTROPHY |  | 11 | 0.20 | 0.75 | 0.792 | 1.000 | 1.000 | 91 | tags=91%, list=72%, signal=299% |
| 366 | HP\_ABNORMAL\_TESTIS\_MORPHOLOGY |  | 97 | 0.13 | 0.75 | 0.774 | 1.000 | 1.000 | 68 | tags=58%, list=54%, signal=29% |
| 367 | GOBP\_PROTEIN\_LOCALIZATION\_TO\_ORGANELLE |  | 16 | 0.17 | 0.75 | 0.779 | 1.000 | 1.000 | 83 | tags=81%, list=66%, signal=208% |
| 368 | HP\_ABNORMAL\_STOMACH\_MORPHOLOGY |  | 12 | 0.18 | 0.75 | 0.775 | 1.000 | 1.000 | 83 | tags=83%, list=66%, signal=221% |
| 369 | HP\_ABNORMAL\_JOINT\_MORPHOLOGY |  | 50 | 0.11 | 0.75 | 0.761 | 1.000 | 1.000 | 61 | tags=56%, list=48%, signal=65% |
| 370 | ZNF449\_TARGET\_GENES |  | 10 | 0.21 | 0.75 | 0.762 | 1.000 | 1.000 | 13 | tags=30%, list=10%, signal=31% |
| 371 | HP\_ABNORMAL\_SIZE\_OF\_THE\_CLITORIS |  | 21 | 0.14 | 0.75 | 0.764 | 1.000 | 1.000 | 86 | tags=81%, list=68%, signal=213% |
| 372 | MIR302C\_5P |  | 12 | 0.18 | 0.74 | 0.792 | 1.000 | 1.000 | 31 | tags=42%, list=25%, signal=50% |
| 373 | PEREZ\_TP53\_TARGETS |  | 11 | 0.18 | 0.74 | 0.744 | 1.000 | 1.000 | 58 | tags=64%, list=46%, signal=108% |
| 374 | HP\_INVOLUNTARY\_MOVEMENTS |  | 22 | 0.14 | 0.74 | 0.795 | 1.000 | 1.000 | 99 | tags=91%, list=79%, signal=350% |
| 375 | HP\_ABNORMALITY\_OF\_THE\_ADRENAL\_GLANDS |  | 22 | 0.14 | 0.74 | 0.799 | 1.000 | 1.000 | 13 | tags=23%, list=10%, signal=21% |
| 376 | HP\_ABNORMALITY\_OF\_THE\_AUTONOMIC\_NERVOUS\_SYSTEM |  | 14 | 0.17 | 0.74 | 0.762 | 1.000 | 1.000 | 34 | tags=43%, list=27%, signal=52% |
| 377 | HP\_HYPERREFLEXIA |  | 11 | 0.19 | 0.74 | 0.804 | 1.000 | 1.000 | 92 | tags=91%, list=73%, signal=307% |
| 378 | HP\_APLASIA\_HYPOPLASIA\_OF\_THE\_THUMB |  | 15 | 0.17 | 0.74 | 0.776 | 1.000 | 1.000 | 73 | tags=73%, list=58%, signal=154% |
| 379 | GOCC\_NUCLEAR\_OUTER\_MEMBRANE\_ENDOPLASMIC\_RETICULUM\_MEMBRANE\_NETWORK |  | 13 | 0.18 | 0.74 | 0.799 | 1.000 | 1.000 | 8 | tags=23%, list=6%, signal=22% |
| 380 | BENPORATH\_SUZ12\_TARGETS |  | 14 | 0.17 | 0.73 | 0.761 | 1.000 | 1.000 | 7 | tags=21%, list=6%, signal=20% |
| 381 | HP\_ABNORMAL\_CEREBRAL\_VENTRICLE\_MORPHOLOGY |  | 41 | 0.11 | 0.73 | 0.816 | 1.000 | 1.000 | 20 | tags=24%, list=16%, signal=20% |
| 382 | HP\_BIFID\_UVULA |  | 16 | 0.16 | 0.73 | 0.785 | 1.000 | 1.000 | 5 | tags=19%, list=4%, signal=17% |
| 383 | HP\_LIMITATION\_OF\_JOINT\_MOBILITY |  | 21 | 0.14 | 0.73 | 0.785 | 1.000 | 1.000 | 74 | tags=71%, list=59%, signal=144% |
| 384 | GOBP\_SMALL\_MOLECULE\_BIOSYNTHETIC\_PROCESS |  | 12 | 0.18 | 0.73 | 0.775 | 1.000 | 1.000 | 21 | tags=33%, list=17%, signal=36% |
| 385 | RNGTGGGC\_UNKNOWN |  | 14 | 0.17 | 0.73 | 0.796 | 1.000 | 1.000 | 61 | tags=64%, list=48%, signal=111% |
| 386 | GOBP\_REGULATION\_OF\_RESPONSE\_TO\_STRESS |  | 15 | 0.16 | 0.73 | 0.794 | 1.000 | 1.000 | 82 | tags=80%, list=65%, signal=202% |
| 387 | HP\_ABNORMAL\_IMMUNE\_SYSTEM\_MORPHOLOGY |  | 14 | 0.17 | 0.73 | 0.785 | 1.000 | 1.000 | 52 | tags=57%, list=41%, signal=86% |
| 388 | HP\_ABNORMALITY\_OF\_THE\_CURVATURE\_OF\_THE\_VERTEBRAL\_COLUMN |  | 37 | 0.12 | 0.73 | 0.828 | 1.000 | 1.000 | 60 | tags=57%, list=48%, signal=77% |
| 389 | GOBP\_NEGATIVE\_REGULATION\_OF\_DEVELOPMENTAL\_PROCESS |  | 18 | 0.15 | 0.72 | 0.809 | 1.000 | 1.000 | 67 | tags=67%, list=53%, signal=122% |
| 390 | HP\_ATROPHY\_DEGENERATION\_AFFECTING\_THE\_CEREBRUM |  | 17 | 0.15 | 0.72 | 0.814 | 1.000 | 1.000 | 42 | tags=47%, list=33%, signal=61% |
| 391 | HP\_INCREASED\_SUSCEPTIBILITY\_TO\_FRACTURES |  | 11 | 0.19 | 0.72 | 0.808 | 1.000 | 1.000 | 12 | tags=27%, list=10%, signal=28% |
| 392 | HP\_PULMONARY\_HYPOPLASIA |  | 20 | 0.15 | 0.72 | 0.839 | 1.000 | 1.000 | 97 | tags=90%, list=77%, signal=329% |
| 393 | GOBP\_DEVELOPMENTAL\_PROCESS\_INVOLVED\_IN\_REPRODUCTION |  | 36 | 0.12 | 0.72 | 0.817 | 1.000 | 1.000 | 41 | tags=42%, list=33%, signal=44% |
| 394 | HP\_RECURRENT\_FRACTURES |  | 11 | 0.19 | 0.71 | 0.813 | 1.000 | 1.000 | 12 | tags=27%, list=10%, signal=28% |
| 395 | HP\_CONSTIPATION |  | 13 | 0.17 | 0.71 | 0.845 | 1.000 | 1.000 | 57 | tags=62%, list=45%, signal=101% |
| 396 | HP\_ABNORMAL\_AORTIC\_MORPHOLOGY |  | 18 | 0.15 | 0.71 | 0.781 | 1.000 | 1.000 | 109 | tags=100%, list=87%, signal=635% |
| 397 | HP\_ABNORMALITY\_OF\_LOWER\_LIP |  | 12 | 0.18 | 0.71 | 0.815 | 1.000 | 1.000 | 42 | tags=50%, list=33%, signal=68% |
| 398 | HP\_APLASIA\_HYPOPLASIA\_OF\_THE\_RIBS |  | 13 | 0.17 | 0.71 | 0.850 | 1.000 | 1.000 | 96 | tags=92%, list=76%, signal=348% |
| 399 | HP\_ABNORMAL\_SKIN\_MORPHOLOGY\_OF\_THE\_PALM |  | 22 | 0.14 | 0.71 | 0.812 | 1.000 | 1.000 | 65 | tags=64%, list=52%, signal=108% |
| 400 | GOBP\_PEPTIDYL\_AMINO\_ACID\_MODIFICATION |  | 18 | 0.15 | 0.71 | 0.790 | 1.000 | 1.000 | 60 | tags=61%, list=48%, signal=100% |
| 401 | NFE2L1\_TARGET\_GENES |  | 10 | 0.19 | 0.70 | 0.813 | 1.000 | 1.000 | 78 | tags=80%, list=62%, signal=193% |
| 402 | FOXP2\_TARGET\_GENES |  | 12 | 0.18 | 0.70 | 0.822 | 1.000 | 1.000 | 42 | tags=50%, list=33%, signal=68% |
| 403 | GOBP\_POSITIVE\_REGULATION\_OF\_LOCOMOTION |  | 10 | 0.18 | 0.70 | 0.813 | 1.000 | 1.000 | 91 | tags=90%, list=72%, signal=298% |
| 404 | HP\_ABNORMALITY\_OF\_THE\_METAPHYSIS |  | 12 | 0.18 | 0.70 | 0.826 | 1.000 | 1.000 | 105 | tags=100%, list=83%, signal=543% |
| 405 | HP\_ABNORMAL\_ORAL\_MUCOSA\_MORPHOLOGY |  | 15 | 0.16 | 0.70 | 0.821 | 1.000 | 1.000 | 15 | tags=27%, list=12%, signal=27% |
| 406 | HP\_RETROGNATHIA |  | 19 | 0.14 | 0.70 | 0.813 | 1.000 | 1.000 | 30 | tags=37%, list=24%, signal=41% |
| 407 | GOBP\_LIPID\_METABOLIC\_PROCESS |  | 20 | 0.14 | 0.70 | 0.828 | 1.000 | 1.000 | 22 | tags=30%, list=17%, signal=31% |
| 408 | HP\_WEAKNESS\_DUE\_TO\_UPPER\_MOTOR\_NEURON\_DYSFUNCTION |  | 14 | 0.16 | 0.70 | 0.830 | 1.000 | 1.000 | 35 | tags=43%, list=28%, signal=53% |
| 409 | HP\_ABNORMALITY\_OF\_THE\_VOICE |  | 25 | 0.13 | 0.69 | 0.841 | 1.000 | 1.000 | 77 | tags=72%, list=61%, signal=148% |
| 410 | HP\_POLYHYDRAMNIOS |  | 17 | 0.15 | 0.69 | 0.867 | 1.000 | 1.000 | 57 | tags=59%, list=45%, signal=93% |
| 411 | HP\_ABNORMAL\_UVULA\_MORPHOLOGY |  | 18 | 0.14 | 0.68 | 0.858 | 1.000 | 1.000 | 5 | tags=17%, list=4%, signal=15% |
| 412 | HP\_DOLICHOCEPHALY |  | 10 | 0.18 | 0.68 | 0.874 | 1.000 | 1.000 | 104 | tags=100%, list=83%, signal=527% |
| 413 | WP\_MALIGNANT\_PLEURAL\_MESOTHELIOMA |  | 10 | 0.18 | 0.68 | 0.851 | 1.000 | 1.000 | 91 | tags=90%, list=72%, signal=298% |
| 414 | HP\_CLEFT\_SOFT\_PALATE |  | 17 | 0.15 | 0.68 | 0.872 | 1.000 | 1.000 | 5 | tags=18%, list=4%, signal=16% |
| 415 | GOCC\_GOLGI\_APPARATUS |  | 18 | 0.14 | 0.68 | 0.861 | 1.000 | 1.000 | 12 | tags=22%, list=10%, signal=21% |
| 416 | HP\_ABNORMALITY\_OF\_THE\_SYNOVIA |  | 21 | 0.13 | 0.67 | 0.837 | 1.000 | 1.000 | 3 | tags=14%, list=2%, signal=12% |
| 417 | GOBP\_INTRACELLULAR\_PROTEIN\_TRANSPORT |  | 10 | 0.18 | 0.67 | 0.856 | 1.000 | 1.000 | 104 | tags=100%, list=83%, signal=527% |
| 418 | HP\_ABNORMALITY\_OF\_THE\_ANTERIOR\_FONTANELLE |  | 16 | 0.15 | 0.67 | 0.862 | 1.000 | 1.000 | 38 | tags=44%, list=30%, signal=55% |
| 419 | HP\_HIRSUTISM |  | 26 | 0.12 | 0.67 | 0.854 | 1.000 | 1.000 | 108 | tags=96%, list=86%, signal=534% |
| 420 | GOCC\_CENTROSOME |  | 15 | 0.15 | 0.66 | 0.854 | 1.000 | 1.000 | 100 | tags=93%, list=79%, signal=398% |
| 421 | HP\_HYPOSMIA |  | 10 | 0.17 | 0.66 | 0.858 | 1.000 | 1.000 | 67 | tags=70%, list=53%, signal=138% |
| 422 | ASH1L\_TARGET\_GENES |  | 14 | 0.15 | 0.66 | 0.872 | 1.000 | 1.000 | 99 | tags=93%, list=79%, signal=385% |
| 423 | NUYTTEN\_NIPP1\_TARGETS\_UP |  | 10 | 0.18 | 0.66 | 0.869 | 1.000 | 1.000 | 54 | tags=60%, list=43%, signal=97% |
| 424 | GOBP\_RESPONSE\_TO\_LIPID |  | 14 | 0.15 | 0.66 | 0.870 | 1.000 | 1.000 | 63 | tags=64%, list=50%, signal=114% |
| 425 | HP\_ABNORMAL\_PENIS\_MORPHOLOGY |  | 93 | 0.11 | 0.66 | 0.885 | 1.000 | 1.000 | 120 | tags=99%, list=95%, signal=544% |
| 426 | GOBP\_RESPONSE\_TO\_GROWTH\_FACTOR |  | 18 | 0.14 | 0.66 | 0.886 | 1.000 | 1.000 | 61 | tags=61%, list=48%, signal=102% |
| 427 | HP\_CONVEX\_NASAL\_RIDGE |  | 11 | 0.17 | 0.66 | 0.864 | 1.000 | 1.000 | 60 | tags=64%, list=48%, signal=111% |
| 428 | HP\_ICHTHYOSIS |  | 10 | 0.17 | 0.65 | 0.879 | 1.000 | 1.000 | 105 | tags=100%, list=83%, signal=552% |
| 429 | GOBP\_RESPONSE\_TO\_TRANSFORMING\_GROWTH\_FACTOR\_BETA |  | 10 | 0.18 | 0.65 | 0.864 | 1.000 | 1.000 | 29 | tags=40%, list=23%, signal=48% |
| 430 | HP\_RECURRENT\_RESPIRATORY\_INFECTIONS |  | 14 | 0.15 | 0.65 | 0.862 | 1.000 | 1.000 | 45 | tags=50%, list=36%, signal=69% |
| 431 | HP\_ABNORMAL\_HAIR\_QUANTITY |  | 67 | 0.10 | 0.65 | 0.875 | 1.000 | 1.000 | 76 | tags=66%, list=60%, signal=77% |
| 432 | HP\_APLASIA\_HYPOPLASIA\_OF\_THE\_EYEBROW |  | 11 | 0.17 | 0.65 | 0.868 | 1.000 | 1.000 | 83 | tags=82%, list=66%, signal=219% |
| 433 | HP\_LOWER\_EXTREMITY\_JOINT\_DISLOCATION |  | 12 | 0.16 | 0.65 | 0.893 | 1.000 | 1.000 | 65 | tags=67%, list=52%, signal=125% |
| 434 | GOBP\_ESTABLISHMENT\_OF\_PROTEIN\_LOCALIZATION |  | 19 | 0.13 | 0.65 | 0.886 | 1.000 | 1.000 | 104 | tags=95%, list=83%, signal=461% |
| 435 | GOBP\_CELL\_CELL\_ADHESION |  | 11 | 0.16 | 0.65 | 0.878 | 1.000 | 1.000 | 3 | tags=18%, list=2%, signal=17% |
| 436 | HP\_ABNORMAL\_HAIR\_MORPHOLOGY |  | 87 | 0.11 | 0.64 | 0.891 | 1.000 | 1.000 | 76 | tags=64%, list=60%, signal=50% |
| 437 | HP\_AMENORRHEA |  | 31 | 0.11 | 0.64 | 0.906 | 1.000 | 1.000 | 13 | tags=19%, list=10%, signal=16% |
| 438 | HP\_ABNORMALITY\_OF\_THE\_LYMPHATIC\_SYSTEM |  | 24 | 0.12 | 0.64 | 0.922 | 1.000 | 1.000 | 76 | tags=71%, list=60%, signal=145% |
| 439 | HP\_HIP\_DISLOCATION |  | 11 | 0.16 | 0.64 | 0.877 | 1.000 | 1.000 | 3 | tags=18%, list=2%, signal=17% |
| 440 | GOBP\_REGULATION\_OF\_CELLULAR\_RESPONSE\_TO\_STRESS |  | 10 | 0.17 | 0.64 | 0.897 | 1.000 | 1.000 | 67 | tags=70%, list=53%, signal=138% |
| 441 | HP\_MIDFACE\_RETRUSION |  | 23 | 0.12 | 0.64 | 0.909 | 1.000 | 1.000 | 96 | tags=87%, list=76%, signal=299% |
| 442 | GOBP\_AMEBOIDAL\_TYPE\_CELL\_MIGRATION |  | 10 | 0.17 | 0.63 | 0.902 | 1.000 | 1.000 | 67 | tags=70%, list=53%, signal=138% |
| 443 | HP\_ABNORMAL\_PERIPHERAL\_NERVOUS\_SYSTEM\_MORPHOLOGY |  | 10 | 0.17 | 0.63 | 0.912 | 1.000 | 1.000 | 105 | tags=100%, list=83%, signal=552% |
| 444 | HP\_POSTNATAL\_GROWTH\_RETARDATION |  | 18 | 0.13 | 0.63 | 0.901 | 1.000 | 1.000 | 27 | tags=33%, list=21%, signal=36% |
| 445 | GOBP\_REGULATION\_OF\_PROTEIN\_PHOSPHORYLATION |  | 14 | 0.14 | 0.63 | 0.913 | 1.000 | 1.000 | 91 | tags=86%, list=72%, signal=274% |
| 446 | HP\_WIDE\_MOUTH |  | 13 | 0.15 | 0.62 | 0.929 | 1.000 | 1.000 | 40 | tags=46%, list=32%, signal=61% |
| 447 | HP\_ABNORMALITY\_OF\_FONTANELLES |  | 23 | 0.12 | 0.62 | 0.912 | 1.000 | 1.000 | 91 | tags=83%, list=72%, signal=243% |
| 448 | HP\_ABNORMAL\_CARDIAC\_ATRIUM\_MORPHOLOGY |  | 36 | 0.10 | 0.62 | 0.915 | 1.000 | 1.000 | 60 | tags=56%, list=48%, signal=76% |
| 449 | HP\_NARROW\_PALATE |  | 16 | 0.14 | 0.62 | 0.909 | 1.000 | 1.000 | 39 | tags=44%, list=31%, signal=55% |
| 450 | HP\_ABNORMAL\_NASAL\_BRIDGE\_MORPHOLOGY |  | 73 | 0.09 | 0.62 | 0.919 | 1.000 | 1.000 | 58 | tags=51%, list=46%, signal=40% |
| 451 | HP\_FUNCTIONAL\_ABNORMALITY\_OF\_THE\_GASTROINTESTINAL\_TRACT |  | 27 | 0.11 | 0.61 | 0.930 | 1.000 | 1.000 | 77 | tags=70%, list=61%, signal=142% |
| 452 | HP\_ABNORMAL\_MORPHOLOGY\_OF\_THE\_GREAT\_VESSELS |  | 36 | 0.10 | 0.61 | 0.917 | 1.000 | 1.000 | 39 | tags=39%, list=31%, signal=40% |
| 453 | HP\_DENTAL\_MALOCCLUSION |  | 15 | 0.14 | 0.60 | 0.919 | 1.000 | 1.000 | 76 | tags=73%, list=60%, signal=163% |
| 454 | HP\_TOOTH\_MALPOSITION |  | 29 | 0.10 | 0.60 | 0.943 | 1.000 | 1.000 | 76 | tags=69%, list=60%, signal=134% |
| 455 | HP\_THORACIC\_HYPOPLASIA |  | 16 | 0.13 | 0.60 | 0.933 | 1.000 | 1.000 | 79 | tags=75%, list=63%, signal=176% |
| 456 | GGATTA\_PITX2\_Q2 |  | 11 | 0.15 | 0.60 | 0.929 | 1.000 | 1.000 | 50 | tags=55%, list=40%, signal=83% |
| 457 | HP\_ABNORMALITY\_OF\_THE\_TARSAL\_BONES |  | 12 | 0.15 | 0.59 | 0.946 | 1.000 | 1.000 | 108 | tags=100%, list=86%, signal=633% |
| 458 | HP\_ABNORMAL\_ESOPHAGUS\_PHYSIOLOGY |  | 27 | 0.11 | 0.59 | 0.929 | 1.000 | 1.000 | 77 | tags=70%, list=61%, signal=142% |
| 459 | HP\_ABNORMAL\_CONNECTION\_OF\_THE\_CARDIAC\_SEGMENTS |  | 13 | 0.14 | 0.59 | 0.948 | 1.000 | 1.000 | 109 | tags=100%, list=87%, signal=665% |
| 460 | HP\_ABNORMAL\_DIAPHYSIS\_MORPHOLOGY |  | 26 | 0.11 | 0.59 | 0.921 | 1.000 | 1.000 | 27 | tags=31%, list=21%, signal=31% |
| 461 | HP\_COARCTATION\_OF\_AORTA |  | 11 | 0.15 | 0.59 | 0.949 | 1.000 | 1.000 | 39 | tags=45%, list=31%, signal=60% |
| 462 | GOBP\_CELL\_MORPHOGENESIS\_INVOLVED\_IN\_NEURON\_DIFFERENTIATION |  | 12 | 0.14 | 0.58 | 0.936 | 1.000 | 1.000 | 67 | tags=67%, list=53%, signal=129% |
| 463 | HP\_PROPTOSIS |  | 21 | 0.11 | 0.58 | 0.933 | 1.000 | 1.000 | 65 | tags=62%, list=52%, signal=107% |
| 464 | ZNF596\_TARGET\_GENES |  | 10 | 0.16 | 0.58 | 0.944 | 1.000 | 1.000 | 107 | tags=100%, list=85%, signal=611% |
| 465 | HP\_ABNORMAL\_DISTAL\_PHALANX\_MORPHOLOGY\_OF\_FINGER |  | 14 | 0.13 | 0.58 | 0.938 | 1.000 | 1.000 | 20 | tags=29%, list=16%, signal=30% |
| 466 | GOBP\_REGULATION\_OF\_RESPONSE\_TO\_EXTERNAL\_STIMULUS |  | 13 | 0.14 | 0.58 | 0.960 | 1.000 | 1.000 | 90 | tags=85%, list=71%, signal=266% |
| 467 | HP\_DYSARTHRIA |  | 17 | 0.13 | 0.58 | 0.941 | 1.000 | 1.000 | 15 | tags=24%, list=12%, signal=23% |
| 468 | HP\_NEPHROTIC\_SYNDROME |  | 18 | 0.12 | 0.58 | 0.934 | 1.000 | 1.000 | 49 | tags=50%, list=39%, signal=70% |
| 469 | GOBP\_NEGATIVE\_REGULATION\_OF\_MULTICELLULAR\_ORGANISMAL\_PROCESS |  | 17 | 0.12 | 0.57 | 0.959 | 1.000 | 1.000 | 67 | tags=65%, list=53%, signal=120% |
| 470 | GOBP\_POSITIVE\_REGULATION\_OF\_CELLULAR\_COMPONENT\_ORGANIZATION |  | 13 | 0.14 | 0.57 | 0.964 | 1.000 | 1.000 | 3 | tags=15%, list=2%, signal=14% |
| 471 | HP\_ABNORMAL\_PHALANGEAL\_JOINT\_MORPHOLOGY\_OF\_THE\_HAND |  | 17 | 0.12 | 0.57 | 0.947 | 1.000 | 1.000 | 60 | tags=59%, list=48%, signal=97% |
| 472 | HP\_ABNORMALITY\_OF\_THE\_CALVARIA |  | 56 | 0.09 | 0.57 | 0.948 | 1.000 | 1.000 | 110 | tags=93%, list=87%, signal=406% |
| 473 | HP\_NAUSEA\_AND\_VOMITING |  | 11 | 0.15 | 0.57 | 0.950 | 1.000 | 1.000 | 5 | tags=18%, list=4%, signal=17% |
| 474 | HP\_ABNORMAL\_JAW\_MORPHOLOGY |  | 56 | 0.09 | 0.57 | 0.957 | 1.000 | 1.000 | 20 | tags=21%, list=16%, signal=14% |
| 475 | HP\_ABNORMAL\_RESPIRATORY\_SYSTEM\_MORPHOLOGY |  | 56 | 0.09 | 0.56 | 0.939 | 1.000 | 1.000 | 119 | tags=100%, list=94%, signal=1000% |
| 476 | HP\_AUTISM |  | 10 | 0.15 | 0.56 | 0.968 | 1.000 | 1.000 | 57 | tags=60%, list=45%, signal=101% |
| 477 | TATAAA\_TATA\_01 |  | 12 | 0.14 | 0.56 | 0.948 | 1.000 | 1.000 | 67 | tags=67%, list=53%, signal=129% |
| 478 | HP\_HYPOTELORISM |  | 10 | 0.15 | 0.56 | 0.952 | 1.000 | 1.000 | 95 | tags=90%, list=75%, signal=337% |
| 479 | HP\_ABNORMALITY\_OF\_THE\_FONTANELLES\_OR\_CRANIAL\_SUTURES |  | 35 | 0.09 | 0.56 | 0.960 | 1.000 | 1.000 | 5 | tags=11%, list=4%, signal=9% |
| 480 | HP\_APLASIA\_HYPOPLASIA\_OF\_THE\_DISTAL\_PHALANGES\_OF\_THE\_HAND |  | 11 | 0.14 | 0.55 | 0.951 | 1.000 | 1.000 | 17 | tags=27%, list=13%, signal=29% |
| 481 | MIR374A\_5P |  | 12 | 0.14 | 0.55 | 0.960 | 1.000 | 1.000 | 15 | tags=25%, list=12%, signal=26% |
| 482 | GOBP\_REGULATION\_OF\_CELL\_CYCLE |  | 15 | 0.12 | 0.54 | 0.987 | 1.000 | 1.000 | 61 | tags=60%, list=48%, signal=102% |
| 483 | GOBP\_GENITALIA\_DEVELOPMENT |  | 11 | 0.14 | 0.53 | 0.977 | 1.000 | 1.000 | 6 | tags=18%, list=5%, signal=17% |
| 484 | HP\_SHORT\_RIBS |  | 10 | 0.14 | 0.53 | 0.976 | 1.000 | 1.000 | 96 | tags=90%, list=76%, signal=348% |
| 485 | HP\_ABNORMAL\_EXTERNAL\_GENITALIA |  | 113 | 0.13 | 0.53 | 0.975 | 1.000 | 1.000 | 120 | tags=97%, list=95%, signal=211% |
| 486 | GOCC\_MICROTUBULE\_CYTOSKELETON |  | 25 | 0.10 | 0.53 | 0.978 | 1.000 | 1.000 | 100 | tags=88%, list=79%, signal=342% |
| 487 | TGGAAA\_NFAT\_Q4\_01 |  | 25 | 0.10 | 0.53 | 0.982 | 1.000 | 1.000 | 85 | tags=76%, list=67%, signal=187% |
| 488 | HP\_CONTRACTURES\_OF\_THE\_JOINTS\_OF\_THE\_LOWER\_LIMBS |  | 10 | 0.14 | 0.52 | 0.983 | 1.000 | 1.000 | 71 | tags=70%, list=56%, signal=148% |
| 489 | GOMF\_RIBONUCLEOTIDE\_BINDING |  | 21 | 0.10 | 0.52 | 0.985 | 1.000 | 1.000 | 108 | tags=95%, list=86%, signal=556% |
| 490 | HP\_ABNORMALITY\_OF\_THE\_EXTERNAL\_NOSE |  | 53 | 0.08 | 0.52 | 0.981 | 0.998 | 1.000 | 60 | tags=53%, list=48%, signal=58% |
| 491 | HP\_WIDE\_ANTERIOR\_FONTANEL |  | 15 | 0.12 | 0.51 | 0.976 | 0.998 | 1.000 | 3 | tags=13%, list=2%, signal=12% |
| 492 | HP\_APLASIA\_HYPOPLASIA\_OF\_THE\_OVARY |  | 31 | 0.09 | 0.50 | 0.998 | 0.999 | 1.000 | 68 | tags=61%, list=54%, signal=100% |
| 493 | HP\_JOINT\_CONTRACTURE\_OF\_THE\_HAND |  | 21 | 0.10 | 0.49 | 0.990 | 0.998 | 1.000 | 61 | tags=57%, list=48%, signal=92% |
| 494 | HP\_ABNORMAL\_VASCULAR\_MORPHOLOGY |  | 41 | 0.07 | 0.48 | 0.998 | 0.999 | 1.000 | 91 | tags=78%, list=72%, signal=190% |
| 495 | HP\_ABNORMAL\_LOCATION\_OF\_EARS |  | 62 | 0.07 | 0.44 | 0.992 | 1.000 | 1.000 | 5 | tags=8%, list=4%, signal=4% |
| 496 | GOMF\_IDENTICAL\_PROTEIN\_BINDING |  | 14 | 0.10 | 0.42 | 0.998 | 1.000 | 1.000 | 51 | tags=50%, list=40%, signal=75% |
| 497 | HP\_ABNORMAL\_OVARIAN\_MORPHOLOGY |  | 37 | 0.07 | 0.41 | 1.000 | 1.000 | 1.000 | 68 | tags=59%, list=54%, signal=91% |
| 498 | GOCC\_MICROTUBULE\_ORGANIZING\_CENTER |  | 21 | 0.07 | 0.33 | 1.000 | 1.000 | 1.000 | 34 | tags=33%, list=27%, signal=38% |
Table: Gene sets enriched in phenotype **na **[plain text format]****

  
