## Supplemental File 3 for "Increased Expression of *ZFPM2* Bypasses *SRY* to Drive 46,XX Testicular Development: A New Mechanism of 46,XX DSD": HP_ABNORMAL_CEREBELLAR_VERMIS_MORPHOLOGY.html

Details for gene set HP\_ABNORMAL\_CEREBELLAR\_VERMIS\_MORPHOLOGY[GSEA]

|  || Dataset | SexDevelopmentGenesPRL\_remapped |
| Phenotype | NoPhenotypeAvailable |
| Upregulated in class | na\_neg |
| GeneSet | HP\_ABNORMAL\_CEREBELLAR\_VERMIS\_MORPHOLOGY |
| Enrichment Score (ES) | -0.34285715 |
| Normalized Enrichment Score (NES) | -1.7424909 |
| Nominal p-value | 0.027667984 |
| FDR q-value | 0.53627074 |
| FWER p-Value | 1.0 |
Table: GSEA Results Summary

  

Fig 1: Enrichment plot: HP\_ABNORMAL\_CEREBELLAR\_VERMIS\_MORPHOLOGY      
 Profile of the Running ES Score & Positions of GeneSet Members on the Rank Ordered List

  

| SYMBOL | TITLE | RANK IN GENE LIST | RANK METRIC SCORE | RUNNING ES | CORE ENRICHMENT || 1 | ZEB2 | zinc finger E-box binding homeobox 2 [Source:HGNC Symbol;Acc:HGNC:14881] | 5 | 36.500 | -0.0000 | No |
| 2 | OPHN1 | oligophrenin 1 [Source:HGNC Symbol;Acc:HGNC:8148] | 11 | 15.900 | -0.0000 | No |
| 3 | BCOR | BCL6 corepressor [Source:HGNC Symbol;Acc:HGNC:20893] | 25 | 9.140 | -0.0762 | No |
| 4 | WDR35 | WD repeat domain 35 [Source:HGNC Symbol;Acc:HGNC:29250] | 54 | 1.420 | -0.2952 | Yes |
| 5 | DHCR7 | 7-dehydrocholesterol reductase [Source:HGNC Symbol;Acc:HGNC:2860] | 57 | 1.040 | -0.2667 | Yes |
| 6 | BMP4 | bone morphogenetic protein 4 [Source:HGNC Symbol;Acc:HGNC:1071] | 61 | 0.750 | -0.2476 | Yes |
| 7 | EPG5 | ectopic P-granules autophagy protein 5 homolog [Source:HGNC Symbol;Acc:HGNC:29331] | 72 | 0.100 | -0.2952 | Yes |
| 8 | NEK1 | NIMA related kinase 1 [Source:HGNC Symbol;Acc:HGNC:7744] | 75 | -0.090 | -0.2667 | Yes |
| 9 | FAT4 | FAT atypical cadherin 4 [Source:HGNC Symbol;Acc:HGNC:23109] | 76 | -0.100 | -0.2190 | Yes |
| 10 | DYNC2H1 | dynein cytoplasmic 2 heavy chain 1 [Source:HGNC Symbol;Acc:HGNC:2962] | 79 | -0.230 | -0.1905 | Yes |
| 11 | CEP41 | centrosomal protein 41 [Source:HGNC Symbol;Acc:HGNC:12370] | 84 | -0.340 | -0.1810 | Yes |
| 12 | MKS1 | MKS transition zone complex subunit 1 [Source:HGNC Symbol;Acc:HGNC:7121] | 88 | -0.480 | -0.1619 | Yes |
| 13 | CDKN1C | cyclin dependent kinase inhibitor 1C [Source:HGNC Symbol;Acc:HGNC:1786] | 89 | -0.530 | -0.1143 | Yes |
| 14 | FGFR1 | fibroblast growth factor receptor 1 [Source:HGNC Symbol;Acc:HGNC:3688] | 90 | -0.570 | -0.0667 | Yes |
| 15 | EVC2 | EvC ciliary complex subunit 2 [Source:HGNC Symbol;Acc:HGNC:19747] | 95 | -0.890 | -0.0571 | Yes |
| 16 | TCTN3 | tectonic family member 3 [Source:HGNC Symbol;Acc:HGNC:24519] | 97 | -1.040 | -0.0190 | Yes |
| 17 | GLI3 | GLI family zinc finger 3 [Source:HGNC Symbol;Acc:HGNC:4319] | 104 | -2.920 | -0.0286 | Yes |
| 18 | CHD7 | chromodomain helicase DNA binding protein 7 [Source:HGNC Symbol;Acc:HGNC:20626] | 105 | -3.230 | 0.0190 | Yes |
| 19 | SPECC1L | sperm antigen with calponin homology and coiled-coil domains 1 like [Source:HGNC Symbol;Acc:HGNC:29022] | 109 | -4.400 | 0.0381 | Yes |
| 20 | EVC | EvC ciliary complex subunit 1 [Source:HGNC Symbol;Acc:HGNC:3497] | 119 | -30.300 | -0.0000 | Yes |
| 21 | HHAT | hedgehog acyltransferase [Source:HGNC Symbol;Acc:HGNC:18270] | 124 | -51.600 | 0.0095 | Yes |
Table: GSEA details [plain text format]

  

Fig 2: HP\_ABNORMAL\_CEREBELLAR\_VERMIS\_MORPHOLOGY: Random ES distribution      
 Gene set null distribution of ES for **HP\_ABNORMAL\_CEREBELLAR\_VERMIS\_MORPHOLOGY**

  
