## Supplemental File 3 for "Increased Expression of *ZFPM2* Bypasses *SRY* to Drive 46,XX Testicular Development: A New Mechanism of 46,XX DSD": HP_ABNORMAL_FEMORAL_NECK_HEAD_MORPHOLOGY.html

Details for gene set HP\_ABNORMAL\_FEMORAL\_NECK\_HEAD\_MORPHOLOGY[GSEA]

|  || Dataset | SexDevelopmentGenesPRL\_remapped |
| Phenotype | NoPhenotypeAvailable |
| Upregulated in class | na\_pos |
| GeneSet | HP\_ABNORMAL\_FEMORAL\_NECK\_HEAD\_MORPHOLOGY |
| Enrichment Score (ES) | 0.43557316 |
| Normalized Enrichment Score (NES) | 1.6832856 |
| Nominal p-value | 0.027833002 |
| FDR q-value | 0.91685236 |
| FWER p-Value | 1.0 |
Table: GSEA Results Summary

  

Fig 1: Enrichment plot: HP\_ABNORMAL\_FEMORAL\_NECK\_HEAD\_MORPHOLOGY      
 Profile of the Running ES Score & Positions of GeneSet Members on the Rank Ordered List

  

| SYMBOL | TITLE | RANK IN GENE LIST | RANK METRIC SCORE | RUNNING ES | CORE ENRICHMENT || 1 | CHD4 | chromodomain helicase DNA binding protein 4 [Source:HGNC Symbol;Acc:HGNC:1919] | 0 | 149.000 | 0.0909 | Yes |
| 2 | FLNA | filamin A [Source:HGNC Symbol;Acc:HGNC:3754] | 3 | 53.800 | 0.1644 | Yes |
| 3 | PDE4D | phosphodiesterase 4D [Source:HGNC Symbol;Acc:HGNC:8783] | 9 | 18.600 | 0.2119 | Yes |
| 4 | PCNT | pericentrin [Source:HGNC Symbol;Acc:HGNC:16068] | 17 | 12.400 | 0.2419 | Yes |
| 5 | CREBBP | CREB binding protein [Source:HGNC Symbol;Acc:HGNC:2348] | 20 | 11.800 | 0.3154 | Yes |
| 6 | ATRX | ATRX chromatin remodeler [Source:HGNC Symbol;Acc:HGNC:886] | 27 | 8.350 | 0.3542 | Yes |
| 7 | FGFR3 | fibroblast growth factor receptor 3 [Source:HGNC Symbol;Acc:HGNC:3690] | 37 | 5.830 | 0.3668 | Yes |
| 8 | HESX1 | HESX homeobox 1 [Source:HGNC Symbol;Acc:HGNC:4877] | 51 | 1.570 | 0.3447 | Yes |
| 9 | LMNA | lamin A/C [Source:HGNC Symbol;Acc:HGNC:6636] | 52 | 1.560 | 0.4356 | Yes |
| 10 | GLI3 | GLI family zinc finger 3 [Source:HGNC Symbol;Acc:HGNC:4319] | 104 | -2.920 | 0.0830 | No |
| 11 | WNT5A | Wnt family member 5A [Source:HGNC Symbol;Acc:HGNC:12784] | 115 | -15.700 | 0.0870 | No |
Table: GSEA details [plain text format]

  

Fig 2: HP\_ABNORMAL\_FEMORAL\_NECK\_HEAD\_MORPHOLOGY: Random ES distribution      
 Gene set null distribution of ES for **HP\_ABNORMAL\_FEMORAL\_NECK\_HEAD\_MORPHOLOGY**

  
