## Supplemental File 3 for "Increased Expression of *ZFPM2* Bypasses *SRY* to Drive 46,XX Testicular Development: A New Mechanism of 46,XX DSD": HP_ABNORMAL_FINGERNAIL_MORPHOLOGY.html

Details for gene set HP\_ABNORMAL\_FINGERNAIL\_MORPHOLOGY[GSEA]

|  || Dataset | SexDevelopmentGenesPRL\_remapped |
| Phenotype | NoPhenotypeAvailable |
| Upregulated in class | na\_neg |
| GeneSet | HP\_ABNORMAL\_FINGERNAIL\_MORPHOLOGY |
| Enrichment Score (ES) | -0.45614034 |
| Normalized Enrichment Score (NES) | -1.8178848 |
| Nominal p-value | 0.016260162 |
| FDR q-value | 0.49166995 |
| FWER p-Value | 1.0 |
Table: GSEA Results Summary

  

Fig 1: Enrichment plot: HP\_ABNORMAL\_FINGERNAIL\_MORPHOLOGY      
 Profile of the Running ES Score & Positions of GeneSet Members on the Rank Ordered List

  

| SYMBOL | TITLE | RANK IN GENE LIST | RANK METRIC SCORE | RUNNING ES | CORE ENRICHMENT || 1 | LMNA | lamin A/C [Source:HGNC Symbol;Acc:HGNC:6636] | 52 | 1.560 | -0.3728 | Yes |
| 2 | EFNB1 | ephrin B1 [Source:HGNC Symbol;Acc:HGNC:3226] | 53 | 1.550 | -0.2895 | Yes |
| 3 | WDR35 | WD repeat domain 35 [Source:HGNC Symbol;Acc:HGNC:29250] | 54 | 1.420 | -0.2061 | Yes |
| 4 | FGFR2 | fibroblast growth factor receptor 2 [Source:HGNC Symbol;Acc:HGNC:3689] | 60 | 0.760 | -0.1667 | Yes |
| 5 | PTDSS1 | phosphatidylserine synthase 1 [Source:HGNC Symbol;Acc:HGNC:9587] | 71 | 0.100 | -0.1711 | Yes |
| 6 | PEX1 | peroxisomal biogenesis factor 1 [Source:HGNC Symbol;Acc:HGNC:8850] | 83 | -0.330 | -0.1842 | Yes |
| 7 | CDKN1C | cyclin dependent kinase inhibitor 1C [Source:HGNC Symbol;Acc:HGNC:1786] | 89 | -0.530 | -0.1447 | Yes |
| 8 | ROR2 | receptor tyrosine kinase like orphan receptor 2 [Source:HGNC Symbol;Acc:HGNC:10257] | 91 | -0.580 | -0.0702 | Yes |
| 9 | EVC2 | EvC ciliary complex subunit 2 [Source:HGNC Symbol;Acc:HGNC:19747] | 95 | -0.890 | -0.0132 | Yes |
| 10 | WNT5A | Wnt family member 5A [Source:HGNC Symbol;Acc:HGNC:12784] | 115 | -15.700 | -0.0965 | Yes |
| 11 | TP63 | tumor protein p63 [Source:HGNC Symbol;Acc:HGNC:15979] | 116 | -20.700 | -0.0132 | Yes |
| 12 | EVC | EvC ciliary complex subunit 1 [Source:HGNC Symbol;Acc:HGNC:3497] | 119 | -30.300 | 0.0526 | Yes |
Table: GSEA details [plain text format]

  

Fig 2: HP\_ABNORMAL\_FINGERNAIL\_MORPHOLOGY: Random ES distribution      
 Gene set null distribution of ES for **HP\_ABNORMAL\_FINGERNAIL\_MORPHOLOGY**

  
