## Supplemental File 3 for "Increased Expression of *ZFPM2* Bypasses *SRY* to Drive 46,XX Testicular Development: A New Mechanism of 46,XX DSD": HP_ABNORMAL_GROWTH_HORMONE_LEVEL.html

Details for gene set HP\_ABNORMAL\_GROWTH\_HORMONE\_LEVEL[GSEA]

|  || Dataset | SexDevelopmentGenesPRL\_remapped |
| Phenotype | NoPhenotypeAvailable |
| Upregulated in class | na\_neg |
| GeneSet | HP\_ABNORMAL\_GROWTH\_HORMONE\_LEVEL |
| Enrichment Score (ES) | -0.55 |
| Normalized Enrichment Score (NES) | -2.0326962 |
| Nominal p-value | 0.0020491802 |
| FDR q-value | 0.34384745 |
| FWER p-Value | 0.897 |
Table: GSEA Results Summary

  

Fig 1: Enrichment plot: HP\_ABNORMAL\_GROWTH\_HORMONE\_LEVEL      
 Profile of the Running ES Score & Positions of GeneSet Members on the Rank Ordered List

  

| SYMBOL | TITLE | RANK IN GENE LIST | RANK METRIC SCORE | RUNNING ES | CORE ENRICHMENT || 1 | PDE4D | phosphodiesterase 4D [Source:HGNC Symbol;Acc:HGNC:8783] | 9 | 18.600 | 0.0224 | No |
| 2 | HESX1 | HESX homeobox 1 [Source:HGNC Symbol;Acc:HGNC:4877] | 51 | 1.570 | -0.2310 | No |
| 3 | CDKN1C | cyclin dependent kinase inhibitor 1C [Source:HGNC Symbol;Acc:HGNC:1786] | 89 | -0.530 | -0.4500 | Yes |
| 4 | FGFR1 | fibroblast growth factor receptor 1 [Source:HGNC Symbol;Acc:HGNC:3688] | 90 | -0.570 | -0.3500 | Yes |
| 5 | GLI3 | GLI family zinc finger 3 [Source:HGNC Symbol;Acc:HGNC:4319] | 104 | -2.920 | -0.3621 | Yes |
| 6 | CHD7 | chromodomain helicase DNA binding protein 7 [Source:HGNC Symbol;Acc:HGNC:20626] | 105 | -3.230 | -0.2621 | Yes |
| 7 | PITX2 | paired like homeodomain 2 [Source:HGNC Symbol;Acc:HGNC:9005] | 107 | -3.730 | -0.1707 | Yes |
| 8 | LEPR | leptin receptor [Source:HGNC Symbol;Acc:HGNC:6554] | 111 | -8.460 | -0.0966 | Yes |
| 9 | TP63 | tumor protein p63 [Source:HGNC Symbol;Acc:HGNC:15979] | 116 | -20.700 | -0.0310 | Yes |
| 10 | ESCO2 | establishment of sister chromatid cohesion N-acetyltransferase 2 [Source:HGNC Symbol;Acc:HGNC:27230] | 120 | -32.100 | 0.0431 | Yes |
Table: GSEA details [plain text format]

  

Fig 2: HP\_ABNORMAL\_GROWTH\_HORMONE\_LEVEL: Random ES distribution      
 Gene set null distribution of ES for **HP\_ABNORMAL\_GROWTH\_HORMONE\_LEVEL**

  
