## Supplemental File 3 for "Increased Expression of *ZFPM2* Bypasses *SRY* to Drive 46,XX Testicular Development: A New Mechanism of 46,XX DSD": HP_ABNORMAL_ILIUM_MORPHOLOGY.html

Details for gene set HP\_ABNORMAL\_ILIUM\_MORPHOLOGY[GSEA]

|  || Dataset | SexDevelopmentGenesPRL\_remapped |
| Phenotype | NoPhenotypeAvailable |
| Upregulated in class | na\_pos |
| GeneSet | HP\_ABNORMAL\_ILIUM\_MORPHOLOGY |
| Enrichment Score (ES) | 0.43275863 |
| Normalized Enrichment Score (NES) | 1.6345174 |
| Nominal p-value | 0.04206501 |
| FDR q-value | 0.8564223 |
| FWER p-Value | 1.0 |
Table: GSEA Results Summary

  

Fig 1: Enrichment plot: HP\_ABNORMAL\_ILIUM\_MORPHOLOGY      
 Profile of the Running ES Score & Positions of GeneSet Members on the Rank Ordered List

  

| SYMBOL | TITLE | RANK IN GENE LIST | RANK METRIC SCORE | RUNNING ES | CORE ENRICHMENT || 1 | SOX9 | SRY-box transcription factor 9 [Source:HGNC Symbol;Acc:HGNC:11204] | 1 | 121.000 | 0.0914 | Yes |
| 2 | FLNA | filamin A [Source:HGNC Symbol;Acc:HGNC:3754] | 3 | 53.800 | 0.1828 | Yes |
| 3 | PCNT | pericentrin [Source:HGNC Symbol;Acc:HGNC:16068] | 17 | 12.400 | 0.1707 | Yes |
| 4 | TWIST2 | twist family bHLH transcription factor 2 [Source:HGNC Symbol;Acc:HGNC:20670] | 19 | 11.800 | 0.2621 | Yes |
| 5 | CREBBP | CREB binding protein [Source:HGNC Symbol;Acc:HGNC:2348] | 20 | 11.800 | 0.3621 | Yes |
| 6 | KAT6B | lysine acetyltransferase 6B [Source:HGNC Symbol;Acc:HGNC:17582] | 28 | 8.070 | 0.4017 | Yes |
| 7 | FGFR3 | fibroblast growth factor receptor 3 [Source:HGNC Symbol;Acc:HGNC:3690] | 37 | 5.830 | 0.4328 | Yes |
| 8 | RIPK4 | receptor interacting serine/threonine kinase 4 [Source:HGNC Symbol;Acc:HGNC:496] | 58 | 0.960 | 0.3603 | No |
| 9 | EVC2 | EvC ciliary complex subunit 2 [Source:HGNC Symbol;Acc:HGNC:19747] | 95 | -0.890 | 0.1500 | No |
| 10 | EVC | EvC ciliary complex subunit 1 [Source:HGNC Symbol;Acc:HGNC:3497] | 119 | -30.300 | 0.0517 | No |
Table: GSEA details [plain text format]

  

Fig 2: HP\_ABNORMAL\_ILIUM\_MORPHOLOGY: Random ES distribution      
 Gene set null distribution of ES for **HP\_ABNORMAL\_ILIUM\_MORPHOLOGY**

  
