## Supplemental File 3 for "Increased Expression of *ZFPM2* Bypasses *SRY* to Drive 46,XX Testicular Development: A New Mechanism of 46,XX DSD": HP_ABNORMAL_LOCALIZATION_OF_KIDNEY.html

Details for gene set HP\_ABNORMAL\_LOCALIZATION\_OF\_KIDNEY[GSEA]

|  || Dataset | SexDevelopmentGenesPRL\_remapped |
| Phenotype | NoPhenotypeAvailable |
| Upregulated in class | na\_neg |
| GeneSet | HP\_ABNORMAL\_LOCALIZATION\_OF\_KIDNEY |
| Enrichment Score (ES) | -0.38863635 |
| Normalized Enrichment Score (NES) | -1.7932663 |
| Nominal p-value | 0.015384615 |
| FDR q-value | 0.47574034 |
| FWER p-Value | 1.0 |
Table: GSEA Results Summary

  

Fig 1: Enrichment plot: HP\_ABNORMAL\_LOCALIZATION\_OF\_KIDNEY      
 Profile of the Running ES Score & Positions of GeneSet Members on the Rank Ordered List

  

| SYMBOL | TITLE | RANK IN GENE LIST | RANK METRIC SCORE | RUNNING ES | CORE ENRICHMENT || 1 | ZEB2 | zinc finger E-box binding homeobox 2 [Source:HGNC Symbol;Acc:HGNC:14881] | 5 | 36.500 | 0.0170 | No |
| 2 | PTPN11 | protein tyrosine phosphatase non-receptor type 11 [Source:HGNC Symbol;Acc:HGNC:9644] | 39 | 4.970 | -0.2205 | No |
| 3 | DHCR7 | 7-dehydrocholesterol reductase [Source:HGNC Symbol;Acc:HGNC:2860] | 57 | 1.040 | -0.3125 | No |
| 4 | RIPK4 | receptor interacting serine/threonine kinase 4 [Source:HGNC Symbol;Acc:HGNC:496] | 58 | 0.960 | -0.2500 | No |
| 5 | FGFR2 | fibroblast growth factor receptor 2 [Source:HGNC Symbol;Acc:HGNC:3689] | 60 | 0.760 | -0.1966 | No |
| 6 | FAT4 | FAT atypical cadherin 4 [Source:HGNC Symbol;Acc:HGNC:23109] | 76 | -0.100 | -0.2705 | No |
| 7 | FGFR1 | fibroblast growth factor receptor 1 [Source:HGNC Symbol;Acc:HGNC:3688] | 90 | -0.570 | -0.3261 | Yes |
| 8 | TCTN3 | tectonic family member 3 [Source:HGNC Symbol;Acc:HGNC:24519] | 97 | -1.040 | -0.3182 | Yes |
| 9 | GLI3 | GLI family zinc finger 3 [Source:HGNC Symbol;Acc:HGNC:4319] | 104 | -2.920 | -0.3102 | Yes |
| 10 | CHD7 | chromodomain helicase DNA binding protein 7 [Source:HGNC Symbol;Acc:HGNC:20626] | 105 | -3.230 | -0.2477 | Yes |
| 11 | SALL1 | spalt like transcription factor 1 [Source:HGNC Symbol;Acc:HGNC:10524] | 106 | -3.630 | -0.1852 | Yes |
| 12 | POR | cytochrome p450 oxidoreductase [Source:HGNC Symbol;Acc:HGNC:9208] | 108 | -3.990 | -0.1318 | Yes |
| 13 | SPECC1L | sperm antigen with calponin homology and coiled-coil domains 1 like [Source:HGNC Symbol;Acc:HGNC:29022] | 109 | -4.400 | -0.0693 | Yes |
| 14 | RBBP8 | "RB binding protein 8, endonuclease [Source:HGNC Symbol;Acc:HGNC:9891]" | 110 | -8.050 | -0.0068 | Yes |
| 15 | PBX1 | PBX homeobox 1 [Source:HGNC Symbol;Acc:HGNC:8632] | 112 | -12.600 | 0.0466 | Yes |
| 16 | ESCO2 | establishment of sister chromatid cohesion N-acetyltransferase 2 [Source:HGNC Symbol;Acc:HGNC:27230] | 120 | -32.100 | 0.0455 | Yes |
Table: GSEA details [plain text format]

  

Fig 2: HP\_ABNORMAL\_LOCALIZATION\_OF\_KIDNEY: Random ES distribution      
 Gene set null distribution of ES for **HP\_ABNORMAL\_LOCALIZATION\_OF\_KIDNEY**

  
