## Supplemental File 3 for "Increased Expression of *ZFPM2* Bypasses *SRY* to Drive 46,XX Testicular Development: A New Mechanism of 46,XX DSD": HP_ABNORMAL_RETINAL_MORPHOLOGY.html

Details for gene set HP\_ABNORMAL\_RETINAL\_MORPHOLOGY[GSEA]

|  || Dataset | SexDevelopmentGenesPRL\_remapped |
| Phenotype | NoPhenotypeAvailable |
| Upregulated in class | na\_neg |
| GeneSet | HP\_ABNORMAL\_RETINAL\_MORPHOLOGY |
| Enrichment Score (ES) | -0.3088954 |
| Normalized Enrichment Score (NES) | -1.8443365 |
| Nominal p-value | 0.016064256 |
| FDR q-value | 0.5147987 |
| FWER p-Value | 1.0 |
Table: GSEA Results Summary

  

Fig 1: Enrichment plot: HP\_ABNORMAL\_RETINAL\_MORPHOLOGY      
 Profile of the Running ES Score & Positions of GeneSet Members on the Rank Ordered List

  

| SYMBOL | TITLE | RANK IN GENE LIST | RANK METRIC SCORE | RUNNING ES | CORE ENRICHMENT || 1 | ZEB2 | zinc finger E-box binding homeobox 2 [Source:HGNC Symbol;Acc:HGNC:14881] | 5 | 36.500 | -0.0235 | No |
| 2 | CREBBP | CREB binding protein [Source:HGNC Symbol;Acc:HGNC:2348] | 20 | 11.800 | -0.1437 | No |
| 3 | BCOR | BCL6 corepressor [Source:HGNC Symbol;Acc:HGNC:20893] | 25 | 9.140 | -0.1564 | No |
| 4 | MKKS | MKKS centrosomal shuttling protein [Source:HGNC Symbol;Acc:HGNC:7108] | 34 | 6.190 | -0.2121 | No |
| 5 | TRIM32 | tripartite motif containing 32 [Source:HGNC Symbol;Acc:HGNC:16380] | 44 | 2.410 | -0.2786 | Yes |
| 6 | DNMT3B | DNA methyltransferase 3 beta [Source:HGNC Symbol;Acc:HGNC:2979] | 45 | 2.110 | -0.2483 | Yes |
| 7 | BBS9 | Bardet-Biedl syndrome 9 [Source:HGNC Symbol;Acc:HGNC:30000] | 47 | 2.000 | -0.2287 | Yes |
| 8 | BBS2 | Bardet-Biedl syndrome 2 [Source:HGNC Symbol;Acc:HGNC:967] | 48 | 1.960 | -0.1984 | Yes |
| 9 | BBS12 | Bardet-Biedl syndrome 12 [Source:HGNC Symbol;Acc:HGNC:26648] | 49 | 1.960 | -0.1681 | Yes |
| 10 | LMNA | lamin A/C [Source:HGNC Symbol;Acc:HGNC:6636] | 52 | 1.560 | -0.1593 | Yes |
| 11 | FGFR2 | fibroblast growth factor receptor 2 [Source:HGNC Symbol;Acc:HGNC:3689] | 60 | 0.760 | -0.2043 | Yes |
| 12 | BMP4 | bone morphogenetic protein 4 [Source:HGNC Symbol;Acc:HGNC:1071] | 61 | 0.750 | -0.1740 | Yes |
| 13 | BBS7 | Bardet-Biedl syndrome 7 [Source:HGNC Symbol;Acc:HGNC:18758] | 66 | 0.400 | -0.1867 | Yes |
| 14 | ARL6 | ADP ribosylation factor like GTPase 6 [Source:HGNC Symbol;Acc:HGNC:13210] | 68 | 0.280 | -0.1672 | Yes |
| 15 | EPG5 | ectopic P-granules autophagy protein 5 homolog [Source:HGNC Symbol;Acc:HGNC:29331] | 72 | 0.100 | -0.1691 | Yes |
| 16 | NEK1 | NIMA related kinase 1 [Source:HGNC Symbol;Acc:HGNC:7744] | 75 | -0.090 | -0.1603 | Yes |
| 17 | DYNC2H1 | dynein cytoplasmic 2 heavy chain 1 [Source:HGNC Symbol;Acc:HGNC:2962] | 79 | -0.230 | -0.1623 | Yes |
| 18 | PEX1 | peroxisomal biogenesis factor 1 [Source:HGNC Symbol;Acc:HGNC:8850] | 83 | -0.330 | -0.1642 | Yes |
| 19 | CEP41 | centrosomal protein 41 [Source:HGNC Symbol;Acc:HGNC:12370] | 84 | -0.340 | -0.1339 | Yes |
| 20 | HCCS | holocytochrome c synthase [Source:HGNC Symbol;Acc:HGNC:4837] | 86 | -0.400 | -0.1144 | Yes |
| 21 | MKS1 | MKS transition zone complex subunit 1 [Source:HGNC Symbol;Acc:HGNC:7121] | 88 | -0.480 | -0.0948 | Yes |
| 22 | FGFR1 | fibroblast growth factor receptor 1 [Source:HGNC Symbol;Acc:HGNC:3688] | 90 | -0.570 | -0.0753 | Yes |
| 23 | TTC8 | tetratricopeptide repeat domain 8 [Source:HGNC Symbol;Acc:HGNC:20087] | 92 | -0.640 | -0.0557 | Yes |
| 24 | TCTN3 | tectonic family member 3 [Source:HGNC Symbol;Acc:HGNC:24519] | 97 | -1.040 | -0.0684 | Yes |
| 25 | WFS1 | wolframin ER transmembrane glycoprotein [Source:HGNC Symbol;Acc:HGNC:12762] | 99 | -1.320 | -0.0489 | Yes |
| 26 | BBS4 | Bardet-Biedl syndrome 4 [Source:HGNC Symbol;Acc:HGNC:969] | 100 | -1.360 | -0.0186 | Yes |
| 27 | BBS10 | Bardet-Biedl syndrome 10 [Source:HGNC Symbol;Acc:HGNC:26291] | 102 | -1.550 | 0.0010 | Yes |
| 28 | CHD7 | chromodomain helicase DNA binding protein 7 [Source:HGNC Symbol;Acc:HGNC:20626] | 105 | -3.230 | 0.0098 | Yes |
| 29 | SALL1 | spalt like transcription factor 1 [Source:HGNC Symbol;Acc:HGNC:10524] | 106 | -3.630 | 0.0401 | Yes |
| 30 | WWOX | WW domain containing oxidoreductase [Source:HGNC Symbol;Acc:HGNC:12799] | 113 | -14.500 | 0.0059 | Yes |
| 31 | BBS5 | Bardet-Biedl syndrome 5 [Source:HGNC Symbol;Acc:HGNC:970] | 114 | -14.600 | 0.0362 | Yes |
| 32 | B3GLCT | beta 3-glucosyltransferase [Source:HGNC Symbol;Acc:HGNC:20207] | 118 | -22.200 | 0.0342 | Yes |
| 33 | HHAT | hedgehog acyltransferase [Source:HGNC Symbol;Acc:HGNC:18270] | 124 | -51.600 | 0.0108 | Yes |
Table: GSEA details [plain text format]

  

Fig 2: HP\_ABNORMAL\_RETINAL\_MORPHOLOGY: Random ES distribution      
 Gene set null distribution of ES for **HP\_ABNORMAL\_RETINAL\_MORPHOLOGY**

  
