## Supplemental File 3 for "Increased Expression of *ZFPM2* Bypasses *SRY* to Drive 46,XX Testicular Development: A New Mechanism of 46,XX DSD": HP_ABNORMALITY_OF_GLOBE_SIZE.html

Details for gene set HP\_ABNORMALITY\_OF\_GLOBE\_SIZE[GSEA]

|  || Dataset | SexDevelopmentGenesPRL\_remapped |
| Phenotype | NoPhenotypeAvailable |
| Upregulated in class | na\_neg |
| GeneSet | HP\_ABNORMALITY\_OF\_GLOBE\_SIZE |
| Enrichment Score (ES) | -0.4181013 |
| Normalized Enrichment Score (NES) | -2.0666394 |
| Nominal p-value | 0.0040241447 |
| FDR q-value | 0.48134091 |
| FWER p-Value | 0.837 |
Table: GSEA Results Summary

  

Fig 1: Enrichment plot: HP\_ABNORMALITY\_OF\_GLOBE\_SIZE      
 Profile of the Running ES Score & Positions of GeneSet Members on the Rank Ordered List

  

| SYMBOL | TITLE | RANK IN GENE LIST | RANK METRIC SCORE | RUNNING ES | CORE ENRICHMENT || 1 | ZEB2 | zinc finger E-box binding homeobox 2 [Source:HGNC Symbol;Acc:HGNC:14881] | 5 | 36.500 | 0.0059 | No |
| 2 | BCOR | BCL6 corepressor [Source:HGNC Symbol;Acc:HGNC:20893] | 25 | 9.140 | -0.1190 | No |
| 3 | RIPK4 | receptor interacting serine/threonine kinase 4 [Source:HGNC Symbol;Acc:HGNC:496] | 58 | 0.960 | -0.3655 | Yes |
| 4 | FGFR2 | fibroblast growth factor receptor 2 [Source:HGNC Symbol;Acc:HGNC:3689] | 60 | 0.760 | -0.3222 | Yes |
| 5 | BMP4 | bone morphogenetic protein 4 [Source:HGNC Symbol;Acc:HGNC:1071] | 61 | 0.750 | -0.2696 | Yes |
| 6 | FIG4 | FIG4 phosphoinositide 5-phosphatase [Source:HGNC Symbol;Acc:HGNC:16873] | 65 | 0.420 | -0.2450 | Yes |
| 7 | FRAS1 | Fraser extracellular matrix complex subunit 1 [Source:HGNC Symbol;Acc:HGNC:19185] | 73 | 0.010 | -0.2577 | Yes |
| 8 | HCCS | holocytochrome c synthase [Source:HGNC Symbol;Acc:HGNC:4837] | 86 | -0.400 | -0.3173 | Yes |
| 9 | MKS1 | MKS transition zone complex subunit 1 [Source:HGNC Symbol;Acc:HGNC:7121] | 88 | -0.480 | -0.2740 | Yes |
| 10 | FGFR1 | fibroblast growth factor receptor 1 [Source:HGNC Symbol;Acc:HGNC:3688] | 90 | -0.570 | -0.2307 | Yes |
| 11 | GRIP1 | glutamate receptor interacting protein 1 [Source:HGNC Symbol;Acc:HGNC:18708] | 94 | -0.750 | -0.2061 | Yes |
| 12 | TCTN3 | tectonic family member 3 [Source:HGNC Symbol;Acc:HGNC:24519] | 97 | -1.040 | -0.1722 | Yes |
| 13 | GLI3 | GLI family zinc finger 3 [Source:HGNC Symbol;Acc:HGNC:4319] | 104 | -2.920 | -0.1756 | Yes |
| 14 | CHD7 | chromodomain helicase DNA binding protein 7 [Source:HGNC Symbol;Acc:HGNC:20626] | 105 | -3.230 | -0.1230 | Yes |
| 15 | SALL1 | spalt like transcription factor 1 [Source:HGNC Symbol;Acc:HGNC:10524] | 106 | -3.630 | -0.0703 | Yes |
| 16 | RBBP8 | "RB binding protein 8, endonuclease [Source:HGNC Symbol;Acc:HGNC:9891]" | 110 | -8.050 | -0.0457 | Yes |
| 17 | FREM2 | FRAS1 related extracellular matrix 2 [Source:HGNC Symbol;Acc:HGNC:25396] | 117 | -21.900 | -0.0492 | Yes |
| 18 | ESCO2 | establishment of sister chromatid cohesion N-acetyltransferase 2 [Source:HGNC Symbol;Acc:HGNC:27230] | 120 | -32.100 | -0.0152 | Yes |
| 19 | FOXL2 | forkhead box L2 [Source:HGNC Symbol;Acc:HGNC:1092] | 122 | -40.400 | 0.0280 | Yes |
Table: GSEA details [plain text format]

  

Fig 2: HP\_ABNORMALITY\_OF\_GLOBE\_SIZE: Random ES distribution      
 Gene set null distribution of ES for **HP\_ABNORMALITY\_OF\_GLOBE\_SIZE**

  
