## Supplemental File 3 for "Increased Expression of *ZFPM2* Bypasses *SRY* to Drive 46,XX Testicular Development: A New Mechanism of 46,XX DSD": HP_ABNORMALITY_OF_HAIR_TEXTURE.html

Details for gene set HP\_ABNORMALITY\_OF\_HAIR\_TEXTURE[GSEA]

|  || Dataset | SexDevelopmentGenesPRL\_remapped |
| Phenotype | NoPhenotypeAvailable |
| Upregulated in class | na\_pos |
| GeneSet | HP\_ABNORMALITY\_OF\_HAIR\_TEXTURE |
| Enrichment Score (ES) | 0.483004 |
| Normalized Enrichment Score (NES) | 1.8550069 |
| Nominal p-value | 0.018329939 |
| FDR q-value | 1.0 |
| FWER p-Value | 0.999 |
Table: GSEA Results Summary

  

Fig 1: Enrichment plot: HP\_ABNORMALITY\_OF\_HAIR\_TEXTURE      
 Profile of the Running ES Score & Positions of GeneSet Members on the Rank Ordered List

  

| SYMBOL | TITLE | RANK IN GENE LIST | RANK METRIC SCORE | RUNNING ES | CORE ENRICHMENT || 1 | FLNA | filamin A [Source:HGNC Symbol;Acc:HGNC:3754] | 3 | 53.800 | 0.0648 | Yes |
| 2 | SOS1 | SOS Ras/Rac guanine nucleotide exchange factor 1 [Source:HGNC Symbol;Acc:HGNC:11187] | 15 | 12.800 | 0.0601 | Yes |
| 3 | PCNT | pericentrin [Source:HGNC Symbol;Acc:HGNC:16068] | 17 | 12.400 | 0.1423 | Yes |
| 4 | TWIST2 | twist family bHLH transcription factor 2 [Source:HGNC Symbol;Acc:HGNC:20670] | 19 | 11.800 | 0.2245 | Yes |
| 5 | KAT6B | lysine acetyltransferase 6B [Source:HGNC Symbol;Acc:HGNC:17582] | 28 | 8.070 | 0.2458 | Yes |
| 6 | MED12 | mediator complex subunit 12 [Source:HGNC Symbol;Acc:HGNC:11957] | 33 | 6.470 | 0.3020 | Yes |
| 7 | PTPN11 | protein tyrosine phosphatase non-receptor type 11 [Source:HGNC Symbol;Acc:HGNC:9644] | 39 | 4.970 | 0.3494 | Yes |
| 8 | EFNB1 | ephrin B1 [Source:HGNC Symbol;Acc:HGNC:3226] | 53 | 1.550 | 0.3273 | Yes |
| 9 | WDR35 | WD repeat domain 35 [Source:HGNC Symbol;Acc:HGNC:29250] | 54 | 1.420 | 0.4182 | Yes |
| 10 | RIPK4 | receptor interacting serine/threonine kinase 4 [Source:HGNC Symbol;Acc:HGNC:496] | 58 | 0.960 | 0.4830 | Yes |
| 11 | TP63 | tumor protein p63 [Source:HGNC Symbol;Acc:HGNC:15979] | 116 | -20.700 | 0.0783 | No |
Table: GSEA details [plain text format]

  

Fig 2: HP\_ABNORMALITY\_OF\_HAIR\_TEXTURE: Random ES distribution      
 Gene set null distribution of ES for **HP\_ABNORMALITY\_OF\_HAIR\_TEXTURE**

  
