## Supplemental File 3 for "Increased Expression of *ZFPM2* Bypasses *SRY* to Drive 46,XX Testicular Development: A New Mechanism of 46,XX DSD": HP_APLASIA_HYPOPLASIA_AFFECTING_THE_EYE.html

Details for gene set HP\_APLASIA\_HYPOPLASIA\_AFFECTING\_THE\_EYE[GSEA]

|  || Dataset | SexDevelopmentGenesPRL\_remapped |
| Phenotype | NoPhenotypeAvailable |
| Upregulated in class | na\_neg |
| GeneSet | HP\_APLASIA\_HYPOPLASIA\_AFFECTING\_THE\_EYE |
| Enrichment Score (ES) | -0.35247526 |
| Normalized Enrichment Score (NES) | -1.9583901 |
| Nominal p-value | 0.0056390977 |
| FDR q-value | 0.420061 |
| FWER p-Value | 0.971 |
Table: GSEA Results Summary

  

Fig 1: Enrichment plot: HP\_APLASIA\_HYPOPLASIA\_AFFECTING\_THE\_EYE      
 Profile of the Running ES Score & Positions of GeneSet Members on the Rank Ordered List

  

| SYMBOL | TITLE | RANK IN GENE LIST | RANK METRIC SCORE | RUNNING ES | CORE ENRICHMENT || 1 | ZEB2 | zinc finger E-box binding homeobox 2 [Source:HGNC Symbol;Acc:HGNC:14881] | 5 | 36.500 | -0.0095 | No |
| 2 | WDR11 | WD repeat domain 11 [Source:HGNC Symbol;Acc:HGNC:13831] | 12 | 15.800 | -0.0289 | No |
| 3 | BCOR | BCL6 corepressor [Source:HGNC Symbol;Acc:HGNC:20893] | 25 | 9.140 | -0.1077 | No |
| 4 | MED12 | mediator complex subunit 12 [Source:HGNC Symbol;Acc:HGNC:11957] | 33 | 6.470 | -0.1370 | No |
| 5 | HESX1 | HESX homeobox 1 [Source:HGNC Symbol;Acc:HGNC:4877] | 51 | 1.570 | -0.2653 | No |
| 6 | DHCR7 | 7-dehydrocholesterol reductase [Source:HGNC Symbol;Acc:HGNC:2860] | 57 | 1.040 | -0.2749 | No |
| 7 | RIPK4 | receptor interacting serine/threonine kinase 4 [Source:HGNC Symbol;Acc:HGNC:496] | 58 | 0.960 | -0.2349 | No |
| 8 | BMP4 | bone morphogenetic protein 4 [Source:HGNC Symbol;Acc:HGNC:1071] | 61 | 0.750 | -0.2147 | No |
| 9 | FIG4 | FIG4 phosphoinositide 5-phosphatase [Source:HGNC Symbol;Acc:HGNC:16873] | 65 | 0.420 | -0.2044 | No |
| 10 | FRAS1 | Fraser extracellular matrix complex subunit 1 [Source:HGNC Symbol;Acc:HGNC:19185] | 73 | 0.010 | -0.2337 | No |
| 11 | HCCS | holocytochrome c synthase [Source:HGNC Symbol;Acc:HGNC:4837] | 86 | -0.400 | -0.3125 | Yes |
| 12 | MKS1 | MKS transition zone complex subunit 1 [Source:HGNC Symbol;Acc:HGNC:7121] | 88 | -0.480 | -0.2824 | Yes |
| 13 | FGFR1 | fibroblast growth factor receptor 1 [Source:HGNC Symbol;Acc:HGNC:3688] | 90 | -0.570 | -0.2523 | Yes |
| 14 | GRIP1 | glutamate receptor interacting protein 1 [Source:HGNC Symbol;Acc:HGNC:18708] | 94 | -0.750 | -0.2420 | Yes |
| 15 | TCTN3 | tectonic family member 3 [Source:HGNC Symbol;Acc:HGNC:24519] | 97 | -1.040 | -0.2218 | Yes |
| 16 | GLI3 | GLI family zinc finger 3 [Source:HGNC Symbol;Acc:HGNC:4319] | 104 | -2.920 | -0.2412 | Yes |
| 17 | CHD7 | chromodomain helicase DNA binding protein 7 [Source:HGNC Symbol;Acc:HGNC:20626] | 105 | -3.230 | -0.2012 | Yes |
| 18 | SALL1 | spalt like transcription factor 1 [Source:HGNC Symbol;Acc:HGNC:10524] | 106 | -3.630 | -0.1612 | Yes |
| 19 | PITX2 | paired like homeodomain 2 [Source:HGNC Symbol;Acc:HGNC:9005] | 107 | -3.730 | -0.1212 | Yes |
| 20 | RBBP8 | "RB binding protein 8, endonuclease [Source:HGNC Symbol;Acc:HGNC:9891]" | 110 | -8.050 | -0.1010 | Yes |
| 21 | TP63 | tumor protein p63 [Source:HGNC Symbol;Acc:HGNC:15979] | 116 | -20.700 | -0.1105 | Yes |
| 22 | FREM2 | FRAS1 related extracellular matrix 2 [Source:HGNC Symbol;Acc:HGNC:25396] | 117 | -21.900 | -0.0705 | Yes |
| 23 | ESCO2 | establishment of sister chromatid cohesion N-acetyltransferase 2 [Source:HGNC Symbol;Acc:HGNC:27230] | 120 | -32.100 | -0.0503 | Yes |
| 24 | FOXL2 | forkhead box L2 [Source:HGNC Symbol;Acc:HGNC:1092] | 122 | -40.400 | -0.0202 | Yes |
| 25 | HHAT | hedgehog acyltransferase [Source:HGNC Symbol;Acc:HGNC:18270] | 124 | -51.600 | 0.0099 | Yes |
Table: GSEA details [plain text format]

  

Fig 2: HP\_APLASIA\_HYPOPLASIA\_AFFECTING\_THE\_EYE: Random ES distribution      
 Gene set null distribution of ES for **HP\_APLASIA\_HYPOPLASIA\_AFFECTING\_THE\_EYE**

  
