## Supplemental File 3 for "Increased Expression of *ZFPM2* Bypasses *SRY* to Drive 46,XX Testicular Development: A New Mechanism of 46,XX DSD": HP_APLASIA_HYPOPLASIA_INVOLVING_BONES_OF_THE_UPPER_LIMBS.html

Details for gene set HP\_APLASIA\_HYPOPLASIA\_INVOLVING\_BONES\_OF\_THE\_UPPER\_LIMBS[GSEA]

|  || Dataset | SexDevelopmentGenesPRL\_remapped |
| Phenotype | NoPhenotypeAvailable |
| Upregulated in class | na\_neg |
| GeneSet | HP\_APLASIA\_HYPOPLASIA\_INVOLVING\_BONES\_OF\_THE\_UPPER\_LIMBS |
| Enrichment Score (ES) | -0.27460548 |
| Normalized Enrichment Score (NES) | -1.7180134 |
| Nominal p-value | 0.03258656 |
| FDR q-value | 0.54115415 |
| FWER p-Value | 1.0 |
Table: GSEA Results Summary

  

Fig 1: Enrichment plot: HP\_APLASIA\_HYPOPLASIA\_INVOLVING\_BONES\_OF\_THE\_UPPER\_LIMBS      
 Profile of the Running ES Score & Positions of GeneSet Members on the Rank Ordered List

  

| SYMBOL | TITLE | RANK IN GENE LIST | RANK METRIC SCORE | RUNNING ES | CORE ENRICHMENT || 1 | FLNA | filamin A [Source:HGNC Symbol;Acc:HGNC:3754] | 3 | 53.800 | -0.0109 | No |
| 2 | PDE4D | phosphodiesterase 4D [Source:HGNC Symbol;Acc:HGNC:8783] | 9 | 18.600 | -0.0453 | No |
| 3 | SETBP1 | SET binding protein 1 [Source:HGNC Symbol;Acc:HGNC:15573] | 14 | 14.100 | -0.0680 | No |
| 4 | PCNT | pericentrin [Source:HGNC Symbol;Acc:HGNC:16068] | 17 | 12.400 | -0.0671 | No |
| 5 | TWIST2 | twist family bHLH transcription factor 2 [Source:HGNC Symbol;Acc:HGNC:20670] | 19 | 11.800 | -0.0545 | No |
| 6 | BCOR | BCL6 corepressor [Source:HGNC Symbol;Acc:HGNC:20893] | 25 | 9.140 | -0.0890 | No |
| 7 | KAT6B | lysine acetyltransferase 6B [Source:HGNC Symbol;Acc:HGNC:17582] | 28 | 8.070 | -0.0881 | No |
| 8 | MED12 | mediator complex subunit 12 [Source:HGNC Symbol;Acc:HGNC:11957] | 33 | 6.470 | -0.1108 | No |
| 9 | FGFR3 | fibroblast growth factor receptor 3 [Source:HGNC Symbol;Acc:HGNC:3690] | 37 | 5.830 | -0.1217 | No |
| 10 | HESX1 | HESX homeobox 1 [Source:HGNC Symbol;Acc:HGNC:4877] | 51 | 1.570 | -0.2502 | Yes |
| 11 | LMNA | lamin A/C [Source:HGNC Symbol;Acc:HGNC:6636] | 52 | 1.560 | -0.2258 | Yes |
| 12 | WDR35 | WD repeat domain 35 [Source:HGNC Symbol;Acc:HGNC:29250] | 54 | 1.420 | -0.2132 | Yes |
| 13 | DHCR7 | 7-dehydrocholesterol reductase [Source:HGNC Symbol;Acc:HGNC:2860] | 57 | 1.040 | -0.2123 | Yes |
| 14 | RIPK4 | receptor interacting serine/threonine kinase 4 [Source:HGNC Symbol;Acc:HGNC:496] | 58 | 0.960 | -0.1879 | Yes |
| 15 | FGF10 | fibroblast growth factor 10 [Source:HGNC Symbol;Acc:HGNC:3666] | 59 | 0.890 | -0.1636 | Yes |
| 16 | FGFR2 | fibroblast growth factor receptor 2 [Source:HGNC Symbol;Acc:HGNC:3689] | 60 | 0.760 | -0.1392 | Yes |
| 17 | BMP4 | bone morphogenetic protein 4 [Source:HGNC Symbol;Acc:HGNC:1071] | 61 | 0.750 | -0.1148 | Yes |
| 18 | FIG4 | FIG4 phosphoinositide 5-phosphatase [Source:HGNC Symbol;Acc:HGNC:16873] | 65 | 0.420 | -0.1257 | Yes |
| 19 | PTDSS1 | phosphatidylserine synthase 1 [Source:HGNC Symbol;Acc:HGNC:9587] | 71 | 0.100 | -0.1601 | Yes |
| 20 | FRAS1 | Fraser extracellular matrix complex subunit 1 [Source:HGNC Symbol;Acc:HGNC:19185] | 73 | 0.010 | -0.1475 | Yes |
| 21 | NEK1 | NIMA related kinase 1 [Source:HGNC Symbol;Acc:HGNC:7744] | 75 | -0.090 | -0.1349 | Yes |
| 22 | FAT4 | FAT atypical cadherin 4 [Source:HGNC Symbol;Acc:HGNC:23109] | 76 | -0.100 | -0.1105 | Yes |
| 23 | DYNC2H1 | dynein cytoplasmic 2 heavy chain 1 [Source:HGNC Symbol;Acc:HGNC:2962] | 79 | -0.230 | -0.1096 | Yes |
| 24 | CDKN1C | cyclin dependent kinase inhibitor 1C [Source:HGNC Symbol;Acc:HGNC:1786] | 89 | -0.530 | -0.1911 | Yes |
| 25 | FGFR1 | fibroblast growth factor receptor 1 [Source:HGNC Symbol;Acc:HGNC:3688] | 90 | -0.570 | -0.1667 | Yes |
| 26 | ROR2 | receptor tyrosine kinase like orphan receptor 2 [Source:HGNC Symbol;Acc:HGNC:10257] | 91 | -0.580 | -0.1423 | Yes |
| 27 | EVC2 | EvC ciliary complex subunit 2 [Source:HGNC Symbol;Acc:HGNC:19747] | 95 | -0.890 | -0.1532 | Yes |
| 28 | CUL7 | cullin 7 [Source:HGNC Symbol;Acc:HGNC:21024] | 96 | -0.950 | -0.1288 | Yes |
| 29 | TCTN3 | tectonic family member 3 [Source:HGNC Symbol;Acc:HGNC:24519] | 97 | -1.040 | -0.1044 | Yes |
| 30 | SCARF2 | scavenger receptor class F member 2 [Source:HGNC Symbol;Acc:HGNC:19869] | 103 | -2.410 | -0.1389 | Yes |
| 31 | GLI3 | GLI family zinc finger 3 [Source:HGNC Symbol;Acc:HGNC:4319] | 104 | -2.920 | -0.1145 | Yes |
| 32 | CHD7 | chromodomain helicase DNA binding protein 7 [Source:HGNC Symbol;Acc:HGNC:20626] | 105 | -3.230 | -0.0901 | Yes |
| 33 | POR | cytochrome p450 oxidoreductase [Source:HGNC Symbol;Acc:HGNC:9208] | 108 | -3.990 | -0.0892 | Yes |
| 34 | SPECC1L | sperm antigen with calponin homology and coiled-coil domains 1 like [Source:HGNC Symbol;Acc:HGNC:29022] | 109 | -4.400 | -0.0648 | Yes |
| 35 | RBBP8 | "RB binding protein 8, endonuclease [Source:HGNC Symbol;Acc:HGNC:9891]" | 110 | -8.050 | -0.0405 | Yes |
| 36 | WNT5A | Wnt family member 5A [Source:HGNC Symbol;Acc:HGNC:12784] | 115 | -15.700 | -0.0631 | Yes |
| 37 | TP63 | tumor protein p63 [Source:HGNC Symbol;Acc:HGNC:15979] | 116 | -20.700 | -0.0387 | Yes |
| 38 | B3GLCT | beta 3-glucosyltransferase [Source:HGNC Symbol;Acc:HGNC:20207] | 118 | -22.200 | -0.0261 | Yes |
| 39 | EVC | EvC ciliary complex subunit 1 [Source:HGNC Symbol;Acc:HGNC:3497] | 119 | -30.300 | -0.0017 | Yes |
| 40 | ESCO2 | establishment of sister chromatid cohesion N-acetyltransferase 2 [Source:HGNC Symbol;Acc:HGNC:27230] | 120 | -32.100 | 0.0227 | Yes |
| 41 | HHAT | hedgehog acyltransferase [Source:HGNC Symbol;Acc:HGNC:18270] | 124 | -51.600 | 0.0118 | Yes |
Table: GSEA details [plain text format]

  

Fig 2: HP\_APLASIA\_HYPOPLASIA\_INVOLVING\_BONES\_OF\_THE\_UPPER\_LIMBS: Random ES distribution      
 Gene set null distribution of ES for **HP\_APLASIA\_HYPOPLASIA\_INVOLVING\_BONES\_OF\_THE\_UPPER\_LIMBS**

  
