## Supplemental File 3 for "Increased Expression of *ZFPM2* Bypasses *SRY* to Drive 46,XX Testicular Development: A New Mechanism of 46,XX DSD": HP_APLASIA_HYPOPLASIA_OF_THE_CEREBELLAR_VERMIS.html

Details for gene set HP\_APLASIA\_HYPOPLASIA\_OF\_THE\_CEREBELLAR\_VERMIS[GSEA]

|  || Dataset | SexDevelopmentGenesPRL\_remapped |
| Phenotype | NoPhenotypeAvailable |
| Upregulated in class | na\_neg |
| GeneSet | HP\_APLASIA\_HYPOPLASIA\_OF\_THE\_CEREBELLAR\_VERMIS |
| Enrichment Score (ES) | -0.34285715 |
| Normalized Enrichment Score (NES) | -1.7636498 |
| Nominal p-value | 0.022900764 |
| FDR q-value | 0.5264081 |
| FWER p-Value | 1.0 |
Table: GSEA Results Summary

  

Fig 2: HP\_APLASIA\_HYPOPLASIA\_OF\_THE\_CEREBELLAR\_VERMIS: Random ES distribution      
 Gene set null distribution of ES for **HP\_APLASIA\_HYPOPLASIA\_OF\_THE\_CEREBELLAR\_VERMIS**

  
