## Supplemental File 3 for "Increased Expression of *ZFPM2* Bypasses *SRY* to Drive 46,XX Testicular Development: A New Mechanism of 46,XX DSD": HP_CLEFT_LIP.html

Details for gene set HP\_CLEFT\_LIP[GSEA]

|  || Dataset | SexDevelopmentGenesPRL\_remapped |
| Phenotype | NoPhenotypeAvailable |
| Upregulated in class | na\_neg |
| GeneSet | HP\_CLEFT\_LIP |
| Enrichment Score (ES) | -0.39139706 |
| Normalized Enrichment Score (NES) | -2.206885 |
| Nominal p-value | 0.0019417476 |
| FDR q-value | 0.3919955 |
| FWER p-Value | 0.525 |
Table: GSEA Results Summary

  

Fig 1: Enrichment plot: HP\_CLEFT\_LIP      
 Profile of the Running ES Score & Positions of GeneSet Members on the Rank Ordered List

  

| SYMBOL | TITLE | RANK IN GENE LIST | RANK METRIC SCORE | RUNNING ES | CORE ENRICHMENT || 1 | BCOR | BCL6 corepressor [Source:HGNC Symbol;Acc:HGNC:20893] | 25 | 9.140 | -0.2232 | No |
| 2 | MID1 | midline 1 [Source:HGNC Symbol;Acc:HGNC:7095] | 26 | 8.770 | -0.1888 | No |
| 3 | MED12 | mediator complex subunit 12 [Source:HGNC Symbol;Acc:HGNC:11957] | 33 | 6.470 | -0.2161 | No |
| 4 | HESX1 | HESX homeobox 1 [Source:HGNC Symbol;Acc:HGNC:4877] | 51 | 1.570 | -0.3569 | Yes |
| 5 | EFNB1 | ephrin B1 [Source:HGNC Symbol;Acc:HGNC:3226] | 53 | 1.550 | -0.3327 | Yes |
| 6 | WDR35 | WD repeat domain 35 [Source:HGNC Symbol;Acc:HGNC:29250] | 54 | 1.420 | -0.2983 | Yes |
| 7 | NSMF | NMDA receptor synaptonuclear signaling and neuronal migration factor [Source:HGNC Symbol;Acc:HGNC:29843] | 55 | 1.360 | -0.2638 | Yes |
| 8 | RIPK4 | receptor interacting serine/threonine kinase 4 [Source:HGNC Symbol;Acc:HGNC:496] | 58 | 0.960 | -0.2499 | Yes |
| 9 | BMP4 | bone morphogenetic protein 4 [Source:HGNC Symbol;Acc:HGNC:1071] | 61 | 0.750 | -0.2360 | Yes |
| 10 | EPG5 | ectopic P-granules autophagy protein 5 homolog [Source:HGNC Symbol;Acc:HGNC:29331] | 72 | 0.100 | -0.3047 | Yes |
| 11 | FRAS1 | Fraser extracellular matrix complex subunit 1 [Source:HGNC Symbol;Acc:HGNC:19185] | 73 | 0.010 | -0.2702 | Yes |
| 12 | IRF6 | interferon regulatory factor 6 [Source:HGNC Symbol;Acc:HGNC:6121] | 74 | -0.040 | -0.2357 | Yes |
| 13 | NEK1 | NIMA related kinase 1 [Source:HGNC Symbol;Acc:HGNC:7744] | 75 | -0.090 | -0.2012 | Yes |
| 14 | DYNC2H1 | dynein cytoplasmic 2 heavy chain 1 [Source:HGNC Symbol;Acc:HGNC:2962] | 79 | -0.230 | -0.1977 | Yes |
| 15 | WNT4 | Wnt family member 4 [Source:HGNC Symbol;Acc:HGNC:12783] | 82 | -0.310 | -0.1838 | Yes |
| 16 | MKS1 | MKS transition zone complex subunit 1 [Source:HGNC Symbol;Acc:HGNC:7121] | 88 | -0.480 | -0.2009 | Yes |
| 17 | FGFR1 | fibroblast growth factor receptor 1 [Source:HGNC Symbol;Acc:HGNC:3688] | 90 | -0.570 | -0.1767 | Yes |
| 18 | GRIP1 | glutamate receptor interacting protein 1 [Source:HGNC Symbol;Acc:HGNC:18708] | 94 | -0.750 | -0.1731 | Yes |
| 19 | EVC2 | EvC ciliary complex subunit 2 [Source:HGNC Symbol;Acc:HGNC:19747] | 95 | -0.890 | -0.1386 | Yes |
| 20 | TCTN3 | tectonic family member 3 [Source:HGNC Symbol;Acc:HGNC:24519] | 97 | -1.040 | -0.1145 | Yes |
| 21 | GLI3 | GLI family zinc finger 3 [Source:HGNC Symbol;Acc:HGNC:4319] | 104 | -2.920 | -0.1418 | Yes |
| 22 | CHD7 | chromodomain helicase DNA binding protein 7 [Source:HGNC Symbol;Acc:HGNC:20626] | 105 | -3.230 | -0.1074 | Yes |
| 23 | SPECC1L | sperm antigen with calponin homology and coiled-coil domains 1 like [Source:HGNC Symbol;Acc:HGNC:29022] | 109 | -4.400 | -0.1038 | Yes |
| 24 | WNT5A | Wnt family member 5A [Source:HGNC Symbol;Acc:HGNC:12784] | 115 | -15.700 | -0.1209 | Yes |
| 25 | TP63 | tumor protein p63 [Source:HGNC Symbol;Acc:HGNC:15979] | 116 | -20.700 | -0.0864 | Yes |
| 26 | FREM2 | FRAS1 related extracellular matrix 2 [Source:HGNC Symbol;Acc:HGNC:25396] | 117 | -21.900 | -0.0519 | Yes |
| 27 | B3GLCT | beta 3-glucosyltransferase [Source:HGNC Symbol;Acc:HGNC:20207] | 118 | -22.200 | -0.0174 | Yes |
| 28 | EVC | EvC ciliary complex subunit 1 [Source:HGNC Symbol;Acc:HGNC:3497] | 119 | -30.300 | 0.0171 | Yes |
| 29 | ESCO2 | establishment of sister chromatid cohesion N-acetyltransferase 2 [Source:HGNC Symbol;Acc:HGNC:27230] | 120 | -32.100 | 0.0515 | Yes |
Table: GSEA details [plain text format]

  

Fig 2: HP\_CLEFT\_LIP: Random ES distribution      
 Gene set null distribution of ES for **HP\_CLEFT\_LIP**

  
