## Supplemental File 3 for "Increased Expression of *ZFPM2* Bypasses *SRY* to Drive 46,XX Testicular Development: A New Mechanism of 46,XX DSD": HP_DECREASED_FERTILITY_IN_MALES.html

Details for gene set HP\_DECREASED\_FERTILITY\_IN\_MALES[GSEA]

|  || Dataset | SexDevelopmentGenesPRL\_remapped |
| Phenotype | NoPhenotypeAvailable |
| Upregulated in class | na\_pos |
| GeneSet | HP\_DECREASED\_FERTILITY\_IN\_MALES |
| Enrichment Score (ES) | 0.4490119 |
| Normalized Enrichment Score (NES) | 1.763321 |
| Nominal p-value | 0.021321962 |
| FDR q-value | 0.8742619 |
| FWER p-Value | 1.0 |
Table: GSEA Results Summary

  

Fig 1: Enrichment plot: HP\_DECREASED\_FERTILITY\_IN\_MALES      
 Profile of the Running ES Score & Positions of GeneSet Members on the Rank Ordered List

  

| SYMBOL | TITLE | RANK IN GENE LIST | RANK METRIC SCORE | RUNNING ES | CORE ENRICHMENT || 1 | SOX9 | SRY-box transcription factor 9 [Source:HGNC Symbol;Acc:HGNC:11204] | 1 | 121.000 | 0.0822 | Yes |
| 2 | AMH | anti-Mullerian hormone [Source:HGNC Symbol;Acc:HGNC:464] | 2 | 55.600 | 0.1731 | Yes |
| 3 | VAMP7 | vesicle associated membrane protein 7 [Source:HGNC Symbol;Acc:HGNC:11486] | 8 | 19.500 | 0.2206 | Yes |
| 4 | MAP3K1 | mitogen-activated protein kinase kinase kinase 1 [Source:HGNC Symbol;Acc:HGNC:6848] | 10 | 17.600 | 0.3028 | Yes |
| 5 | LHCGR | luteinizing hormone/choriogonadotropin receptor [Source:HGNC Symbol;Acc:HGNC:6585] | 18 | 11.900 | 0.3328 | Yes |
| 6 | ZFPM2 | "zinc finger protein, FOG family member 2 [Source:HGNC Symbol;Acc:HGNC:16700]" | 31 | 7.040 | 0.3194 | Yes |
| 7 | CYB5A | cytochrome b5 type A [Source:HGNC Symbol;Acc:HGNC:2570] | 36 | 5.850 | 0.3755 | Yes |
| 8 | PTPN11 | protein tyrosine phosphatase non-receptor type 11 [Source:HGNC Symbol;Acc:HGNC:9644] | 39 | 4.970 | 0.4490 | Yes |
| 9 | AR | androgen receptor [Source:HGNC Symbol;Acc:HGNC:644] | 63 | 0.490 | 0.3399 | No |
| 10 | CYP19A1 | cytochrome P450 family 19 subfamily A member 1 [Source:HGNC Symbol;Acc:HGNC:2594] | 81 | -0.300 | 0.2830 | No |
| 11 | WWOX | WW domain containing oxidoreductase [Source:HGNC Symbol;Acc:HGNC:12799] | 113 | -14.500 | 0.1043 | No |
Table: GSEA details [plain text format]

  

Fig 2: HP\_DECREASED\_FERTILITY\_IN\_MALES: Random ES distribution      
 Gene set null distribution of ES for **HP\_DECREASED\_FERTILITY\_IN\_MALES**

  
