## Supplemental File 3 for "Increased Expression of *ZFPM2* Bypasses *SRY* to Drive 46,XX Testicular Development: A New Mechanism of 46,XX DSD": HP_DUPLICATION_OF_HAND_BONES.html

Details for gene set HP\_DUPLICATION\_OF\_HAND\_BONES[GSEA]

|  || Dataset | SexDevelopmentGenesPRL\_remapped |
| Phenotype | NoPhenotypeAvailable |
| Upregulated in class | na\_neg |
| GeneSet | HP\_DUPLICATION\_OF\_HAND\_BONES |
| Enrichment Score (ES) | -0.29444444 |
| Normalized Enrichment Score (NES) | -1.804519 |
| Nominal p-value | 0.0198915 |
| FDR q-value | 0.4717949 |
| FWER p-Value | 1.0 |
Table: GSEA Results Summary

  

Fig 1: Enrichment plot: HP\_DUPLICATION\_OF\_HAND\_BONES      
 Profile of the Running ES Score & Positions of GeneSet Members on the Rank Ordered List

  

| SYMBOL | TITLE | RANK IN GENE LIST | RANK METRIC SCORE | RUNNING ES | CORE ENRICHMENT || 1 | FLNA | filamin A [Source:HGNC Symbol;Acc:HGNC:3754] | 3 | 53.800 | -0.0056 | No |
| 2 | SETBP1 | SET binding protein 1 [Source:HGNC Symbol;Acc:HGNC:15573] | 14 | 14.100 | -0.0889 | No |
| 3 | BCOR | BCL6 corepressor [Source:HGNC Symbol;Acc:HGNC:20893] | 25 | 9.140 | -0.1722 | No |
| 4 | MKKS | MKKS centrosomal shuttling protein [Source:HGNC Symbol;Acc:HGNC:7108] | 34 | 6.190 | -0.2333 | No |
| 5 | FGFR3 | fibroblast growth factor receptor 3 [Source:HGNC Symbol;Acc:HGNC:3690] | 37 | 5.830 | -0.2278 | No |
| 6 | TRIM32 | tripartite motif containing 32 [Source:HGNC Symbol;Acc:HGNC:16380] | 44 | 2.410 | -0.2667 | Yes |
| 7 | BBS9 | Bardet-Biedl syndrome 9 [Source:HGNC Symbol;Acc:HGNC:30000] | 47 | 2.000 | -0.2611 | Yes |
| 8 | BBS2 | Bardet-Biedl syndrome 2 [Source:HGNC Symbol;Acc:HGNC:967] | 48 | 1.960 | -0.2333 | Yes |
| 9 | BBS12 | Bardet-Biedl syndrome 12 [Source:HGNC Symbol;Acc:HGNC:26648] | 49 | 1.960 | -0.2056 | Yes |
| 10 | HESX1 | HESX homeobox 1 [Source:HGNC Symbol;Acc:HGNC:4877] | 51 | 1.570 | -0.1889 | Yes |
| 11 | EFNB1 | ephrin B1 [Source:HGNC Symbol;Acc:HGNC:3226] | 53 | 1.550 | -0.1722 | Yes |
| 12 | WDR35 | WD repeat domain 35 [Source:HGNC Symbol;Acc:HGNC:29250] | 54 | 1.420 | -0.1444 | Yes |
| 13 | DHCR7 | 7-dehydrocholesterol reductase [Source:HGNC Symbol;Acc:HGNC:2860] | 57 | 1.040 | -0.1389 | Yes |
| 14 | FGF10 | fibroblast growth factor 10 [Source:HGNC Symbol;Acc:HGNC:3666] | 59 | 0.890 | -0.1222 | Yes |
| 15 | FGFR2 | fibroblast growth factor receptor 2 [Source:HGNC Symbol;Acc:HGNC:3689] | 60 | 0.760 | -0.0944 | Yes |
| 16 | BMP4 | bone morphogenetic protein 4 [Source:HGNC Symbol;Acc:HGNC:1071] | 61 | 0.750 | -0.0667 | Yes |
| 17 | BBS7 | Bardet-Biedl syndrome 7 [Source:HGNC Symbol;Acc:HGNC:18758] | 66 | 0.400 | -0.0833 | Yes |
| 18 | ARL6 | ADP ribosylation factor like GTPase 6 [Source:HGNC Symbol;Acc:HGNC:13210] | 68 | 0.280 | -0.0667 | Yes |
| 19 | NEK1 | NIMA related kinase 1 [Source:HGNC Symbol;Acc:HGNC:7744] | 75 | -0.090 | -0.1056 | Yes |
| 20 | DYNC2H1 | dynein cytoplasmic 2 heavy chain 1 [Source:HGNC Symbol;Acc:HGNC:2962] | 79 | -0.230 | -0.1111 | Yes |
| 21 | CEP41 | centrosomal protein 41 [Source:HGNC Symbol;Acc:HGNC:12370] | 84 | -0.340 | -0.1278 | Yes |
| 22 | MKS1 | MKS transition zone complex subunit 1 [Source:HGNC Symbol;Acc:HGNC:7121] | 88 | -0.480 | -0.1333 | Yes |
| 23 | FGFR1 | fibroblast growth factor receptor 1 [Source:HGNC Symbol;Acc:HGNC:3688] | 90 | -0.570 | -0.1167 | Yes |
| 24 | ROR2 | receptor tyrosine kinase like orphan receptor 2 [Source:HGNC Symbol;Acc:HGNC:10257] | 91 | -0.580 | -0.0889 | Yes |
| 25 | TTC8 | tetratricopeptide repeat domain 8 [Source:HGNC Symbol;Acc:HGNC:20087] | 92 | -0.640 | -0.0611 | Yes |
| 26 | EVC2 | EvC ciliary complex subunit 2 [Source:HGNC Symbol;Acc:HGNC:19747] | 95 | -0.890 | -0.0556 | Yes |
| 27 | TCTN3 | tectonic family member 3 [Source:HGNC Symbol;Acc:HGNC:24519] | 97 | -1.040 | -0.0389 | Yes |
| 28 | BBS4 | Bardet-Biedl syndrome 4 [Source:HGNC Symbol;Acc:HGNC:969] | 100 | -1.360 | -0.0333 | Yes |
| 29 | BBS10 | Bardet-Biedl syndrome 10 [Source:HGNC Symbol;Acc:HGNC:26291] | 102 | -1.550 | -0.0167 | Yes |
| 30 | GLI3 | GLI family zinc finger 3 [Source:HGNC Symbol;Acc:HGNC:4319] | 104 | -2.920 | -0.0000 | Yes |
| 31 | CHD7 | chromodomain helicase DNA binding protein 7 [Source:HGNC Symbol;Acc:HGNC:20626] | 105 | -3.230 | 0.0278 | Yes |
| 32 | SALL1 | spalt like transcription factor 1 [Source:HGNC Symbol;Acc:HGNC:10524] | 106 | -3.630 | 0.0556 | Yes |
| 33 | BBS5 | Bardet-Biedl syndrome 5 [Source:HGNC Symbol;Acc:HGNC:970] | 114 | -14.600 | 0.0056 | Yes |
| 34 | WNT5A | Wnt family member 5A [Source:HGNC Symbol;Acc:HGNC:12784] | 115 | -15.700 | 0.0333 | Yes |
| 35 | EVC | EvC ciliary complex subunit 1 [Source:HGNC Symbol;Acc:HGNC:3497] | 119 | -30.300 | 0.0278 | Yes |
| 36 | ESCO2 | establishment of sister chromatid cohesion N-acetyltransferase 2 [Source:HGNC Symbol;Acc:HGNC:27230] | 120 | -32.100 | 0.0556 | Yes |
Table: GSEA details [plain text format]

  

Fig 2: HP\_DUPLICATION\_OF\_HAND\_BONES: Random ES distribution      
 Gene set null distribution of ES for **HP\_DUPLICATION\_OF\_HAND\_BONES**

  
