## Supplemental File 3 for "Increased Expression of *ZFPM2* Bypasses *SRY* to Drive 46,XX Testicular Development: A New Mechanism of 46,XX DSD": HP_FINGER_SYNDACTYLY.html

Details for gene set HP\_FINGER\_SYNDACTYLY[GSEA]

|  || Dataset | SexDevelopmentGenesPRL\_remapped |
| Phenotype | NoPhenotypeAvailable |
| Upregulated in class | na\_neg |
| GeneSet | HP\_FINGER\_SYNDACTYLY |
| Enrichment Score (ES) | -0.35720602 |
| Normalized Enrichment Score (NES) | -2.2442381 |
| Nominal p-value | 0.0 |
| FDR q-value | 0.61183876 |
| FWER p-Value | 0.438 |
Table: GSEA Results Summary

  

Fig 1: Enrichment plot: HP\_FINGER\_SYNDACTYLY      
 Profile of the Running ES Score & Positions of GeneSet Members on the Rank Ordered List

  

| SYMBOL | TITLE | RANK IN GENE LIST | RANK METRIC SCORE | RUNNING ES | CORE ENRICHMENT || 1 | BCOR | BCL6 corepressor [Source:HGNC Symbol;Acc:HGNC:20893] | 25 | 9.140 | -0.2617 | No |
| 2 | MED12 | mediator complex subunit 12 [Source:HGNC Symbol;Acc:HGNC:11957] | 33 | 6.470 | -0.3165 | No |
| 3 | MKKS | MKKS centrosomal shuttling protein [Source:HGNC Symbol;Acc:HGNC:7108] | 34 | 6.190 | -0.2909 | No |
| 4 | FGFR3 | fibroblast growth factor receptor 3 [Source:HGNC Symbol;Acc:HGNC:3690] | 37 | 5.830 | -0.2882 | No |
| 5 | TRIM32 | tripartite motif containing 32 [Source:HGNC Symbol;Acc:HGNC:16380] | 44 | 2.410 | -0.3316 | Yes |
| 6 | BBS9 | Bardet-Biedl syndrome 9 [Source:HGNC Symbol;Acc:HGNC:30000] | 47 | 2.000 | -0.3289 | Yes |
| 7 | BBS2 | Bardet-Biedl syndrome 2 [Source:HGNC Symbol;Acc:HGNC:967] | 48 | 1.960 | -0.3033 | Yes |
| 8 | BBS12 | Bardet-Biedl syndrome 12 [Source:HGNC Symbol;Acc:HGNC:26648] | 49 | 1.960 | -0.2776 | Yes |
| 9 | EFNB1 | ephrin B1 [Source:HGNC Symbol;Acc:HGNC:3226] | 53 | 1.550 | -0.2865 | Yes |
| 10 | WDR35 | WD repeat domain 35 [Source:HGNC Symbol;Acc:HGNC:29250] | 54 | 1.420 | -0.2608 | Yes |
| 11 | DHCR7 | 7-dehydrocholesterol reductase [Source:HGNC Symbol;Acc:HGNC:2860] | 57 | 1.040 | -0.2582 | Yes |
| 12 | RIPK4 | receptor interacting serine/threonine kinase 4 [Source:HGNC Symbol;Acc:HGNC:496] | 58 | 0.960 | -0.2325 | Yes |
| 13 | FGF10 | fibroblast growth factor 10 [Source:HGNC Symbol;Acc:HGNC:3666] | 59 | 0.890 | -0.2069 | Yes |
| 14 | FGFR2 | fibroblast growth factor receptor 2 [Source:HGNC Symbol;Acc:HGNC:3689] | 60 | 0.760 | -0.1813 | Yes |
| 15 | BMP4 | bone morphogenetic protein 4 [Source:HGNC Symbol;Acc:HGNC:1071] | 61 | 0.750 | -0.1556 | Yes |
| 16 | CKAP2L | cytoskeleton associated protein 2 like [Source:HGNC Symbol;Acc:HGNC:26877] | 64 | 0.460 | -0.1530 | Yes |
| 17 | BBS7 | Bardet-Biedl syndrome 7 [Source:HGNC Symbol;Acc:HGNC:18758] | 66 | 0.400 | -0.1388 | Yes |
| 18 | ARL6 | ADP ribosylation factor like GTPase 6 [Source:HGNC Symbol;Acc:HGNC:13210] | 68 | 0.280 | -0.1247 | Yes |
| 19 | PTDSS1 | phosphatidylserine synthase 1 [Source:HGNC Symbol;Acc:HGNC:9587] | 71 | 0.100 | -0.1220 | Yes |
| 20 | FRAS1 | Fraser extracellular matrix complex subunit 1 [Source:HGNC Symbol;Acc:HGNC:19185] | 73 | 0.010 | -0.1079 | Yes |
| 21 | IRF6 | interferon regulatory factor 6 [Source:HGNC Symbol;Acc:HGNC:6121] | 74 | -0.040 | -0.0822 | Yes |
| 22 | NEK1 | NIMA related kinase 1 [Source:HGNC Symbol;Acc:HGNC:7744] | 75 | -0.090 | -0.0566 | Yes |
| 23 | FAT4 | FAT atypical cadherin 4 [Source:HGNC Symbol;Acc:HGNC:23109] | 76 | -0.100 | -0.0309 | Yes |
| 24 | MKS1 | MKS transition zone complex subunit 1 [Source:HGNC Symbol;Acc:HGNC:7121] | 88 | -0.480 | -0.1317 | Yes |
| 25 | FGFR1 | fibroblast growth factor receptor 1 [Source:HGNC Symbol;Acc:HGNC:3688] | 90 | -0.570 | -0.1176 | Yes |
| 26 | ROR2 | receptor tyrosine kinase like orphan receptor 2 [Source:HGNC Symbol;Acc:HGNC:10257] | 91 | -0.580 | -0.0920 | Yes |
| 27 | TTC8 | tetratricopeptide repeat domain 8 [Source:HGNC Symbol;Acc:HGNC:20087] | 92 | -0.640 | -0.0663 | Yes |
| 28 | GRIP1 | glutamate receptor interacting protein 1 [Source:HGNC Symbol;Acc:HGNC:18708] | 94 | -0.750 | -0.0522 | Yes |
| 29 | TCTN3 | tectonic family member 3 [Source:HGNC Symbol;Acc:HGNC:24519] | 97 | -1.040 | -0.0495 | Yes |
| 30 | BBS4 | Bardet-Biedl syndrome 4 [Source:HGNC Symbol;Acc:HGNC:969] | 100 | -1.360 | -0.0469 | Yes |
| 31 | BBS10 | Bardet-Biedl syndrome 10 [Source:HGNC Symbol;Acc:HGNC:26291] | 102 | -1.550 | -0.0327 | Yes |
| 32 | GLI3 | GLI family zinc finger 3 [Source:HGNC Symbol;Acc:HGNC:4319] | 104 | -2.920 | -0.0186 | Yes |
| 33 | SALL1 | spalt like transcription factor 1 [Source:HGNC Symbol;Acc:HGNC:10524] | 106 | -3.630 | -0.0044 | Yes |
| 34 | SPECC1L | sperm antigen with calponin homology and coiled-coil domains 1 like [Source:HGNC Symbol;Acc:HGNC:29022] | 109 | -4.400 | -0.0018 | Yes |
| 35 | BBS5 | Bardet-Biedl syndrome 5 [Source:HGNC Symbol;Acc:HGNC:970] | 114 | -14.600 | -0.0221 | Yes |
| 36 | WNT5A | Wnt family member 5A [Source:HGNC Symbol;Acc:HGNC:12784] | 115 | -15.700 | 0.0035 | Yes |
| 37 | TP63 | tumor protein p63 [Source:HGNC Symbol;Acc:HGNC:15979] | 116 | -20.700 | 0.0292 | Yes |
| 38 | FREM2 | FRAS1 related extracellular matrix 2 [Source:HGNC Symbol;Acc:HGNC:25396] | 117 | -21.900 | 0.0548 | Yes |
| 39 | ESCO2 | establishment of sister chromatid cohesion N-acetyltransferase 2 [Source:HGNC Symbol;Acc:HGNC:27230] | 120 | -32.100 | 0.0575 | Yes |
Table: GSEA details [plain text format]

  

Fig 2: HP\_FINGER\_SYNDACTYLY: Random ES distribution      
 Gene set null distribution of ES for **HP\_FINGER\_SYNDACTYLY**

  
