## Supplemental File 3 for "Increased Expression of *ZFPM2* Bypasses *SRY* to Drive 46,XX Testicular Development: A New Mechanism of 46,XX DSD": HP_GONOSOMAL_INHERITANCE.html

Details for gene set HP\_GONOSOMAL\_INHERITANCE[GSEA]

|  || Dataset | SexDevelopmentGenesPRL\_remapped |
| Phenotype | NoPhenotypeAvailable |
| Upregulated in class | na\_pos |
| GeneSet | HP\_GONOSOMAL\_INHERITANCE |
| Enrichment Score (ES) | 0.43859652 |
| Normalized Enrichment Score (NES) | 1.7867757 |
| Nominal p-value | 0.009784736 |
| FDR q-value | 1.0 |
| FWER p-Value | 1.0 |
Table: GSEA Results Summary

  

Fig 1: Enrichment plot: HP\_GONOSOMAL\_INHERITANCE      
 Profile of the Running ES Score & Positions of GeneSet Members on the Rank Ordered List

  

| SYMBOL | TITLE | RANK IN GENE LIST | RANK METRIC SCORE | RUNNING ES | CORE ENRICHMENT || 1 | FLNA | filamin A [Source:HGNC Symbol;Acc:HGNC:3754] | 3 | 53.800 | 0.0570 | Yes |
| 2 | ANOS1 | anosmin 1 [Source:HGNC Symbol;Acc:HGNC:6211] | 7 | 25.200 | 0.1140 | Yes |
| 3 | OPHN1 | oligophrenin 1 [Source:HGNC Symbol;Acc:HGNC:8148] | 11 | 15.900 | 0.1711 | Yes |
| 4 | BCOR | BCL6 corepressor [Source:HGNC Symbol;Acc:HGNC:20893] | 25 | 9.140 | 0.1404 | Yes |
| 5 | MID1 | midline 1 [Source:HGNC Symbol;Acc:HGNC:7095] | 26 | 8.770 | 0.2237 | Yes |
| 6 | ATRX | ATRX chromatin remodeler [Source:HGNC Symbol;Acc:HGNC:886] | 27 | 8.350 | 0.3070 | Yes |
| 7 | MAMLD1 | mastermind like domain containing 1 [Source:HGNC Symbol;Acc:HGNC:2568] | 30 | 7.350 | 0.3728 | Yes |
| 8 | MED12 | mediator complex subunit 12 [Source:HGNC Symbol;Acc:HGNC:11957] | 33 | 6.470 | 0.4386 | Yes |
| 9 | EFNB1 | ephrin B1 [Source:HGNC Symbol;Acc:HGNC:3226] | 53 | 1.550 | 0.3553 | No |
| 10 | AR | androgen receptor [Source:HGNC Symbol;Acc:HGNC:644] | 63 | 0.490 | 0.3596 | No |
| 11 | ARL6 | ADP ribosylation factor like GTPase 6 [Source:HGNC Symbol;Acc:HGNC:13210] | 68 | 0.280 | 0.4079 | No |
| 12 | HCCS | holocytochrome c synthase [Source:HGNC Symbol;Acc:HGNC:4837] | 86 | -0.400 | 0.3421 | No |
Table: GSEA details [plain text format]

  

Fig 2: HP\_GONOSOMAL\_INHERITANCE: Random ES distribution      
 Gene set null distribution of ES for **HP\_GONOSOMAL\_INHERITANCE**

  
