## Supplemental File 3 for "Increased Expression of *ZFPM2* Bypasses *SRY* to Drive 46,XX Testicular Development: A New Mechanism of 46,XX DSD": HP_HETEROTROPIA.html

Details for gene set HP\_HETEROTROPIA[GSEA]

|  || Dataset | SexDevelopmentGenesPRL\_remapped |
| Phenotype | NoPhenotypeAvailable |
| Upregulated in class | na\_pos |
| GeneSet | HP\_HETEROTROPIA |
| Enrichment Score (ES) | 0.45172414 |
| Normalized Enrichment Score (NES) | 1.6808664 |
| Nominal p-value | 0.036 |
| FDR q-value | 0.874847 |
| FWER p-Value | 1.0 |
Table: GSEA Results Summary

  

Fig 1: Enrichment plot: HP\_HETEROTROPIA      
 Profile of the Running ES Score & Positions of GeneSet Members on the Rank Ordered List

  

| SYMBOL | TITLE | RANK IN GENE LIST | RANK METRIC SCORE | RUNNING ES | CORE ENRICHMENT || 1 | ZEB2 | zinc finger E-box binding homeobox 2 [Source:HGNC Symbol;Acc:HGNC:14881] | 5 | 36.500 | 0.0569 | Yes |
| 2 | PDE4D | phosphodiesterase 4D [Source:HGNC Symbol;Acc:HGNC:8783] | 9 | 18.600 | 0.1310 | Yes |
| 3 | BCOR | BCL6 corepressor [Source:HGNC Symbol;Acc:HGNC:20893] | 25 | 9.140 | 0.1017 | Yes |
| 4 | ATRX | ATRX chromatin remodeler [Source:HGNC Symbol;Acc:HGNC:886] | 27 | 8.350 | 0.1931 | Yes |
| 5 | CYB5A | cytochrome b5 type A [Source:HGNC Symbol;Acc:HGNC:2570] | 36 | 5.850 | 0.2241 | Yes |
| 6 | FGFR3 | fibroblast growth factor receptor 3 [Source:HGNC Symbol;Acc:HGNC:3690] | 37 | 5.830 | 0.3241 | Yes |
| 7 | EFNB1 | ephrin B1 [Source:HGNC Symbol;Acc:HGNC:3226] | 53 | 1.550 | 0.2948 | Yes |
| 8 | FGF10 | fibroblast growth factor 10 [Source:HGNC Symbol;Acc:HGNC:3666] | 59 | 0.890 | 0.3517 | Yes |
| 9 | FGFR2 | fibroblast growth factor receptor 2 [Source:HGNC Symbol;Acc:HGNC:3689] | 60 | 0.760 | 0.4517 | Yes |
| 10 | TCTN3 | tectonic family member 3 [Source:HGNC Symbol;Acc:HGNC:24519] | 97 | -1.040 | 0.2414 | No |
Table: GSEA details [plain text format]

  

Fig 2: HP\_HETEROTROPIA: Random ES distribution      
 Gene set null distribution of ES for **HP\_HETEROTROPIA**

  
