## Supplemental File 3 for "Increased Expression of *ZFPM2* Bypasses *SRY* to Drive 46,XX Testicular Development: A New Mechanism of 46,XX DSD": HP_HYPERACTIVITY.html

Details for gene set HP\_HYPERACTIVITY[GSEA]

|  || Dataset | SexDevelopmentGenesPRL\_remapped |
| Phenotype | NoPhenotypeAvailable |
| Upregulated in class | na\_pos |
| GeneSet | HP\_HYPERACTIVITY |
| Enrichment Score (ES) | 0.3839286 |
| Normalized Enrichment Score (NES) | 1.6395985 |
| Nominal p-value | 0.041257367 |
| FDR q-value | 0.87353474 |
| FWER p-Value | 1.0 |
Table: GSEA Results Summary

  

Fig 1: Enrichment plot: HP\_HYPERACTIVITY      
 Profile of the Running ES Score & Positions of GeneSet Members on the Rank Ordered List

  

| SYMBOL | TITLE | RANK IN GENE LIST | RANK METRIC SCORE | RUNNING ES | CORE ENRICHMENT || 1 | PDE4D | phosphodiesterase 4D [Source:HGNC Symbol;Acc:HGNC:8783] | 9 | 18.600 | -0.0089 | Yes |
| 2 | OPHN1 | oligophrenin 1 [Source:HGNC Symbol;Acc:HGNC:8148] | 11 | 15.900 | 0.0536 | Yes |
| 3 | SETBP1 | SET binding protein 1 [Source:HGNC Symbol;Acc:HGNC:15573] | 14 | 14.100 | 0.1071 | Yes |
| 4 | PCNT | pericentrin [Source:HGNC Symbol;Acc:HGNC:16068] | 17 | 12.400 | 0.1607 | Yes |
| 5 | LHCGR | luteinizing hormone/choriogonadotropin receptor [Source:HGNC Symbol;Acc:HGNC:6585] | 18 | 11.900 | 0.2321 | Yes |
| 6 | CREBBP | CREB binding protein [Source:HGNC Symbol;Acc:HGNC:2348] | 20 | 11.800 | 0.2946 | Yes |
| 7 | ATRX | ATRX chromatin remodeler [Source:HGNC Symbol;Acc:HGNC:886] | 27 | 8.350 | 0.3125 | Yes |
| 8 | MED12 | mediator complex subunit 12 [Source:HGNC Symbol;Acc:HGNC:11957] | 33 | 6.470 | 0.3393 | Yes |
| 9 | FGFR3 | fibroblast growth factor receptor 3 [Source:HGNC Symbol;Acc:HGNC:3690] | 37 | 5.830 | 0.3839 | Yes |
| 10 | DHCR7 | 7-dehydrocholesterol reductase [Source:HGNC Symbol;Acc:HGNC:2860] | 57 | 1.040 | 0.2857 | No |
| 11 | CDKN1C | cyclin dependent kinase inhibitor 1C [Source:HGNC Symbol;Acc:HGNC:1786] | 89 | -0.530 | 0.0804 | No |
| 12 | FGFR1 | fibroblast growth factor receptor 1 [Source:HGNC Symbol;Acc:HGNC:3688] | 90 | -0.570 | 0.1518 | No |
| 13 | CHD7 | chromodomain helicase DNA binding protein 7 [Source:HGNC Symbol;Acc:HGNC:20626] | 105 | -3.230 | 0.0982 | No |
| 14 | WWOX | WW domain containing oxidoreductase [Source:HGNC Symbol;Acc:HGNC:12799] | 113 | -14.500 | 0.1071 | No |
Table: GSEA details [plain text format]

  

Fig 2: HP\_HYPERACTIVITY: Random ES distribution      
 Gene set null distribution of ES for **HP\_HYPERACTIVITY**

  
