## Supplemental File 3 for "Increased Expression of *ZFPM2* Bypasses *SRY* to Drive 46,XX Testicular Development: A New Mechanism of 46,XX DSD": HP_HYPERTONIA.html

Details for gene set HP\_HYPERTONIA[GSEA]

|  || Dataset | SexDevelopmentGenesPRL\_remapped |
| Phenotype | NoPhenotypeAvailable |
| Upregulated in class | na\_pos |
| GeneSet | HP\_HYPERTONIA |
| Enrichment Score (ES) | 0.34052888 |
| Normalized Enrichment Score (NES) | 1.6227976 |
| Nominal p-value | 0.049212597 |
| FDR q-value | 0.8681112 |
| FWER p-Value | 1.0 |
Table: GSEA Results Summary

  

Fig 1: Enrichment plot: HP\_HYPERTONIA      
 Profile of the Running ES Score & Positions of GeneSet Members on the Rank Ordered List

  

| SYMBOL | TITLE | RANK IN GENE LIST | RANK METRIC SCORE | RUNNING ES | CORE ENRICHMENT || 1 | FLNA | filamin A [Source:HGNC Symbol;Acc:HGNC:3754] | 3 | 53.800 | 0.0313 | Yes |
| 2 | DHCR24 | 24-dehydrocholesterol reductase [Source:HGNC Symbol;Acc:HGNC:2859] | 4 | 47.700 | 0.0901 | Yes |
| 3 | ZEB2 | zinc finger E-box binding homeobox 2 [Source:HGNC Symbol;Acc:HGNC:14881] | 5 | 36.500 | 0.1489 | Yes |
| 4 | OPHN1 | oligophrenin 1 [Source:HGNC Symbol;Acc:HGNC:8148] | 11 | 15.900 | 0.1619 | Yes |
| 5 | HSD17B4 | hydroxysteroid 17-beta dehydrogenase 4 [Source:HGNC Symbol;Acc:HGNC:5213] | 13 | 15.000 | 0.2115 | Yes |
| 6 | SETBP1 | SET binding protein 1 [Source:HGNC Symbol;Acc:HGNC:15573] | 14 | 14.100 | 0.2704 | Yes |
| 7 | BCOR | BCL6 corepressor [Source:HGNC Symbol;Acc:HGNC:20893] | 25 | 9.140 | 0.2375 | Yes |
| 8 | ATRX | ATRX chromatin remodeler [Source:HGNC Symbol;Acc:HGNC:886] | 27 | 8.350 | 0.2871 | Yes |
| 9 | TOE1 | "target of EGR1, exonuclease [Source:HGNC Symbol;Acc:HGNC:15954]" | 35 | 5.880 | 0.2817 | Yes |
| 10 | CYB5A | cytochrome b5 type A [Source:HGNC Symbol;Acc:HGNC:2570] | 36 | 5.850 | 0.3405 | Yes |
| 11 | DHCR7 | 7-dehydrocholesterol reductase [Source:HGNC Symbol;Acc:HGNC:2860] | 57 | 1.040 | 0.2159 | No |
| 12 | CKAP2L | cytoskeleton associated protein 2 like [Source:HGNC Symbol;Acc:HGNC:26877] | 64 | 0.460 | 0.2196 | No |
| 13 | FIG4 | FIG4 phosphoinositide 5-phosphatase [Source:HGNC Symbol;Acc:HGNC:16873] | 65 | 0.420 | 0.2785 | No |
| 14 | NEK1 | NIMA related kinase 1 [Source:HGNC Symbol;Acc:HGNC:7744] | 75 | -0.090 | 0.2547 | No |
| 15 | PEX1 | peroxisomal biogenesis factor 1 [Source:HGNC Symbol;Acc:HGNC:8850] | 83 | -0.330 | 0.2493 | No |
| 16 | FGFR1 | fibroblast growth factor receptor 1 [Source:HGNC Symbol;Acc:HGNC:3688] | 90 | -0.570 | 0.2531 | No |
| 17 | WWOX | WW domain containing oxidoreductase [Source:HGNC Symbol;Acc:HGNC:12799] | 113 | -14.500 | 0.1101 | No |
Table: GSEA details [plain text format]

  

Fig 2: HP\_HYPERTONIA: Random ES distribution      
 Gene set null distribution of ES for **HP\_HYPERTONIA**

  
