## Supplemental File 3 for "Increased Expression of *ZFPM2* Bypasses *SRY* to Drive 46,XX Testicular Development: A New Mechanism of 46,XX DSD": HP_HYPOGONADOTROPIC_HYPOGONADISM.html

Details for gene set HP\_HYPOGONADOTROPIC\_HYPOGONADISM[GSEA]

|  || Dataset | SexDevelopmentGenesPRL\_remapped |
| Phenotype | NoPhenotypeAvailable |
| Upregulated in class | na\_pos |
| GeneSet | HP\_HYPOGONADOTROPIC\_HYPOGONADISM |
| Enrichment Score (ES) | 0.32415152 |
| Normalized Enrichment Score (NES) | 1.5754095 |
| Nominal p-value | 0.048780486 |
| FDR q-value | 0.9964663 |
| FWER p-Value | 1.0 |
Table: GSEA Results Summary

  

Fig 1: Enrichment plot: HP\_HYPOGONADOTROPIC\_HYPOGONADISM      
 Profile of the Running ES Score & Positions of GeneSet Members on the Rank Ordered List

  

| SYMBOL | TITLE | RANK IN GENE LIST | RANK METRIC SCORE | RUNNING ES | CORE ENRICHMENT || 1 | SOX9 | SRY-box transcription factor 9 [Source:HGNC Symbol;Acc:HGNC:11204] | 1 | 121.000 | 0.0433 | Yes |
| 2 | HSD17B3 | hydroxysteroid 17-beta dehydrogenase 3 [Source:HGNC Symbol;Acc:HGNC:5212] | 6 | 27.700 | 0.0585 | Yes |
| 3 | ANOS1 | anosmin 1 [Source:HGNC Symbol;Acc:HGNC:6211] | 7 | 25.200 | 0.1112 | Yes |
| 4 | MAP3K1 | mitogen-activated protein kinase kinase kinase 1 [Source:HGNC Symbol;Acc:HGNC:6848] | 10 | 17.600 | 0.1451 | Yes |
| 5 | WDR11 | WD repeat domain 11 [Source:HGNC Symbol;Acc:HGNC:13831] | 12 | 15.800 | 0.1884 | Yes |
| 6 | SOS1 | SOS Ras/Rac guanine nucleotide exchange factor 1 [Source:HGNC Symbol;Acc:HGNC:11187] | 15 | 12.800 | 0.2223 | Yes |
| 7 | DMRT1 | doublesex and mab-3 related transcription factor 1 [Source:HGNC Symbol;Acc:HGNC:2934] | 16 | 12.800 | 0.2750 | Yes |
| 8 | GNRH1 | gonadotropin releasing hormone 1 [Source:HGNC Symbol;Acc:HGNC:4419] | 23 | 9.790 | 0.2715 | Yes |
| 9 | HS6ST1 | heparan sulfate 6-O-sulfotransferase 1 [Source:HGNC Symbol;Acc:HGNC:5201] | 24 | 9.710 | 0.3242 | Yes |
| 10 | PTPN11 | protein tyrosine phosphatase non-receptor type 11 [Source:HGNC Symbol;Acc:HGNC:9644] | 39 | 4.970 | 0.2459 | No |
| 11 | HFE | homeostatic iron regulator [Source:HGNC Symbol;Acc:HGNC:4886] | 43 | 2.890 | 0.2705 | No |
| 12 | HESX1 | HESX homeobox 1 [Source:HGNC Symbol;Acc:HGNC:4877] | 51 | 1.570 | 0.2577 | No |
| 13 | NSMF | NMDA receptor synaptonuclear signaling and neuronal migration factor [Source:HGNC Symbol;Acc:HGNC:29843] | 55 | 1.360 | 0.2823 | No |
| 14 | SEMA3A | semaphorin 3A [Source:HGNC Symbol;Acc:HGNC:10723] | 67 | 0.310 | 0.2322 | No |
| 15 | CBX2 | chromobox 2 [Source:HGNC Symbol;Acc:HGNC:1552] | 87 | -0.410 | 0.1072 | No |
| 16 | FGFR1 | fibroblast growth factor receptor 1 [Source:HGNC Symbol;Acc:HGNC:3688] | 90 | -0.570 | 0.1412 | No |
| 17 | IL17RD | interleukin 17 receptor D [Source:HGNC Symbol;Acc:HGNC:17616] | 98 | -1.150 | 0.1284 | No |
| 18 | CHD7 | chromodomain helicase DNA binding protein 7 [Source:HGNC Symbol;Acc:HGNC:20626] | 105 | -3.230 | 0.1249 | No |
| 19 | TP63 | tumor protein p63 [Source:HGNC Symbol;Acc:HGNC:15979] | 116 | -20.700 | 0.0841 | No |
Table: GSEA details [plain text format]

  

Fig 2: HP\_HYPOGONADOTROPIC\_HYPOGONADISM: Random ES distribution      
 Gene set null distribution of ES for **HP\_HYPOGONADOTROPIC\_HYPOGONADISM**

  
