## Supplemental File 3 for "Increased Expression of *ZFPM2* Bypasses *SRY* to Drive 46,XX Testicular Development: A New Mechanism of 46,XX DSD": HP_INFERTILITY.html

Details for gene set HP\_INFERTILITY[GSEA]

|  || Dataset | SexDevelopmentGenesPRL\_remapped |
| Phenotype | NoPhenotypeAvailable |
| Upregulated in class | na\_pos |
| GeneSet | HP\_INFERTILITY |
| Enrichment Score (ES) | 0.4351852 |
| Normalized Enrichment Score (NES) | 2.1027203 |
| Nominal p-value | 0.002 |
| FDR q-value | 0.4899425 |
| FWER p-Value | 0.788 |
Table: GSEA Results Summary

  

Fig 1: Enrichment plot: HP\_INFERTILITY      
 Profile of the Running ES Score & Positions of GeneSet Members on the Rank Ordered List

  

| SYMBOL | TITLE | RANK IN GENE LIST | RANK METRIC SCORE | RUNNING ES | CORE ENRICHMENT || 1 | SOX9 | SRY-box transcription factor 9 [Source:HGNC Symbol;Acc:HGNC:11204] | 1 | 121.000 | 0.0463 | Yes |
| 2 | AMH | anti-Mullerian hormone [Source:HGNC Symbol;Acc:HGNC:464] | 2 | 55.600 | 0.1019 | Yes |
| 3 | HSD17B3 | hydroxysteroid 17-beta dehydrogenase 3 [Source:HGNC Symbol;Acc:HGNC:5212] | 6 | 27.700 | 0.1296 | Yes |
| 4 | VAMP7 | vesicle associated membrane protein 7 [Source:HGNC Symbol;Acc:HGNC:11486] | 8 | 19.500 | 0.1759 | Yes |
| 5 | MAP3K1 | mitogen-activated protein kinase kinase kinase 1 [Source:HGNC Symbol;Acc:HGNC:6848] | 10 | 17.600 | 0.2222 | Yes |
| 6 | WDR11 | WD repeat domain 11 [Source:HGNC Symbol;Acc:HGNC:13831] | 12 | 15.800 | 0.2685 | Yes |
| 7 | LHCGR | luteinizing hormone/choriogonadotropin receptor [Source:HGNC Symbol;Acc:HGNC:6585] | 18 | 11.900 | 0.2778 | Yes |
| 8 | H6PD | hexose-6-phosphate dehydrogenase/glucose 1-dehydrogenase [Source:HGNC Symbol;Acc:HGNC:4795] | 21 | 11.600 | 0.3148 | Yes |
| 9 | GNRH1 | gonadotropin releasing hormone 1 [Source:HGNC Symbol;Acc:HGNC:4419] | 23 | 9.790 | 0.3611 | Yes |
| 10 | NR3C1 | nuclear receptor subfamily 3 group C member 1 [Source:HGNC Symbol;Acc:HGNC:7978] | 29 | 7.690 | 0.3704 | Yes |
| 11 | ZFPM2 | "zinc finger protein, FOG family member 2 [Source:HGNC Symbol;Acc:HGNC:16700]" | 31 | 7.040 | 0.4167 | Yes |
| 12 | PTPN11 | protein tyrosine phosphatase non-receptor type 11 [Source:HGNC Symbol;Acc:HGNC:9644] | 39 | 4.970 | 0.4074 | Yes |
| 13 | HFE | homeostatic iron regulator [Source:HGNC Symbol;Acc:HGNC:4886] | 43 | 2.890 | 0.4352 | Yes |
| 14 | HESX1 | HESX homeobox 1 [Source:HGNC Symbol;Acc:HGNC:4877] | 51 | 1.570 | 0.4259 | No |
| 15 | AR | androgen receptor [Source:HGNC Symbol;Acc:HGNC:644] | 63 | 0.490 | 0.3796 | No |
| 16 | CYP19A1 | cytochrome P450 family 19 subfamily A member 1 [Source:HGNC Symbol;Acc:HGNC:2594] | 81 | -0.300 | 0.2778 | No |
| 17 | WWOX | WW domain containing oxidoreductase [Source:HGNC Symbol;Acc:HGNC:12799] | 113 | -14.500 | 0.0463 | No |
| 18 | FOXL2 | forkhead box L2 [Source:HGNC Symbol;Acc:HGNC:1092] | 122 | -40.400 | 0.0278 | No |
Table: GSEA details [plain text format]

  

Fig 2: HP\_INFERTILITY: Random ES distribution      
 Gene set null distribution of ES for **HP\_INFERTILITY**

  
