## Supplemental File 3 for "Increased Expression of *ZFPM2* Bypasses *SRY* to Drive 46,XX Testicular Development: A New Mechanism of 46,XX DSD": HP_INTELLECTUAL_DISABILITY_SEVERE.html

Details for gene set HP\_INTELLECTUAL\_DISABILITY\_SEVERE[GSEA]

|  || Dataset | SexDevelopmentGenesPRL\_remapped |
| Phenotype | NoPhenotypeAvailable |
| Upregulated in class | na\_pos |
| GeneSet | HP\_INTELLECTUAL\_DISABILITY\_SEVERE |
| Enrichment Score (ES) | 0.4664032 |
| Normalized Enrichment Score (NES) | 1.7795436 |
| Nominal p-value | 0.013888889 |
| FDR q-value | 0.9612762 |
| FWER p-Value | 1.0 |
Table: GSEA Results Summary

  

Fig 1: Enrichment plot: HP\_INTELLECTUAL\_DISABILITY\_SEVERE      
 Profile of the Running ES Score & Positions of GeneSet Members on the Rank Ordered List

  

| SYMBOL | TITLE | RANK IN GENE LIST | RANK METRIC SCORE | RUNNING ES | CORE ENRICHMENT || 1 | ZEB2 | zinc finger E-box binding homeobox 2 [Source:HGNC Symbol;Acc:HGNC:14881] | 5 | 36.500 | 0.0474 | Yes |
| 2 | OPHN1 | oligophrenin 1 [Source:HGNC Symbol;Acc:HGNC:8148] | 11 | 15.900 | 0.0949 | Yes |
| 3 | SETBP1 | SET binding protein 1 [Source:HGNC Symbol;Acc:HGNC:15573] | 14 | 14.100 | 0.1684 | Yes |
| 4 | ATRX | ATRX chromatin remodeler [Source:HGNC Symbol;Acc:HGNC:886] | 27 | 8.350 | 0.1549 | Yes |
| 5 | KAT6B | lysine acetyltransferase 6B [Source:HGNC Symbol;Acc:HGNC:17582] | 28 | 8.070 | 0.2458 | Yes |
| 6 | MED12 | mediator complex subunit 12 [Source:HGNC Symbol;Acc:HGNC:11957] | 33 | 6.470 | 0.3020 | Yes |
| 7 | CYB5A | cytochrome b5 type A [Source:HGNC Symbol;Acc:HGNC:2570] | 36 | 5.850 | 0.3755 | Yes |
| 8 | FGFR3 | fibroblast growth factor receptor 3 [Source:HGNC Symbol;Acc:HGNC:3690] | 37 | 5.830 | 0.4664 | Yes |
| 9 | FAT4 | FAT atypical cadherin 4 [Source:HGNC Symbol;Acc:HGNC:23109] | 76 | -0.100 | 0.2269 | No |
| 10 | PEX1 | peroxisomal biogenesis factor 1 [Source:HGNC Symbol;Acc:HGNC:8850] | 83 | -0.330 | 0.2656 | No |
| 11 | GLI3 | GLI family zinc finger 3 [Source:HGNC Symbol;Acc:HGNC:4319] | 104 | -2.920 | 0.1826 | No |
Table: GSEA details [plain text format]

  

Fig 2: HP\_INTELLECTUAL\_DISABILITY\_SEVERE: Random ES distribution      
 Gene set null distribution of ES for **HP\_INTELLECTUAL\_DISABILITY\_SEVERE**

  
