## Supplemental File 3 for "Increased Expression of *ZFPM2* Bypasses *SRY* to Drive 46,XX Testicular Development: A New Mechanism of 46,XX DSD": HP_OLIGOHYDRAMNIOS.html

Details for gene set HP\_OLIGOHYDRAMNIOS[GSEA]

|  || Dataset | SexDevelopmentGenesPRL\_remapped |
| Phenotype | NoPhenotypeAvailable |
| Upregulated in class | na\_neg |
| GeneSet | HP\_OLIGOHYDRAMNIOS |
| Enrichment Score (ES) | -0.48965517 |
| Normalized Enrichment Score (NES) | -1.836674 |
| Nominal p-value | 0.016563147 |
| FDR q-value | 0.5006044 |
| FWER p-Value | 1.0 |
Table: GSEA Results Summary

  

Fig 1: Enrichment plot: HP\_OLIGOHYDRAMNIOS      
 Profile of the Running ES Score & Positions of GeneSet Members on the Rank Ordered List

  

| SYMBOL | TITLE | RANK IN GENE LIST | RANK METRIC SCORE | RUNNING ES | CORE ENRICHMENT || 1 | TMEM70 | transmembrane protein 70 [Source:HGNC Symbol;Acc:HGNC:26050] | 40 | 3.900 | -0.2448 | No |
| 2 | TALDO1 | transaldolase 1 [Source:HGNC Symbol;Acc:HGNC:11559] | 69 | 0.210 | -0.3862 | No |
| 3 | WNT4 | Wnt family member 4 [Source:HGNC Symbol;Acc:HGNC:12783] | 82 | -0.310 | -0.3897 | Yes |
| 4 | MKS1 | MKS transition zone complex subunit 1 [Source:HGNC Symbol;Acc:HGNC:7121] | 88 | -0.480 | -0.3328 | Yes |
| 5 | CDKN1C | cyclin dependent kinase inhibitor 1C [Source:HGNC Symbol;Acc:HGNC:1786] | 89 | -0.530 | -0.2328 | Yes |
| 6 | TCTN3 | tectonic family member 3 [Source:HGNC Symbol;Acc:HGNC:24519] | 97 | -1.040 | -0.1931 | Yes |
| 7 | GLI3 | GLI family zinc finger 3 [Source:HGNC Symbol;Acc:HGNC:4319] | 104 | -2.920 | -0.1448 | Yes |
| 8 | POR | cytochrome p450 oxidoreductase [Source:HGNC Symbol;Acc:HGNC:9208] | 108 | -3.990 | -0.0707 | Yes |
| 9 | PBX1 | PBX homeobox 1 [Source:HGNC Symbol;Acc:HGNC:8632] | 112 | -12.600 | 0.0034 | Yes |
| 10 | BNC2 | basonuclin 2 [Source:HGNC Symbol;Acc:HGNC:30988] | 123 | -42.000 | 0.0172 | Yes |
Table: GSEA details [plain text format]

  

Fig 2: HP\_OLIGOHYDRAMNIOS: Random ES distribution      
 Gene set null distribution of ES for **HP\_OLIGOHYDRAMNIOS**

  
