## Supplemental File 3 for "Increased Expression of *ZFPM2* Bypasses *SRY* to Drive 46,XX Testicular Development: A New Mechanism of 46,XX DSD": HP_THICK_VERMILION_BORDER.html

Details for gene set HP\_THICK\_VERMILION\_BORDER[GSEA]

|  || Dataset | SexDevelopmentGenesPRL\_remapped |
| Phenotype | NoPhenotypeAvailable |
| Upregulated in class | na\_pos |
| GeneSet | HP\_THICK\_VERMILION\_BORDER |
| Enrichment Score (ES) | 0.524138 |
| Normalized Enrichment Score (NES) | 1.9732567 |
| Nominal p-value | 0.0124223605 |
| FDR q-value | 0.82326853 |
| FWER p-Value | 0.973 |
Table: GSEA Results Summary

  

Fig 1: Enrichment plot: HP\_THICK\_VERMILION\_BORDER      
 Profile of the Running ES Score & Positions of GeneSet Members on the Rank Ordered List

  

| SYMBOL | TITLE | RANK IN GENE LIST | RANK METRIC SCORE | RUNNING ES | CORE ENRICHMENT || 1 | FLNA | filamin A [Source:HGNC Symbol;Acc:HGNC:3754] | 3 | 53.800 | 0.0741 | Yes |
| 2 | ZEB2 | zinc finger E-box binding homeobox 2 [Source:HGNC Symbol;Acc:HGNC:14881] | 5 | 36.500 | 0.1655 | Yes |
| 3 | SOS1 | SOS Ras/Rac guanine nucleotide exchange factor 1 [Source:HGNC Symbol;Acc:HGNC:11187] | 15 | 12.800 | 0.1879 | Yes |
| 4 | TWIST2 | twist family bHLH transcription factor 2 [Source:HGNC Symbol;Acc:HGNC:20670] | 19 | 11.800 | 0.2621 | Yes |
| 5 | ATRX | ATRX chromatin remodeler [Source:HGNC Symbol;Acc:HGNC:886] | 27 | 8.350 | 0.3017 | Yes |
| 6 | MED12 | mediator complex subunit 12 [Source:HGNC Symbol;Acc:HGNC:11957] | 33 | 6.470 | 0.3586 | Yes |
| 7 | TOE1 | "target of EGR1, exonuclease [Source:HGNC Symbol;Acc:HGNC:15954]" | 35 | 5.880 | 0.4500 | Yes |
| 8 | PTPN11 | protein tyrosine phosphatase non-receptor type 11 [Source:HGNC Symbol;Acc:HGNC:9644] | 39 | 4.970 | 0.5241 | Yes |
| 9 | PTDSS1 | phosphatidylserine synthase 1 [Source:HGNC Symbol;Acc:HGNC:9587] | 71 | 0.100 | 0.3569 | No |
| 10 | CUL7 | cullin 7 [Source:HGNC Symbol;Acc:HGNC:21024] | 96 | -0.950 | 0.2500 | No |
Table: GSEA details [plain text format]

  

Fig 2: HP\_THICK\_VERMILION\_BORDER: Random ES distribution      
 Gene set null distribution of ES for **HP\_THICK\_VERMILION\_BORDER**

  
