## Supplemental File 3 for "Increased Expression of *ZFPM2* Bypasses *SRY* to Drive 46,XX Testicular Development: A New Mechanism of 46,XX DSD": HP_UPPER_MOTOR_NEURON_DYSFUNCTION.html

Details for gene set HP\_UPPER\_MOTOR\_NEURON\_DYSFUNCTION[GSEA]

|  || Dataset | SexDevelopmentGenesPRL\_remapped |
| Phenotype | NoPhenotypeAvailable |
| Upregulated in class | na\_pos |
| GeneSet | HP\_UPPER\_MOTOR\_NEURON\_DYSFUNCTION |
| Enrichment Score (ES) | 0.3322772 |
| Normalized Enrichment Score (NES) | 1.790156 |
| Nominal p-value | 0.010309278 |
| FDR q-value | 1.0 |
| FWER p-Value | 1.0 |
Table: GSEA Results Summary

  

Fig 1: Enrichment plot: HP\_UPPER\_MOTOR\_NEURON\_DYSFUNCTION      
 Profile of the Running ES Score & Positions of GeneSet Members on the Rank Ordered List

  

| SYMBOL | TITLE | RANK IN GENE LIST | RANK METRIC SCORE | RUNNING ES | CORE ENRICHMENT || 1 | FLNA | filamin A [Source:HGNC Symbol;Acc:HGNC:3754] | 3 | 53.800 | 0.0103 | Yes |
| 2 | DHCR24 | 24-dehydrocholesterol reductase [Source:HGNC Symbol;Acc:HGNC:2859] | 4 | 47.700 | 0.0503 | Yes |
| 3 | ZEB2 | zinc finger E-box binding homeobox 2 [Source:HGNC Symbol;Acc:HGNC:14881] | 5 | 36.500 | 0.0903 | Yes |
| 4 | ANOS1 | anosmin 1 [Source:HGNC Symbol;Acc:HGNC:6211] | 7 | 25.200 | 0.1204 | Yes |
| 5 | OPHN1 | oligophrenin 1 [Source:HGNC Symbol;Acc:HGNC:8148] | 11 | 15.900 | 0.1307 | Yes |
| 6 | WDR11 | WD repeat domain 11 [Source:HGNC Symbol;Acc:HGNC:13831] | 12 | 15.800 | 0.1707 | Yes |
| 7 | HSD17B4 | hydroxysteroid 17-beta dehydrogenase 4 [Source:HGNC Symbol;Acc:HGNC:5213] | 13 | 15.000 | 0.2107 | Yes |
| 8 | SETBP1 | SET binding protein 1 [Source:HGNC Symbol;Acc:HGNC:15573] | 14 | 14.100 | 0.2507 | Yes |
| 9 | HS6ST1 | heparan sulfate 6-O-sulfotransferase 1 [Source:HGNC Symbol;Acc:HGNC:5201] | 24 | 9.710 | 0.2016 | Yes |
| 10 | BCOR | BCL6 corepressor [Source:HGNC Symbol;Acc:HGNC:20893] | 25 | 9.140 | 0.2416 | Yes |
| 11 | ATRX | ATRX chromatin remodeler [Source:HGNC Symbol;Acc:HGNC:886] | 27 | 8.350 | 0.2717 | Yes |
| 12 | MED12 | mediator complex subunit 12 [Source:HGNC Symbol;Acc:HGNC:11957] | 33 | 6.470 | 0.2622 | Yes |
| 13 | TOE1 | "target of EGR1, exonuclease [Source:HGNC Symbol;Acc:HGNC:15954]" | 35 | 5.880 | 0.2923 | Yes |
| 14 | CYB5A | cytochrome b5 type A [Source:HGNC Symbol;Acc:HGNC:2570] | 36 | 5.850 | 0.3323 | Yes |
| 15 | HESX1 | HESX homeobox 1 [Source:HGNC Symbol;Acc:HGNC:4877] | 51 | 1.570 | 0.2337 | No |
| 16 | CKAP2L | cytoskeleton associated protein 2 like [Source:HGNC Symbol;Acc:HGNC:26877] | 64 | 0.460 | 0.1549 | No |
| 17 | FIG4 | FIG4 phosphoinositide 5-phosphatase [Source:HGNC Symbol;Acc:HGNC:16873] | 65 | 0.420 | 0.1949 | No |
| 18 | SEMA3A | semaphorin 3A [Source:HGNC Symbol;Acc:HGNC:10723] | 67 | 0.310 | 0.2250 | No |
| 19 | NEK1 | NIMA related kinase 1 [Source:HGNC Symbol;Acc:HGNC:7744] | 75 | -0.090 | 0.1956 | No |
| 20 | PEX1 | peroxisomal biogenesis factor 1 [Source:HGNC Symbol;Acc:HGNC:8850] | 83 | -0.330 | 0.1663 | No |
| 21 | FGFR1 | fibroblast growth factor receptor 1 [Source:HGNC Symbol;Acc:HGNC:3688] | 90 | -0.570 | 0.1469 | No |
| 22 | IL17RD | interleukin 17 receptor D [Source:HGNC Symbol;Acc:HGNC:17616] | 98 | -1.150 | 0.1176 | No |
| 23 | CHD7 | chromodomain helicase DNA binding protein 7 [Source:HGNC Symbol;Acc:HGNC:20626] | 105 | -3.230 | 0.0982 | No |
| 24 | WWOX | WW domain containing oxidoreductase [Source:HGNC Symbol;Acc:HGNC:12799] | 113 | -14.500 | 0.0689 | No |
| 25 | STAR | steroidogenic acute regulatory protein [Source:HGNC Symbol;Acc:HGNC:11359] | 125 | -56.800 | -0.0000 | No |
Table: GSEA details [plain text format]

  

Fig 2: HP\_UPPER\_MOTOR\_NEURON\_DYSFUNCTION: Random ES distribution      
 Gene set null distribution of ES for **HP\_UPPER\_MOTOR\_NEURON\_DYSFUNCTION**

  
