## Supplemental File 3 for "Increased Expression of *ZFPM2* Bypasses *SRY* to Drive 46,XX Testicular Development: A New Mechanism of 46,XX DSD": HP_VERTEBRAL_SEGMENTATION_DEFECT.html

Details for gene set HP\_VERTEBRAL\_SEGMENTATION\_DEFECT[GSEA]

|  || Dataset | SexDevelopmentGenesPRL\_remapped |
| Phenotype | NoPhenotypeAvailable |
| Upregulated in class | na\_neg |
| GeneSet | HP\_VERTEBRAL\_SEGMENTATION\_DEFECT |
| Enrichment Score (ES) | -0.45336962 |
| Normalized Enrichment Score (NES) | -1.8642881 |
| Nominal p-value | 0.01705757 |
| FDR q-value | 0.5388096 |
| FWER p-Value | 0.996 |
Table: GSEA Results Summary

  

Fig 1: Enrichment plot: HP\_VERTEBRAL\_SEGMENTATION\_DEFECT      
 Profile of the Running ES Score & Positions of GeneSet Members on the Rank Ordered List

  

| SYMBOL | TITLE | RANK IN GENE LIST | RANK METRIC SCORE | RUNNING ES | CORE ENRICHMENT || 1 | FLNA | filamin A [Source:HGNC Symbol;Acc:HGNC:3754] | 3 | 53.800 | 0.0504 | No |
| 2 | ATRX | ATRX chromatin remodeler [Source:HGNC Symbol;Acc:HGNC:886] | 27 | 8.350 | -0.0762 | No |
| 3 | FGFR2 | fibroblast growth factor receptor 2 [Source:HGNC Symbol;Acc:HGNC:3689] | 60 | 0.760 | -0.2825 | No |
| 4 | FRAS1 | Fraser extracellular matrix complex subunit 1 [Source:HGNC Symbol;Acc:HGNC:19185] | 73 | 0.010 | -0.3118 | No |
| 5 | FGFR1 | fibroblast growth factor receptor 1 [Source:HGNC Symbol;Acc:HGNC:3688] | 90 | -0.570 | -0.3764 | Yes |
| 6 | ROR2 | receptor tyrosine kinase like orphan receptor 2 [Source:HGNC Symbol;Acc:HGNC:10257] | 91 | -0.580 | -0.2995 | Yes |
| 7 | GRIP1 | glutamate receptor interacting protein 1 [Source:HGNC Symbol;Acc:HGNC:18708] | 94 | -0.750 | -0.2403 | Yes |
| 8 | GLI3 | GLI family zinc finger 3 [Source:HGNC Symbol;Acc:HGNC:4319] | 104 | -2.920 | -0.2430 | Yes |
| 9 | CHD7 | chromodomain helicase DNA binding protein 7 [Source:HGNC Symbol;Acc:HGNC:20626] | 105 | -3.230 | -0.1661 | Yes |
| 10 | POR | cytochrome p450 oxidoreductase [Source:HGNC Symbol;Acc:HGNC:9208] | 108 | -3.990 | -0.1069 | Yes |
| 11 | WNT5A | Wnt family member 5A [Source:HGNC Symbol;Acc:HGNC:12784] | 115 | -15.700 | -0.0830 | Yes |
| 12 | FREM2 | FRAS1 related extracellular matrix 2 [Source:HGNC Symbol;Acc:HGNC:25396] | 117 | -21.900 | -0.0150 | Yes |
| 13 | B3GLCT | beta 3-glucosyltransferase [Source:HGNC Symbol;Acc:HGNC:20207] | 118 | -22.200 | 0.0619 | Yes |
Table: GSEA details [plain text format]

  

Fig 2: HP\_VERTEBRAL\_SEGMENTATION\_DEFECT: Random ES distribution      
 Gene set null distribution of ES for **HP\_VERTEBRAL\_SEGMENTATION\_DEFECT**

  
