## Supplemental File 3 for "Increased Expression of *ZFPM2* Bypasses *SRY* to Drive 46,XX Testicular Development: A New Mechanism of 46,XX DSD": index.html

Index for xtools.gsea.GseaPreranked my\_analysis.GseaPreranked.1653344355978

### GSEA Report for Dataset SexDevelopmentGenesPRL

#### Enrichment in phenotype: **na**

- 498 / 1033 gene sets are upregulated in phenotype **na\_pos**- 1 gene sets are significant at FDR < 25%- 5 gene sets are significantly enriched at nominal pvalue < 1%- 26 gene sets are significantly enriched at nominal pvalue < 5%- Snapshot of enrichment results- Detailed enrichment results in html format- Detailed enrichment results in TSV format (tab delimited text)- Guide to interpret results

#### Enrichment in phenotype: **na**

- 535 / 1033 gene sets are upregulated in phenotype **na\_neg**- 0 gene sets are significantly enriched at FDR < 25%- 10 gene sets are significantly enriched at nominal pvalue < 1%- 54 gene sets are significantly enriched at nominal pvalue < 5%- Snapshot of enrichment results- Detailed enrichment results in html format- Detailed enrichment results in TSV format (tab delimited text)- Guide to interpret results

#### Dataset details

- The dataset has 126 native features- After collapsing features into gene symbols, there are: 126 genes

#### Gene set details

- Gene set size filters (min=10, max=500) resulted in filtering out 31847 / 32880 gene sets- The remaining 1033 gene sets were used in the analysis- List of gene sets used and their sizes (restricted to features in the specified dataset)

#### Gene markers for the **na\_pos** *versus* **na\_neg** comparison

- The dataset has 126 features (genes)- Detailed rank ordered gene list for all features in the dataset

#### Global statistics and plots

- Plot of p-values *vs.* NES- Global ES histogram

#### Other

- Parameters used for this analysis

#### Comments

- Timestamp used as the random seed: 1653344355978

#### Warnings

- Loaded RNK with 126 features. This may be too few for GSEA, which expects data for all expressed genes for a proper analysis.- Collapsed ranked list results in 126 features. This may be too few for GSEA, which expects data for all expressed genes for a proper analysis.

#### Citing GSEA and MSigDB

To cite your use of the GSEA software please reference the following:

- Subramanian, A., Tamayo, P., et al. (2005, PNAS). - Mootha, V. K., Lindgren, C. M., et al. (2003, Nature Genetics).

For use of the Molecular Signatures Database (MSigDB), to cite please reference   
one or more of the following as appropriate, along with the source for the gene set as listed on the gene set page:

- Liberzon A, et al. (Bioinformatics, 2011). - Liberzon A, et al. (Cell Systems 2015).

---

Report: my\_analysis.GseaPreranked.1653344355978.rpt   by user: leahragno

xtools.gsea.GseaPreranked [Mon, May 23, '22 6 PM 19]

Website: www.gsea-msigdb.org/gsea
Questions & Suggestions: Contact page
