## Supplemental File 3 for "Increased Expression of *ZFPM2* Bypasses *SRY* to Drive 46,XX Testicular Development: A New Mechanism of 46,XX DSD": NAB2_TARGET_GENES.html

Details for gene set NAB2\_TARGET\_GENES[GSEA]

|  || Dataset | SexDevelopmentGenesPRL\_remapped |
| Phenotype | NoPhenotypeAvailable |
| Upregulated in class | na\_pos |
| GeneSet | NAB2\_TARGET\_GENES |
| Enrichment Score (ES) | 0.52068967 |
| Normalized Enrichment Score (NES) | 1.9444038 |
| Nominal p-value | 0.006185567 |
| FDR q-value | 0.7806952 |
| FWER p-Value | 0.987 |
Table: GSEA Results Summary

  

Fig 1: Enrichment plot: NAB2\_TARGET\_GENES      
 Profile of the Running ES Score & Positions of GeneSet Members on the Rank Ordered List

  

| SYMBOL | TITLE | RANK IN GENE LIST | RANK METRIC SCORE | RUNNING ES | CORE ENRICHMENT || 1 | MAP3K1 | mitogen-activated protein kinase kinase kinase 1 [Source:HGNC Symbol;Acc:HGNC:6848] | 10 | 17.600 | 0.0138 | Yes |
| 2 | SOS1 | SOS Ras/Rac guanine nucleotide exchange factor 1 [Source:HGNC Symbol;Acc:HGNC:11187] | 15 | 12.800 | 0.0793 | Yes |
| 3 | PCNT | pericentrin [Source:HGNC Symbol;Acc:HGNC:16068] | 17 | 12.400 | 0.1707 | Yes |
| 4 | H6PD | hexose-6-phosphate dehydrogenase/glucose 1-dehydrogenase [Source:HGNC Symbol;Acc:HGNC:4795] | 21 | 11.600 | 0.2448 | Yes |
| 5 | HS6ST1 | heparan sulfate 6-O-sulfotransferase 1 [Source:HGNC Symbol;Acc:HGNC:5201] | 24 | 9.710 | 0.3276 | Yes |
| 6 | NR3C1 | nuclear receptor subfamily 3 group C member 1 [Source:HGNC Symbol;Acc:HGNC:7978] | 29 | 7.690 | 0.3931 | Yes |
| 7 | PTPN11 | protein tyrosine phosphatase non-receptor type 11 [Source:HGNC Symbol;Acc:HGNC:9644] | 39 | 4.970 | 0.4155 | Yes |
| 8 | DNMT3B | DNA methyltransferase 3 beta [Source:HGNC Symbol;Acc:HGNC:2979] | 45 | 2.110 | 0.4724 | Yes |
| 9 | LMNA | lamin A/C [Source:HGNC Symbol;Acc:HGNC:6636] | 52 | 1.560 | 0.5207 | Yes |
| 10 | CBX2 | chromobox 2 [Source:HGNC Symbol;Acc:HGNC:1552] | 87 | -0.410 | 0.3276 | No |
Table: GSEA details [plain text format]

  

Fig 2: NAB2\_TARGET\_GENES: Random ES distribution      
 Gene set null distribution of ES for **NAB2\_TARGET\_GENES**

  
