## Supplemental File 3 for "Increased Expression of *ZFPM2* Bypasses *SRY* to Drive 46,XX Testicular Development: A New Mechanism of 46,XX DSD": RYTTCCTG_ETS2_B.html

Details for gene set RYTTCCTG\_ETS2\_B[GSEA]

|  || Dataset | SexDevelopmentGenesPRL\_remapped |
| Phenotype | NoPhenotypeAvailable |
| Upregulated in class | na\_pos |
| GeneSet | RYTTCCTG\_ETS2\_B |
| Enrichment Score (ES) | 0.4166667 |
| Normalized Enrichment Score (NES) | 1.6693673 |
| Nominal p-value | 0.032786883 |
| FDR q-value | 0.87744224 |
| FWER p-Value | 1.0 |
Table: GSEA Results Summary

  

Fig 1: Enrichment plot: RYTTCCTG\_ETS2\_B      
 Profile of the Running ES Score & Positions of GeneSet Members on the Rank Ordered List

  

| SYMBOL | TITLE | RANK IN GENE LIST | RANK METRIC SCORE | RUNNING ES | CORE ENRICHMENT || 1 | DHCR24 | 24-dehydrocholesterol reductase [Source:HGNC Symbol;Acc:HGNC:2859] | 4 | 47.700 | 0.0482 | Yes |
| 2 | ZEB2 | zinc finger E-box binding homeobox 2 [Source:HGNC Symbol;Acc:HGNC:14881] | 5 | 36.500 | 0.1316 | Yes |
| 3 | PDE4D | phosphodiesterase 4D [Source:HGNC Symbol;Acc:HGNC:8783] | 9 | 18.600 | 0.1886 | Yes |
| 4 | WDR11 | WD repeat domain 11 [Source:HGNC Symbol;Acc:HGNC:13831] | 12 | 15.800 | 0.2544 | Yes |
| 5 | MID1 | midline 1 [Source:HGNC Symbol;Acc:HGNC:7095] | 26 | 8.770 | 0.2237 | Yes |
| 6 | PTPN11 | protein tyrosine phosphatase non-receptor type 11 [Source:HGNC Symbol;Acc:HGNC:9644] | 39 | 4.970 | 0.2018 | Yes |
| 7 | HOXA10 | homeobox A10 [Source:HGNC Symbol;Acc:HGNC:5100] | 41 | 3.830 | 0.2763 | Yes |
| 8 | HFE | homeostatic iron regulator [Source:HGNC Symbol;Acc:HGNC:4886] | 43 | 2.890 | 0.3509 | Yes |
| 9 | HOXB6 | homeobox B6 [Source:HGNC Symbol;Acc:HGNC:5117] | 46 | 2.100 | 0.4167 | Yes |
| 10 | FGFR2 | fibroblast growth factor receptor 2 [Source:HGNC Symbol;Acc:HGNC:3689] | 60 | 0.760 | 0.3860 | No |
| 11 | HOXA4 | homeobox A4 [Source:HGNC Symbol;Acc:HGNC:5105] | 85 | -0.350 | 0.2588 | No |
| 12 | PITX2 | paired like homeodomain 2 [Source:HGNC Symbol;Acc:HGNC:9005] | 107 | -3.730 | 0.1579 | No |
Table: GSEA details [plain text format]

  

Fig 2: RYTTCCTG\_ETS2\_B: Random ES distribution      
 Gene set null distribution of ES for **RYTTCCTG\_ETS2\_B**

  
