## Supplemental File 3 for "Increased Expression of *ZFPM2* Bypasses *SRY* to Drive 46,XX Testicular Development: A New Mechanism of 46,XX DSD": WP_CILIOPATHIES.html

Details for gene set WP\_CILIOPATHIES[GSEA]

|  || Dataset | SexDevelopmentGenesPRL\_remapped |
| Phenotype | NoPhenotypeAvailable |
| Upregulated in class | na\_neg |
| GeneSet | WP\_CILIOPATHIES |
| Enrichment Score (ES) | -0.35566038 |
| Normalized Enrichment Score (NES) | -1.7389268 |
| Nominal p-value | 0.02739726 |
| FDR q-value | 0.5234723 |
| FWER p-Value | 1.0 |
Table: GSEA Results Summary

  

Fig 1: Enrichment plot: WP\_CILIOPATHIES      
 Profile of the Running ES Score & Positions of GeneSet Members on the Rank Ordered List

  

| SYMBOL | TITLE | RANK IN GENE LIST | RANK METRIC SCORE | RUNNING ES | CORE ENRICHMENT || 1 | MKKS | MKKS centrosomal shuttling protein [Source:HGNC Symbol;Acc:HGNC:7108] | 34 | 6.190 | -0.2708 | No |
| 2 | TRIM32 | tripartite motif containing 32 [Source:HGNC Symbol;Acc:HGNC:16380] | 44 | 2.410 | -0.3057 | Yes |
| 3 | BBS9 | Bardet-Biedl syndrome 9 [Source:HGNC Symbol;Acc:HGNC:30000] | 47 | 2.000 | -0.2745 | Yes |
| 4 | BBS2 | Bardet-Biedl syndrome 2 [Source:HGNC Symbol;Acc:HGNC:967] | 48 | 1.960 | -0.2245 | Yes |
| 5 | BBS12 | Bardet-Biedl syndrome 12 [Source:HGNC Symbol;Acc:HGNC:26648] | 49 | 1.960 | -0.1745 | Yes |
| 6 | WDR35 | WD repeat domain 35 [Source:HGNC Symbol;Acc:HGNC:29250] | 54 | 1.420 | -0.1623 | Yes |
| 7 | BBS7 | Bardet-Biedl syndrome 7 [Source:HGNC Symbol;Acc:HGNC:18758] | 66 | 0.400 | -0.2160 | Yes |
| 8 | ARL6 | ADP ribosylation factor like GTPase 6 [Source:HGNC Symbol;Acc:HGNC:13210] | 68 | 0.280 | -0.1755 | Yes |
| 9 | NEK1 | NIMA related kinase 1 [Source:HGNC Symbol;Acc:HGNC:7744] | 75 | -0.090 | -0.1821 | Yes |
| 10 | DYNC2H1 | dynein cytoplasmic 2 heavy chain 1 [Source:HGNC Symbol;Acc:HGNC:2962] | 79 | -0.230 | -0.1604 | Yes |
| 11 | CEP41 | centrosomal protein 41 [Source:HGNC Symbol;Acc:HGNC:12370] | 84 | -0.340 | -0.1481 | Yes |
| 12 | MKS1 | MKS transition zone complex subunit 1 [Source:HGNC Symbol;Acc:HGNC:7121] | 88 | -0.480 | -0.1264 | Yes |
| 13 | TTC8 | tetratricopeptide repeat domain 8 [Source:HGNC Symbol;Acc:HGNC:20087] | 92 | -0.640 | -0.1047 | Yes |
| 14 | EVC2 | EvC ciliary complex subunit 2 [Source:HGNC Symbol;Acc:HGNC:19747] | 95 | -0.890 | -0.0736 | Yes |
| 15 | TCTN3 | tectonic family member 3 [Source:HGNC Symbol;Acc:HGNC:24519] | 97 | -1.040 | -0.0330 | Yes |
| 16 | BBS4 | Bardet-Biedl syndrome 4 [Source:HGNC Symbol;Acc:HGNC:969] | 100 | -1.360 | -0.0019 | Yes |
| 17 | BBS10 | Bardet-Biedl syndrome 10 [Source:HGNC Symbol;Acc:HGNC:26291] | 102 | -1.550 | 0.0387 | Yes |
| 18 | GLI3 | GLI family zinc finger 3 [Source:HGNC Symbol;Acc:HGNC:4319] | 104 | -2.920 | 0.0792 | Yes |
| 19 | BBS5 | Bardet-Biedl syndrome 5 [Source:HGNC Symbol;Acc:HGNC:970] | 114 | -14.600 | 0.0443 | Yes |
| 20 | EVC | EvC ciliary complex subunit 1 [Source:HGNC Symbol;Acc:HGNC:3497] | 119 | -30.300 | 0.0566 | Yes |
Table: GSEA details [plain text format]

  

Fig 2: WP\_CILIOPATHIES: Random ES distribution      
 Gene set null distribution of ES for **WP\_CILIOPATHIES**

  
