## Supplementary figures and images for "Increased Expression of *ZFPM2* Bypasses *SRY* to Drive 46,XX Testicular Development: A New Mechanism of 46,XX DSD"

### enplot_BUSSLINGER_GASTRIC_IMMUNE_CELLS_451.png

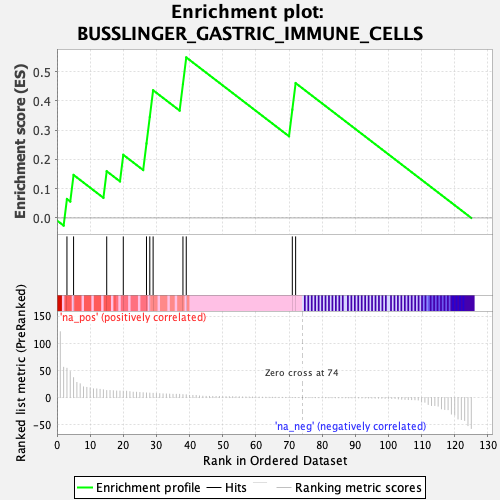

### enplot_DACOSTA_UV_RESPONSE_VIA_ERCC3_DN_449.png

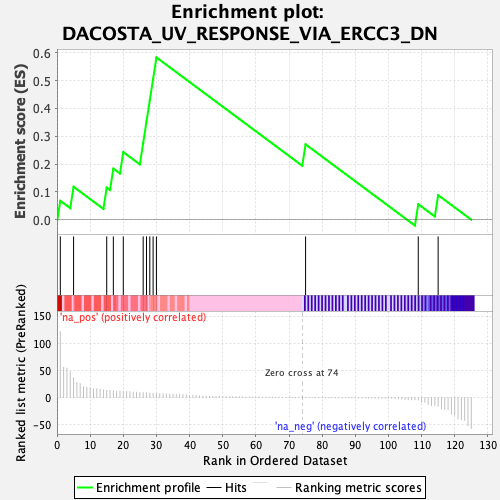

### enplot_GOBP_EMBRYONIC_APPENDAGE_MORPHOGENESIS_549.png

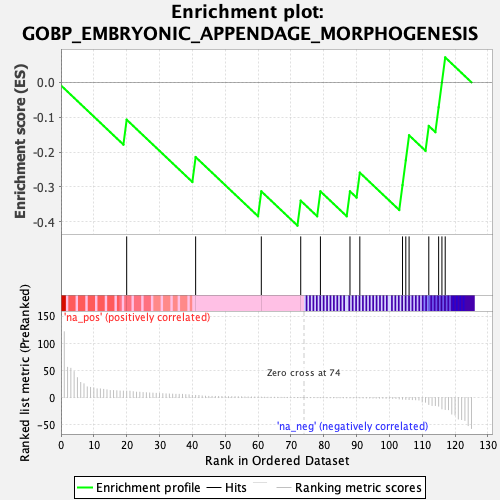

### enplot_GOBP_IMMUNE_RESPONSE_469.png

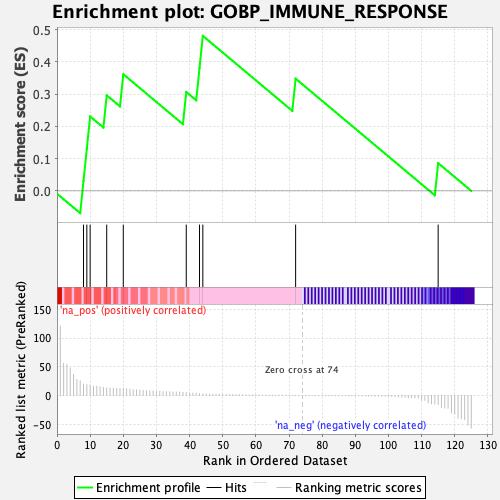

### enplot_GOBP_MALE_SEX_DIFFERENTIATION_461.png

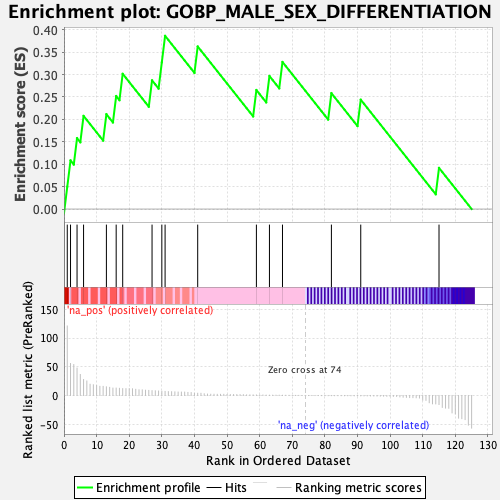

### enplot_GOBP_POSITIVE_REGULATION_OF_GENE_EXPRESSION_487.png

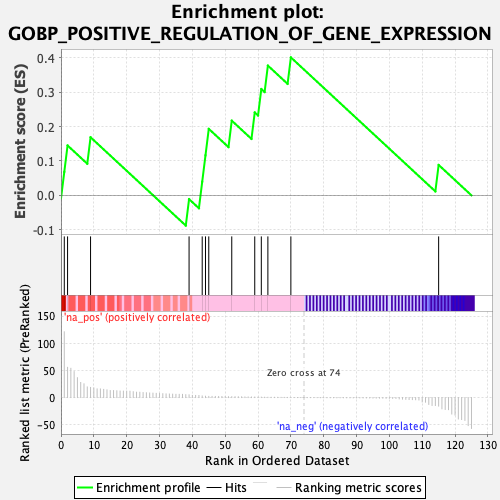

### enplot_GOBP_SKELETAL_SYSTEM_DEVELOPMENT_535.png

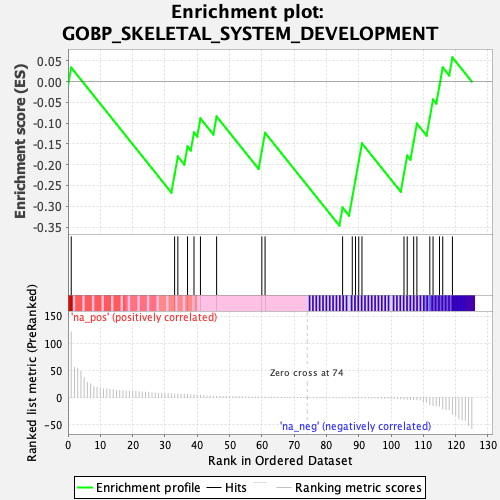

### enplot_GOBP_UROGENITAL_SYSTEM_DEVELOPMENT_525.png

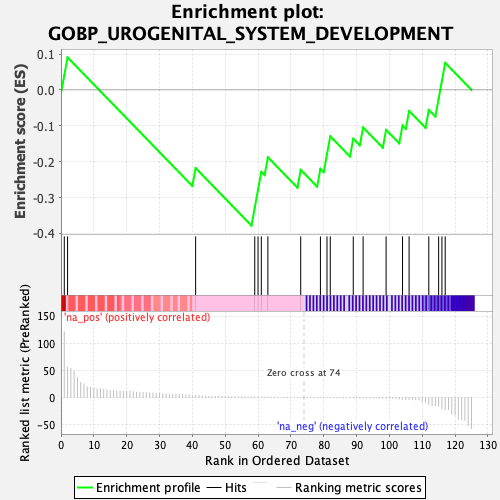

### enplot_GOCC_CATALYTIC_COMPLEX_479.png

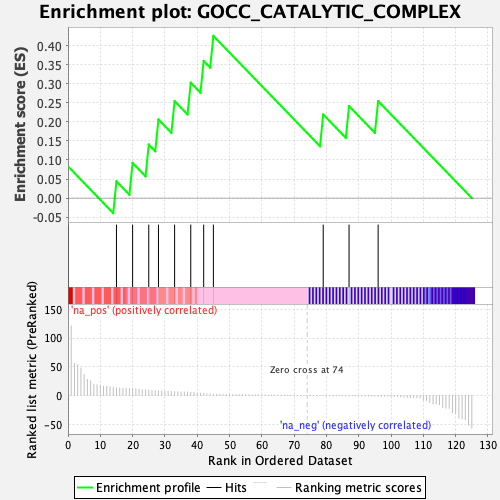

### enplot_GOCC_PLASMA_MEMBRANE_REGION_543.png

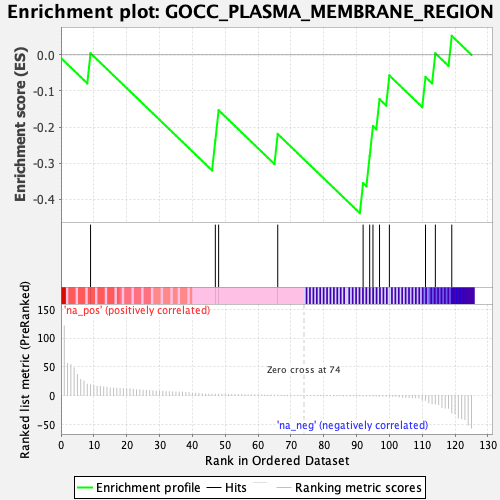

### enplot_GOMF_TRANSITION_METAL_ION_BINDING_477.png

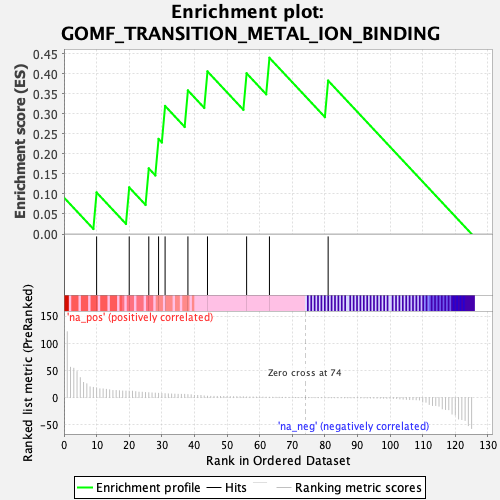

### enplot_HP_ABNORMAL_CEREBELLAR_VERMIS_MORPHOLOGY_545.png

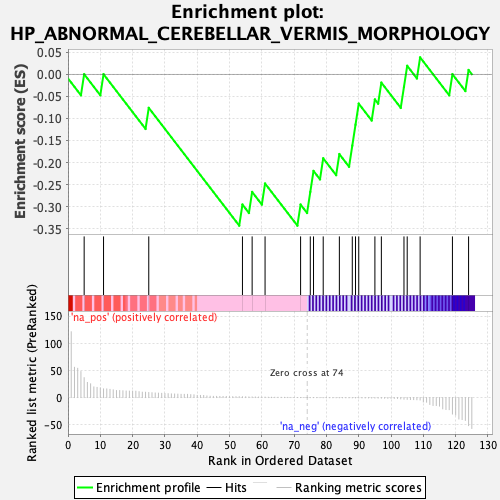

### enplot_HP_ABNORMAL_FINGERNAIL_MORPHOLOGY_533.png

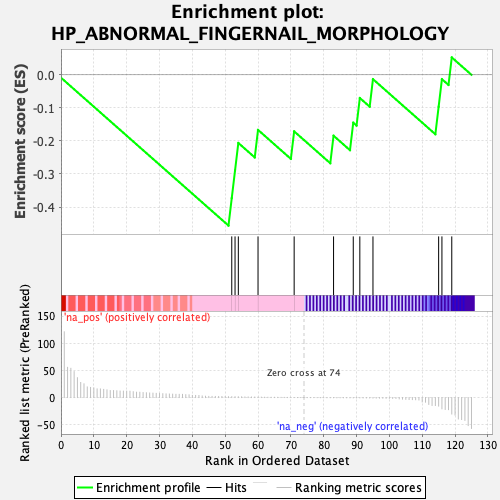

### enplot_HP_ABNORMAL_GROWTH_HORMONE_LEVEL_513.png

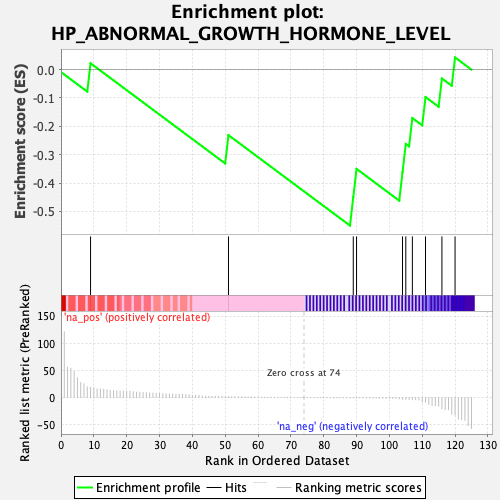

### enplot_HP_ABNORMAL_ILIUM_MORPHOLOGY_493.png

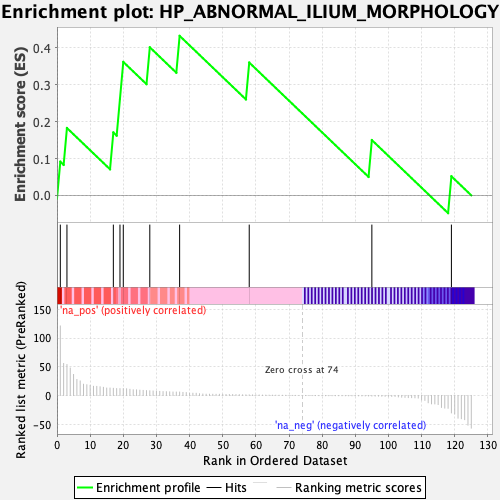

### enplot_HP_ABNORMAL_LOCALIZATION_OF_KIDNEY_539.png

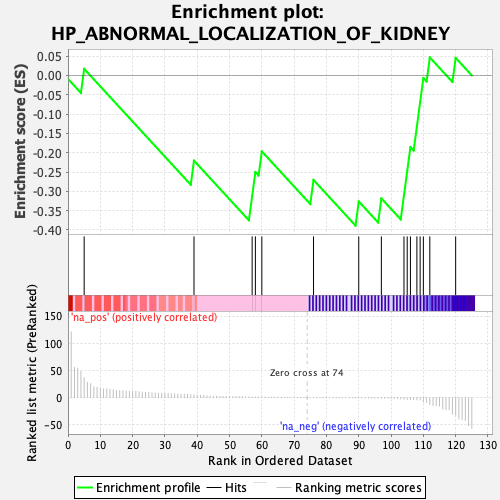

### enplot_HP_ABNORMAL_RETINAL_MORPHOLOGY_527.png

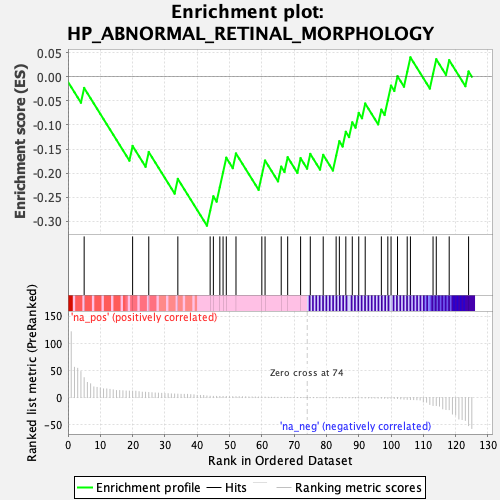

### enplot_HP_ABNORMALITY_OF_GLOBE_SIZE_507.png

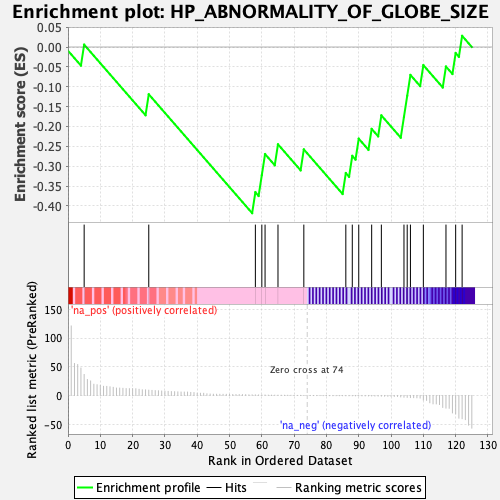

### enplot_HP_ABNORMALITY_OF_HAIR_TEXTURE_459.png

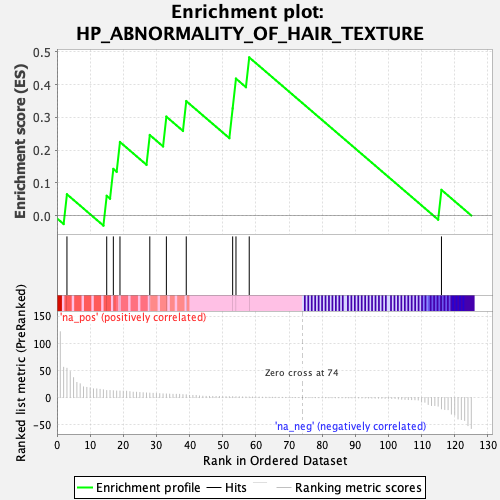

### enplot_HP_ABNORMALITY_OF_THE_BREAST_489.png

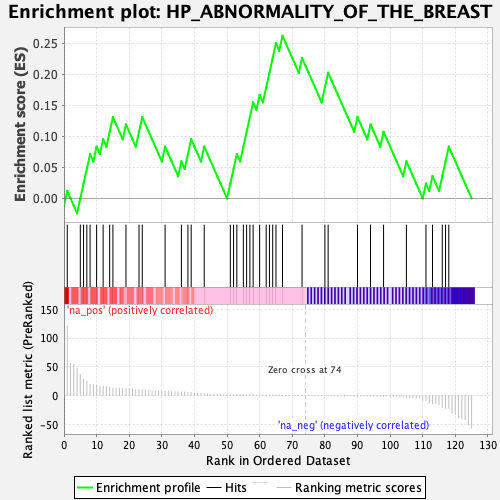

### enplot_HP_APLASIA_HYPOPLASIA_AFFECTING_THE_EYE_517.png

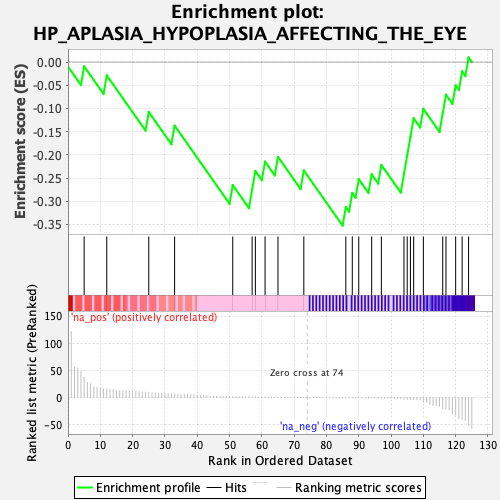

### enplot_HP_APLASIA_HYPOPLASIA_INVOLVING_BONES_OF_THE_UPPER_LIMBS_551.png

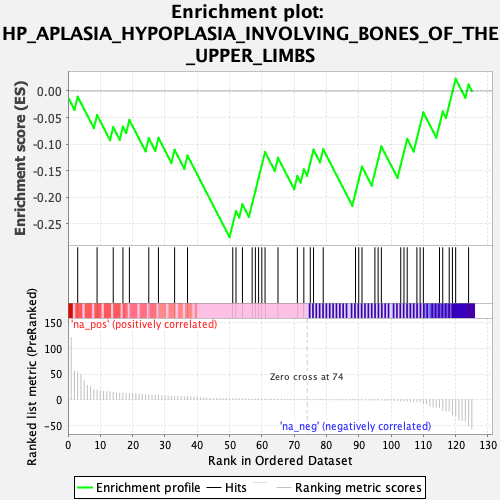

### enplot_HP_APLASIA_HYPOPLASIA_OF_THE_CEREBELLAR_VERMIS_541.png

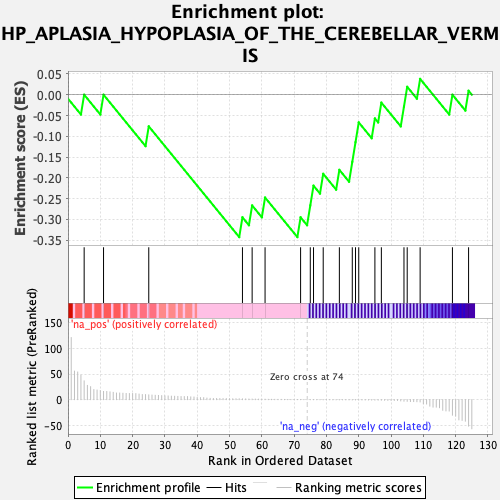

### enplot_HP_APLASIA_HYPOPLASIA_OF_THE_NAILS_515.png

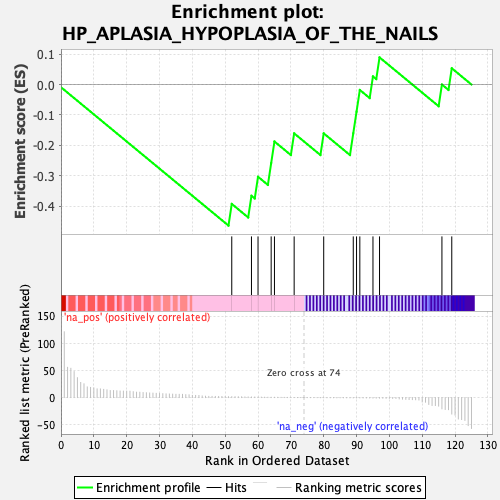

### enplot_HP_CLEFT_LIP_503.png

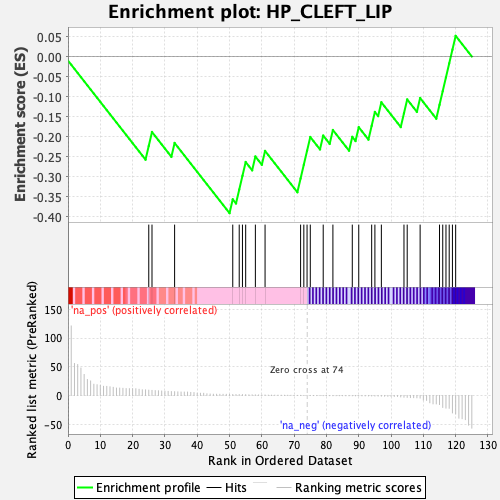

### enplot_HP_CLEFT_UPPER_LIP_505.png

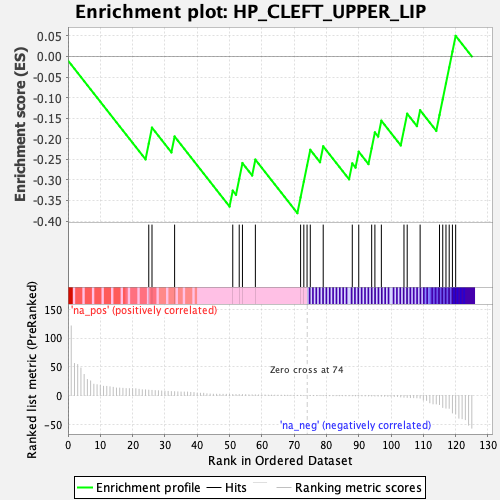

### enplot_HP_DECREASED_FERTILITY_IN_MALES_471.png

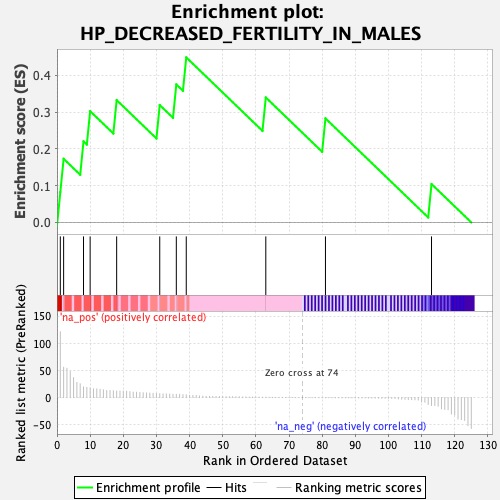

### enplot_HP_DUPLICATION_OF_HAND_BONES_537.png

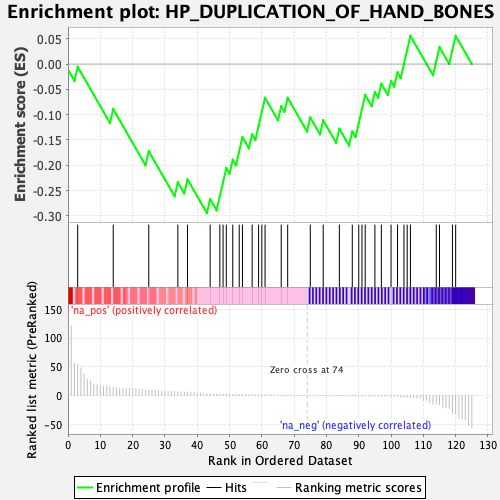

### enplot_HP_FINGER_SYNDACTYLY_501.png

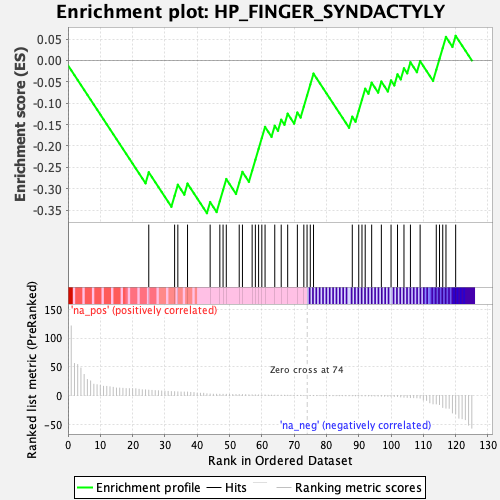

### enplot_HP_GONOSOMAL_INHERITANCE_465.png

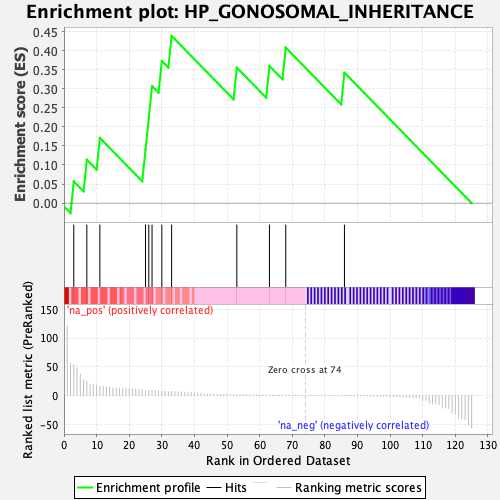

### enplot_HP_HAND_POLYDACTYLY_521.png

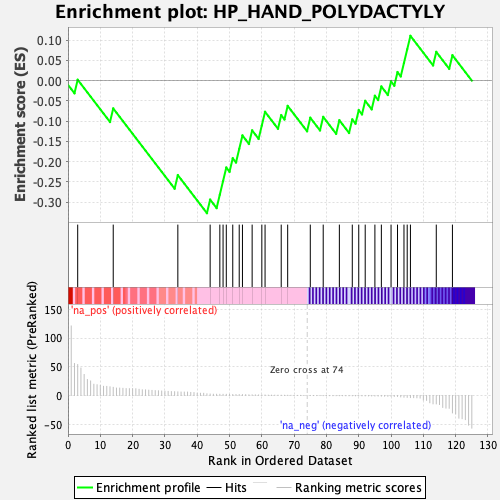

### enplot_HP_HETEROTROPIA_483.png

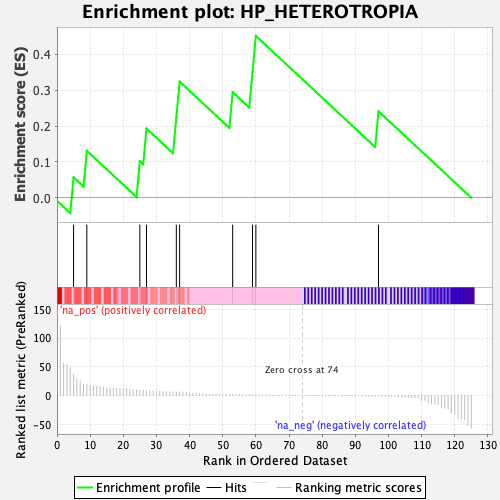

### enplot_HP_HYPERACTIVITY_491.png

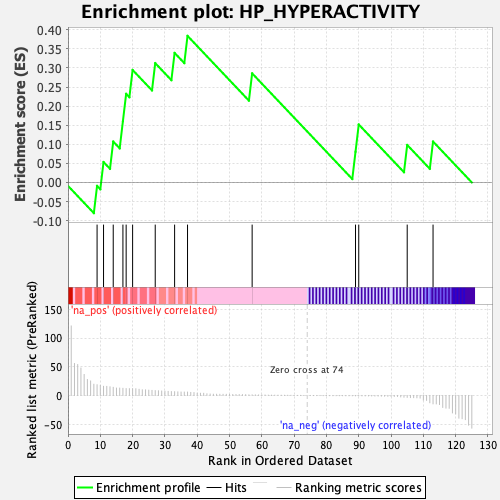

### enplot_HP_HYPOGONADOTROPIC_HYPOGONADISM_499.png

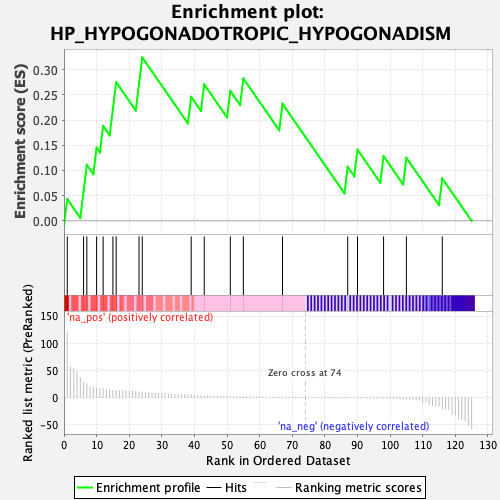

### enplot_HP_INFERTILITY_453.png

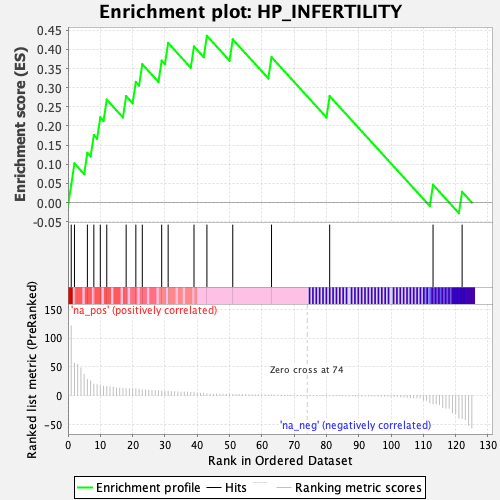

### enplot_HP_INTELLECTUAL_DISABILITY_SEVERE_467.png

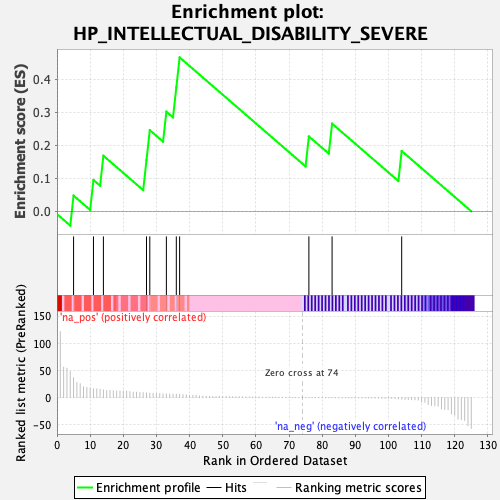

### enplot_HP_SHORT_THUMB_473.png

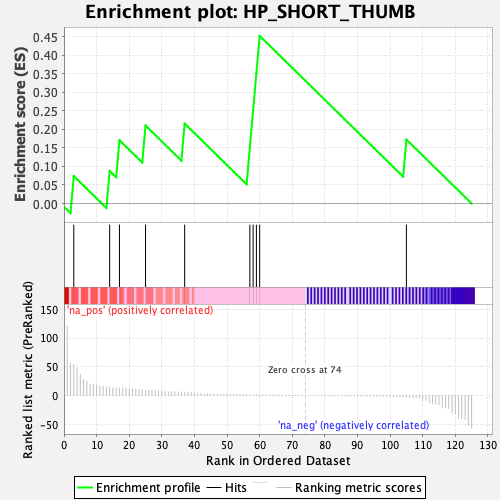

### enplot_HP_SMALL_NAIL_511.png

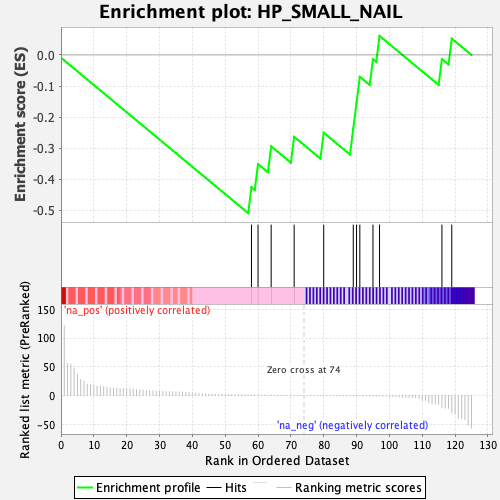

### enplot_HP_SPASTICITY_475.png

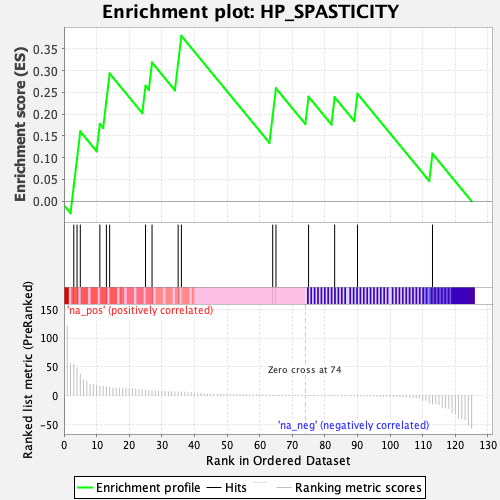

### enplot_HP_UPPER_MOTOR_NEURON_DYSFUNCTION_463.png

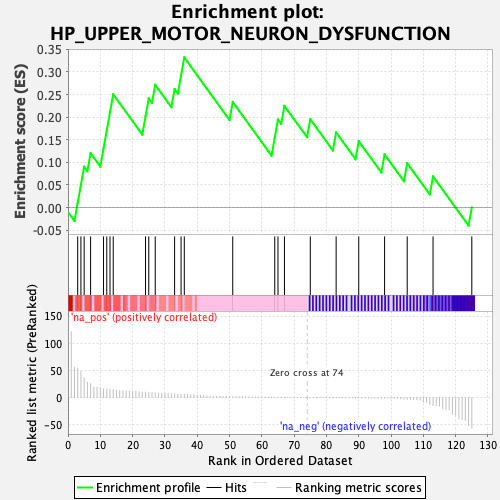

### enplot_HP_VERTEBRAL_SEGMENTATION_DEFECT_523.png

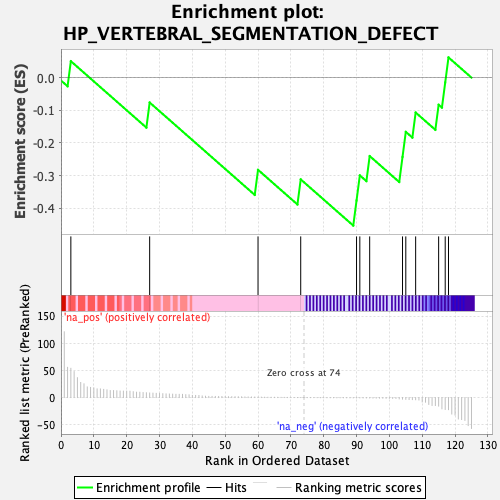

### enplot_RYTTCCTG_ETS2_B_485.png

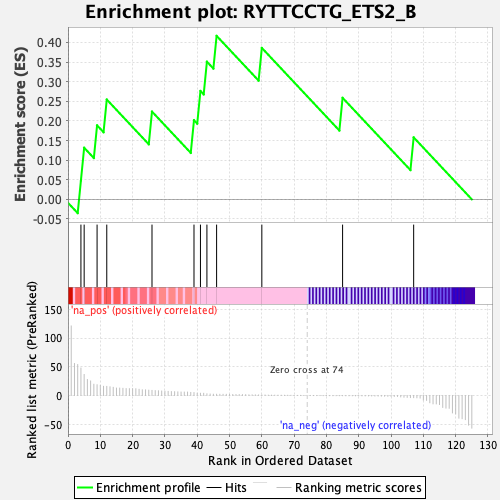

### enplot_WP_CILIOPATHIES_547.png

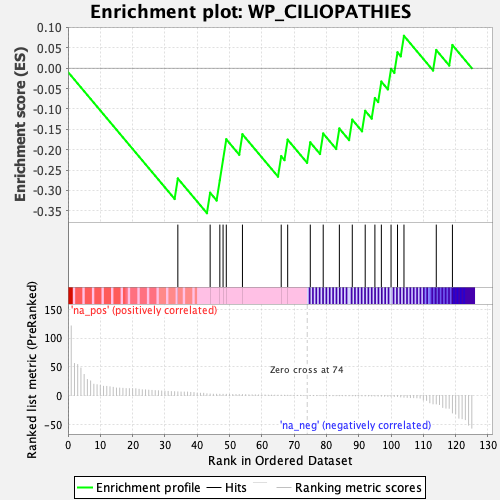

### global_es_histogram.png

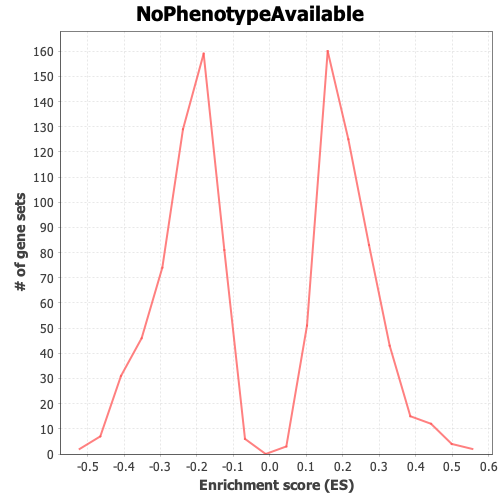

### gset_rnd_es_dist_450.png

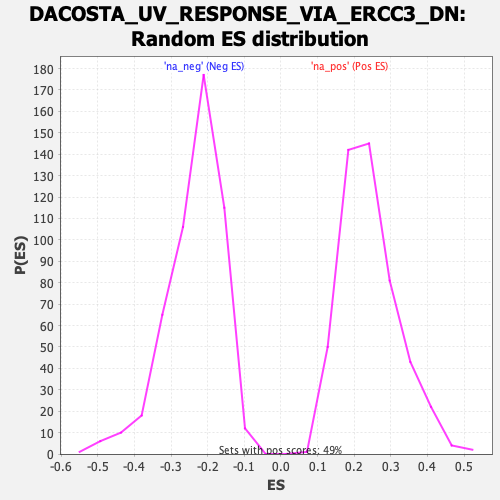

### gset_rnd_es_dist_452.png

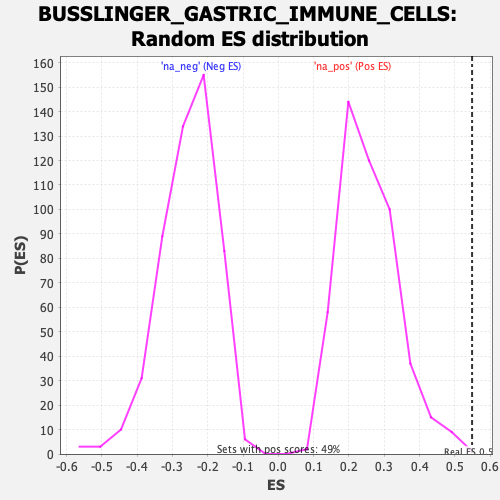

### gset_rnd_es_dist_454.png

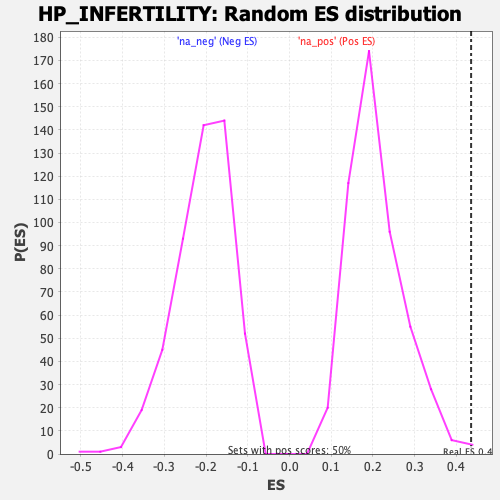

### gset_rnd_es_dist_456.png

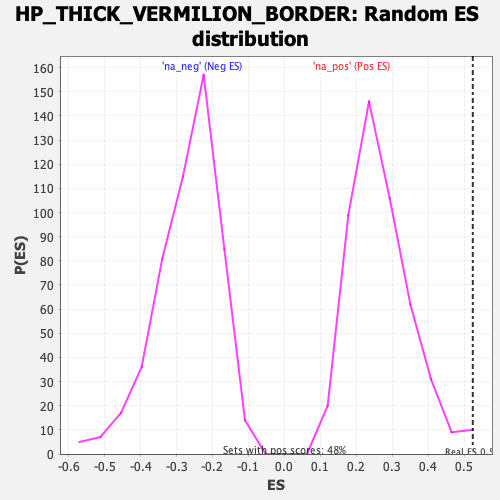

### gset_rnd_es_dist_458.png

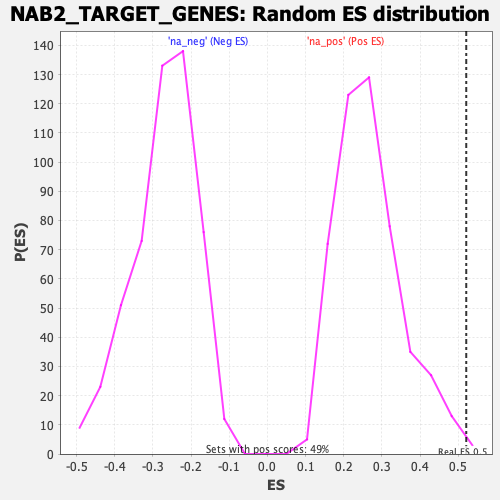

### gset_rnd_es_dist_460.png

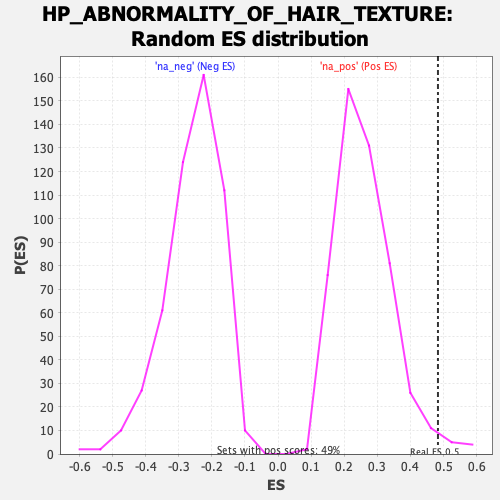

### gset_rnd_es_dist_462.png

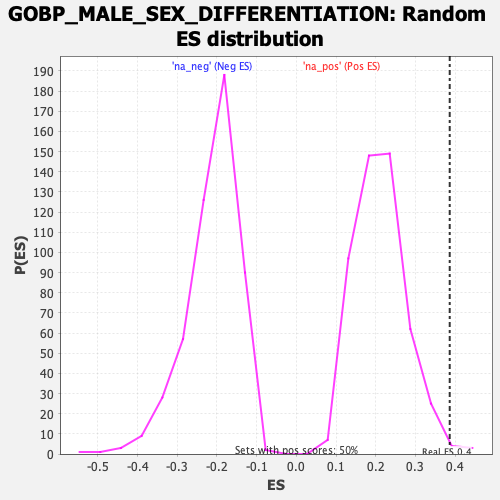

### gset_rnd_es_dist_464.png

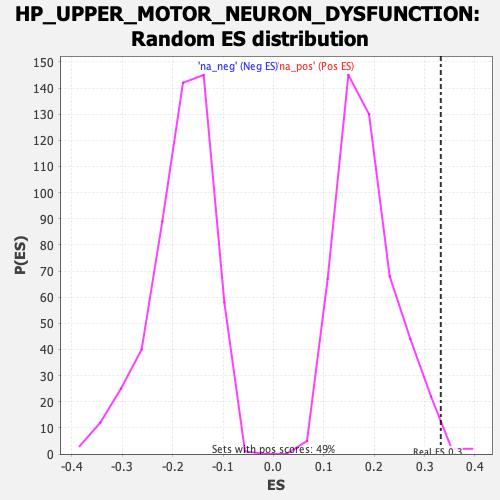

### gset_rnd_es_dist_466.png

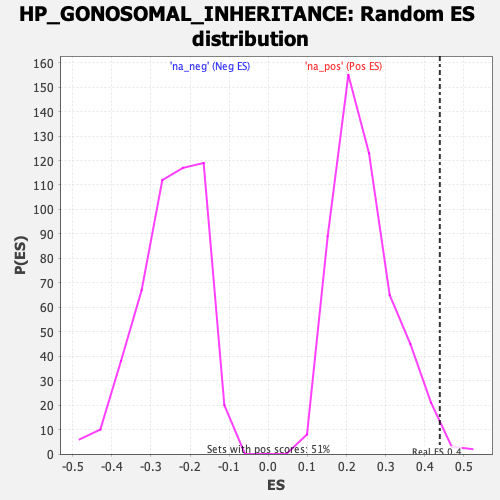

### gset_rnd_es_dist_468.png

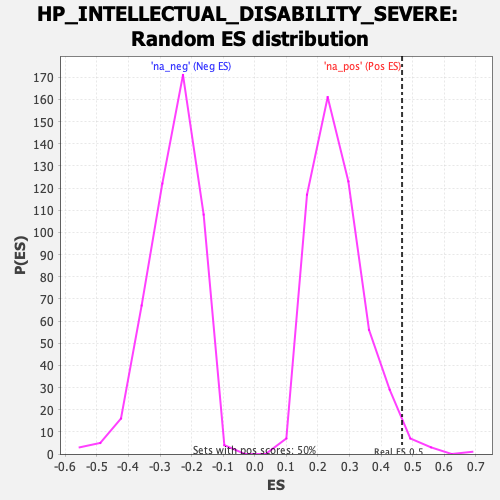

### gset_rnd_es_dist_470.png

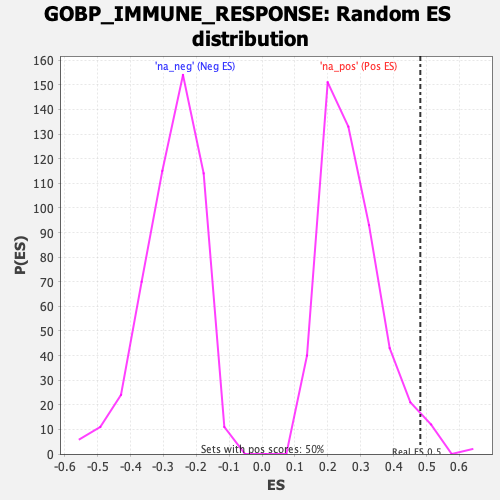

### gset_rnd_es_dist_472.png

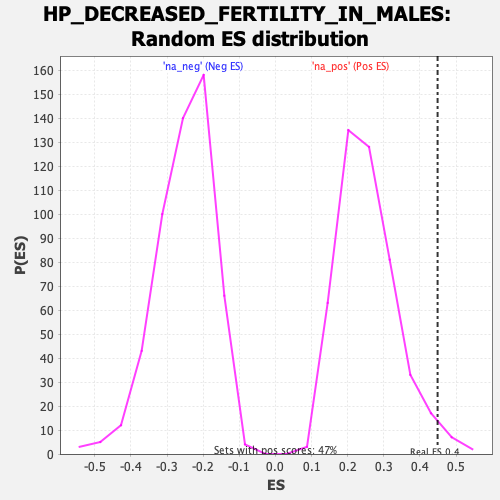

### gset_rnd_es_dist_474.png

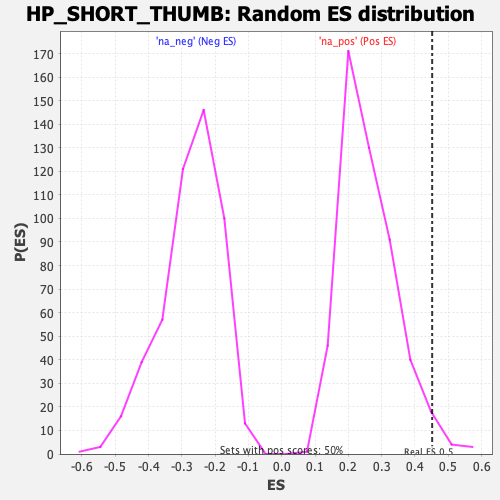

### gset_rnd_es_dist_476.png

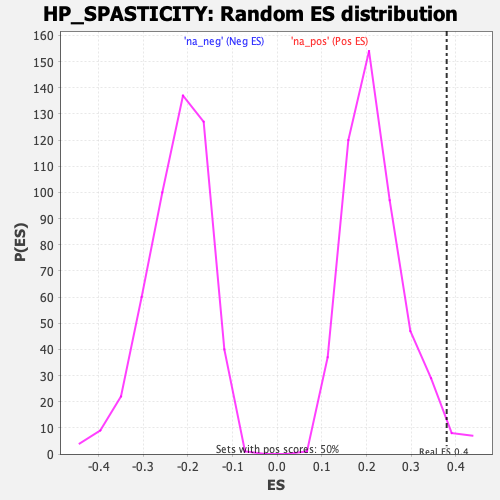

### gset_rnd_es_dist_478.png

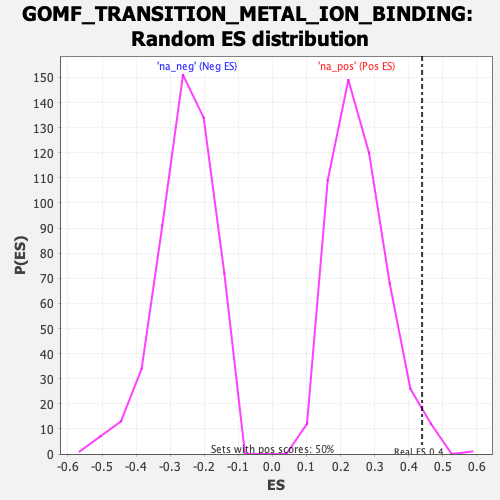

### gset_rnd_es_dist_480.png

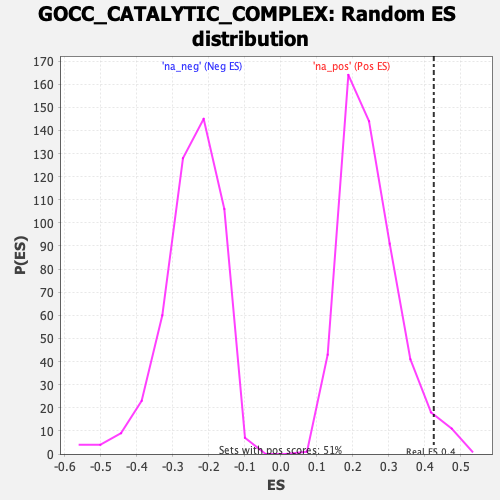

### gset_rnd_es_dist_482.png

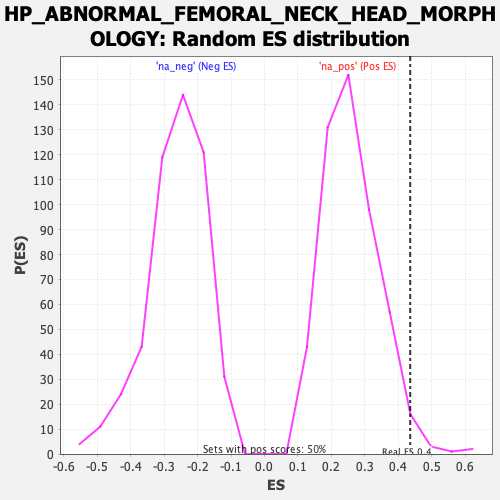

### gset_rnd_es_dist_484.png

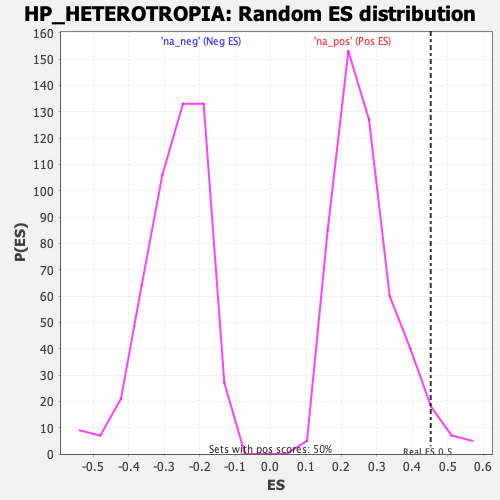

### gset_rnd_es_dist_486.png

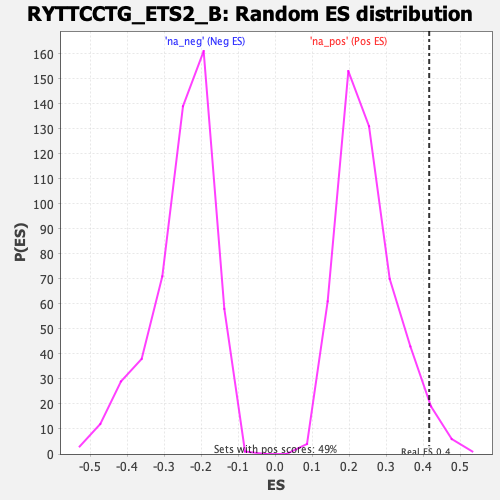

### gset_rnd_es_dist_488.png

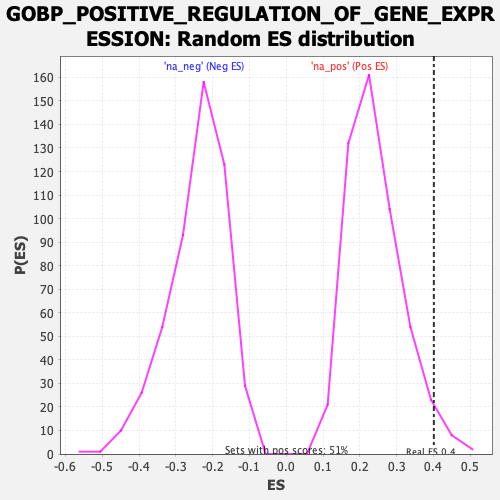

### gset_rnd_es_dist_490.png

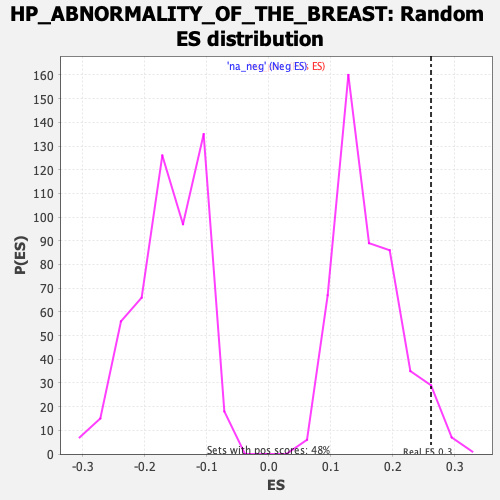

### gset_rnd_es_dist_492.png

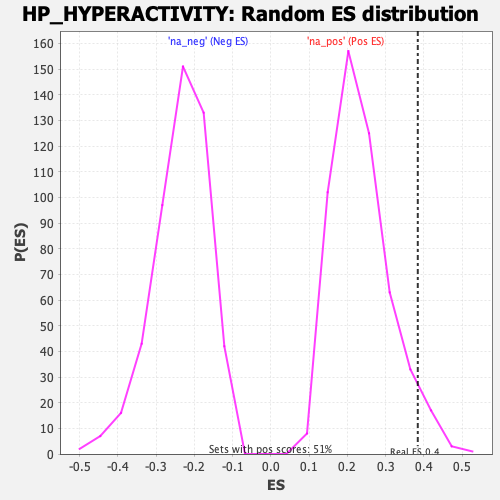

### gset_rnd_es_dist_494.png

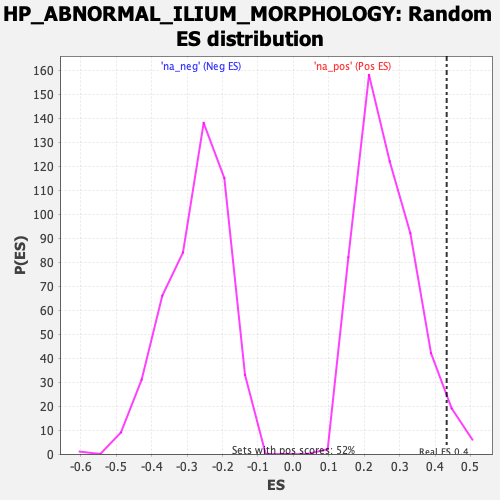

### gset_rnd_es_dist_496.png

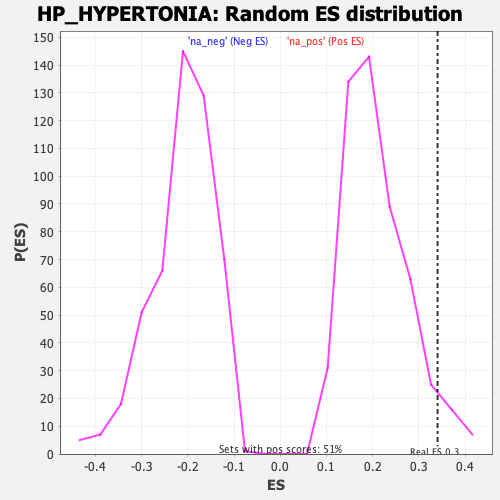

### gset_rnd_es_dist_498.png

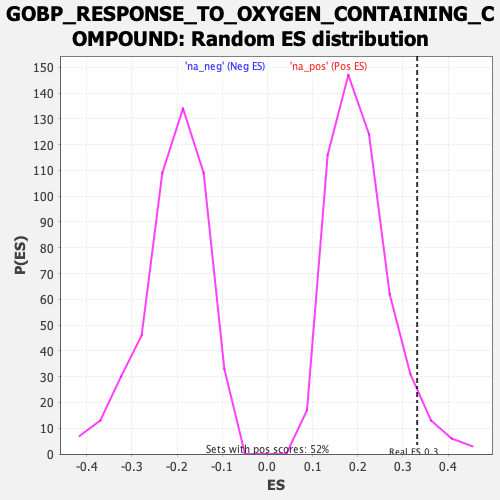

### gset_rnd_es_dist_500.png

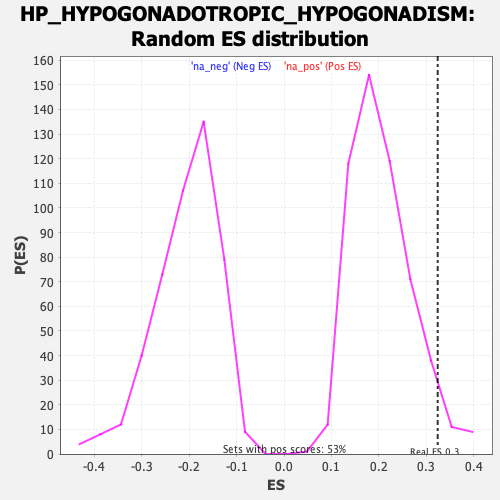

### gset_rnd_es_dist_502.png

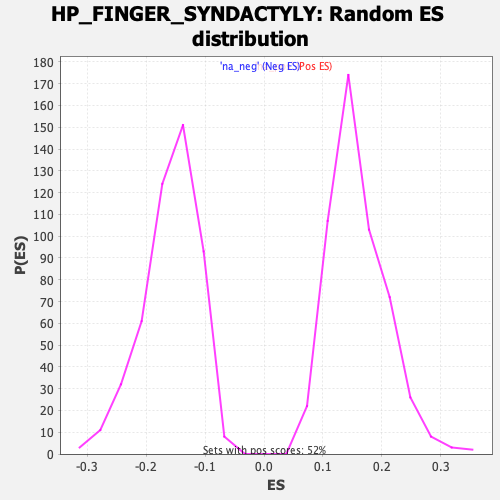

### gset_rnd_es_dist_504.png

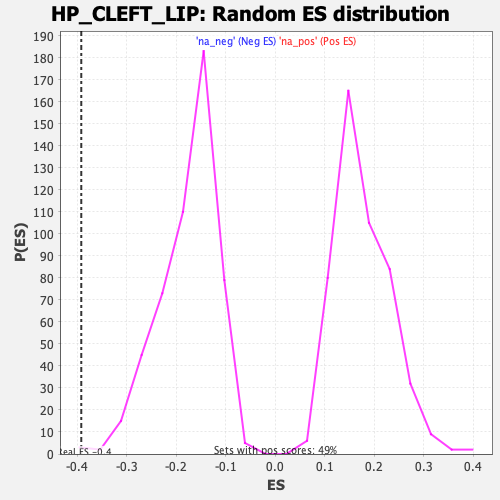

### gset_rnd_es_dist_506.png

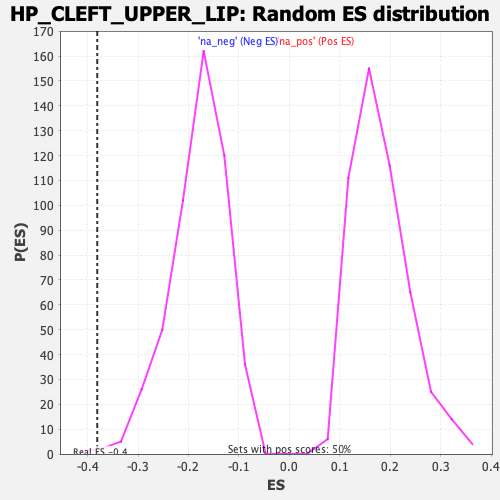

### gset_rnd_es_dist_508.png

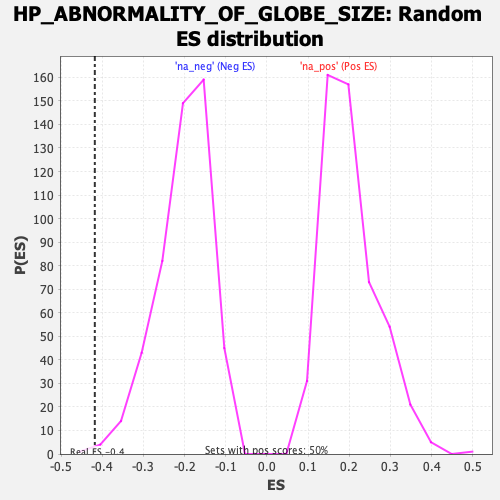

### gset_rnd_es_dist_510.png

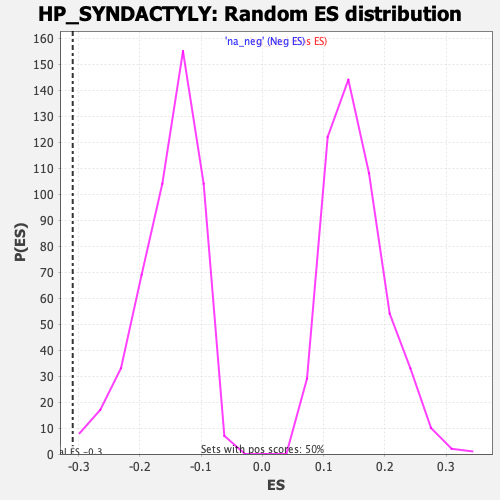

### gset_rnd_es_dist_512.png

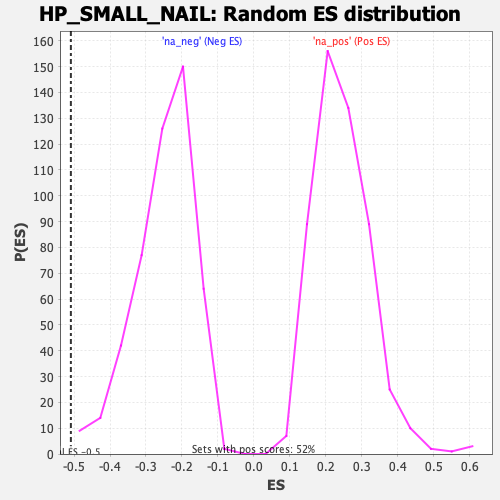

### gset_rnd_es_dist_514.png

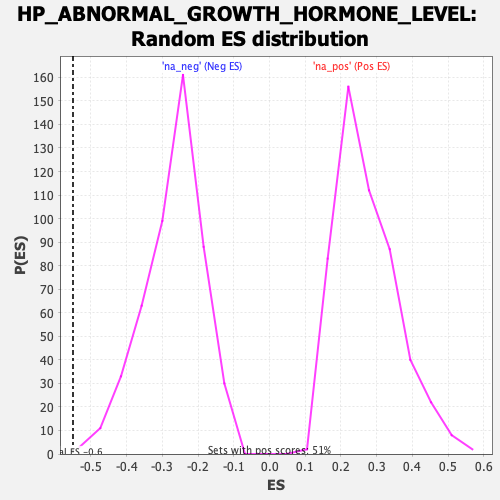

### gset_rnd_es_dist_516.png

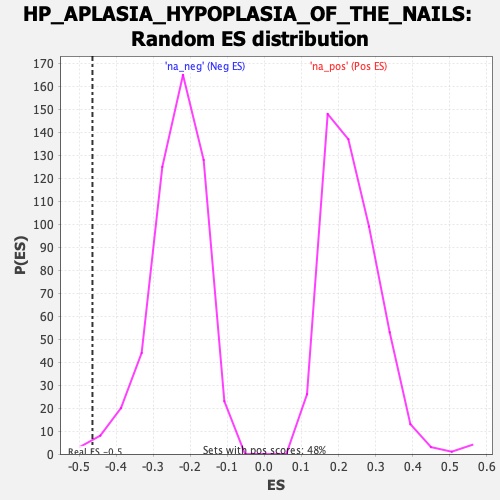

### gset_rnd_es_dist_518.png

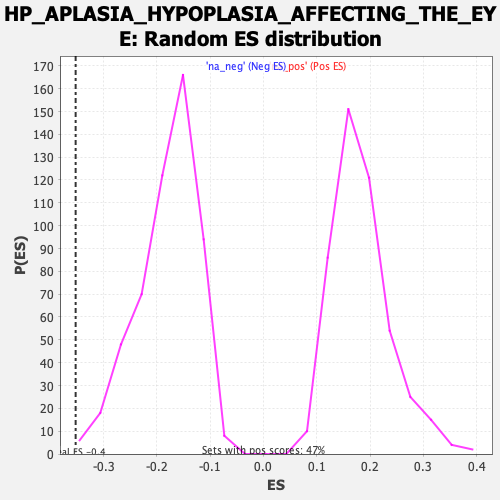

### gset_rnd_es_dist_520.png

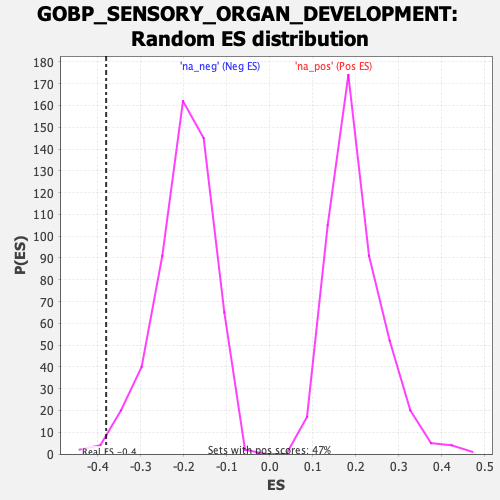

### gset_rnd_es_dist_522.png

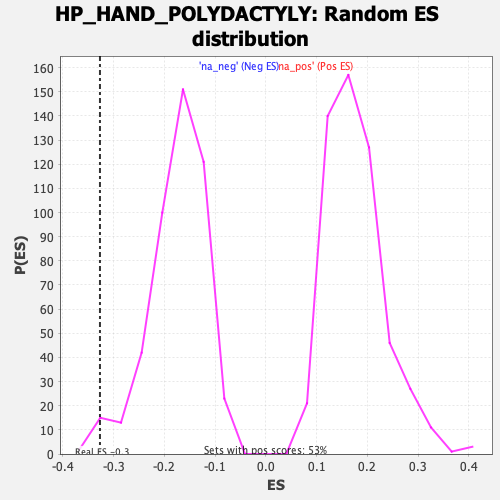

### gset_rnd_es_dist_524.png

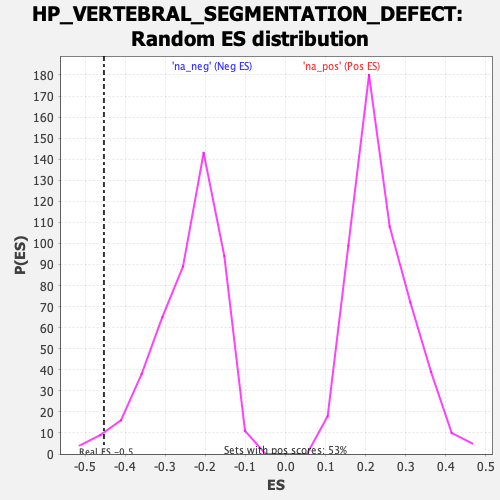

### gset_rnd_es_dist_526.png

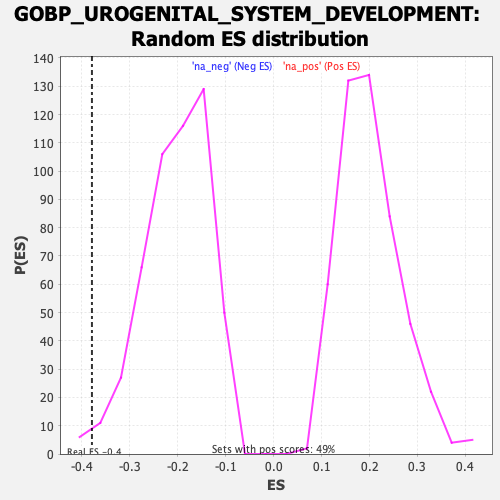

### gset_rnd_es_dist_528.png

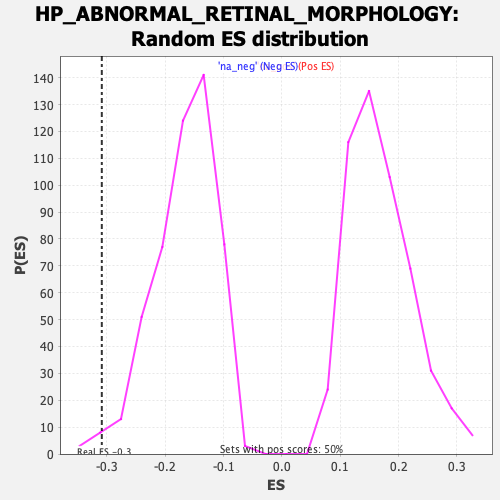

### gset_rnd_es_dist_530.png

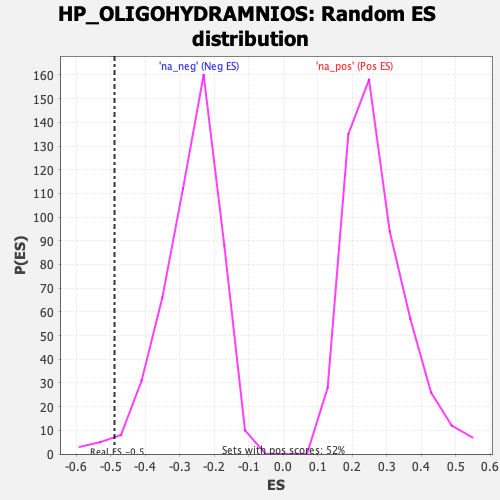

### gset_rnd_es_dist_532.png

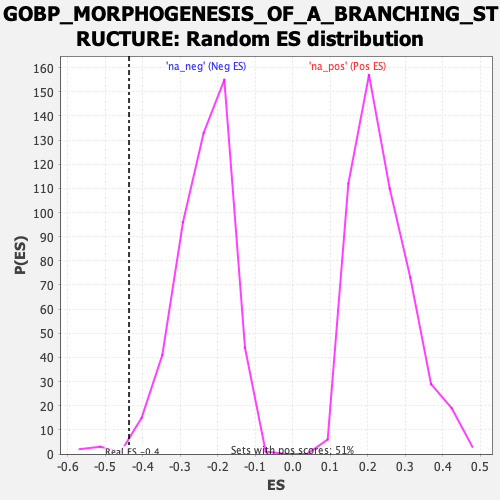

### gset_rnd_es_dist_534.png

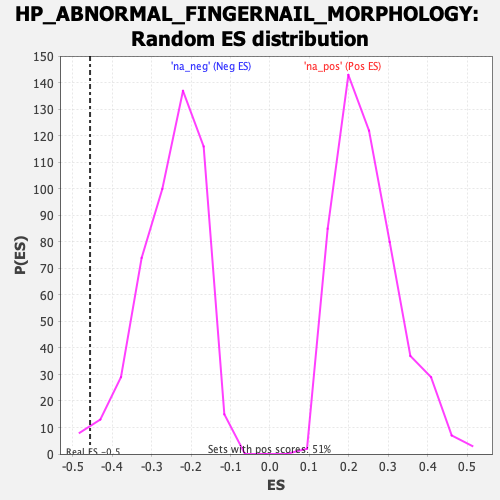

### gset_rnd_es_dist_536.png

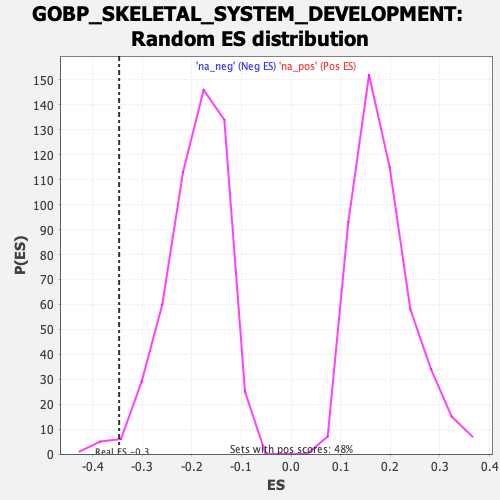

### gset_rnd_es_dist_538.png

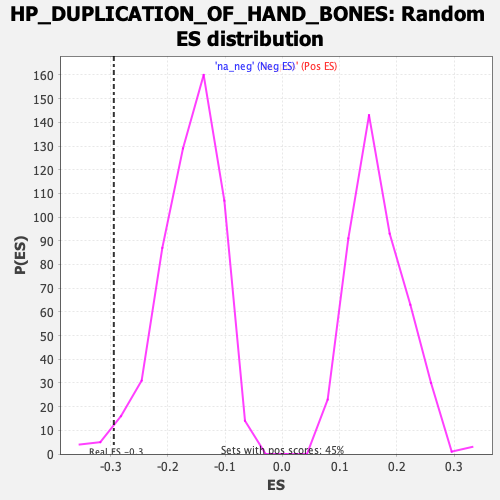

### gset_rnd_es_dist_540.png

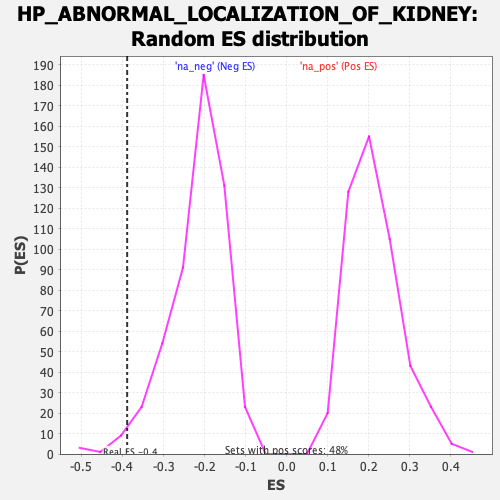

### gset_rnd_es_dist_542.png

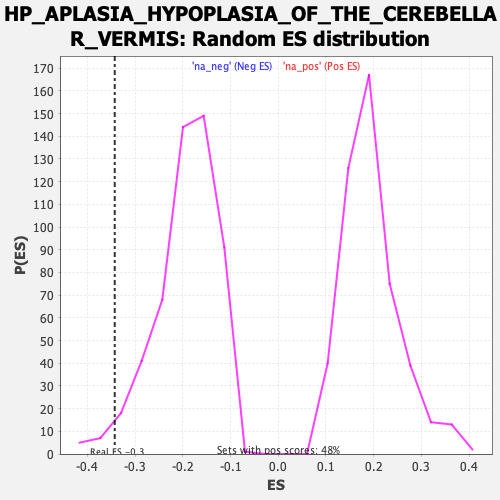

### gset_rnd_es_dist_544.png

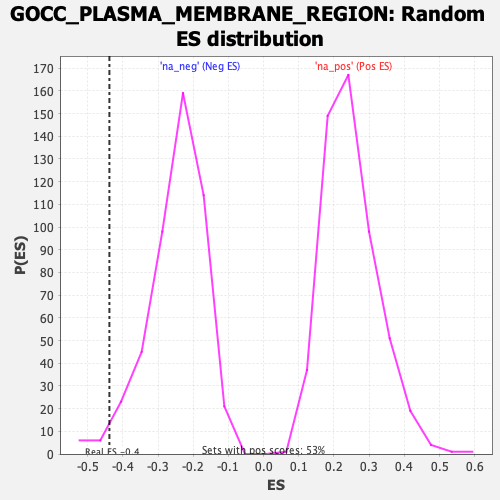

### gset_rnd_es_dist_546.png

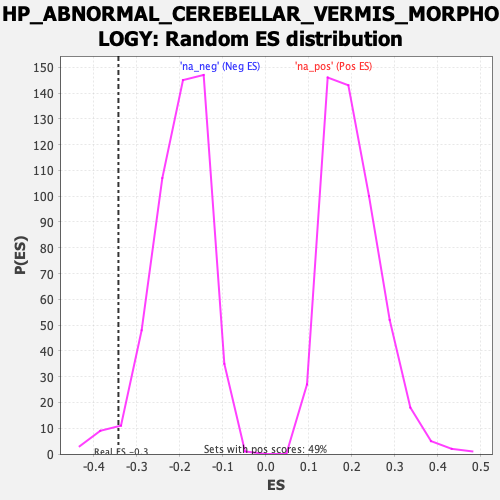

### gset_rnd_es_dist_550.png

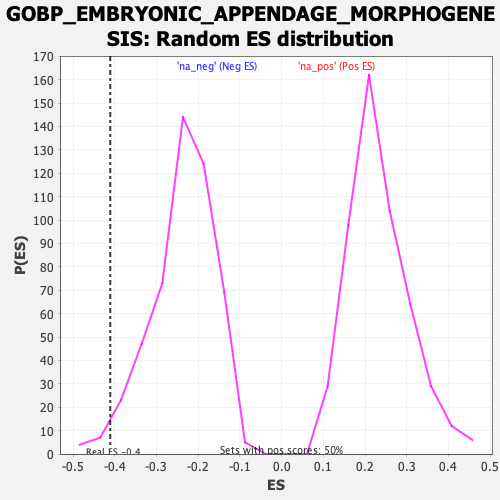

### gset_rnd_es_dist_552.png

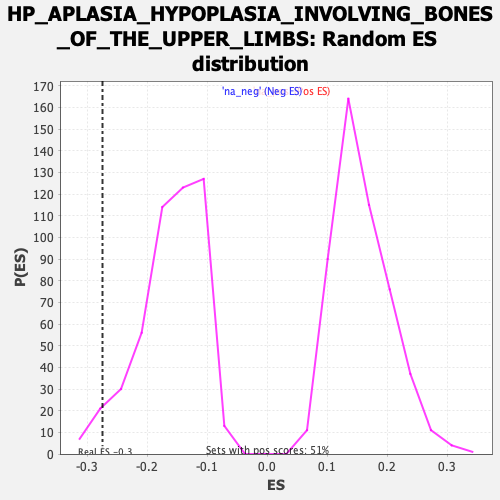

### pvalues_vs_nes_plot.png

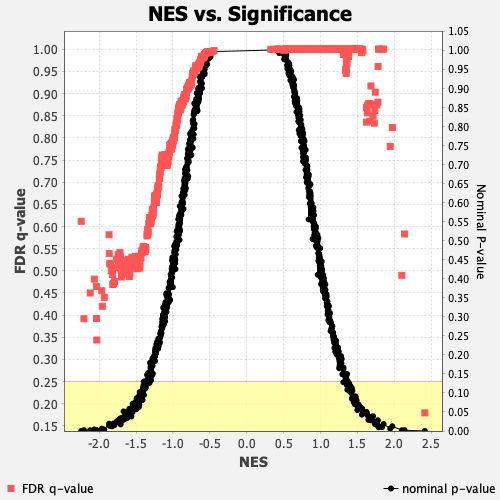
